# Exploring health in the UK Biobank: associations with sociodemographic characteristics, psychosocial factors, lifestyle and environmental exposures

**DOI:** 10.1101/2020.04.15.20066035

**Authors:** Julian Mutz, Charlotte J. Roscoe, Cathryn M. Lewis

## Abstract

A greater understanding of factors associated with good health may help increase longevity and healthy life expectancy. Here we report associations between multiple health indicators and sociodemographic (age, sex, ethnicity, education, income and deprivation), psychosocial (loneliness and social isolation), lifestyle (smoking, alcohol intake, sleep, BMI, physical activity and stair climbing) and environmental (air pollution, noise and greenspace) factors, using data from 307,378 UK Biobank participants. Low income, being male, neighbourhood deprivation, loneliness, social isolation, short or long sleep duration, low or high BMI and smoking was associated with poor health. Walking, vigorous-intensity physical activity and more frequent alcohol intake was associated with good health. There was some evidence that airborne pollutants (PM_2.5_, PM_10_, and NO_2_) and noise (L_den_) were associated with poor health, though findings were inconsistent in adjusted models. Our findings highlight the multifactorial nature of health, the importance of non-medical factors, such as loneliness, healthy lifestyle behaviours and weight management, and the need to examine efforts to improve health outcomes of individuals with low income.

## Introduction

Substantial improvements in human health and significant increases in average life expectancy are amongst the main achievements of civilization over the past two centuries^1^. Life expectancy at birth in England and Wales has doubled from about 40 years in 1850 to more than 80 years in 2013^2^. Future life expectancy is projected to increase across 35 industrialised nations, with a median increase in life expectancy at birth of approximately 3 years for women and 4 years for men in the United Kingdom between 2010–2030^3^.

Many adults, however, now spend a substantial portion of their lives with late-life morbidities and decreased sensory, motor and cognitive functioning, which may lead to lower quality of life^4,5^. More than 80% of the population of England and Wales aged 85 and above report a disability and 50% require care or help with daily activities^6^. Nevertheless, some individuals experience very little functional decline in old age and rate their health as good or excellent^7^. A greater understanding of the factors associated with good health may help increase longevity and healthspan, i.e. the length of time that a person is healthy^8^.

The health effects of lifestyle factors such as smoking^9^, diet^10^, excessive alcohol intake^11^ and physical activity^12^ are well established. However, the number and type of potential confounders that have been adjusted for in previous analyses vary widely^13^, and few studies have jointly examined sociodemographic, psychosocial, lifestyle and environmental factors. In addition, research has primarily focused on predicting mortality or disease incidence instead of overall health status, despite its potential to provide insights into the factors associated with increased healthspan.

The UK Biobank study provides an unprecedented data resource to investigate determinants of health and ageing trajectories. One of its strengths is that data collection was not merely focused on established predictors of health and disease but also included relevant non-medical factors such as social isolation and loneliness, as well as road traffic noise and ambient air pollution^14,15^.

The aim of this study was to explore potential predictors of health status. More specifically, we examined how health status was associated with (i) sociodemographic characteristics and psychosocial factors, (ii) lifestyle factors, and (iii) environmental exposures, both independently and jointly. We also examined, as a secondary aim, whether there was evidence of similar associations between these factors and (i) long-standing illness and (ii) self-rated health.

## Methods

### Study population

The UK Biobank is a prospective study of >500,000 UK residents aged 37–73 at baseline, recruited between 2006–2010. Details of the study rationale and design have been reported elsewhere^16^. Briefly, individuals registered with the UK National Health Service (NHS) and living within a 25- mile (∼40 km) radius of one of 22 assessment centres were invited to participate (9,238,453 postal invitations sent). At the baseline assessment, participants completed questionnaires and were interviewed by nurses regarding sociodemographic characteristics, health behaviours and their medical history. A small subset of participants completed repeat measurements: first revisit between 2012–2013 (20,344 participants); second revisit as part of the UK Biobank Imaging Study between 2014–2019 (43,190 participants). Participants consented to use of their de-identified data. See Supplement-e1 for the UK Biobank data fields used.

### Exposures

We considered individual-level sociodemographic characteristics including age at baseline assessment, sex, ethnicity (White, Asian, Black, Chinese, mixed-race or other), highest educational or professional qualification (four categories: (1) College/University degree; (2) A levels/AS levels or equivalent, NVQ/HND/HNC or equivalent, other professional qualifications; (3) O levels/GCSEs or equivalent, CSEs or equivalent; (4) none)^17^ and gross annual household income (<£18,000, £18,000– 30,999, £31,000–51,999, £52,000–100,000 or >£100 000). We also included the Index of Multiple Deprivation for England which is a small-area level measure derived from data on income deprivation, employment deprivation, health deprivation and disability, education skills and training deprivation, barriers to housing and services, living environment deprivation and crime^18^. Higher values on the index reflect greater deprivation.

Loneliness was assessed using two questions: “Do you often feel lonely?” (no=0/yes=1) and “How often are you able to confide in someone close to you?” (almost daily to about once a month=0/once every few months to never or almost never=1). Individuals who received a sum score of 2 were classified as lonely^19^. Social isolation was assessed using three questions: “Including yourself, how many people are living together in your household?” (living alone=1), “How often do you visit friends or family or have them visit you?” (less than once a month=1) and “Which of the following [leisure/social activities] do you attend once a week or more often?” (none of the above=1). Individuals who received a sum score of 2–3 were classified as socially isolated^19^.

Smoking status was assessed using two questions summarising current and past smoking behaviour. Individuals who responded “Yes, on most or all days” or “Only occasionally” to current tobacco smoking were coded as “current”. Individuals who responded “Smoked on most or all days” or “Smoked occasionally” to past tobacco smoking were coded as “former”. Individuals who responded “No” to current tobacco smoking and “Just tried once or twice” or “I have never smoked” to past tobacco smoking were coded as “never”. Alcohol intake frequency was assessed using one question: “About how often do you drink alcohol?”. Response options included “Daily or almost daily”, “Three or four times a week”, “Once or twice a week”, “One to three times a month”, “Special occasions only” and “Never”. Sleep duration was assessed using one question: “About how many hours sleep do you get in every 24 hours? (please include naps)”. Values below 1 hour or above 23 hours were rejected, and participants were asked to confirm values below 3 hours or above 12 hours. Body mass index (BMI) was calculated as weight divided by height squared (kg/m^2^). Weight measurements were obtained with a Tanita BC-418 MA body composition analyser. Standing height measurements were obtained using a Seca 202 height measure. Physical activity was assessed using the International Physical Activity Questionnaire (IPAQ) short form^20^. Specifically, we included data on the number of days per week spent walking, engaging in moderate-intensity physical activity (e.g. “carrying light loads, cycling at normal pace”) or engaging in vigorous-intensity physical activity (i.e. “activities that make you sweat or breathe hard such as fast cycling, aerobics, heavy lifting”) for ≥10 minutes continuously. Daily frequency of stair climbing at home was assessed using one question: “At home, during the last 4 weeks, about how many times a day do you climb a flight of stairs? (approx 10 steps)”. These data were only collected from individuals who indicated that they were able to walk. Response options included “None”, “1–5 times a day”, “6–10 times a day”, “11–15 times a day”, “16– 20 times a day” and “More than 20 times a day”.

Exposure to airborne pollutants was estimated using a Land Use Regression model developed as part of the European Study of Cohorts for Air Pollution Effects (ESCAPE) project^21,22^. We included annual average concentration of particulate matter with an aerodynamic diameter of <2.5 μm (PM_2.5_) and <10 μm (PM_10_), as well as nitrogen dioxide (NO_2_) modelled at participants’ residential addresses for the year 2010. Residential road traffic noise was modelled for the year 2009 using the Common Noise Assessment Methods (CNOSSOS-EU) algorithm^23,24^. We used L_den_ (day-evening-night noise level) which is an annual average 24-hour sound pressure level in decibels with a 10-decibel penalty added between 11pm and 7am. This penalty has previously been added in epidemiological analyses to account for annoyance/sleep disruption at night^25^. The percentage of the home location classed as greenspace, as a proportion of all land use types, was modelled using 2005 data from the Generalized Land Use Database for England (GLUD)^26^ for the 2001 Census Output Areas in England. Each residential address was allocated a circular distance buffer of 1000m, representing wider-area greenspace.

### Health status

Data on 81 cancer and 443 non-cancer illnesses (past and current) were ascertained through touchscreen self-report questionnaire and confirmed during a verbal interview by a trained nurse. In order to provide a single health indicator (“health status”) based on a previously defined algorithm, we used a classification developed by the Reinsurance Group of America (RGA) in which an experienced underwriter classified each illness according to whether it was “likely acceptable for standard life insurance”^27^. Participants were thus classified as healthy or unhealthy based on their reported cancer and non-cancer illnesses (Supplement-e1; Supplementary file).

Two secondary health outcomes were assessed. Firstly, whether patients had a long-standing illness, disability or infirmity was assessed using the question “Do you have any long-standing illness, disability or infirmity?” to which individuals could respond “Yes” or “No”. Secondly, participants’ perceived health was assessed using the question “In general how would you rate your overall health?”. Response options included “Poor”, “Fair”, “Good” and “Excellent”. These health outcomes will be termed “long-standing illness” and “self-rated health”.

### Exclusion criteria

Women who were pregnant at the time of assessment were excluded from the analysis based on the assumption that lifestyle patterns change during pregnancy^28^. Participants for whom their genetic sex, inferred from the relative intensity of biological markers on the Y and X chromosomes, and self-reported sex did not match were also excluded.

### Statistical analyses

Participants with baseline data on all explanatory variables and health indicators were selected for cross-sectional analyses. Participants with missing data or who responded “do not know” or “prefer not to answer” were excluded.

Logistic regression analyses were used to estimate associations between sociodemographic characteristics, psychosocial factors, lifestyle factors and environmental exposures with health status and long-standing illness. Ordinal logistic regression analyses were used to estimate associations between these factors and self-rated health. Across these outcomes, poor health was the reference group. In defining the reference groups for categorical explanatory variables, we focused on interpretability of the results for middle aged and older UK residents.

We fitted three incremental models using the baseline UK Biobank data: Model 1–individual explanatory variables; Model 2–individual explanatory variables plus age and sex; Model 3–all explanatory variables. All models were fitted in the analytical sample without missing data to ensure that any differences between models were not due to inclusion of different participants. We calculated odds ratios and Bonferroni-adjusted (∼99.9%) confidence intervals. Adjusted *p*-values were calculated to account for multiple testing within each model (i.e., typically for 39 tests). Two methods were used: (1) Bonferroni and (2) Benjamini & Hochberg^29^, all two-tailed with *α* = .05. We report in-text the most conservative correction at which statistical significance was reached. If the unadjusted *p*- value was greater than .05, in which case neither Bonferroni nor Benjamini & Hochberg-adjusted *p*- values would reach criteria for statistical significance, we report the unadjusted *p*-value. Multicollinearity was assessed using generalised variance inflation factors^30^.

To compare the magnitudes of association between the explanatory variables and health indicators, we calculated standardised regression coefficients by rescaling all variables included in Model 3 to have a mean of zero and by dividing the coefficients of numeric variables with more than two values by twice their standard deviation^31^.

To assess whether any associations between explanatory variables and health indicators were modified by sex or age, we stratified analyses by sex and by age at baseline assessment (<65 years and ≥65 years) and assessed potential age- and sex-interactions by adding cross-product terms to Model 3. The choice of age strata was based on the current UK retirement age.

We used ordinal logistic regression analyses to model associations between the explanatory variables at baseline and self-rated health at the first and second revisit (t1 and t2, respectively). We applied the same modelling strategy, except that we additionally adjusted for the number of days between baseline and follow-up assessment.

As variable selection and classification might impact results, we conducted multiple additional analyses to examine related variables, categorised continuous variables according to health guidelines or previously determined cut-offs and tested the robustness of our findings to additional exclusion criteria. We repeated the main analysis (i) with sleep duration as categorical variable (<7, 7-8 [reference] and ≥9 hours of sleep/day)^32^; (ii) with BMI as categorical variable (<18.5, 18.5-24.9 [reference] and >24.9 kg/m^2^); (iii) with body fat percentage (estimated by electrical bio-impedance measurement) instead of BMI as measure of body composition; (iv) excluding individuals who reported that they stopped drinking alcohol because of “Illness or ill health” or “Doctor’s advice”; (v) sub-dividing those individuals who reported that they never drink alcohol into current and lifetime abstainers; (vi) with current tobacco smoking only (three categories: “Yes, on most or all days”, “Only occasionally” and “No”); (vii) with Metabolic Equivalent Task (MET) minutes per week for walking, moderate physical activity and vigorous physical activity^33^ instead of the number of days per week spent engaging in these activities for ≥10 minutes; (viii) reducing the greenspace percentage circular distance buffer from 1000m to 300m to examine nearby greenspace; (ix) with air pollution and noise exposure estimates dichotomised according to WHO recommendation thresholds^34,35^: PM_2.5_ ≤10 μg/m^3^, PM_10_ ≤20 μg/m^3^, NO_2_ ≤40 μg/m^3^ and L_den_ ≤53 decibels; (x) restricting analyses to participants assessed after 31 December 2008 (regarding noise estimates) and restricting analyses to participants assessed after 31 December 2009 (regarding air pollution estimates); (xi) restricting analyses to those individuals who had lived at their current address for at least 10 years; (xii) truncating continuous explanatory variables at the 1st and 99th %ile of the distribution.

Statistical analyses were conducted using R (version 3.5.0).

## Results

### Study population

Of the *N* = 502,521 UK Biobank participants, 5.32% (*n* = 26,757) had missing health data and 33.51% (*n* = 168,386) had missing data on explanatory variables or did not meet our inclusion criteria. Hence our analytical sample included *n* = 307,378 adults (Supplement-e2).

Descriptive statistics are presented in Supplement-e3–e6. There were no substantial differences between the full and analytical sample. Of the participants in the analytical sample, 8.18% (*n* = 25,142) reported at least one cancer and 73.30% (*n* = 225,312) at least one non-cancer illness (Supplement-e4). Approximately two thirds of participants (69.04%, *n* = 212,201) were classified as healthy, while 30.96% (*n* = 95,177) were unhealthy. A similar percentage of participants (30.5%, *n* = 93,757) reported having a long-standing illness (72.9% agreement with health status). Finally, 3.60% (*n* = 11,066) rated their health as poor, while 19.25% (*n* = 59,169), 59.44% (*n* = 182,699) and 17.71% (*n* = 54,444) rated their health as fair, good and excellent, respectively.

### Regression analyses

We found weak correlations between most continuous explanatory variables and moderate to strong correlations between the environmental exposures (Supplement-e5). Generalised variance inflation factor (VIF) values for Model 3 were within the range of 1.02–2.62 for 19/21 explanatory variables. The VIFs for NO_2_ and PM_2.5_ were 6.08 and 4.27, respectively. Excluding NO_2_ from the model reduced the highest VIF to 2.42 (for PM_2.5_). Fitting Model 3 without NO_2_ or with each air pollutant separately had little impact on their associations with health status. As such, we report the results for Model 3 that included all explanatory variables. Unless indicated otherwise, results presented below correspond to multivariable-adjusted odds ratios and Bonferroni-adjusted (∼99.9%) confidence intervals from Model 3. A simplified overview of our findings is presented in Supplement-e7.

Increased age was associated with lower odds of favourable health status (OR = 0.953, 99.9% CI 0.951-0.955, *p*_Bonf._<0.001). For every 1-year increase in age, the log-odds of being healthy decreased by −0.048 (99.9% CI −0.050 to −0.046). Men had lower odds of being healthy (OR = 0.88, 99.9% CI 0.861-0.909, *p*_Bonf._<0.001). Individuals with high income had higher odds of being healthy (OR = 1.05, 99.9% CI 1.010-1.094, *p*_Bonf._ = 0.003 [£52,000–100,000 vs £31,000–51,999]), while those with lower levels of income had lower odds of being healthy (e.g. OR = 0.74, 99.9% CI 0.715-0.776, *p*_Bonf._<0.001 [<£18,000 vs £31,000–51,999]). Increased neighbourhood deprivation was associated with lower odds of being healthy (OR = 0.995, 99.9% CI 0.994-0.996, *p*_Bonf._<0.001). Compared to Whites, individuals of Black, Chinese, Mixed-race, and “other” ethnic background had higher odds of being healthy in Model 1, but only Chinese ethnicity was associated with higher odds of being healthy across all models (Model 3: OR = 1.83, 99.9% CI 1.365-2.511, *p*_Bonf._<0.001). Compared to individuals without educational or professional qualification, participants with any qualification had higher odds of being healthy in Model 1 and after adjustment for age and sex (Model 2). However, there was only limited evidence of associations between highest qualification and health in Model 3 (Table-1).

**Table 1.**
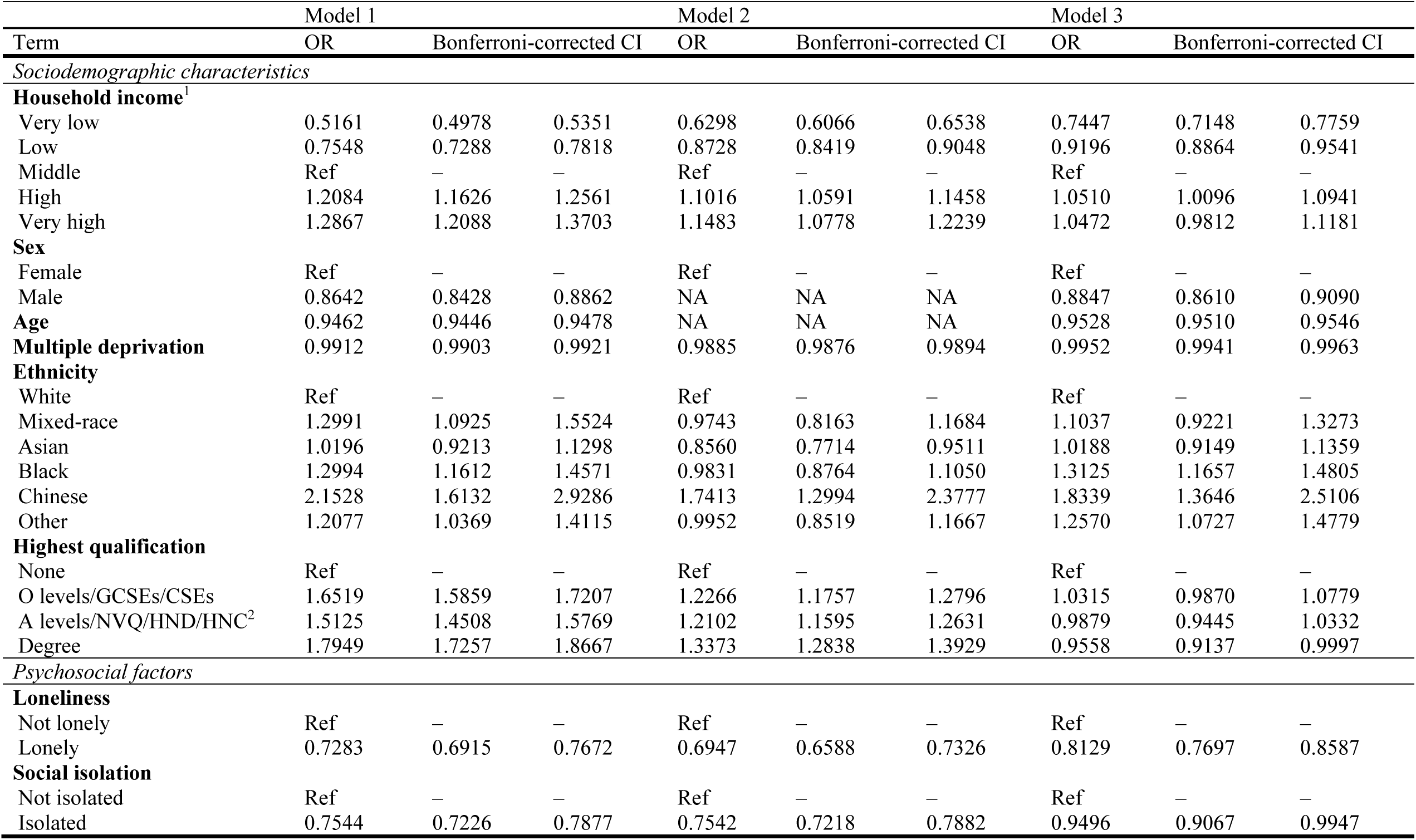
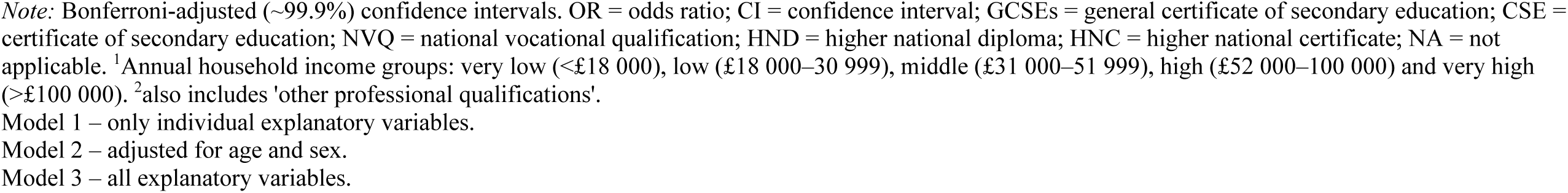
Sociodemographic characteristics and psychosocial factors associated with health status.

|  |  |  |  |  |  |  |  |  |  |
| --- | --- | --- | --- | --- | --- | --- | --- | --- | --- |
|  | Model 1 |  |  | Model 2 |  |  | Model 3 |  |  |
| Term | OR | Bonferroni-corrected CI |  | OR | Bonferroni-corrected CI |  | OR | Bonferroni-corrected CI |  |
| *Sociodemographic characteristics* |  |  |  |  |  |  |  |  |  |
| **Household income<sup>1</sup>** |  |  |  |  |  |  |  |  |  |
| Very low | 0.5161 | 0.4978 | 0.5351 | 0.6298 | 0.6066 | 0.6538 | 0.7447 | 0.7148 | 0.7759 |
| Low | 0.7548 | 0.7288 | 0.7818 | 0.8728 | 0.8419 | 0.9048 | 0.9196 | 0.8864 | 0.9541 |
| Middle | Ref | — | — | Ref | — | — | Ref | — | — |
| High | 1.2084 | 1.1626 | 1.2561 | 1.1016 | 1.0591 | 1.1458 | 1.0510 | 1.0096 | 1.0941 |
| Very high | 1.2867 | 1.2088 | 1.3703 | 1.1483 | 1.0778 | 1.2239 | 1.0472 | 0.9812 | 1.1181 |
| **Sex** |  |  |  |  |  |  |  |  |  |
| Female | Ref | — | — | Ref | — | — | Ref | — | — |
| Male | 0.8642 | 0.8428 | 0.8862 | NA | NA | NA | 0.8847 | 0.8610 | 0.9090 |
| **Age** | 0.9462 | 0.9446 | 0.9478 | NA | NA | NA | 0.9528 | 0.9510 | 0.9546 |
| **Multiple deprivation** | 0.9912 | 0.9903 | 0.9921 | 0.9885 | 0.9876 | 0.9894 | 0.9952 | 0.9941 | 0.9963 |
| **Ethnicity** |  |  |  |  |  |  |  |  |  |
| White | Ref | — | — | Ref | — | — | Ref | — | — |
| Mixed-race | 1.2991 | 1.0925 | 1.5524 | 0.9743 | 0.8163 | 1.1684 | 1.1037 | 0.9221 | 1.3273 |
| Asian | 1.0196 | 0.9213 | 1.1298 | 0.8560 | 0.7714 | 0.9511 | 1.0188 | 0.9149 | 1.1359 |
| Black | 1.2994 | 1.1612 | 1.4571 | 0.9831 | 0.8764 | 1.1050 | 1.3125 | 1.1657 | 1.4805 |
| Chinese | 2.1528 | 1.6132 | 2.9286 | 1.7413 | 1.2994 | 2.3777 | 1.8339 | 1.3646 | 2.5106 |
| Other | 1.2077 | 1.0369 | 1.4115 | 0.9952 | 0.8519 | 1.1667 | 1.2570 | 1.0727 | 1.4779 |
| **Highest qualification** |  |  |  |  |  |  |  |  |  |
| None | Ref | — | — | Ref | — | — | Ref | — | — |
| O levels/GCSEs/CSEs | 1.6519 | 1.5859 | 1.7207 | 1.2266 | 1.1757 | 1.2796 | 1.0315 | 0.9870 | 1.0779 |
| A levels/NVQ/HND/HNC<sup>2</sup> | 1.5125 | 1.4508 | 1.5769 | 1.2102 | 1.1595 | 1.2631 | 0.9879 | 0.9445 | 1.0332 |
| Degree | 1.7949 | 1.7257 | 1.8667 | 1.3373 | 1.2838 | 1.3929 | 0.9558 | 0.9137 | 0.9997 |
| *Psychosocial factors* |  |  |  |  |  |  |  |  |  |
| **Loneliness** |  |  |  |  |  |  |  |  |  |
| Not lonely | Ref | — | — | Ref | — | — | Ref | — | — |
| Lonely | 0.7283 | 0.6915 | 0.7672 | 0.6947 | 0.6588 | 0.7326 | 0.8129 | 0.7697 | 0.8587 |
| **Social isolation** |  |  |  |  |  |  |  |  |  |
| Not isolated | Ref | — | — | Ref | — | — | Ref | — | — |
| Isolated | 0.7544 | 0.7226 | 0.7877 | 0.7542 | 0.7218 | 0.7882 | 0.9496 | 0.9067 | 0.9947 |

*Note:* Bonferroni-adjusted (~99.9%) confidence intervals. OR = odds ratio; CI = confidence interval; GCSEs = general certificate of secondary education; CSE = certificate of secondary education; NVQ = national vocational qualification; HND = higher national diploma; HNC = higher national certificate; NA = not applicable. <sup>1</sup>Annual household income groups: very low (<£18 000), low (£18 000–30 999), middle (£31 000–51 999), high (£52 000–100 000) and very high (>£100 000). <sup>2</sup>also includes 'other professional qualifications'.
Model 1 – only individual explanatory variables.
Model 2 – adjusted for age and sex.
Model 3 – all explanatory variables.

Loneliness was associated with lower odds of favourable health status (OR = 0.81, 99.9% CI 0.770- 0.859, *p*_Bonf._<0.001). Socially isolated individuals also had lower odds of being healthy, but there was greater attenuation in the strength of association in Model 3 (OR = 0.95, 99.9% CI 0.907-0.995, *p*_Bonf._ = 0.013) (Table-1).

Longer sleep duration was associated with lower odds of favourable health status (OR = 0.97, 99.9% CI 0.963-0.987, *p*_Bonf._<0.001). For every 1-hour increase in sleep duration, the log-odds of being healthy decreased by −0.03 (99.9% CI −0.038 to −0.013). Walking frequently and engaging in frequent vigorous physical activity was also associated with higher odds of being healthy (OR = 1.01, 99.9% CI 1.003-1.018, *p*_Bonf._<0.001 and OR = 1.03, 99.9% CI 1.024-1.040, *p*_Bonf._<0.001, respectively). For every one-day increase in ≥10 minutes of vigorous physical activity, the log-odds of favourable health status increased by 0.03 (99.9% CI 0.024-0.039). Although moderate physical activity was associated with higher odds of being healthy in Model 1–2, we did not find evidence of an association in Model 3 (OR = 1.00, 99.9% CI 0.996-1.010, *p*_unadj._ = 0.13). Frequent daily stair climbing at home was associated with higher odds of being healthy (ranging from OR = 1.07, 99.9% CI 1.015-1.129, *p*_Bonf._ = 0.002 to OR = 1.19, 99.9% CI 1.110-1.272, *p*_Bonf._<0.001 [1-5 times/day and >20 times/day vs none, respectively]). We found some evidence that frequent alcohol intake was associated with higher odds of being healthy (e.g. OR = 1.06, 99.9% CI 1.020-1.010, *p*_Bonf._<0.001 [3-4 vs 1-2 times/week]), although for daily/almost daily alcohol drinking only in Model 2 (OR = 1.06, 99.9% CI 1.023-1.104, *p*_Bonf._<0.001), while infrequent alcohol intake was associated with lower odds of favourable health status (ranging from OR = 0.91, 99.9% CI 0.871-0.957, *p*_Bonf._<0.001 to OR = 0.63, 99.9% CI 0.595- 0.664, *p*_Bonf._<0.001 [1-3 times/month and never vs 1-2 times/week, respectively]). Higher BMI was associated with lower odds of being healthy (OR = 0.968, 99.9% CI 0.966-0.971, *p*_Bonf._<0.001). For every one-unit increase in kg/m^2^, the log-odds of being healthy decreased by −0.03 (99.9% CI −0.035 to −0.029). Past and current tobacco smoking was associated with lower odds of being healthy (OR = 0.79, 99.9% CI 0.770-0.815, *p*_Bonf._<0.001 and OR = 0.75, 99.9% CI 0.718-0.787, *p*_Bonf._<0.001, respectively) (Table-2).

**Table 2.** Lifestyle factors associated with health status.

| Table 2. Lifestyle factors associated with health status |  |  |  |  |  |  |  |  |  |
| --- | --- | --- | --- | --- | --- | --- | --- | --- | --- |
|  | Model 1 |  |  | Model 2 |  |  | Model 3 |  |  |
| Term | OR | Bonferroni-corrected CI |  | OR | Bonferroni-corrected CI |  | OR | Bonferroni-corrected CI |  |
| **Sleep duration** (hours/day) | 0.9574 | 0.9460 | 0.9688 | 0.9817 | 0.9699 | 0.9937 | 0.9748 | 0.9631 | 0.9868 |
| **Physical activity** (days/week)<sup>1</sup> |  |  |  |  |  |  |  |  |  |
| Walking | 1.0099 | 1.0034 | 1.0164 | 1.0258 | 1.0190 | 1.0326 | 1.0105 | 1.0032 | 1.0178 |
| Moderate activity | 1.0095 | 1.0040 | 1.0149 | 1.0253 | 1.0197 | 1.0310 | 1.0032 | 0.9964 | 1.0100 |
| Vigorous activity | 1.0571 | 1.0502 | 1.0642 | 1.0502 | 1.0431 | 1.0573 | 1.0319 | 1.0238 | 1.0400 |
| **Stair climbing frequency** |  |  |  |  |  |  |  |  |  |
| None | Ref | – | – | Ref | – | – | Ref | – | – |
| 1-5/day | 1.3009 | 1.2362 | 1.3688 | 1.0980 | 1.0422 | 1.1568 | 1.0704 | 1.0148 | 1.1289 |
| 6-10/day | 1.5529 | 1.4813 | 1.6278 | 1.3354 | 1.2725 | 1.4013 | 1.1756 | 1.1188 | 1.2352 |
| 11-15/day | 1.6271 | 1.5457 | 1.7128 | 1.3923 | 1.3211 | 1.4672 | 1.1714 | 1.1098 | 1.2363 |
| 16-20/day | 1.6494 | 1.5530 | 1.7520 | 1.4123 | 1.3280 | 1.5020 | 1.1650 | 1.0937 | 1.2410 |
| 20+/day | 1.7494 | 1.6390 | 1.8675 | 1.4402 | 1.3474 | 1.5396 | 1.1881 | 1.1097 | 1.2723 |
| **Alcohol intake frequency** |  |  |  |  |  |  |  |  |  |
| Never | 0.5651 | 0.5364 | 0.5954 | 0.5849 | 0.5545 | 0.6171 | 0.6285 | 0.5946 | 0.6644 |
| Special occasions | 0.7015 | 0.6705 | 0.7341 | 0.7109 | 0.6785 | 0.7448 | 0.7792 | 0.7429 | 0.8174 |
| 1-3/month | 0.9082 | 0.8678 | 0.9505 | 0.8730 | 0.8333 | 0.9146 | 0.9129 | 0.8708 | 0.9570 |
| 1-2/week | Ref | – | – | Ref | – | – | Ref | – | – |
| 3-4/week | 1.0652 | 1.0271 | 1.1047 | 1.1059 | 1.0656 | 1.1478 | 1.0590 | 1.0198 | 1.0998 |
| Daily/almost daily | 0.9293 | 0.8957 | 0.9643 | 1.0627 | 1.0232 | 1.1037 | 1.0334 | 0.9939 | 1.0745 |
| **BMI** (kg/m<sup>2</sup>) | 0.9561 | 0.9536 | 0.9587 | 0.9580 | 0.9554 | 0.9606 | 0.9683 | 0.9656 | 0.9711 |
| **Smoking status** |  |  |  |  |  |  |  |  |  |
| Never | Ref | – | – | Ref | – | – | Ref | – | – |
| Former | 0.6977 | 0.6792 | 0.7168 | 0.7879 | 0.7663 | 0.8100 | 0.7922 | 0.7698 | 0.8152 |
| Current | 0.7309 | 0.7003 | 0.7629 | 0.6850 | 0.6556 | 0.7158 | 0.7518 | 0.7183 | 0.7869 |

*Note:* Bonferroni-adjusted (~99.9%) confidence intervals. OR = odds ratio; CI = confidence interval; BMI = body mass index. <sup>1</sup>number of days per week engaging in these activities for 10+ minutes continuously.
Model 1 – only individual explanatory variables.
Model 2 – adjusted for age and sex.
Model 3 – all explanatory variables.

In our analyses of environmental exposures, higher PM_2.5_ concentration was associated with lower odds of favourable health status (OR = 0.97, 99.9% CI 0.941-0.991, *p*_Bonf._<0.001). PM_10_ was also associated with lower odds of being healthy in Model 1–2, but there was no evidence of an association between PM_10_ and health status in Model 3 (OR = 1.002, 99.9% CI 0.993-1.010, *p*_unadj._ = 0.47). NO_2_ was associated with lower odds of being healthy in Model 1–2. However, in Model 3, there was limited evidence that NO_2_ was associated with higher odds of being healthy (OR = 1.004, 99.9% CI 0.999-1.008, *p*_BH_ = 0.009). Ambient sound level was associated with lower odds of being healthy in Model 1–2, but there was no evidence of an association with health status in Model 3 (OR = 0.998, 99.9% CI 0.995-1.002, *p*_unadj._ = 0.09). Finally, we did not find consistent evidence of an association between percentage greenspace within a 1000m circular distance buffer and health status (Table-3).

**Table 3.**
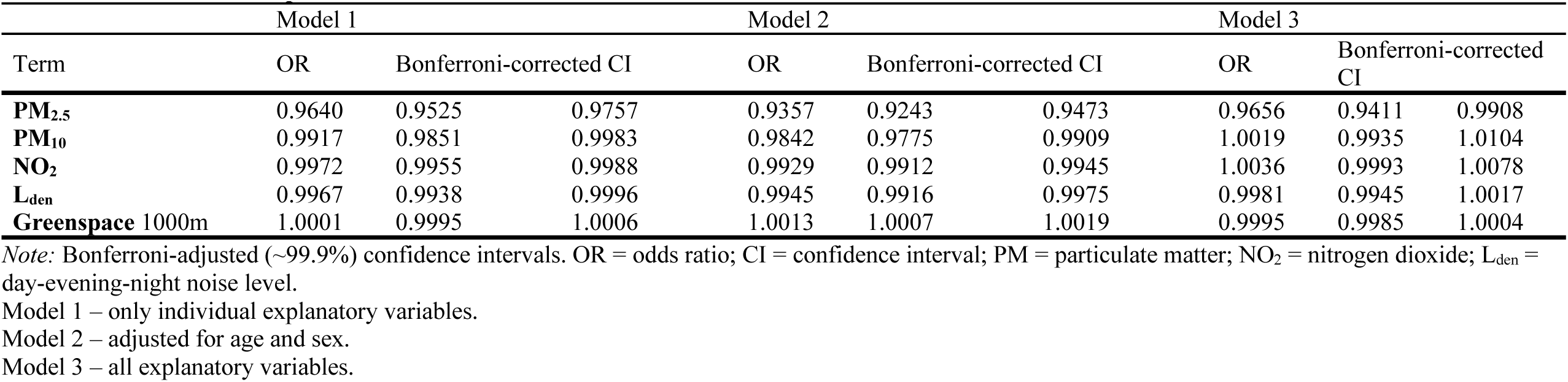
Environmental exposures associated with health status.

| Table 3. Environmental exposures associated with health status |  |  |  |  |  |  |  |  |  |
| --- | --- | --- | --- | --- | --- | --- | --- | --- | --- |
| Term | Model 1 |  |  | Model 2 |  |  | Model 3 |  |  |
|  | OR | Bonferroni-corrected CI |  | OR | Bonferroni-corrected CI |  | OR | Bonferroni-corrected CI |  |
| **PM<sub>2.5</sub>** | 0.9640 | 0.9525 | 0.9757 | 0.9357 | 0.9243 | 0.9473 | 0.9656 | 0.9411 | 0.9908 |
| **PM<sub>10</sub>** | 0.9917 | 0.9851 | 0.9983 | 0.9842 | 0.9775 | 0.9909 | 1.0019 | 0.9935 | 1.0104 |
| **NO<sub>2</sub>** | 0.9972 | 0.9955 | 0.9988 | 0.9929 | 0.9912 | 0.9945 | 1.0036 | 0.9993 | 1.0078 |
| **L<sub>den</sub>** | 0.9967 | 0.9938 | 0.9996 | 0.9945 | 0.9916 | 0.9975 | 0.9981 | 0.9945 | 1.0017 |
| **Greenspace 1000m** | 1.0001 | 0.9995 | 1.0006 | 1.0013 | 1.0007 | 1.0019 | 0.9995 | 0.9985 | 1.0004 |

*Note:* Bonferroni-adjusted (~99.9%) confidence intervals. OR = odds ratio; CI = confidence interval; PM = particulate matter; NO<sub>2</sub> = nitrogen dioxide; L<sub>den</sub> = day-evening-night noise level.
Model 1 – only individual explanatory variables.
Model 2 – adjusted for age and sex.
Model 3 – all explanatory variables.

Results for long-standing illness and self-rated health are described in the Supplementary text and presented in Supplement-e8–e10. Model 3 results across health indicators are presented in Figure-1–3.

**Figure 1.**
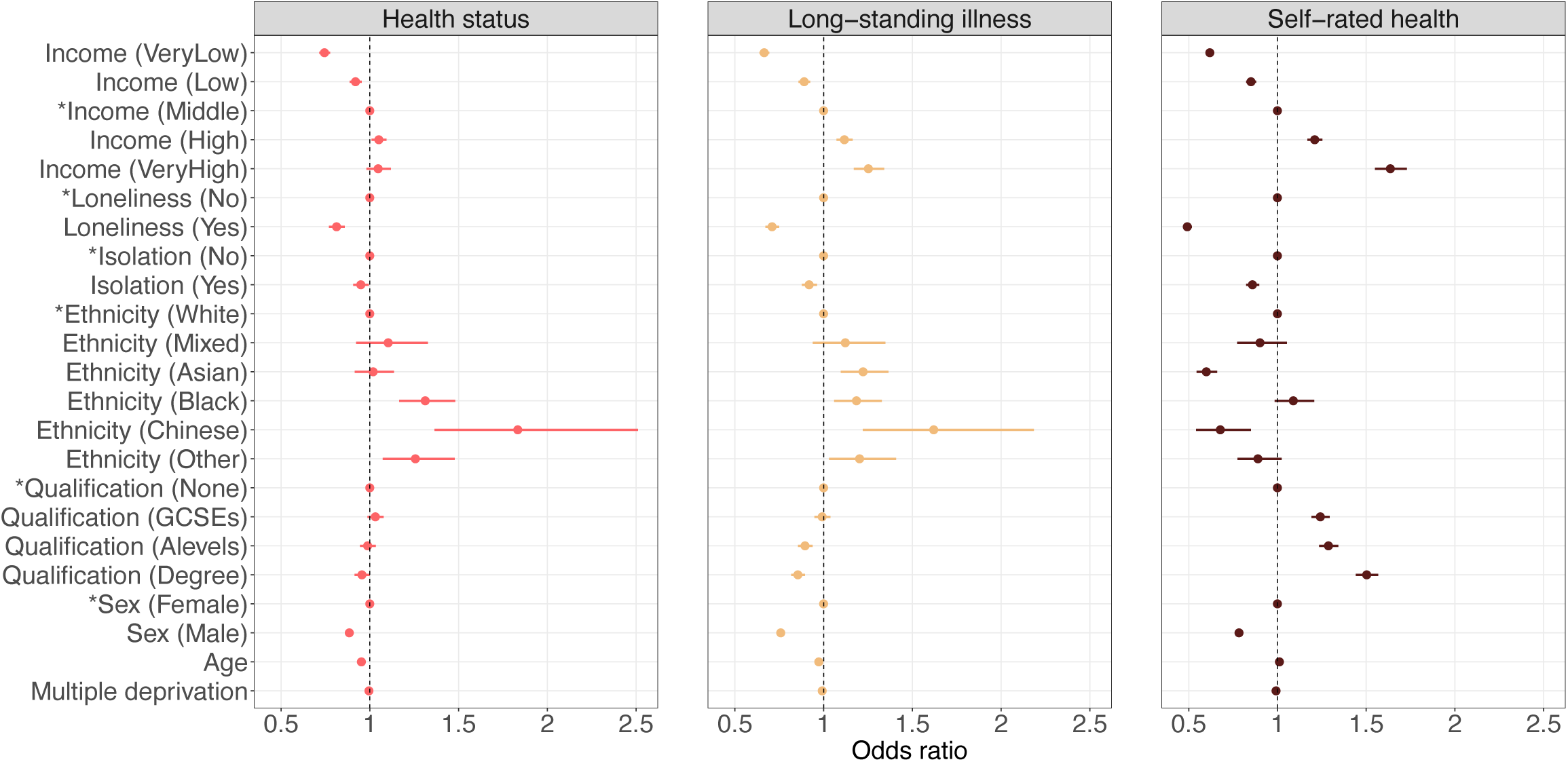
Sociodemographic characteristics and psychosocial factors associated with health indicators. Confidence interval plot (odds ratio ± Bonferroni-adjusted (∼99.9%) confidence intervals) for Model 3 (i.e. including all explanatory variables). GCSEs = general certificate of secondary education. ^*^Indicates reference group for categorical explanatory variables. For categorical explanatory variables the odds ratios for self-rated health indicate the changes in odds of reporting better self-rated health associated with the explanatory variable group relative to the reference group. Odds ratios for continuous explanatory variables for self-rated health indicate proportional odds ratios for a 1-unit increase in the explanatory variable on level of self-rated health. Annual household income groups: very low (<£18 000), low (£18 000–30 999), middle (£31 000–51 999), high (£52 000–100 000) and very high (>£100 000). ‘GCSEs’ also includes O levels and certificate of secondary education (CSE). ‘A levels’ also includes national vocational qualification (NVQ), higher national diploma (HND), higher national certificate (HNC) and ‘other professional qualifications’.

**Figure 2.**
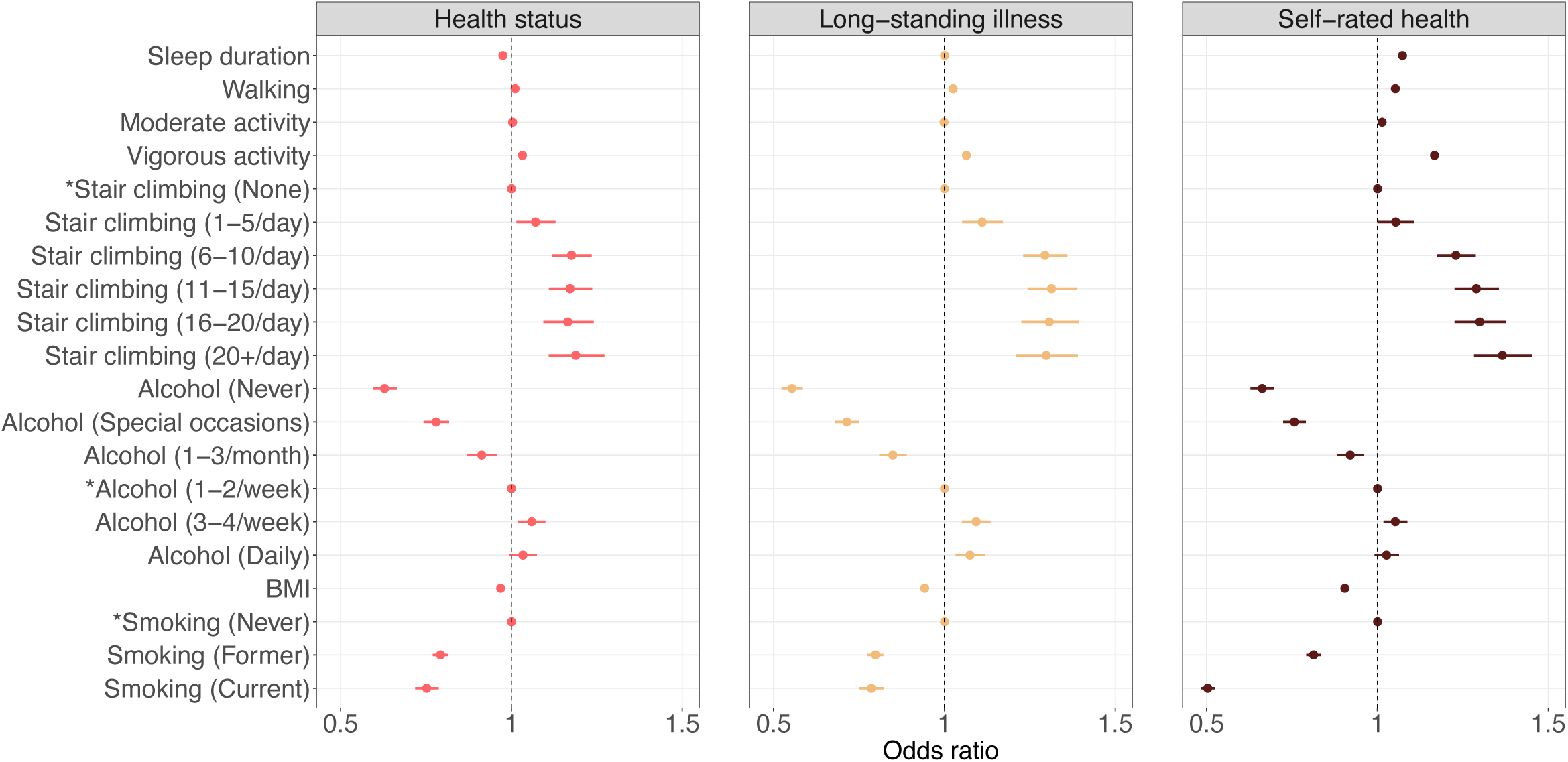
Lifestyle factors associated with health indicators. Confidence interval plot (odds ratio ± Bonferroni-adjusted (∼99.9%) confidence intervals) for Model 3 (i.e. including all explanatory variables). BMI = body mass index. ^*^Indicates reference group for categorical explanatory variables. For categorical explanatory variables the odds ratios for self-rated health indicate the changes in odds of reporting better self-rated health associated with the explanatory variable group relative to the reference group. Odds ratios for continuous explanatory variables for self-rated health indicate proportional odds ratios for a 1-unit increase in the explanatory variable on level of self-rated health.

**Figure 3.**
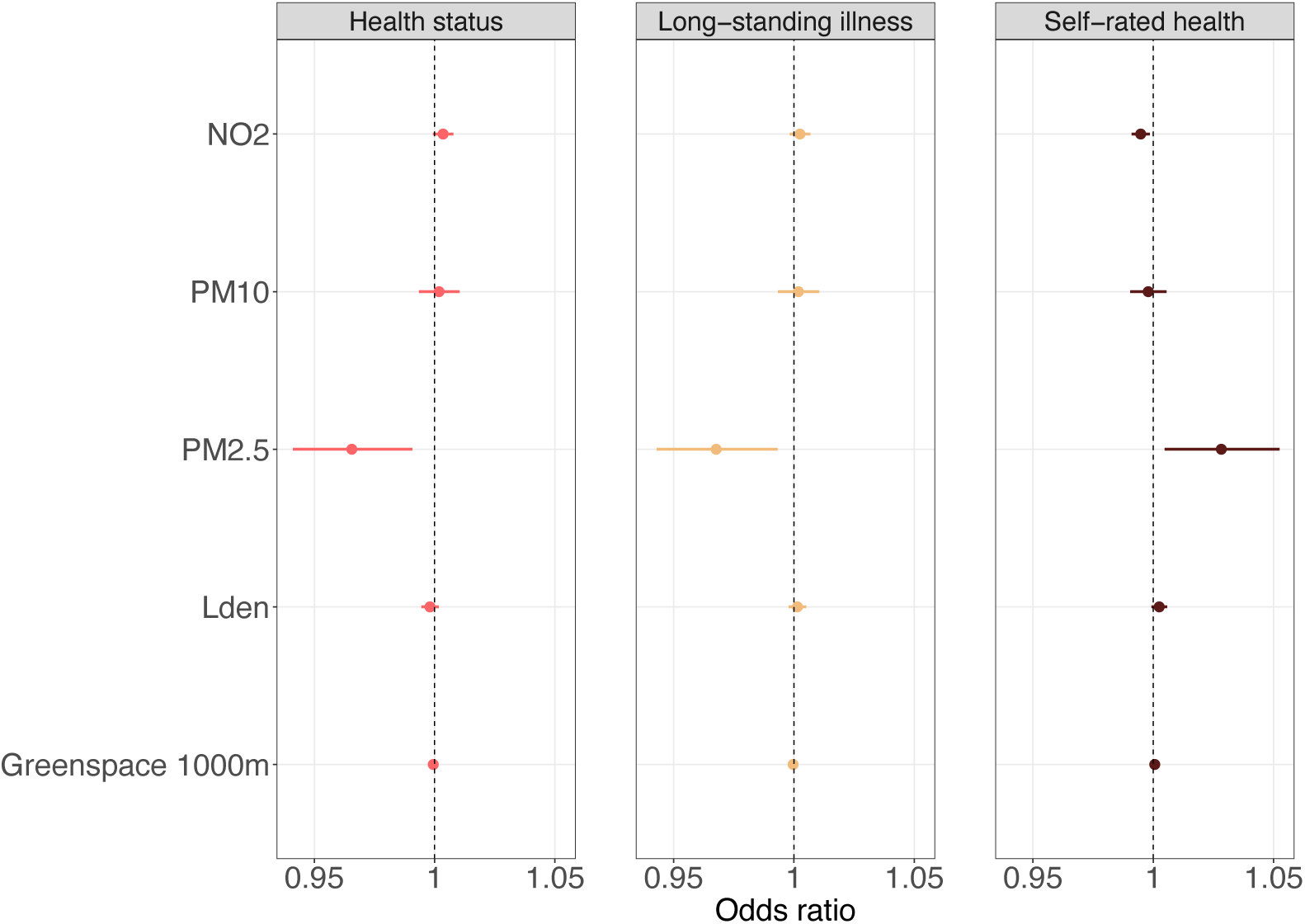
Environmental exposures associated with health indicators. Confidence interval plot (odds ratio ± Bonferroni-adjusted (∼99.9%) confidence intervals) for Model 3 (i.e. including all explanatory variables). PM = particulate matter; NO_2_ = nitrogen dioxide; L_den_ = day-evening-night noise level. Odds ratios for self-rated health indicate proportional odds ratios for a 1-unit increase in the explanatory variable on level of self-rated health.

### Magnitude of associations

Standardised regression coefficients for health status are presented in Figure-4. Most notably, the magnitude of association between BMI (*β* = −0.30, 99.9% CI −0.33 to −0.27) or very low income (*β* = - 0.29, 99.9% CI −0.34 to −0.25) and health status was comparable to that of current smoking (*β* = −0.29, 99.9% CI −0.33 to −0.24). There was also a substantial difference in magnitude of association with health status between the very low and low income groups (*β* = −0.29, 99.9% CI −0.34 to −0.25, and *β* = −0.08, 99.9% CI −0.12 to −0.05, respectively). The magnitude of association between loneliness and health status was substantially larger than that of social isolation (*β* = −0.21, 99.9% CI −0.26 to −0.15, and *β* = −0.05, 99.9% CI −0.10 to −0.01, respectively). Finally, associations between environmental exposures and health status were relatively small compared to those observed for sociodemographic, psychosocial and lifestyle factors (Supplement-e11).

**Figure 4.**
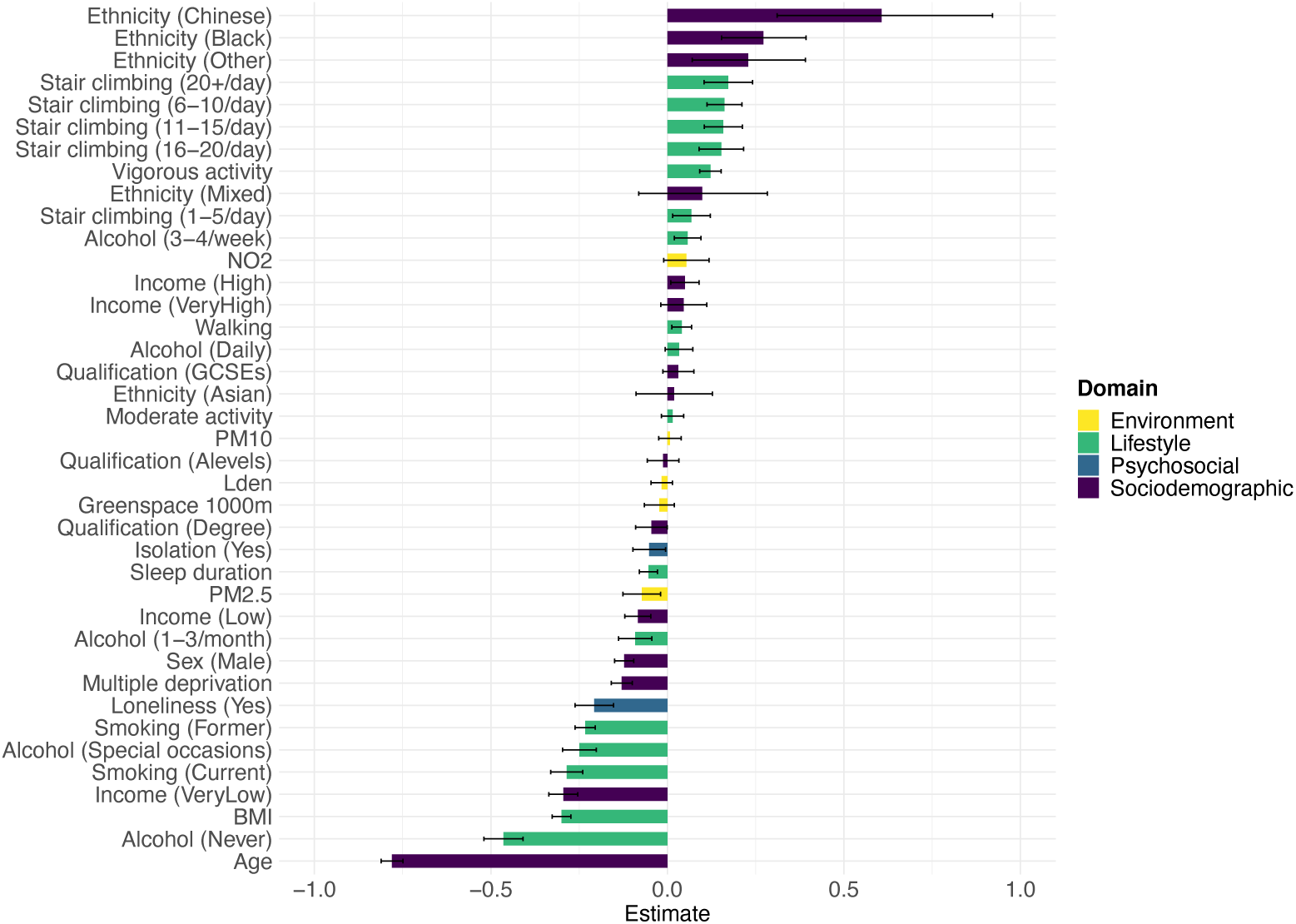
Horizontal bar plot for health status. *β* estimates and Bonferroni-adjusted (∼99.9%) confidence intervals. Model 3 regression coefficients rescaled to have a mean equal to zero and, for numeric variables with more than two values, divided by two standard deviations.

Associations with long-standing illness and self-rated health compared to health status were stronger for household income, sex, neighbourhood deprivation, loneliness, social isolation, walking frequency, vigorous physical activity, BMI and stair climbing frequency (Supplement-e11–e12).

### Stratified and interaction analyses

Descriptive statistics of the analytical sample stratified by sex and age group are presented in Supplement-e13–e14.

The odds of being healthy were lower for men than for women in the lowest income group (*p*_Bonf.(interaction)_<0.001). The association between increased age and lower odds of being healthy was stronger in men (*p*_Bonf.(interaction)_<0.001). Asian women had higher odds of being healthy (*p*_BH(interaction)_ = 0.007). The association between neighbourhood deprivation and lower odds of being healthy was stronger in men (*p*_Bonf.(interaction)_<0.001). Men who were lonely had lower odds of being healthy (*p*_Bonf.(interaction)_ = 0.008) and social isolation was associated with lower odds of being healthy only in men (*p*_BH(interaction)_ = 0.006). Longer sleep duration was associated with lower odds of being healthy only in men (*p*_Bonf.(interaction)_<0.001). Reporting frequent alcohol intake was associated with higher odds of being healthy only in men (*p*_BH(interaction)_ = 0.023 for 3-4 times/week and *p*_BH(interaction)_ = 0.039 for daily/almost daily). The association between higher BMI and lower odds of being healthy was stronger in men (*p*_Bonf.(interaction)_<0.001). The association between former smoking and health status was stronger in men (*p*_Bonf.(interaction)_<0.001), while the association between current smoking and health status was stronger in women (*p*_BH(interaction)_ = 0.005). PM_2.5_ was associated with lower odds of being healthy only in men (*p*_BH(ineraction)_ = 0.011). Finally, we found that NO_2_ was associated with higher odds of favourable health status only in women (*p*_BH(interaction)_ = 0.024) (Supplement-e15–e16).

The association between very low income and lower odds of being healthy was stronger in participants below the age of 65 than in participants aged 65 and above (*p*_Bonf.(interaction)_<0.001). Reporting an income of £52,000–100,000 was associated with higher odds of being healthy only in participants younger than 65 (*p*_BH(interaction)_ = 0.016). Men aged 65 and above had lower odds of being healthy (*p*_Bonf.(interaction)_<0.001). Loneliness and social isolation were associated with lower odds of being healthy only in individuals younger than 65 (*p*_BH(interaction)_ = 0.024 and *p*_Bonf.(interaction)_ = 0.011, respectively). The association between walking frequency and higher odds of being healthy was stronger in individuals aged 65 and above (*p*_Bonf.(interaction)_<0.001). Climbing stairs 1-5 times/day was associated with favourable health status only in individuals younger than 65 (*p*_Bonf.(interaction)_<0.001). There was some evidence that the association between never drinking alcohol and poor health status was stronger in individuals younger than 65 (*p*_Bonf.(interaction)_ = 0.002). The association between higher BMI and lower odds of being healthy was stronger in individuals aged 65 and above (*p*_BH(interaction)_<0.001). Finally, the association between current smoking and poor health status was stronger in individuals younger than 65 (*p*_BH(interaction)_ = 0.038) (Supplement-e17–e18).

Although there were some differences, stratified analyses of long-standing illness and self-rated health indicated a large degree of consistency with the results observed for health status (Supplement- e19–e26).

### Longitudinal analyses

We repeated our analyses of self-rated health in participants with follow-up data collected between 2012–2013 (*n* = 16,058; mean follow-up = 4.26 years, SD = 0.87) and between 2014–2019 (*n* = 32,617; mean follow-up = 8.57 years, SD = 1.64). Descriptive statistics and full results are presented in Supplement-e27–e29. The findings were fully consistent with cross-sectional analyses for very low and high to very high levels of income, sex, loneliness, sleep duration, walking frequency, vigorous physical activity, infrequent alcohol intake, BMI and smoking status. Although many results were consistent across timepoints, we found some differences between cross-sectional and prospective analyses for low income, age, neighbourhood deprivation, ethnicity, highest qualification, social isolation, moderate physical activity, stair climbing frequency, regular alcohol intake and environmental exposures.

### Additional analyses

Descriptive statistics are presented in Supplement-e30.

For sleep duration and BMI, we found evidence of non-linearity in the association with health status (Supplement-e31). Hence, we also examined these explanatory variables as categorical variables. Compared to individuals with optimal sleep duration (7-8 hours/day), those who slept <7 or ≥9 hours/day had lower odds of being healthy (OR = 0.90, 99.9% CI 0.868-0.923, *p*_Bonf._<0.001 and OR = 0.70, 99.9% CI 0.667-0.736, *p*_Bonf._<0.001, respectively). We also found that low (<18.5 kg/m^2^) and high (>24.9 kg/m^2^) BMI was associated with lower odds of being healthy, compared to a BMI of 18.5-24.9 kg/m^2^ (OR = 0.70, 99.9% CI 0.584-0.852, *p*_Bonf._<0.001 and OR = 0.85, 99.9% CI 0.827- 0.877, *p*_Bonf._<0.001, respectively). There was no evidence of substantial departure from a linear association with health status for all other numerical variables with more than two values (Supplement-e31). Repeating the main analysis with body fat percentage as measure of body composition led to similar conclusions as for BMI: higher fat percentage was associated with lower odds of being healthy (*n* = 307,202; OR = 0.978, 99.9% CI 0.975-0.980, *p*_Bonf._<0.001). Excluding individuals who reported that they had stopped drinking alcohol due to illness or their doctor’s advice (*n* = 2,790) attenuated the association between never drinking alcohol and health status (*n* = 304,588; OR = 0.74, 99.9% CI 0.701-0.790, *p*_Bonf._<0.001). When sub-dividing individuals who reported that they never drink alcohol into current and lifetime abstainers, the association with health status was stronger in current abstainers (*n* = 307,346; OR = 0.53, 99.9% CI 0.491-0.568, *p*_Bonf._<0.001 and OR = 0.75, 99.9% CI 0.698-0.810, *p*_Bonf._<0.001, respectively). Restricting analyses to current smoking, we found that regular smoking and smoking only occasionally was associated with lower odds of being healthy, although the magnitude of association was stronger for regular smoking (*n* = 307,372; OR = 0.81, 99.9% CI 0.766-0.848, *p*_Bonf._<0.001 and OR = 0.90, 99.9% CI 0.835-0.980, *p*_Bonf._ = 0.002, respectively). Examining MET minutes instead of the number of days per week spent walking, engaging in moderate or vigorous physical activity led to similar results (*n* = 268,674; data not shown).

When we examined air pollution and noise exposure estimates dichotomised according to WHO recommendation thresholds, we found some evidence that being exposed to ≤10 μg/m^3^ PM_2.5_ on average annually was associated with higher odds of being healthy (OR = 1.03, 99.9% CI 0.997- 1.062, *p*_BH_ = 0.005). There was no evidence of an association with health status for exposures to ≤20 μg/m^3^ PM_10_, ≤40 μg/m^3^ NO_2_ or ≤53 decibels L_den_ in Model 3. We also did not find evidence of a threshold in the association between PM_10_ or NO_2_ and health status when including quintiles of these exposures in Model 3 (data not shown). Restricting analyses to participants assessed after 2008 (regarding noise estimates; *n* = 182,342) or 2009 (regarding air pollution estimates; *n* = 60,521) or to those who had lived at their current address for at least 10 years (*n* = 209,239) did not lead to different conclusions regarding the associations between environmental exposures and health status. There was some evidence that percentage greenspace within a 300m circular distance buffer was associated with health status (*n* = 307,378; OR = 0.999, 99.9% CI 0.9985-1.0001, *p*_BH_ = 0.009). Truncating continuous explanatory variables at the 1st and 99th %ile did not materially change our results (*n* = 278,065; data not shown).

## Discussion

Increased age was strongly associated with unfavourable health status and reporting a long-standing illness. However, older participants generally rated their health positive, which is consistent with several smaller studies^36,37^, though not all^38,39^. Ageing might be the single most important factor underlying disease^40^, with an almost universally-accepted expectation of declining health as people get older. As attainable health states shift with age^41^, older participants might evaluate their health more favourable, despite higher rates of illness and disability.

Although women, on average, report more illnesses, disabilities, and limitations in daily life^42-44^, one of the most robust findings in human biology is that they live longer^45^. Our findings are consistent with results from the Newcastle 85+ cohort study in which women rated their health more favourably than men^42^. However, other studies reported that women rated their health less favourably than men^46-48^, or did not find evidence of sex differences in self-rated health^49,50^. Sex differences in health status could result from differences in the frequency of specific illnesses, sex-specific reporting patterns, or biological and social factors^48^. Discrepancies in findings between studies might reflect differences in age group or socio-cultural factors.

High income and low levels of neighbourhood deprivation were associated with better health, broadly consistent with previous studies^51-53^. Notably, we found only a small difference in the strength of association with health status and long-standing illness between the high-income groups. The difference between the low-income groups, however, was substantial, supporting previous findings which suggested a non-linear association between family income and mortality^54^. For self-rated health, we also found evidence of substantial differences between the high-income groups.

Our study provides limited evidence that education was associated with favourable health status once other factors had been accounted for, although higher levels of qualification were associated with better self-rated health, consistent with previous research^38^. A recent UK Biobank analysis showed that remaining longer in school causally reduced participants’ risk of diabetes and mortality^55^. A potential explanation for why we did not find a consistent pattern in the full model is that differences in health status result from educated individuals engaging in healthier lifestyle behaviours^56^ that we had accounted for, or they could be due to socioeconomic or genomic factors.

Social isolation and loneliness were associated with poor health. The strength of association was greater for loneliness, particularly in men and individuals below the age of 65. Social isolation and loneliness are not always correlated^57,58^ and represent different aspects of social relations (scarcity of contact with others and discrepancies between the need for, and the fulfilment of, social interaction, respectively). In a meta-analysis of 70 studies, social isolation, loneliness, and living alone were associated with a 26-32% increased mortality risk^14^. There was no evidence of differences in mortality between these measures, although the strength of association was greater in individuals below the age of 65. A recent UK Biobank analysis found that socially isolated and lonely individuals had an increased risk of death, but only social isolation predicted all-cause mortality in a joint model^19^. The discrepancy with our finding (loneliness was more strongly associated with poor health) might reflect differences in outcome measures (general health in the present study vs mortality in previous investigations).

Long sleep duration, higher BMI as well as past and current smoking was associated with poor health. Sleeping less than 7 hours/day was also associated with poor health, consistent with a meta-analysis that provided evidence of a U-shaped association between sleep duration and all-cause mortality^32^. A BMI outside the optimal range of 18.5-24.9 kg/m^2^ was also associated with poor health.

Physical activity is a key lifestyle factor recommended for primary and secondary prevention of chronic health conditions^59^, and is associated with lower mortality risk^60^. In this study, walking frequency, especially in individuals aged 65 and above, stair climbing, and engaging in vigorous physical activity was associated with good health. Moderate physical activity was associated with better self-rated health, especially in men. A study in middle-aged British men found evidence of an association between vigorous, but not moderate, physical activity and reduced mortality^61^. Reviews of the literature report mixed findings on the relative contributions of moderate and vigorous physical activity^62,63^, with some evidence suggesting stronger associations for vigorous activity^64^.

A more frequent drinking pattern was associated with good health. Alcohol drinking often occurs in a social context and might therefore constitute a proxy for social wellness, supported by the finding that non-drinkers tend to be characterised by poor psychosocial health and low socioeconomic status. Moderate drinkers also perform more sports than lifelong abstainers^65^. Drinking less frequent than 1-2 times/week was associated with poor health and could not be fully accounted for by excluding individuals who had discontinued alcohol intake for health reasons or because of their doctor’s advice. However, the association with health status was stronger for current abstainers, suggesting that some individuals are non-drinkers in later life due to illness^66^. Current abstainers who quit drinking for health reasons could exaggerate poor health outcomes associated with not drinking alcohol^67^. Those who never drink might also differ from current drinkers in other characteristics^68^.

Road traffic noise and ambient air pollution are environmental risk factors for health. While we found some evidence that higher levels of all airborne pollutants were associated with poor health, only PM_2.5_ was associated with unfavourable health status and long-standing illness after adjustment for other factors. Higher levels of NO_2_ were associated with less favourable self-rated health. A joint analysis of UK Biobank, HUNT, and EPIC-Oxford data did not find evidence of an association between NO_2_ and any disease outcome, although both PM_2.5_ and PM_10_ were associated with higher incident cardiovascular disease^69^. Adjustment for physical activity and neighbourhood deprivation in the present study could have contributed to differences in findings between studies. Few individuals in our study were exposed to high levels of PM_10_ and NO_2_, which might explain why we did not find strong associations between these pollutants and health. The only exception was PM_2.5_, for which 45% of our sample were exposed to levels above the WHO recommendation threshold of ≤10 μg/m^3^. Positive associations between PM_2.5_ and self-rated health and between NO_2_ and favourable health status might represent spurious associations or statistical artefacts; we are not aware of any mechanisms linking higher levels of pollution to good health. While higher levels of residential noise were associated with poor health in Model 1-2, there was no evidence of an association after adjustment for other factors. Our findings are consistent with a previous analysis that did not find evidence of an independent association between L_den_ and cardiovascular disease, ischemic heart disease or cerebrovascular disease^69^.

We found some evidence that greenspace was associated with better self-rated health, consistent with a recent meta-analysis^70^, although not after adjustment for other factors. There were also some inconsistencies in the prospective analyses and regarding health status and long-standing illness. Several previous studies examining associations between greenspace and health did not adjust for physical activity and most did not include other environmental exposures such as air pollution or noise^70^. These differences could explain why we did not find consistent associations in this study.

### Strengths and limitations

The large sample size enabled high precision in the estimation of associations and it allowed us to explore a range of explanatory variables, sex and age-specific associations, interactions and additional analyses with further classification of explanatory variables. Many of the findings from previous studies that we replicated in our analyses were conducted in smaller populations and with one exposure at a time, often providing limited insight into the multifactorial nature of health. One of the contributions of this study is that it allows for systematic comparisons of a broad range of factors associated with health. We examined three health indicators with slightly different ascertainment. While our findings were fairly consistent across these outcomes, we also identified differences between objective and subjective health. Overall health is arguably what matters most to people, and exploring factors associated with health indicators that transcend traditional disease-boundaries could represent an effective strategy to increase longevity and improve healthy life expectancy. Previous studies have often used composite scores of healthy lifestyles^13^ in which individual behaviours were weighted equally, even though the strength of their respective associations with health might differ. In the present study, we estimated associations separately for each lifestyle factor.

Nevertheless, our study has limitations. Cross-sectional analyses prevent examination of temporal sequences. Reverse causality such as poor health and disability leading to unhealthy lifestyle, less favourable socioeconomic outcomes, fewer social interactions or living in more deprived and polluted areas needs to be considered, except for fixed characteristics such as sex and ethnicity. Associations reported here cannot be interpreted as causal effects, given that several variables included in Model 3 could lie on the causal pathway of other variables. Model 3 might also be over-adjusted. Hence, we have focussed on associations that are consistent across Model 1-3 in the main body of the text and highlighted inconsistencies. As many explanatory variables were self-reported, we cannot exclude the possibility that social desirability might have influenced responses. For example, previous research has shown that up to half of participants who claim to never have had an alcoholic drink reported alcohol intake during previous surveys^71^. For most participants, explanatory variables were measured at a single time point, which could increase the potential for measurement error and did not allow us to model changes across the life course. However, there is evidence that lifestyle is fairly stable in this age group, with few people newly adopting a healthy lifestyle in middle age^72^. There might be some misclassification in the reporting of medical illnesses. However, participants were asked to report illnesses that had been diagnosed by a doctor and these were confirmed during a nurse-led interview.

### Generalisability

The overall response rate of the UK Biobank was low (5.5%) and compared with non-responders, participants were older, more likely to be female and lived in less deprived neighbourhoods. Participants were also less likely to be obese, to smoke, to never drink or to drink alcohol daily and they reported fewer health conditions compared with data from a nationally representative survey^73^. While the UK Biobank states that “valid assessment of exposure-disease relationships are nonetheless widely generalizable and do not require participants to be representative of the population at large”^74^, concordant with the finding that there is little evidence of considerable bias due to non-participation in epidemiological research^75^, the magnitude of exposure-disease associations may depend on prevalence of effect modifiers^76^. A recent empirical investigation comparing the UK Biobank with data from 18 prospective cohort studies with conventional response rates showed that the direction of risk factor associations were similar, although with differences in magnitude^77^.

### Implications

Our findings, while partly confirmatory in nature, shed light on some of the behavioural, psychosocial and environmental pathways to health and are thus relevant to public health policies aimed at promoting health in later life. Public health and medicine could put greater focus on non-medical factors such as loneliness, further encourage healthy lifestyle behaviours and weight management, and examine efforts to improve the health outcomes of individuals with low income. Our findings support the view that promoting individual responsibility for one’s health (e.g. engaging in healthy lifestyle behaviours) and policy-level commitments (e.g. reducing long-term exposure to environmental pollutants^78^ or improving neighbourhoods) both need to be considered in population health. While associations between lifestyle and health might reflect a life-long commitment to healthy behaviours, it is not too late to newly adopt healthy behaviours later in life^72^. Prevention, in addition to drug discovery and disease treatment, should be the top priority for health policy.

## Data Availability

The data used in the present study are available to all bona fide researchers for health-related research that is in the public interest, subject to an application process and approval criteria. Study materials are publicly available online at http://www.ukbiobank.ac.uk.

## Acknowledgments

JM acknowledges current studentship funding from the Biotechnology and Biological Sciences Research Council (BBSRC) (ref: 2050702) and Eli Lilly and Company Limited in support of this work. CML is part-funded by the National Institute for Health Research (NIHR) Biomedical Research Centre at South London and Maudsley NHS Foundation Trust and Kings College London. The views expressed are those of the authors and not necessarily those of the NHS, the NIHR or the Department of Health and Social Care. This project made use of time on Rosalind HPC, funded by Guy’s & St Thomas’ Hospital NHS Trust Biomedical Research Centre (GSTT-BRC), South London & Maudsley NHS Trust Biomedical Research Centre (SLAM-BRC), and Faculty of Natural Mathematics & Science (NMS) at King’s College London.

## Authorship contributions

CML acquired the studentship funding. JM and CML conceived the idea of the study; JM acquired the data; JM carried out the statistical analysis; JM and CML interpreted the findings; JM wrote the manuscript. CJR and CML critically reviewed the manuscript and JM revised the manuscript for final submission. All authors read and approved the final manuscript. JM had full access to all the data in the study and takes responsibility for the integrity of the data and the accuracy of the data analysis.

## Ethics

Ethical approval for the UK Biobank study has been granted by the National Information Governance Board for Health and Social Care and the NHS North West Multicentre Research Ethics Committee (11/NW/0382). No project-specific ethical approval is needed. Data access permission has been granted under UK Biobank application 45514.

## References

1 Partridge, L., Deelen, J. & Slagboom, P. E. Facing up to the global challenges of ageing. Nature 561, 45 (2018).

2 Roser, M. Life expectancy, 2013).

3 Kontis, V. et al. Future life expectancy in 35 industrialised countries: projections with a Bayesian model ensemble. The Lancet 389, 1323–1335 (2017).

4 Netuveli, G., Wiggins, R. D., Hildon, Z., Montgomery, S. M. & Blane, D. Quality of life at older ages: evidence from the English longitudinal study of aging (wave 1). Journal of Epidemiology & Community Health 60, 357–363 (2006).

5 Van Hecke, O., Torrance, N. & Smith, B. Chronic pain epidemiology and its clinical relevance. British journal of anaesthesia 111, 13–18 (2013).

6 Brown, G. C. Living too long. EMBO reports 16, 137–141 (2015).

7 Nybo, H. et al. Functional status and self-rated health in 2,262 nonagenarians: the Danish 1905 Cohort Survey. Journal of the American Geriatrics Society 49, 601–609 (2001).

8 Olshansky, S. J. From Lifespan to Healthspan. JAMA (2018).

9 Carter, B. D. et al. Smoking and mortality—beyond established causes. New England journal of medicine 372, 631–640 (2015).

10 Lai, H. T. et al. Serial circulating omega 3 polyunsaturated fatty acids and healthy ageing among older adults in the Cardiovascular Health Study: prospective cohort study. BMJ 363, k4067 (2018).

11 Room, R., Babor, T. & Rehm, J. Alcohol and public health. The lancet 365, 519–530 (2005).

12 Cooper, R., Kuh, D. & Hardy, R. Objectively measured physical capability levels and mortality: systematic review and meta-analysis. Bmj 341, c4467 (2010).

13 Loef, M. & Walach, H. The combined effects of healthy lifestyle behaviors on all cause mortality: a systematic review and meta-analysis. Preventive medicine 55, 163–170 (2012).

14 Holt-Lunstad, J., Smith, T. B., Baker, M., Harris, T. & Stephenson, D. Loneliness and social isolation as risk factors for mortality: a meta-analytic review. Perspectives on Psychological Science 10, 227–237 (2015).

15 Smith, R. B. et al. Impact of London’s road traffic air and noise pollution on birth weight: retrospective population based cohort study. bmj 359, j5299 (2017).

16 Bycroft, C. et al. The UK Biobank resource with deep phenotyping and genomic data. Nature 562, 203 (2018).

17 Guggenheim, J. A. & Williams, C. Childhood febrile illness and the risk of myopia in UK Biobank participants. Eye 30, 608 (2016).

18 The English indices of deprivation 2010. (2011).

19 Elovainio, M. et al. Contribution of risk factors to excess mortality in isolated and lonely individuals: an analysis of data from the UK Biobank cohort study. The Lancet Public Health 2, e260–e266 (2017).

20 IPAQ-Group. IPAQ scoring protocol - International Physical Activity Questionnaire, <https://sites.google.com/site/theipaq/scoring-protocol> (

21 Beelen, R. et al. Development of NO2 and NOx land use regression models for estimating air pollution exposure in 36 study areas in Europe–The ESCAPE project. Atmospheric Environment 72, 10–23 (2013).

22 Eeftens, M. et al. Development of land use regression models for PM2. 5, PM2. 5 absorbance, PM10 and PMcoarse in 20 European study areas; results of the ESCAPE project. Environmental science & technology 46, 11195–11205 (2012).

23 Kephalopoulos, S. et al. Advances in the development of common noise assessment methods in Europe: The CNOSSOS-EU framework for strategic environmental noise mapping. Science of the Total Environment 482, 400–410 (2014).

24 Morley, D. et al. International scale implementation of the CNOSSOS-EU road traffic noise prediction model for epidemiological studies. Environmental pollution 206, 332–341 (2015).

25 Vienneau, D., Schindler, C., Perez, L., Probst-Hensch, N. & Röösli, M. The relationship between transportation noise exposure and ischemic heart disease: a meta-analysis. Environmental research 138, 372–380 (2015).

26 Department-for-Communities-and-Local-Government. (https://data.gov.uk/dataset/land_use_statistics_generalised_land_use_database, 2007).

27 Maxwell, J. M. et al. Multifactorial disorders and polygenic risk scores: predicting common diseases and the possibility of adverse selection in life and protection insurance. Annals of Actuarial Science, 1–16 (2020).

28 Fell, D. B., Joseph, K., Armson, B. A. & Dodds, L. The impact of pregnancy on physical activity level. Maternal and child health journal 13, 597 (2009).

29 Benjamini, Y. & Hochberg, Y. Controlling the false discovery rate: a practical and powerful approach to multiple testing. Journal of the Royal statistical society: series B (Methodological) 57, 289–300 (1995).

30 Fox, J. & Monette, G. Generalized collinearity diagnostics. Journal of the American Statistical Association 87, 178–183 (1992).

31 Gelman, A. Scaling regression inputs by dividing by two standard deviations. Statistics in medicine 27, 2865–2873 (2008).

32 Cappuccio, F. P., D’Elia, L., Strazzullo, P. & Miller, M. A. Sleep duration and all-cause mortality: a systematic review and meta-analysis of prospective studies. Sleep 33, 585–592 (2010).

33 Cassidy, S., Chau, J. Y., Catt, M., Bauman, A. & Trenell, M. I. Cross-sectional study of diet, physical activity, television viewing and sleep duration in 233 110 adults from the UK Biobank; the behavioural phenotype of cardiovascular disease and type 2 diabetes. BMJ open 6, e010038 (2016).

34 WorldHealthOrganization. Air quality guidelines: global update 2005: particulate matter, ozone, nitrogen dioxide, and sulfur dioxide. (World Health Organization, 2006).

35 Mayor, S. (British Medical Journal Publishing Group, 2018).

36 Cockerham, W. C., Sharp, K. & Wilcox, J. A. Aging and perceived health status. Journal of Gerontology 38, 349–355 (1983).

37 Ferraro, K. F. Self-ratings of health among the old and the old-old. Journal of Health and Social Behavior, 377–383 (1980).

38 Wu, S. et al. The relationship between self-rated health and objective health status: a population-based study. BMC public health 13, 320 (2013).

39 Kelleher, C., Friel, S., Gabhainn, S. N. & Tay, J. B. Socio-demographic predictors of selfrated health in the Republic of Ireland: findings from the National Survey on Lifestyle, Attitudes and Nutrition, SLAN. Social science & medicine 57, 477–486 (2003).

40 Sinclair, D. A. & LaPlante, M. D. Lifespan: Why We Age—and Why We Don’t Have To. (Atria Books, 2019).

41 Brouwer, W. B., van Exel, N. J. A. & Stolk, E. A. Acceptability of less than perfect health states. Social Science & Medicine 60, 237–246 (2005).

42 Collerton, J. et al. Health and disease in 85 year olds: baseline findings from the Newcastle 85+ cohort study. Bmj 339, b4904 (2009).

43 Gorman, B. K. & Read, J. n. G. Gender disparities in adult health: an examination of three measures of morbidity. Journal of health and social behavior 47, 95–110 (2006).

44 McDonough, P. & Walters, V. Gender and health: reassessing patterns and explanations. Social science & medicine 52, 547–559 (2001).

45 Austad, S. N. & Bartke, A. Sex differences in longevity and in responses to anti-aging interventions: a mini-review. Gerontology 62, 40–46 (2016).

46 Phaswana-Mafuya, N. et al. Self-rated health and associated factors among older South Africans: evidence from the study on global ageing and adult health. Global Health Action 6, 19880 (2013).

47 Benyamini, Y., Blumstein, T., Lusky, A. & Modan, B. Gender differences in the self-rated health–mortality association: Is it poor self-rated health that predicts mortality or excellent self-rated health that predicts survival? The gerontologist 43, 396–405 (2003).

48 Singh, L., Arokiasamy, P., Singh, P. K. & Rai, R. K. Determinants of gender differences in self-rated health among older population: evidence from India. Sage Open 3, 2158244013487914 (2013).

49 Jylhä, M., Guralnik, J. M., Ferrucci, L., Jokela, J. & Heikkinen, E. Is self-rated health comparable across cultures and genders? The Journals of Gerontology Series B: Psychological Sciences and Social Sciences 53, S144–S152 (1998).

50 Gold, C. H., Malmberg, B., McClearn, G. E., Pedersen, N. L. & Berg, S. Gender and health: a study of older unlike-sex twins. The Journals of Gerontology Series B: Psychological Sciences and Social Sciences 57, S168–S176 (2002).

51 Shibuya, K., Hashimoto, H. & Yano, E. Individual income, income distribution, and self rated health in Japan: cross sectional analysis of nationally representative sample. Bmj 324, 16 (2002).

52 Der, G., Macintyre, S., Ford, G., Hunt, K. & West, P. The relationship of household income to a range of health measures in three age cohorts from the West of Scotland. The European Journal of Public Health 9, 271–277 (1999).

53 Godhwani, S., Jivraj, S., Marshall, A. & Bécares, L. Comparing subjective and objective neighbourhood deprivation and their association with health over time among older adults in England. Health & place 55, 51–58 (2019).

54 Rehkopf, D. H., Berkman, L. F., Coull, B. & Krieger, N. The non-linear risk of mortality by income level in a healthy population: US National Health and Nutrition Examination Survey mortality follow-up cohort, 1988–2001. BMC Public Health 8, 383 (2008).

55 Davies, N. M., Dickson, M., Smith, G. D., Van Den Berg, G. J. & Windmeijer, F. The causal effects of education on health outcomes in the UK Biobank. Nature human behaviour 2, 117 (2018).

56 Ding, D., Rogers, K., van der Ploeg, H., Stamatakis, E. & Bauman, A. E. Traditional and emerging lifestyle risk behaviors and all-cause mortality in middle-aged and older adults: evidence from a large population-based Australian cohort. PLoS medicine 12 (2015).

57 Coyle, C. E. & Dugan, E. Social isolation, loneliness and health among older adults. Journal of aging and health 24, 1346–1363 (2012).

58 Perissinotto, C. M., Cenzer, I. S. & Covinsky, K. E. Loneliness in older persons: a predictor of functional decline and death. Archives of internal medicine 172, 1078–1084 (2012).

59 Warburton, D. E., Nicol, C. W. & Bredin, S. S. Health benefits of physical activity: the evidence. Cmaj 174, 801–809 (2006).

60 Arem, H. et al. Leisure time physical activity and mortality: a detailed pooled analysis of the dose-response relationship. JAMA internal medicine 175, 959–967 (2015).

61 Yu, S., Yarnell, J., Sweetnam, P. & Murray, L. What level of physical activity protects against premature cardiovascular death? The Caerphilly study. Heart 89, 502–506 (2003).

62 Lee, I., Paffenbarger, R. & Hennekens, C. Physical activity, physical fitness and longevity. Aging Clinical and Experimental Research 9, 2–11 (1997).

63 Oguma, Y., Sesso, H., Paffenbarger, R. & Lee, I. Physical activity and all cause mortality in women: a review of the evidence. British journal of sports medicine 36, 162–172 (2002).

64 Lee, I.-M. & Paffenbarger Jr, R S. Associations of light, moderate, and vigorous intensity physical activity with longevity: the Harvard Alumni Health Study. American journal of epidemiology 151, 293–299 (2000).

65 Piazza-Gardner, A. K. & Barry, A. E. Examining physical activity levels and alcohol consumption: are people who drink more active? American Journal of Health Promotion 26, e95–e104 (2012).

66 Shaper, A. G., Wannamethee, G. & Walker, M. Alcohol and mortality in British men: explaining the U-shaped curve. The Lancet 332, 1267–1273 (1988).

67 Stockwell, T. et al. Do “moderate” drinkers have reduced mortality risk? A systematic review and meta-analysis of alcohol consumption and all-cause mortality. Journal of studies on alcohol and drugs 77, 185–198 (2016).

68 Fat, L. N., Cable, N., Marmot, M. & Shelton, N. Persistent long-standing illness and non-drinking over time, implications for the use of lifetime abstainers as a control group. J Epidemiol Community Health 68, 71–77 (2014).

69 Cai, Y. et al. Road traffic noise, air pollution and incident cardiovascular disease: a joint analysis of the HUNT, EPIC-Oxford and UK Biobank cohorts. Environment international 114, 191–201 (2018).

70 Twohig-Bennett, C. & Jones, A. The health benefits of the great outdoors: A systematic review and meta-analysis of greenspace exposure and health outcomes. Environmental research 166, 628–637 (2018).

71 Rehm, J. et al. Are lifetime abstainers the best control group in alcohol epidemiology? On the stability and validity of reported lifetime abstention. American journal of epidemiology 168, 866–871 (2008).

72 King, D. E., Mainous III, A.G. & Geesey, M. E. Turning back the clock: adopting a healthy lifestyle in middle age. The American journal of medicine 120, 598–603 (2007).

73 Fry, A. et al. Comparison of sociodemographic and health-related characteristics of UK Biobank participants with those of the general population. American Journal of Epidemiology 186, 1026–1034 (2017).

74 UK-Biobank. http://www.ukbiobank.ac.uk/wp-content/uploads/2017/03/access-matters-representativeness-1.pdf, <http://www.ukbiobank.ac.uk/wp-content/uploads/2017/03/access-matters-representativeness-1.pdf> (

75 Galea, S. & Tracy, M. Participation rates in epidemiologic studies. Annals of epidemiology 17, 643–653 (2007).

76 Keyes, K. M. & Westreich, D. UK Biobank, big data, and the consequences of non-representativeness. The Lancet 393, 1297 (2019).

77 Battya, G. D., Galec, C. R., Kivimäkia, M., Dearyd, I. J. & Belle, S. Generalisability of Results from UK Biobank: Comparison With a Pooling of 18 Cohort Studies.

78 Holgate, S. T. ‘Every breath we take: the lifelong impact of air pollution’–a call for action. Clinical Medicine 17, 8–12 (2017).

