## Supplement for "Exploring health in the UK Biobank: associations with sociodemographic characteristics, psychosocial factors, lifestyle and environmental exposures"

### Table of contents


**SUPPLEMENTARY TEXT** 4
**SUPPLEMENT E1. DATA FIELDS** 5
**SUPPLEMENT E2. STUDY FLOWCHART** 6
**SUPPLEMENT E3. BASELINE CHARACTERISTICS** 6
**SUPPLEMENT E4. SELF-REPORTED ILLNESSES** 9
Cancer illnesses 9
Non-cancer illnesses 10
**SUPPLEMENT E5. CORRELATION MATRIX CONTINUOUS VARIABLES** 11
**SUPPLEMENT E6. DESCRIPTIVE STATISTICS STRATIFIED BY HEALTH INDICATORS** 12
Stratified by health status 12
Stratified by long-standing illness 13
Baseline characteristics stratified by self-rated health 14
**SUPPLEMENT E7. VISUAL SUMMARY OF FINDINGS** 16
Lifestyle factors 16
Environmental exposures 17
Psychosocial factors 18
Sociodemographic characteristics 19
**SUPPLEMENT E8. REGRESSION TABLES LONG-STANDING ILLNESS** 20
Sociodemographic characteristics 20
Psychosocial factors 21
Lifestyle factors 22
Environmental exposures 23
**SUPPLEMENT E9. REGRESSION TABLES SELF-RATED HEALTH** 24
Sociodemographic characteristics 24
Psychosocial factors 25
Lifestyle factors 26
Environmental exposures 27
**SUPPLEMENT E10. REGRESSION TABLES HEALTH INDICATORS** 28
Sociodemographic characteristics 28
Psychosocial factors 29
Lifestyle factors 30
Environmental exposures 31
**SUPPLEMENT E11. STANDARDISED REGRESSION COEFFICIENTS - TABLES** 32
Health status 32
Long-standing illness 33
Self-rated health 34
**SUPPLEMENT E12. STANDARDISED REGRESSION COEFFICIENTS – PLOTS** 35
Long-standing illness 35
Self-rated health 36
**SUPPLEMENT E13. BASELINE CHARACTERISTICS STRATIFIED BY SEX** 37
**SUPPLEMENT E14. BASELINE CHARACTERISTICS STRATIFIED BY AGE** 39
**SUPPLEMENT E15. REGRESSION TABLES HEALTH STATUS STRATIFIED BY SEX** 41
Sociodemographic characteristics 41
Psychosocial factors 42
Lifestyle factors 43
Environmental exposures 44
**SUPPLEMENT E16. CONFIDENCE INTERVAL PLOTS HEALTH STATUS STRATIFIED BY SEX** 45
Sociodemographic and psychosocial factors 45
Lifestyle factors 46
Environmental exposures 47
**SUPPLEMENT E17. REGRESSION TABLES HEALTH STATUS STRATIFIED BY AGE** 48
Sociodemographic characteristics 48
Psychosocial factors 49
Lifestyle factors 50
Environmental exposures 51
**SUPPLEMENT E18. CONFIDENCE INTERVAL PLOTS HEALTH STATUS STRATIFIED BY AGE** 52
Sociodemographic and psychosocial factors 52
Lifestyle factors 53
Environmental exposures 54
**SUPPLEMENT E19. REGRESSION TABLES LONG-STANDING ILLNESS STRATIFIED BY SEX** 55
Sociodemographic characteristics 55
Psychosocial factors 56
Lifestyle factors 57
Environmental exposures 58
**SUPPLEMENT E20. CONFIDENCE INTERVAL PLOTS LONG-STANDING ILLNESS STRATIFIED BY SEX** 59
Sociodemographic and psychosocial factors 59
Lifestyle factors 60
Environmental exposures 61
**SUPPLEMENT E21. REGRESSION TABLES LONG-STANDING ILLNESS STRATIFIED BY AGE** 62
Sociodemographic characteristics 62
Psychosocial factors 63
Lifestyle factors 64
Environmental exposures 65

**SUPPLEMENT E22. CONFIDENCE INTERVAL PLOTS LONG-STANDING ILLNESS STRATIFIED BY AGE ..... 66**
SOCIODEMOGRAPHIC AND PSYCHOSOCIAL FACTORS ..... 66
LIFESTYLE FACTORS ..... 67
ENVIRONMENTAL EXPOSURES ..... 68
**SUPPLEMENT E23. REGRESSION TABLES SELF-RATED HEALTH STRATIFIED BY SEX ..... 69**
SOCIODEMOGRAPHIC CHARACTERISTICS ..... 69
PSYCHOSOCIAL FACTORS ..... 70
LIFESTYLE FACTORS ..... 71
ENVIRONMENTAL EXPOSURES ..... 72
**SUPPLEMENT E24. CONFIDENCE INTERVAL PLOTS SELF-RATED HEALTH STRATIFIED BY SEX ..... 73**
SOCIODEMOGRAPHIC AND PSYCHOSOCIAL FACTORS ..... 73
LIFESTYLE FACTORS ..... 74
ENVIRONMENTAL EXPOSURES ..... 75
**SUPPLEMENT E25. REGRESSION TABLES SELF-RATED HEALTH STRATIFIED BY AGE ..... 76**
SOCIODEMOGRAPHIC CHARACTERISTICS ..... 76
PSYCHOSOCIAL FACTORS ..... 77
LIFESTYLE FACTORS ..... 78
ENVIRONMENTAL EXPOSURES ..... 79
**SUPPLEMENT E26. CONFIDENCE INTERVAL PLOTS SELF-RATED HEALTH STRATIFIED BY AGE ..... 80**
SOCIODEMOGRAPHIC AND PSYCHOSOCIAL FACTORS ..... 80
LIFESTYLE FACTORS ..... 81
ENVIRONMENTAL EXPOSURES ..... 82
**SUPPLEMENT E27. BASELINE CHARACTERISTICS PROSPECTIVE SAMPLES ..... 83**
**SUPPLEMENT E28. REGRESSION TABLES SELF-RATED HEALTH T1 ..... 85**
SOCIODEMOGRAPHIC CHARACTERISTICS ..... 85
PSYCHOSOCIAL FACTORS ..... 86
LIFESTYLE FACTORS ..... 87
ENVIRONMENTAL EXPOSURES ..... 88
**SUPPLEMENT E29. REGRESSION TABLES SELF-RATED HEALTH T2 ..... 89**
SOCIODEMOGRAPHIC CHARACTERISTICS ..... 89
PSYCHOSOCIAL FACTORS ..... 90
LIFESTYLE FACTORS ..... 91
ENVIRONMENTAL EXPOSURES ..... 92
**SUPPLEMENT E30. ADDITIONAL ANALYSES ..... 93**
DESCRIPTIVE STATISTICS ..... 93
**SUPPLEMENT E31. FITTED PROBABILITIES ..... 94**
SOCIODEMOGRAPHIC CHARACTERISTICS ..... 94
PSYCHOSOCIAL FACTORS ..... 96
LIFESTYLE FACTORS ..... 97
ENVIRONMENTAL EXPOSURES ..... 99

**Supplementary text**

**Summary description of results for long-standing illness and self-rated health**

The full results are presented in Supplement-e8–e10.

Findings regarding income, sex and neighbourhood deprivation were mostly consistent across health indicators, although with some variation in the magnitude of associations. Compared to Whites, participants of Chinese ethnicity had lower odds of rating their health favourable (OR = 0.68, 99.9% CI 0.541-0.851, $p_{Bonf.} < 0.001$) but had higher odds of being classified healthy (OR = 1.83, 99.9% CI 1.365-2.511, $p_{Bonf.} < 0.001$) and being free from long-standing illness (OR = 1.62, 99.9% CI 1.221-2.184, $p_{Bonf.} < 0.001$). Individuals of non-White ethnic backgrounds tended to rate their health less favourable although there was some evidence that they had higher odds of being free from long-standing illness, especially after adjustment for covariates. Individuals with any qualification had higher odds of rating their health more favourable (e.g. OR = 1.50, 99.9% CI 1.441-1.568, $p_{Bonf.} < 0.001$ [university/college degree vs no qualification]), while there was no consistent pattern for health status and long-standing illness. Finally, older individuals tended to rate their health more favourable (OR = 1.011, 99.9% CI 1.010-1.013, $p_{Bonf.} < 0.001$).

Findings regarding psychosocial factors were consistent across health indicators, but the association was strongest for loneliness and self-rated health (OR = 0.49, 99.9% CI 0.468-0.518, $p_{Bonf.} < 0.001$).

Findings regarding lifestyle factors were consistent across health indicators, except that there were some inconsistencies in the associations between daily alcohol intake and health and that more frequent moderate physical activity was associated with better self-rated health also in Model 3 (OR = 1.01, 99.9% CI 1.007-1.019, $p_{Bonf.} < 0.001$). Longer sleep duration was also associated with better self-rated health (OR = 1.07, 99.9% CI 1.061-1.085, $p_{Bonf.} < 0.001$) but there was little evidence of an association with long-standing illness.

Higher levels of $PM_{2.5}$ were also associated with lower odds of not having a long-standing illness (OR = 0.97, 99.9% CI 0.943-0.993, $p_{Bonf.} = 0.002$). While we found similar associations with self-rated health in Model 1–2, $PM_{2.5}$ was associated with more favourable self-rated health in Model 3 (OR = 1.03, 99.9% CI 1.005-1.052, $p_{Bonf.} = 0.004$). There was no evidence of an association between $PM_{10}$ and any health indicator in Model 3, although $PM_{10}$ was associated with poor health across outcomes in Model 1–2. Higher $NO_2$ concentration was associated with less favourable self-rated health (OR = 0.99, 99.9% CI 0.991-0.999, $p_{Bonf.} < 0.001$), while there was no evidence of an association with health status and long-standing illness after adjusting for covariates.

Higher $L_{den}$ was mostly associated with poor health indicators in Model 1–2, but we found no evidence of an association in Model 3. Percentage greenspace was associated with better self-rated health and higher odds of being free from long-standing illness in Model 1–2, but we found only limited evidence of an association in Model 3 (but OR = 1.0007, 99.9% CI 0.998-1.0015, $p_{BH} = 0.015$ for self-rated health).

**Supplement e1. Data fields**

Supplement e1. **Data fields used in the present study.**

| UK Biobank data field | Variable name |
|---|---|
| **Sociodemographic characteristics** | |
| 31 | Sex |
| 738 | Average total household income before tax |
| 6138[1] | Qualifications |
| 21000[1] | Ethnic background |
| 21003 | Age when attended assessment centre |
| 22001 | Genetic sex |
| 26410 | Index of Multiple Deprivation (England) |
| **Psychosocial factors** | |
| 709[2] | Number in household |
| 1031[2] | Frequency of friend/family visits |
| 6160[2] | Leisure/social activities |
| 2020[3] | Loneliness, isolation |
| 2110[3] | Able to confide |
| **Lifestyle factors** | |
| 864 | Number of days per week walked 10+ minutes |
| 884 | Number of days per week of moderate physical activity 10+ minutes |
| 904 | Number of days per week of vigorous physical activity 10+ minutes |
| 943 | Frequency of stair climbing in last 4 weeks |
| 1160 | Sleep duration |
| 1239 | Current tobacco smoking |
| 1558 | Alcohol intake frequency |
| 3731 | Former alcohol drinker |
| 3859 | Reason former drinker stopped drinking alcohol |
| 20116 | Smoking status |
| 22037 | MET minutes per week for walking |
| 22038 | MET minutes per week for moderate activity |
| 22039 | MET minutes per week for vigorous activity |
| 23099 | Body fat percentage |
| 23104 | Body mass index (BMI) |
| **Environmental exposures** | |
| 24003 | Nitrogen dioxide air pollution; 2010 |
| 24005 | Particulate matter air pollution (pm10); 2010 |
| 24006 | Particulate matter air pollution (pm2.5); 2010 |
| 24024 | Average 24-hour sound level of noise pollution |
| 24500 | Greenspace percentage, buffer 1000m |
| 24503 | Greenspace percentage, buffer 300m |
| **Health indicators** | |
| 2178 | Overall health rating |
| 2188 | Long-standing illness, disability of infirmity |
| 20001 | Cancer code, self-reported |
| 20002 | Non-cancer illness code, self-reported |
| **Other** | |
| 53 | Date of attending assessment centre |
| 699 | Length of time at current address |
| 3140 | Pregnant |

*Note:* [1] these variables have been further processed and categories used in the present study are described in the main body of the text. [2] variables used to derive social isolation index. [3] variables used to derive loneliness index.

**Loneliness index:**

1. If the response to one question was missing, “don’t know” or “prefer not to answer” and the response to the second question was scored 0, individuals were classified as not lonely.
2. If the response to one question was missing, “don’t know” or “prefer not to answer” and the response to the second question was scored 1, individuals were classified as missing data on loneliness.
3. If responses to both questions were missing, “don’t know” or “prefer not to answer”, individuals were classified as missing data on loneliness.

**Social isolation index:**

1. If the response to two questions were scored 1 and the response to the third question was missing, “don’t know” or “prefer not to answer”, individuals were classified as socially isolated.
2. If two or more responses to questions were missing, “don’t know” or “prefer not to answer”, individuals were classified as missing data on social isolation.
3. If the response to two questions were scored 0 and the response to the third question was missing, “don’t know” or “prefer not to answer”, individuals were classified as not socially isolated.
4. If the response to one question was scored 0, the response to one question was scored 1 and the response to the third question was missing, “don’t know” or “prefer not to answer”, individuals were classified as missing data on social isolation.

**Health status:**

Individuals who reported one 'unclassifiable' non-cancer illness and no cancer illness may be classified healthy or unhealthy. To take the most conservative approach, we classified these as 'missing data'. Any 'unclassifiable' cancer was assumed to be unhealthy. A complete list of the non-cancer illness and cancer classification is provided in a separate supplementary file.

**Supplement e2. Study flowchart**

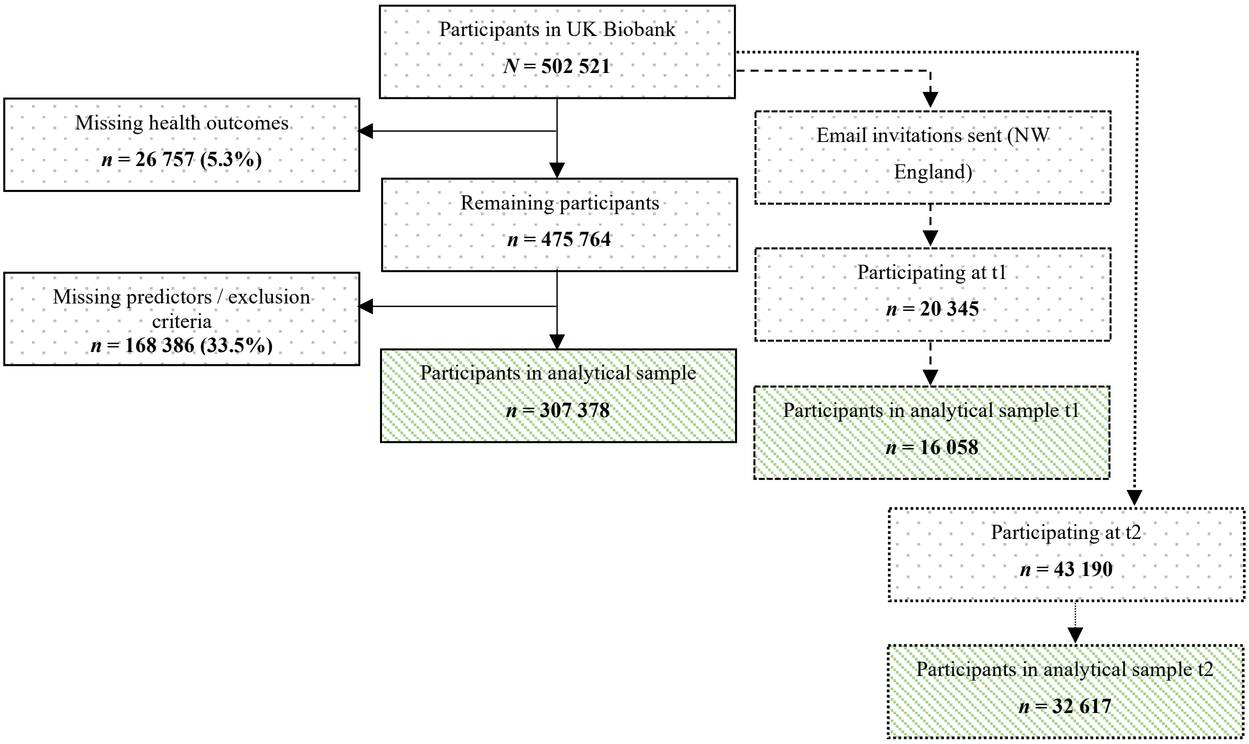


**Supplement e2.** Flowchart of study population. t1 = follow-up during first revisit between 2012–2013; t2 = follow-up during UK Biobank Imaging Study between 2014-2019.

**Supplement e3. Baseline characteristics**

Supplement e3. **Baseline characteristics**

| | Full sample (*N* = 502 521) | Analytical sample (*n* = 307 378) |
|---|---|---|
| Health indicators | | |
| **Health status** | | |
| Healthy | 162 096 (32.3%) | 95 177 (31.0%) |
| Unhealthy | 328 148 (65.3%) | 212 201 (69.0%) |
| Missing | 12 277 (2.4%) | |
| **Long-standing illness** | | |
| Yes | 159 904 (31.8%) | 93 757 (30.5%) |
| No | 329 260 (65.5%) | 213 621 (69.5%) |
| Prefer not to answer | 1052 (0.2%) | |
| Do not know | 11 387 (2.3%) | |
| Missing | 918 (0.2%) | |
| **Self-rated health** | | |
| Poor | 22 778 (4.5%) | 11 066 (3.6%) |
| Fair | 105 373 (21.0%) | 59 169 (19.2%) |
| Good | 289 023 (57.5%) | 182 699 (59.4%) |
| Excellent | 81 860 (16.3%) | 54 444 (17.7%) |
| Prefer not to answer | 365 (0.1%) | |
| Do not know | 2 204 (0.4%) | |
| Missing | 918 (0.2%) | |
| Sociodemographic characteristics | | |
| **Age** | | |
| Mean (SD) | 56.53 (8.10) | 56.11 (8.01) |
| Range | 37-73 | 38-73 |
| **Sex** | | |
| Female | 273 394 (54.4%) | 159 574 (51.9%) |
| Male | 229 127 (45.6%) | 147 804 (48.1%) |
| **Ethnicity** | | |
| White | 472 711 (94.1%) | 293 565 (95.5%) |
| Mixed-race | 2 958 (0.6%) | 1 766 (0.6%) |
| Black | 8 061 (1.6%) | 4 257 (1.4%) |
| Asian | 9 882 (2.0%) | 4 755 (1.5%) |
| Chinese | 1 574 (0.3%) | 818 (0.3%) |
| Other | 4 558 (0.9%) | 2 217 (0.7%) |

| | | |
|---|---|---|
| Prefer not to answer | 1 662 (0.3%) | |
| Do not know | 217 (0.0%) | |
| Missing | 898 (0.2%) | |
| **Highest qualification** | | |
| None | 85 274 (17.0%) | 39 828 (13.0%) |
| O levels/GCSEs/CSEs | 132 086 (26.3%) | 84 448 (27.5%) |
| A levels/NVQ/HND/HNC[1] | 113 859 (22.7%) | 72 584 (23.6%) |
| Degree | 161 168 (32.1%) | 110 518 (36.0%) |
| Prefer not to answer | 5 493 (1.1%) | |
| Missing | 4 641 (0.9%) | |
| **Household income**[2] | | |
| Very low | 97 205 (19.3%) | 63 099 (20.5%) |
| Low | 108 177 (21.5%) | 77 931 (25.4%) |
| Medium | 110 774 (22.0%) | 82 338 (26.8%) |
| High | 86 269 (17.2%) | 66 106 (21.5%) |
| Very high | 22 930 (4.6%) | 17 904 (5.8%) |
| Prefer not to answer | 49 848 (9.9%) | |
| Do not know | 21 305 (4.2%) | |
| Missing | 6 013 (1.2%) | |
| **Multiple deprivation** | | |
| Mean (SD) | 17.68 (14.01) | 16.77 (13.33) |
| Range | 0.61-82 | 0.61-82 |
| Missing | 69 774 (13.1%) | |
| Psychosocial factors | | |
| **Loneliness** | | |
| Not lonely | 466 182 (92.8%) | 289 901 (94.3%) |
| Lonely | 30 395 (6.0%) | 17 477 (5.7%) |
| Missing | 5 944 (1.2%) | |
| **Social isolation** | | |
| Not isolated | 453 401 (90.2%) | 280 931 (91.4%) |
| Isolated | 46 143 (9.2%) | 26 447 (8.6%) |
| Missing | 2 977 (0.6%) | |
| Lifestyle factors | | |
| **Smoking status** | | |
| Never | 273 528 (54.4%) | 168 475 (54.8%) |
| Former | 173 064 (34.4%) | 108 638 (35.3%) |
| Current | 52 979 (10.5%) | 30 265 (9.8%) |
| Prefer not to answer | 2 059 (0.4%) | |
| Missing | 891 (0.2%) | |
| **Stair climbing frequency** | | |
| None | 44 988 (9.0%) | 24 049 (7.8%) |
| 1-5/day | 100 569 (20.0%) | 58 267 (19.0%) |
| 6-10/day | 178 969 (35.6%) | 115 982 (37.7%) |
| 11-15/day | 91 352 (18.2%) | 60 315 (19.6%) |
| 16-20/day | 42 477 (8.5%) | 27 609 (9.0%) |
| 20+/day | 34 566 (6.9%) | 21 156 (6.9%) |
| Prefer not to answer | 461 (0.1%) | |
| Do not know | 2 580 (0.5%) | |
| Missing | 6 559 (1.3%) | |
| **Alcohol intake frequency** | | |
| Never | 40 645 (8.1%) | 20 423 (6.6%) |
| Special occasions | 58 011 (11.5%) | 31 526 (10.3%) |
| 1-3/month | 55 856 (11.1%) | 33 798 (11.0%) |
| 1-2/week | 129 294 (25.7%) | 78 777 (25.6%) |
| 3-4/week | 115443 (23.0%) | 75 251 (24.5%) |
| Daily/almost daily | 10 770 (20.3%) | 67 603 (22.0%) |
| Prefer not to answer | 605 (0.1%) | |
| Missing | 897 (0.2%) | |
| **Sleep duration** (hours/day) | | |
| Mean (SD) | 7.15 (1.11)* | 7.16 (1.06) |
| Range | 1-23* | 1-20 |
| Prefer not to answer | 386 (0.1%) | |
| Do not know | 2943 (0.6%) | |
| Missing | 887 (0.2%) | |
| **BMI** ($kg/m^2$) | | |
| Mean (SD) | 27.43 (4.79) | 27.27 (4.67) |
| Range | 12.80-68.40 | 12.80-67.30 |
| Missing | 10 136 (2.0%) | |
| **Walking** (days/week)[3] | | |
| Mean (SD) | 5.39 (1. 93)§ | 5.36 (1.95) |
| Range | 0-7§ | 0-7 |
| Prefer not to answer | 979 (0.2%) | |
| Do not know | 6 687 (1.3%) | |
| Unable to walk | 1 929 (0.4%) | |
| Missing | 874 (0.2%) | |

| | | |
|---|---|---|
| **Moderate activity** (days/week)[3] | | |
| Mean (SD) | 3.63 (2.33)[±] | 3.59 (2.32) |
| Range | 0-7[±] | 0-7 |
| Prefer not to answer | 2 273 (0.5%) | |
| Do not know | 24 120 (4.8%) | |
| Missing | 878 (0.2%) | |
| **Vigorous activity** (days/week)[3] | | |
| Mean (SD) | 1.84 (1.96)[$] | 1.876 (1.94) |
| Range | 0-7[$] | 0-7 |
| Prefer not to answer | 4 116 (0.8%) | |
| Do not know | 22 582 (4.5%) | |
| Missing | 878 (0.2%) | |
| Environmental exposures | | |
| **$PM_{2.5}$** | | |
| Mean (SD) | 9.99 (1.06) | 9.95 (1.04) |
| Range | 8.17-21.31 | 8.17-21.25 |
| Missing | 41 304 (8.2%) | |
| **$PM_{10}$** | | |
| Mean (SD) | 16.24 (1. 90) | 16.19 (1.88) |
| Range | 11.78-31.39 | 11.78-30.65 |
| Missing | 41 304 (8.2%) | |
| **$NO_2$** | | |
| Mean (SD) | 26.71 (7.58) | 26.44 (7.56) |
| Range | 12.93-108.49 | 12.93-108.49 |
| Missing | 7 381 (1.5%) | |
| **$L_{den}$** | | |
| Mean (SD) | 56.06 (4.28) | 56.01 (4.24) |
| Range | 51.54-93.36 | 51.55-89.29 |
| Missing | 7 381 (1.5%) | |
| **Greenspace** 1000m | | |
| Mean (SD) | 44.99 (21.62) | 45.52 (21.77) |
| Range | 4.42-99.19 | 4.49-99.19 |
| Missing | 61 631 (12.3%) | |

*Note:* GCSEs = general certificate of secondary education; CSE = certificate of secondary education; NVQ = national vocational qualification; HND = higher national diploma; HNC = higher national certificate; BMI = body mass index; PM = particulate matter; $NO_2$ = nitrogen dioxide; $L_{den}$ = day-evening-night noise level. [1]also includes 'other professional qualifications'. [2]Annual household income groups: very low (<£18 000), low (£18 000–30 999), middle (£31 000–51 999), high (£52 000–100 000) and very high (>£100 000). [3]number of days per week engaging in these activities for 10+ minutes continuously. [*]*n*=498 305; [§]*n*=492 052; [±]*n*=475 250; [$]*n*=474 945.

**Additional information**

Descriptive statistic in manuscript on duration living at current address does not include participants who lived at their current address for <1 year (*n* = 4,781), but these individuals were included in all other analyses. The average number of days between the baseline assessment and the air pollution exposure estimate (calculated based on the midpoint between 26 January 2010 and 18 January 2011) was 491 days (SD = 285; range = -56 to 1312). The corresponding figures for traffic noise were 105 days (SD = 285; range = -442 to 926).

**Supplement e4. Self-reported illnesses**

Cancer illnesses

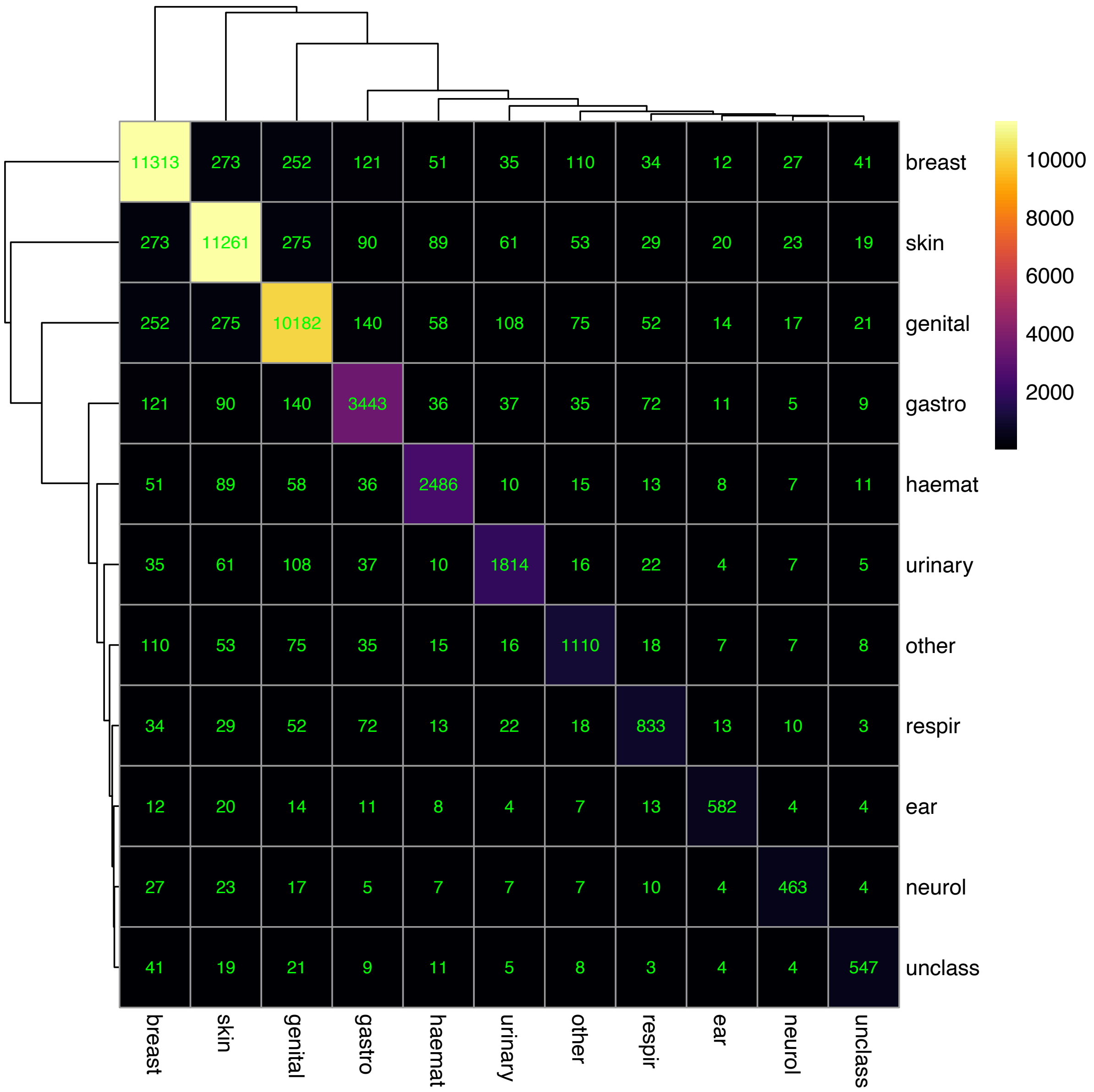


**Supplement e4-A.** Self-reported cancer illnesses. Cell entries reflect the number of individuals with ≥1 cancer illness of the group in the corresponding row and ≥1 cancer illness of the group in the corresponding column (e.g. 273 participants report ≥1 breast cancer and ≥1 skin cancer). Cell entries in the diagonal reflect the number of participants with ≥1 cancer illness of the group in the corresponding row or column (e.g. 11 313 participants report ≥1 breast cancer). breast = breast cancer; skin = skin cancer; genital = genital tract cancer; gastro = gastrointestinal cancer; haemat = haematological malignancy; urinary = urinary tract cancer; other = other cancer; respir = respiratory / intrathoracic cancer; ear = ear/nose/throat cancer; neurol = neurological system cancer; unclass = unclassifiable.

Non-cancer illnesses

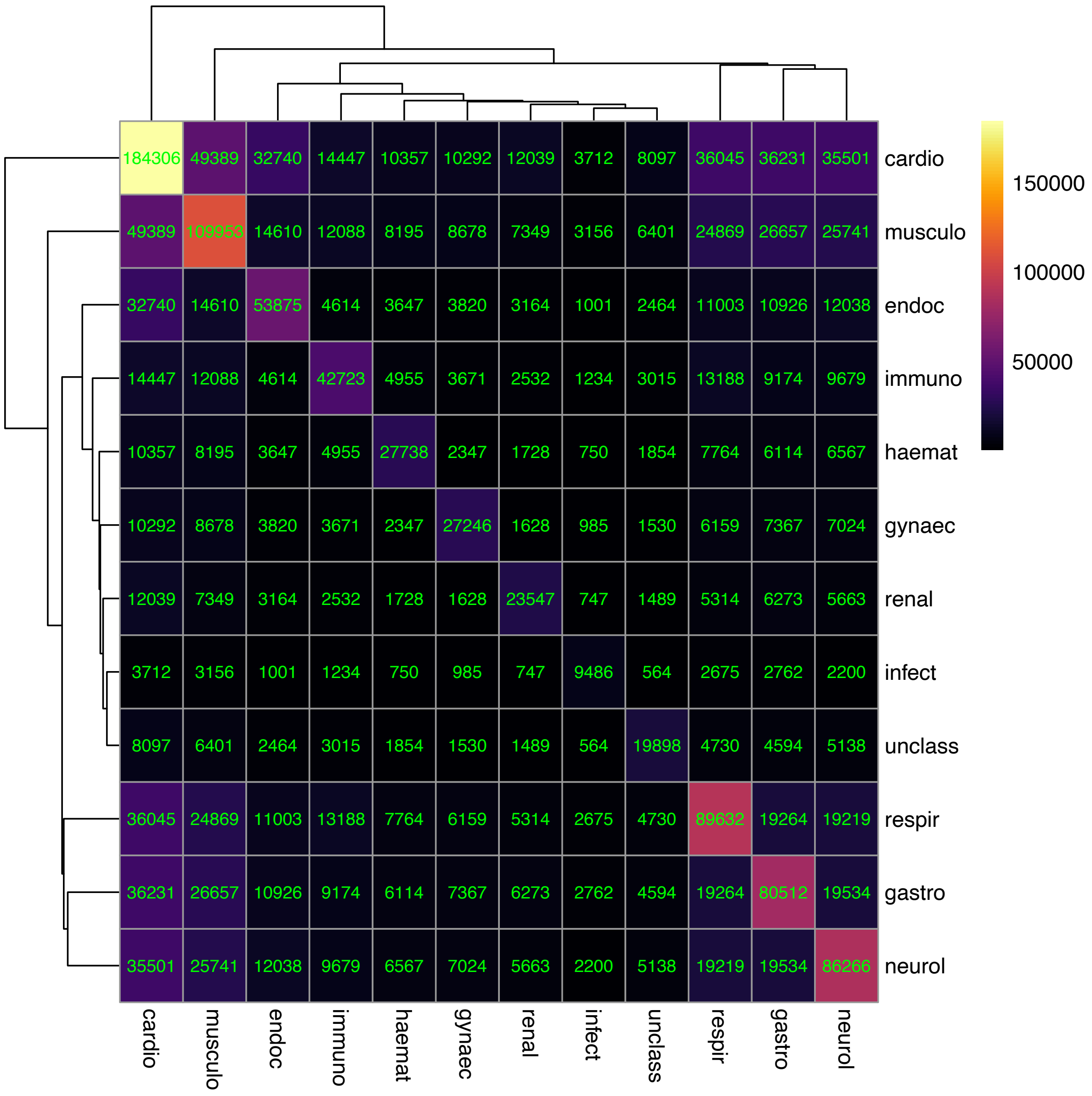


**Supplement e4-B.** Self-reported non-cancer illnesses. Cell entries reflect the number of individuals with ≥1 non-cancer illness of the group in the corresponding row and ≥1 non-cancer illness of the group in the corresponding column (e.g. 3 712 participants report ≥1 cardiovascular illness and ≥1 infection). Cell entries in the diagonal reflect the number of participants with ≥1 non-cancer illness of the group in the corresponding row or column (e.g. 184 306 participants report ≥1 cardiovascular illness). cardio = cardiovascular; musculo = musculoskeletal/trauma; endoc = endocrine/diabetes; immuno = immunological/systemic disorders; haemat = haematology/dermatology; gynaec = gynaecology/breast; renal = renal/urology; infect = infections; unclass = unclassifiable; respir = respiratory/ent; gastro = gastrointestinal/abdominal; neurol = neurology/eye/psychiatry.

**Supplement e5. Correlation matrix continuous variables**

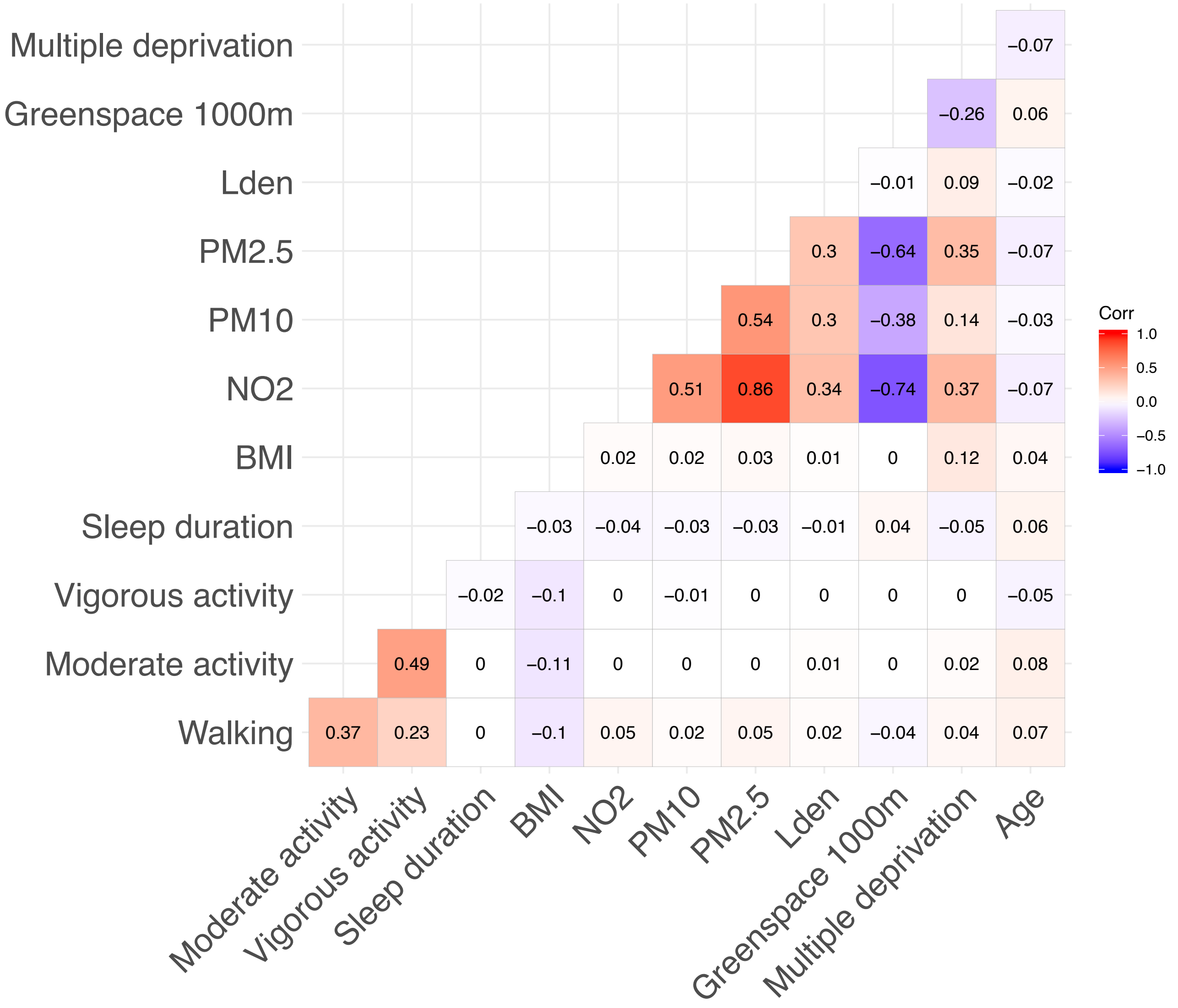


**Supplement e5.** Correlation matrix of continuous explanatory variables.

**Supplement e6. Descriptive statistics stratified by health indicators**

Stratified by health status

Supplement e6-A. **Baseline characteristics stratified by health status**

| | Unhealthy ($n$ = 95 177) | Healthy ($n$ = 212 201) | p value |
|---|---|---|---|
| Sociodemographic characteristics | | | |
| **Age** | | | < 0.001 |
| Mean (SD) | 58.46 (7.57) | 55.05 (8.06) | |
| Range | 38.00-72.00 | 39.00-73.00 | |
| **Sex** | | | < 0.001 |
| Female | 47 016 (49.4%) | 112 558 (53.0%) | |
| Male | 48 161 (50.6%) | 99 643 (47.0%) | |
| **Ethnicity** | | | < 0.001 |
| White | 91 415 (96.0%) | 202 150 (95.3%) | |
| Mixed-race | 456 (0.5%) | 1 310 (0.6%) | |
| Black | 1 099 (1.2%) | 3 158 (1.5%) | |
| Asian | 1 461 (1.5%) | 3 294 (1.6%) | |
| Chinese | 142 (0.1%) | 676 (0.3%) | |
| Other | 604 (0.6%) | 1 613 (0.8%) | |
| **Highest qualification** | | | < 0.001 |
| None | 16 351 (17.2%) | 23 477 (11.1%) | |
| O levels/GCSEs/CSEs | 25 045 (26.3%) | 59 403 (28.0%) | |
| A levels/NVQ/HND/HNC[1] | 22 885 (24.0%) | 49 699 (23.4%) | |
| Degree | 30 896 (32.5%) | 79 622 (37.5%) | |
| **Household income**[2] | | | < 0.001 |
| Very low | 26 699 (28.1%) | 36 400 (17.2%) | |
| Low | 26 029 (27.3%) | 51 902 (24.5%) | |
| Medium | 22 610 (23.8%) | 59 728 (28.1%) | |
| High | 15 769 (16.6%) | 50 337 (23.7%) | |
| Very high | 4 070 (4.3%) | 13 834 (6.5%) | |
| **Multiple deprivation** | | | < 0.001 |
| Mean (SD) | 17.89 (14.15) | 16.27 (12.92) | |
| Range | 0.61-82.00 | 0.61-82.00 | |
| Psychosocial factors | | | |
| **Loneliness** | | | < 0.001 |
| Not lonely | 88 594 (93.1%) | 201 307 (94.9%) | |
| Lonely | 6 583 (6.9%) | 10 894 (5.1%) | |
| **Social isolation** | | | < 0.001 |
| Not isolated | 85 472 (89.8%) | 195 459 (92.1%) | |
| Isolated | 9 705 (10.2%) | 16 742 (7.9%) | |
| Lifestyle factors | | | |
| **Smoking status** | | | < 0.001 |
| Never | 46 457 (48.8%) | 122 018 (57.5%) | |
| Former | 38 354 (40.3%) | 70 284 (33.1%) | |
| Current | 10 366 (10.9%) | 19 899 (9.4%) | |
| **Stair climbing frequency** | | | < 0.001 |
| None | 9 590 (10.1%) | 14 459 (6.8%) | |
| 1-5/day | 19 676 (20.7%) | 38 591 (18.2%) | |
| 6-10/day | 34 711 (36.5%) | 81 271 (38.3%) | |
| 11-15/day | 17 466 (18.4%) | 42 849 (20.2%) | |
| 16-20/day | 7 918 (8.3%) | 19 691 (9.3%) | |
| 20+/day | 5 816 (6.1%) | 15 340 (7.2%) | |
| **Alcohol intake frequency** | | | < 0.001 |
| Never | 8 582 (9.0%) | 11 841 (5.6%) | |
| Special occasions | 11 621 (12.2%) | 19 905 (9.4%) | |
| 1-3/month | 10 505 (11.0%) | 23 293 (11.0%) | |
| 1-2/week | 22 890 (24.0%) | 55 887 (26.3%) | |
| 3-4/week | 20 899 (22.0%) | 54 352 (25.6%) | |
| Daily/almost daily | 20 680 (21.7%) | 46 923 (22.1%) | |
| **Sleep duration** (hours/day) | | | < 0.001 |
| Mean (SD) | 7.19 (1.19) | 7.14 (0.99) | |
| Range | 1.00-20.00 | 1.00-20.00 | |
| **BMI** (kg/m$^2$) | | | < 0.001 |
| Mean (SD) | 27.96 (5.06) | 26.96 (4.44) | |
| Range | 12.80-66.20 | 12.80-67.30 | |
| **Walking** (days/week)[3] | | | < 0.001 |
| Mean (SD) | 5.34 (1.98) | 5.37 (1.93) | |
| Range | 0.00-7.00 | 0.00-7.00 | |
| **Moderate activity** (days/week)[3] | | | < 0.001 |
| Mean (SD) | 3.55 (2.37) | 3.60 (2.30) | |
| Range | 0.00-7.00 | 0.00-7.00 | |
| **Vigorous activity** (days/week)[3] | | | < 0.001 |
| Mean (SD) | 1.73 (1.96) | 1.94 (1.93) | |

| | | | |
|---|---|---|---|
| Range | 0.00-7.00 | 0.00-7.00 | |
| Environmental exposures | | | |
| **$PM_{2.5}$** | | | < 0.001 |
| Mean (SD) | 9.98 (1.05) | 9.94 (1.03) | |
| Range | 8.17-21.25 | 8.17-19.89 | |
| **$PM_{10}$** | | | < 0.001 |
| Mean (SD) | 16.21 (1.88) | 16.18 (1.88) | |
| Range | 11.78-30.65 | 11.78-30.65 | |
| **$NO_2$** | | | < 0.001 |
| Mean (SD) | 26.55 (7.60) | 26.38 (7.54) | |
| Range | 12.93-107.07 | 12.93-108.49 | |
| **$L_{den}$** | | | < 0.001 |
| Mean (SD) | 56.05 (4.29) | 55.99 (4.22) | |
| Range | 51.55-86.50 | 51.55-89.29 | |
| **Greenspace** 1000m | | | 0.750 |
| Mean (SD) | 45.50 (21.57) | 45.53 (21.86) | |
| Range | 4.67-99.18 | 4.49-99.19 | |

*Note:* GCSEs = general certificate of secondary education; CSE = certificate of secondary education; NVQ = national vocational qualification; HND = higher national diploma; HNC = higher national certificate; BMI = body mass index; PM = particulate matter; $NO_2$ = nitrogen dioxide; $L_{den}$ = day-evening-night noise level. [1]also includes 'other professional qualifications'. [2]Annual household income groups: very low (<£18 000), low (£18 000–30 999), middle (£31 000–51 999), high (£52 000–100 000) and very high (>£100 000). [3]number of days per week engaging in these activities for 10+ minutes continuously.

Stratified by long-standing illness

Supplement e6-B. **Baseline characteristics stratified by long-standing illness**

| | Yes<br>(*n* = 93 757) | No<br>(*n* = 213 621) | p value |
|---|---|---|---|
| Sociodemographic characteristics | | | |
| **Age** | | | < 0.001 |
| Mean (SD) | 57.70 (7.67) | 55.41 (8.14) | |
| Range | 39.00-72.00 | 38.00-73.00 | |
| **Sex** | | | < 0.001 |
| Female | 44 607 (47.6%) | 114 967 (53.8%) | |
| Male | 49 150 (52.4%) | 98 654 (46.2%) | |
| **Ethnicity** | | | < 0.001 |
| White | 89 561 (95.5%) | 204 004 (95.5%) | |
| Mixed-race | 487 (0.5%) | 1 279 (0.6%) | |
| Black | 1 419 (1.5%) | 2 838 (1.3%) | |
| Asian | 1 429 (1.5%) | 3 326 (1.6%) | |
| Chinese | 163 (0.2%) | 655 (0.3%) | |
| Other | 698 (0.7%) | 1 519 (0.7%) | |
| **Highest qualification** | | | < 0.001 |
| None | 15 643 (16.7%) | 24 185 (11.3%) | |
| O levels/GCSEs/CSEs | 24 847 (26.5%) | 59 601 (27.9%) | |
| A levels/NVQ/HND/HNC[1] | 23 010 (24.5%) | 49 574 (23.2%) | |
| Degree | 30 257 (32.3%) | 80 261 (37.6%) | |
| **Household income**[2] | | | < 0.001 |
| Very low | 27 247 (29.1%) | 35 852 (16.8%) | |
| Low | 25 264 (26.9%) | 52 667 (24.7%) | |
| Medium | 22 387 (23.9%) | 59 951 (28.1%) | |
| High | 15 291 (16.3%) | 50 815 (23.8%) | |
| Very high | 3 568 (3.8%) | 14 336 (6.7%) | |
| **Multiple deprivation** | | | < 0.001 |
| Mean (SD) | 18.79 (14.62) | 15.89 (12.63) | |
| Range | 0.61-82.00 | 0.61-82.00 | |
| Psychosocial factors | | | |
| **Loneliness** | | | < 0.001 |
| Not lonely | 86 392 (92.1%) | 203 509 (95.3%) | |
| Lonely | 7 365 (7.9%) | 10 112 (4.7%) | |
| **Social isolation** | | | < 0.001 |
| Not isolated | 83 385 (88.9%) | 197 546 (92.5%) | |
| Isolated | 10 372 (11.1%) | 16 075 (7.5%) | |
| Lifestyle factors | | | |
| **Smoking status** | | | < 0.001 |
| Never | 45 946 (49.0%) | 122 529 (57.4%) | |
| Former | 37 269 (39.8%) | 71 369 (33.4%) | |
| Current | 10 542 (11.2%) | 19 723 (9.2%) | |
| **Stair climbing frequency** | | | < 0.001 |
| None | 9 858 (10.5%) | 14 191 (6.6%) | |
| 1-5/day | 20 726 (22.1%) | 37 541 (17.6%) | |
| 6-10/day | 33 819 (36.1%) | 82 163 (38.5%) | |
| 11-15/day | 16 446 (17.5%) | 43 869 (20.5%) | |
| 16-20/day | 7 373 (7.9%) | 20 236 (9.5%) | |
| 20+/day | 5 535 (5.9%) | 15 621 (7.3%) | |

| | | | |
|---|---|---|---|
| **Alcohol intake frequency** | | | < 0.001 |
| Never | 9 151 (9.8%) | 11 272 (5.3%) | |
| Special occasions | 12 274 (13.1%) | 19 252 (9.0%) | |
| 1-3/month | 11 083 (11.8%) | 22 715 (10.6%) | |
| 1-2/week | 22 588 (24.1%) | 56 189 (26.3%) | |
| 3-4/week | 19 669 (21.0%) | 55 582 (26.0%) | |
| Daily/almost daily | 18 992 (20.3%) | 48 611 (22.8%) | |
| **Sleep duration** (hours/day) | | | 0.671 |
| Mean (SD) | 7.16 (1.24) | 7.16 (0.96) | |
| Range | 1.00-20.00 | 1.00-20.00 | |
| **BMI** (kg/m$^2$) | | | < 0.001 |
| Mean (SD) | 28.46 (5.23) | 26.75 (4.29) | |
| Range | 12.80-67.30 | 12.80-66.00 | |
| **Walking** (days/week)[3] | | | < 0.001 |
| Mean (SD) | 5.26 (2.04) | 5.41 (1.90) | |
| Range | 0.00-7.00 | 0.00-7.00 | |
| **Moderate activity** (days/week)[3] | | | < 0.001 |
| Mean (SD) | 3.47 (2.39) | 3.64 (2.28) | |
| Range | 0.00-7.00 | 0.00-7.00 | |
| **Vigorous activity** (days/week)[3] | | | < 0.001 |
| Mean (SD) | 1.65 (1.95) | 1.97 (1.93) | |
| Range | 0.00-7.00 | 0.00-7.00 | |
| Environmental exposures | | | |
| **$PM_{2.5}$** | | | < 0.001 |
| Mean (SD) | 10.01 (1.05) | 9.93 (1.03) | |
| Range | 8.17-21.25 | 8.17-19.89 | |
| **$PM_{10}$** | | | < 0.001 |
| Mean (SD) | 16.24 (1.86) | 16.17 (1.88) | |
| Range | 11.78-30.65 | 11.78-30.65 | |
| **$NO_2$** | | | < 0.001 |
| Mean (SD) | 26.77 (7.53) | 26.29 (7.57) | |
| Range | 12.93-108.49 | 12.93-107.81 | |
| **$L_{den}$** | | | 0.002 |
| Mean (SD) | 56.05 (4.28) | 56.00 (4.22) | |
| Range | 51.55-86.50 | 51.55-89.29 | |
| **Greenspace** 1000m | | | < 0.001 |
| Mean (SD) | 44.97 (21.27) | 45.76 (21.98) | |
| Range | 4.54-99.19 | 4.49-99.18 | |

*Note:* GCSEs = general certificate of secondary education; CSE = certificate of secondary education; NVQ = national vocational qualification; HND = higher national diploma; HNC = higher national certificate; BMI = body mass index; PM = particulate matter; $NO_2$ = nitrogen dioxide; $L_{den}$ = day-evening-night noise level. [1]also includes 'other professional qualifications'. [2]Annual household income groups: very low (<£18 000), low (£18 000–30 999), middle (£31 000–51 999), high (£52 000–100 000) and very high (>£100 000). [3]number of days per week engaging in these activities for 10+ minutes continuously.

Baseline characteristics stratified by self-rated health

Supplement e6-C. **Baseline characteristics stratified by self-rated health**

| | Poor (*n* = 11 066) | Fair (*n* = 59 169) | Good (*n* = 182 699) | Excellent (*n* = 54 444) | p value |
|---|---|---|---|---|---|
| Sociodemographic characteristics | | | | | |
| **Age** | | | | | < 0.001 |
| Mean (SD) | 55.91 (7.77) | 56.19 (8.09) | 56.25 (8.07) | 55.58 (8.06) | |
| Range | 40.00-72.00 | 39.00-73.00 | 38.00-72.00 | 39.00-71.00 | |
| **Sex** | | | | | < 0.001 |
| Female | 4 991 (45.1%) | 27 657 (46.7%) | 97 197 (53.2%) | 29 729 (54.6%) | |
| Male | 6 075 (54.9%) | 31 512 (53.3%) | 85 502 (46.8%) | 24 715 (45.4%) | |
| **Ethnicity** | | | | | < 0.001 |
| White | 10 299 (93.1%) | 55 498 (93.8%) | 175 085 (95.8%) | 52 683 (96.8%) | |
| Mixed-race | 82 (0.7%) | 401 (0.7%) | 999 (0.5%) | 284 (0.5%) | |
| Black | 236 (2.1%) | 1 127 (1.9%) | 2 359 (1.3%) | 535 (1.0%) | |
| Asian | 311 (2.8%) | 1 391 (2.4%) | 2 538 (1.4%) | 515 (0.9%) | |
| Chinese | 21 (0.2%) | 184 (0.3%) | 499 (0.3%) | 114 (0.2%) | |
| Other | 117 (1.1%) | 568 (1.0%) | 1 219 (0.7%) | 313 (0.6%) | |
| **Highest qualification** | | | | | < 0.001 |
| None | 2 723 (24.6%) | 10 888 (18.4%) | 21 910 (12.0%) | 4 307 (7.9%) | |
| O levels/GCSEs/CSEs | 3 133 (28.3%) | 17 564 (29.7%) | 51 129 (28.0%) | 12 622 (23.2%) | |
| A levels/NVQ/HND/HNC[1] | 2 664 (24.1%) | 14 193 (24.0%) | 43 648 (23.9%) | 12 079 (22.2%) | |
| Degree | 2 546 (23.0%) | 16 524 (27.9%) | 66 012 (36.1%) | 25 436 (46.7%) | |
| **Household income**[2] | | | | | < 0.001 |
| Very low | 5 224 (47.2%) | 16 903 (28.6%) | 33 777 (18.5%) | 7 195 (13.2%) | |
| Low | 2 673 (24.2%) | 15 853 (26.8%) | 47 574 (26.0%) | 11 831 (21.7%) | |
| Medium | 1 889 (17.1%) | 14 817 (25.0%) | 50 698 (27.7%) | 14 934 (27.4%) | |
| High | 1 072 (9.7%) | 9 699 (16.4%) | 40 407 (22.1%) | 14 928 (27.4%) | |
| Very high | 208 (1.9%) | 1 897 (3.2%) | 10 243 (5.6%) | 5 556 (10.2%) | |
| **Multiple deprivation** | | | | | < 0.001 |

| | | | | | |
|---|---|---|---|---|---|
| Mean (SD) | 24.45 (16.97) | 19.66 (14.84) | 16.08 (12.72) | 14.39 (11.60) | |
| Range | 0.61-82.00 | 0.61-82.00 | 0.61-82.00 | 0.61-82.00 | |
| Psychosocial factors | | | | | |
| **Loneliness** | | | | | < 0.001 |
| Not lonely | 9 116 (82.4%) | 53 565 (90.5%) | 174 338 (95.4%) | 52 882 (97.1%) | |
| Lonely | 1 950 (17.6%) | 5 604 (9.5%) | 8 361 (4.6%) | 1 562 (2.9%) | |
| **Social isolation** | | | | | < 0.001 |
| Not isolated | 8 848 (80.0%) | 52 191 (88.2%) | 168 805 (92.4%) | 51 087 (93.8%) | |
| Isolated | 2 218 (20.0%) | 6 978 (11.8%) | 13 894 (7.6%) | 3 357 (6.2%) | |
| Lifestyle factors | | | | | |
| **Smoking status** | | | | | < 0.001 |
| Never | 4 517 (40.8%) | 28 086 (47.5%) | 102 052 (55.9%) | 33 820 (62.1%) | |
| Former | 4 259 (38.5%) | 22 211 (37.5%) | 64 552 (35.3%) | 17 616 (32.4%) | |
| Current | 2 290 (20.7%) | 8 872 (15.0%) | 16 095 (8.8%) | 3 008 (5.5%) | |
| **Stair climbing frequency** | | | | | < 0.001 |
| None | 1 830 (16.5%) | 5 687 (9.6%) | 13 311 (7.3%) | 3 221 (5.9%) | |
| 1-5/day | 3 674 (33.2%) | 13 892 (23.5%) | 32 466 (17.8%) | 8 235 (15.1%) | |
| 6-10/day | 3 263 (29.5%) | 22 079 (37.3%) | 70 409 (38.5%) | 20 231 (37.2%) | |
| 11-15/day | 1 270 (11.5%) | 9 948 (16.8%) | 37 158 (20.3%) | 11 939 (21.9%) | |
| 16-20/day | 568 (5.1%) | 4 328 (7.3%) | 16 862 (9.2%) | 5 851 (10.7%) | |
| 20+/day | 461 (4.2%) | 3 235 (5.5%) | 12 493 (6.8%) | 4 967 (9.1%) | |
| **Alcohol intake frequency** | | | | | < 0.001 |
| Never | 1 944 (17.6%) | 5 319 (9.0%) | 10 392 (5.7%) | 2 768 (5.1%) | |
| Special occasions | 2 081 (18.8%) | 8 077 (13.7%) | 17 346 (9.5%) | 4 022 (7.4%) | |
| 1-3/month | 1 421 (12.8%) | 7 222 (12.2%) | 19 906 (10.9%) | 5 249 (9.6%) | |
| 1-2/week | 2 298 (20.8%) | 14 795 (25.0%) | 47 880 (26.2%) | 13 804 (25.4%) | |
| 3-4/week | 1 525 (13.8%) | 12 256 (20.7%) | 46 384 (25.4%) | 15 086 (27.7%) | |
| Daily/almost daily | 1 797 (16.2%) | 11 500 (19.4%) | 40 791 (22.3%) | 13 515 (24.8%) | |
| **Sleep duration** (hours/day) | | | | | < 0.001 |
| Mean (SD) | 7.13 (1.80) | 7.07 (1.19) | 7.17 (0.98) | 7.23 (0.92) | |
| Range | 1.00-20.00 | 1.00-16.00 | 1.00-20.00 | 2.00-18.00 | |
| **BMI** ($kg/m^2$) | | | | | < 0.001 |
| Mean (SD) | 30.68 (6.72) | 29.13 (5.29) | 27.00 (4.27) | 25.46 (3.50) | |
| Range | 12.80-67.30 | 13.80-66.00 | 12.80-63.40 | 14.90-51.90 | |
| **Walking** (days/week)[3] | | | | | < 0.001 |
| Mean (SD) | 4.53 (2.44) | 5.14 (2.06) | 5.42 (1.88) | 5.58 (1.84) | |
| Range | 0.00-7.00 | 0.00-7.00 | 0.00-7.00 | 0.00-7.00 | |
| **Moderate activity** (days/week)[3] | | | | | < 0.001 |
| Mean (SD) | 2.65 (2.52) | 3.29 (2.38) | 3.63 (2.28) | 3.97 (2.26) | |
| Range | 0.00-7.00 | 0.00-7.00 | 0.00-7.00 | 0.00-7.00 | |
| **Vigorous activity** (days/week)[3] | | | | | < 0.001 |
| Mean (SD) | 0.98 (1.75) | 1.46 (1.88) | 1.89 (1.90) | 2.45 (2.02) | |
| Range | 0.00-7.00 | 0.00-7.00 | 0.00-7.00 | 0.00-7.00 | |
| Environmental exposures | | | | | |
| **$PM_{2.5}$** | | | | | < 0.001 |
| Mean (SD) | 10.19 (1.07) | 10.05 (1.04) | 9.93 (1.03) | 9.88 (1.05) | |
| Range | 8.17-17.24 | 8.17-21.25 | 8.17-19.76 | 8.17-19.89 | |
| **$PM_{10}$** | | | | | < 0.001 |
| Mean (SD) | 16.37 (1.83) | 16.29 (1.85) | 16.17 (1.88) | 16.10 (1.91) | |
| Range | 11.78-25.48 | 11.78-30.65 | 11.78-30.65 | 11.78-29.90 | |
| **$NO_2$** | | | | | < 0.001 |
| Mean (SD) | 28.01 (7.53) | 27.13 (7.43) | 26.27 (7.51) | 25.93 (7.77) | |
| Range | 12.93-98.49 | 12.93-105.88 | 12.93-108.49 | 12.93-107.81 | |
| **$L_{den}$** | | | | | < 0.001 |
| Mean (SD) | 56.17 (4.37) | 56.08 (4.31) | 55.99 (4.23) | 55.97 (4.17) | |
| Range | 51.56-84.41 | 51.56-89.29 | 51.55-86.50 | 51.55-84.65 | |
| **Greenspace** 1000m | | | | | < 0.001 |
| Mean (SD) | 42.57 (20.15) | 44.00 (20.81) | 45.88 (21.81) | 46.54 (22.81) | |
| Range | 6.45-98.60 | 6.08-99.14 | 4.49-99.18 | 4.67-99.19 | |

*Note:* GCSEs = general certificate of secondary education; CSE = certificate of secondary education; NVQ = national vocational qualification; HND = higher national diploma; HNC = higher national certificate; BMI = body mass index; PM = particulate matter; $NO_2$ = nitrogen dioxide; $L_{den}$ = day-evening-night noise level. [1]also includes 'other professional qualifications'. [2]Annual household income groups: very low (<£18 000), low (£18 000–30 999), middle (£31 000–51 999), high (£52 000–100 000) and very high (>£100 000). [3]number of days per week engaging in these activities for 10+ minutes continuously.

**Supplement e7. Visual summary of findings**

Lifestyle factors

| | Outcome | | | | | Health status | | | | | | Long-standing illness | | | | | | Self-rated health | | | | | |
|---|---|---|---|---|---|---|---|---|---|---|---|---|---|---|---|---|---|---|---|---|---|---|---|
| | Cross-sectional | | | Prospective | | Sex | | | Age | | | Sex | | | Age | | | Sex | | | Age | | |
| Variable | Health status | Long-standing illness | Self-rated health | Self-rated health t1 | Self-rated health t2 | Stratified (m/f) | | Interaction term | Stratified (<65/65+) | | Interaction term | Stratified (m/f) | | Interaction term | Stratified (<65/65+) | | Interaction term | Stratified (m/f) | | Interaction term | Stratified (<65/65+) | | Interaction term |
| **Sleep** | ↓ | -- | ↑ | ↑ | ↑ | ↓ | -- | √√ | ↓ | -- | ns | -- | -- | ns | -- | -- | √ | ↑ | ↑ | ns | ↑ | ↑ | √√ |
| **Physical activity** | | | | | | | | | | | | | | | | | | | | | | | |
| Walking | ↑ | ↑ | ↑ | ↑ | ↑ | -- | ↑ | ns | ↑ | ↑ | √√ | ↑ | ↑ | √√ | ↑ | ↑ | √√ | ↑ | ↑ | √ | ↑ | ↑ | √√ |
| Moderate | -- | -- | ↑ | -- | -- | -- | -- | √ | -- | -- | ns | -- | ↓ | √ | -- | -- | √√ | ↑ | -- | √√ | ↑ | ↑ | ns |
| Vigorous | ↑ | ↑ | ↑ | ↑ | ↑ | ↑ | ↑ | √√ | ↑ | ↑ | ns | ↑ | ↑ | ns | ↑ | ↑ | ns | ↑ | ↑ | √√ | ↑ | ↑ | √√ |
| **Stair climbing frequency** | | | | | | | | | | | | | | | | | | | | | | | |
| None | | | | | | | | | | | | | | | | | | | | | | | |
| 1-5/day | ↑ | ↑ | ↑ | -- | -- | -- | -- | ns | ↑ | -- | √√ | -- | ↑ | ns | ↑ | -- | √√ | -- | -- | ns | ↑ | -- | √√ |
| 6-10/day | ↑ | ↑ | ↑ | -- | -- | ↑ | ↑ | √ | ↑ | ↑ | √√ | ↑ | ↑ | ns | ↑ | ↑ | √√ | ↑ | ↑ | √ | ↑ | ↑ | √ |
| 11-15/day | ↑ | ↑ | ↑ | -- | -- | ↑ | ↑ | √ | ↑ | ↑ | √√ | ↑ | ↑ | ns | ↑ | ↑ | √√ | ↑ | ↑ | √√ | ↑ | ↑ | ns |
| 16-20/day | ↑ | ↑ | ↑ | -- | -- | ↑ | ↑ | ns | ↑ | ↑ | √ | ↑ | ↑ | ns | ↑ | ↑ | ns | ↑ | ↑ | √√ | ↑ | ↑ | ns |
| 20+/day | ↑ | ↑ | ↑ | -- | -- | ↑ | ↑ | √√ | ↑ | -- | √√ | ↑ | ↑ | ns | ↑ | ↑ | ns | ↑ | ↑ | √√ | ↑ | ↑ | √ |
| **Alcohol intake frequency** | | | | | | | | | | | | | | | | | | | | | | | |
| Never | ↓ | ↓ | ↓ | ↓ | ↓ | ↓ | ↓ | √ | ↓ | ↓ | √√ | ↓ | ↓ | √√ | ↓ | ↓ | √ | ↓ | ↓ | √√ | ↓ | ↓ | √√ |
| Special | ↓ | ↓ | ↓ | ↓ | ↓ | ↓ | ↓ | ns | ↓ | ↓ | √√ | ↓ | ↓ | ns | ↓ | ↓ | √√ | ↓ | ↓ | ns | ↓ | ↓ | ns |
| 1-3/month | ↓ | ↓ | ↓ | -- | -- | ↓ | ↓ | ns | ↓ | -- | ns | ↓ | ↓ | ns | ↓ | ↓ | √ | ↓ | ↓ | ns | ↓ | -- | √ |
| 1-2/week | | | | | | | | | | | | | | | | | | | | | | | |
| 3-4/week | ↑ | ↑ | ↑ | -- | -- | ↑ | -- | √ | ↑ | -- | ns | ↑ | ↑ | ns | ↑ | ↑ | ns | -- | ↑ | ns | ↑ | ↑ | ns |
| Daily | -- | ↑ | -- | -- | -- | ↑ | -- | √ | -- | -- | ns | ↑ | ↑ | ns | ↑ | ↑ | ns | -- | ↑ | ns | -- | ↑ | √√ |
| **BMI** | ↓ | ↓ | ↓ | ↓ | ↓ | ↓ | ↓ | √√ | ↓ | ↓ | √√ | ↓ | ↓ | √√ | ↓ | ↓ | √√ | ↓ | ↓ | √√ | ↓ | ↓ | √√ |
| **Smoking status** | | | | | | | | | | | | | | | | | | | | | | | |
| Never | | | | | | | | | | | | | | | | | | | | | | | |
| Former | ↓ | ↓ | ↓ | ↓ | ↓ | ↓ | ↓ | √√ | ↓ | ↓ | √√ | ↓ | ↓ | √√ | ↓ | ↓ | √√ | ↓ | ↓ | √√ | ↓ | ↓ | ns |
| Current | ↓ | ↓ | ↓ | ↓ | ↓ | ↓ | ↓ | √ | ↓ | ↓ | √ | ↓ | ↓ | ns | ↓ | ↓ | ns | ↓ | ↓ | √√ | ↓ | ↓ | √√ |

Table legend

| | |
|---|---|
| ↓ | associated with unfavourable health |
| ↓ | associated with unfavourable health; stronger in this stratum |
| ↑ | associated with favourable health |
| ↑ | associated with favourable health; stronger in this stratum |
| -- | no evidence of association with health |
| √√ | statistically significant – Bonferroni correction |
| √ | statistically significant – Benjamini & Hochberg correction |
| ns | not statistically significant |

Environmental exposures

| | Outcome | | | | | Health status | | | | | | Long-standing illness | | | | | | Self-rated health | | | | | |
|---|---|---|---|---|---|---|---|---|---|---|---|---|---|---|---|---|---|---|---|---|---|---|---|
| | Cross-sectional | | | Prospective | | Sex | | | Age | | | Sex | | | Age | | | Sex | | | Age | | |
| Variable | Health status | Long-standing illness | Self-rated health | Self-rated health t1 | Self-rated health t2 | Stratified (m/f) | | Interaction term | Stratified (<65/65+) | | Interaction term | Stratified (m/f) | | Interaction term | Stratified (<65/65+) | | Interaction term | Stratified (m/f) | | Interaction term | Stratified (<65/65+) | | Interaction term |
| **$PM_{2.5}$** | ↓ | ↓ | ↑ | -- | -- | ↓ | -- | ✓ | ↓ | -- | ns | ↓ | -- | ns | ↓ | -- | ns | -- | -- | ✓✓ | ↑ | -- | ✓✓ |
| **$PM_{10}$** | -- | -- | -- | -- | -- | -- | -- | ns | -- | -- | ns | -- | -- | ns | -- | -- | ns | -- | -- | ✓ | -- | -- | ns |
| **$NO_2$** | -- | -- | ↓ | -- | -- | -- | ↑ | ✓ | -- | -- | ns | -- | -- | ns | -- | -- | ns | -- | ↓ | ✓ | -- | -- | ✓✓ |
| **$L_{den}$** | -- | -- | -- | -- | -- | -- | -- | ns | -- | -- | ✓ | -- | -- | ns | -- | -- | ns | -- | -- | ns | -- | -- | ✓✓ |
| **Greenspace** | -- | -- | -- | -- | -- | -- | -- | ns | -- | -- | ns | -- | -- | ns | -- | -- | ns | -- | -- | ns | -- | -- | ✓ |

| Table legend | |
|---|---|
| ↓ (light orange) | associated with unfavourable health |
| ↓ (dark orange) | associated with unfavourable health; stronger in this stratum |
| ↑ (light green) | associated with favourable health |
| ↑ (dark green) | associated with favourable health; stronger in this stratum |
| -- | no evidence of association with health |
| ✓✓ | statistically significant – Bonferroni correction |
| ✓ | statistically significant – Benjamini & Hochberg correction |
| ns | not statistically significant |

Psychosocial factors

| | Outcome | | | | | Health status | | | | Long-standing illness | | | | Self-rated health | | | |
|---|---|---|---|---|---|---|---|---|---|---|---|---|---|---|---|---|---|
| | Cross-sectional | | | Prospective | | Sex | | Age | | Sex | | Age | | Sex | | Age | |
| Variable | Health status | Long-standing illness | Self-rated health | Self-rated health t1 | Self-rated health t2 | Stratified (m/f) | Interaction term | Stratified (<65/65+) | Interaction term | Stratified (m/f) | Interaction term | Stratified (<65/65+) | Interaction term | Stratified (m/f) | Interaction term | Stratified (<65/65+) | Interaction term |
| **Loneliness** | | | | | | | | | | | | | | | | | |
| Not lonely | | | | | | | | | | | | | | | | | |
| Lonely | ↓ | ↓ | ↓ | ↓ | ↓ | ↓ ↓ | ✓✓ | ↓ -- | ✓ | ↓ ↓ | ✓✓ | ↓ ↓ | ns | ↓ ↓ | ✓✓ | ↓ ↓ | ✓✓ |
| **Social isolation** | | | | | | | | | | | | | | | | | |
| Not isolated | | | | | | | | | | | | | | | | | |
| Isolated | ↓ | ↓ | ↓ | -- | ↓ | ↓ -- | ✓ | ↓ -- | ✓✓ | ↓ ↓ | ns | ↓ -- | ✓✓ | ↓ ↓ | ✓✓ | ↓ -- | ✓✓ |

Table legend

| Symbol | Meaning |
|---|---|
| ↓ | associated with unfavourable health |
| ↓ | associated with unfavourable health; stronger in this stratum |
| ↑ | associated with favourable health |
| ↑ | associated with favourable health; stronger in this stratum |
| -- | no evidence of association with health |
| ✓✓ | statistically significant – Bonferroni correction |
| ✓ | statistically significant – Benjamini & Hochberg correction |
| ns | not statistically significant |

Sociodemographic characteristics

| Variable | Outcome | | | | | Health status | | | | | | Long-standing illness | | | | | | Self-rated health | | | | | |
|---|---|---|---|---|---|---|---|---|---|---|---|---|---|---|---|---|---|---|---|---|---|---|---|
| | Cross-sectional | | | Prospective | | Sex | | | Age | | | Sex | | | Age | | | Sex | | | Age | | |
| | Health status | Long-standing illness | Self-rated health | Self-rated health t1 | Self-rated health t2 | Stratified (m/f) | | Interaction term | Stratified (<65/65+) | | Interaction term | Stratified (m/f) | | Interaction term | Stratified (<65/65+) | | Interaction term | Stratified (m/f) | | Interaction term | Stratified (<65/65+) | | Interaction term |
| **Household income** | | | | | | | | | | | | | | | | | | | | | | | |
| Very low | ↓ | ↓ | ↓ | ↓ | ↓ | ↓ | ↓ | ✓✓ | ↓ | ↓ | ✓✓ | ↓ | ↓ | ✓✓ | ↓ | ↓ | ✓✓ | ↓ | ↓ | ✓✓ | ↓ | ↓ | ✓✓ |
| Low | ↓ | ↓ | ↓ | -- | ↓ | ↓ | ↓ | ✓✓ | ↓ | -- | ns | ↓ | ↓ | ✓✓ | ↓ | -- | ✓ | ↓ | ↓ | ✓ | ↓ | ↓ | ✓ |
| Middle | | | | | | | | | | | | | | | | | | | | | | | |
| High | ↑ | ↑ | ↑ | ↑ | ↑ | -- | -- | ✓ | ↑ | -- | ✓ | ↑ | ↑ | ns | ↑ | -- | ns | ↑ | ↑ | ✓✓ | ↑ | -- | ✓✓ |
| Very high | -- | ↑ | ↑ | ↑ | ↑ | -- | -- | ✓ | -- | -- | ns | ↑ | ↑ | ns | ↑ | -- | ns | ↑ | ↑ | ✓✓ | ↑ | -- | ✓✓ |
| **Sex** | | | | | | | | | | | | | | | | | | | | | | | |
| Female | | | | | | | | | | | | | | | | | | | | | | | |
| Male | ↓ | ↓ | ↓ | ↓ | ↓ | | | | ↓ | ↓ | ✓✓ | | | | ↓ | ↓ | ✓✓ | | | | ↓ | ↓ | ✓✓ |
| **Age** | ↓ | ↓ | ↑ | -- | ↑ | ↓ | ↓ | ✓✓ | ↓ | ↓ | | ↓ | ↓ | ✓✓ | ↓ | ↓ | | ↑ | ↑ | ✓✓ | ↑ | -- | |
| **Deprivation** | ↓ | ↓ | ↓ | -- | ↓ | ↓ | ↓ | ✓✓ | ↓ | ↓ | ns | ↓ | ↓ | ✓ | ↓ | ↓ | ns | ↓ | ↓ | ✓✓ | ↓ | ↓ | ns |
| **Ethnicity** | | | | | | | | | | | | | | | | | | | | | | | |
| White | | | | | | | | | | | | | | | | | | | | | | | |
| Mixed-race | -- | -- | -- | -- | -- | -- | -- | ns | -- | -- | ns | -- | -- | ns | -- | -- | ns | -- | -- | ✓ | -- | -- | ns |
| Asian | -- | ↑ | ↓ | -- | ↓ | -- | ↑ | ✓ | -- | -- | ✓✓ | ↑ | ↑ | ns | ↑ | -- | ✓✓ | ↓ | ↓ | ✓ | ↓ | ↓ | ✓✓ |
| Black | ↑ | ↑ | -- | -- | -- | ↑ | ↑ | ns | ↑ | -- | ns | ↑ | -- | ✓✓ | ↑ | -- | ✓✓ | ↑ | -- | ✓✓ | ↑ | ↓ | ✓✓ |
| Chinese | ↑ | ↑ | ↓ | -- | -- | ↑ | ↑ | ns | ↑ | -- | ns | ↑ | ↑ | ns | ↑ | -- | ns | -- | ↓ | ns | ↓ | -- | ns |
| Other | ↑ | ↑ | -- | -- | -- | ↑ | -- | ns | ↑ | -- | ns | ↑ | -- | ns | ↑ | -- | ✓✓ | -- | ↓ | ✓✓ | -- | ↓ | ✓✓ |
| **Highest qualification** | | | | | | | | | | | | | | | | | | | | | | | |
| None | | | | | | | | | | | | | | | | | | | | | | | |
| GCSEs | -- | -- | ↑ | -- | -- | -- | -- | ✓✓ | -- | -- | ns | -- | -- | ✓✓ | -- | -- | ✓✓ | ↑ | ↑ | ✓ | ↑ | ↑ | ✓✓ |
| A levels | -- | ↓ | ↑ | -- | -- | -- | -- | ✓✓ | -- | -- | ✓ | ↓ | ↓ | ✓✓ | ↓ | ↓ | ✓✓ | ↑ | ↑ | ✓✓ | ↑ | ↑ | ✓✓ |
| Degree | ↓ | ↓ | ↑ | -- | ↑ | -- | -- | ✓✓ | -- | -- | ✓ | ↓ | ↓ | ✓✓ | ↓ | ↓ | ✓✓ | ↑ | ↑ | ✓✓ | ↑ | ↑ | ✓✓ |

Table legend

| Symbol | Meaning |
|---|---|
| ↓ | associated with unfavourable health |
| ↓ | associated with unfavourable health; stronger in this stratum |
| ↑ | associated with favourable health |
| ↑ | associated with favourable health; stronger in this stratum |
| -- | no evidence of association with health |
| ✓✓ | statistically significant – Bonferroni correction |
| ✓ | statistically significant – Benjamini & Hochberg correction |
| ns | not statistically significant |

**Supplement e8. Regression tables long-standing illness**

Sociodemographic characteristics

| Supplement e8-A. **Sociodemographic characteristics associated with long-standing illness** | | | | | | | | | |
|---|---|---|---|---|---|---|---|---|---|
| | Model 1 | | | Model 2 | | | Model 3 | | |
| Term | OR | Bonferroni-corrected CI | | OR | Bonferroni-corrected CI | | OR | Bonferroni-corrected CI | |
| **Household income**[1] | | | | | | | | | |
| Very low | 0.4914 | 0.4739 | 0.5094 | 0.5362 | 0.5166 | 0.5565 | 0.6654 | 0.6384 | 0.6934 |
| Low | 0.7785 | 0.7515 | 0.8064 | 0.8323 | 0.8029 | 0.8628 | 0.8906 | 0.8581 | 0.9244 |
| Middle | Ref | – | – | Ref | – | – | Ref | – | – |
| High | 1.2410 | 1.1936 | 1.2903 | 1.1933 | 1.1473 | 1.2412 | 1.1171 | 1.0726 | 1.1634 |
| Very high | 1.5004 | 1.4059 | 1.6021 | 1.4327 | 1.3419 | 1.5305 | 1.2524 | 1.1703 | 1.3410 |
| **Sex** | | | | | | | | | |
| Female | Ref | – | – | Ref | – | – | Ref | – | – |
| Male | 0.7788 | 0.7594 | 0.7987 | – | – | – | 0.7585 | 0.7380 | 0.7797 |
| **Age** | 0.9644 | 0.9629 | 0.9660 | – | – | – | 0.9730 | 0.9711 | 0.9748 |
| **Multiple deprivation** | 0.9845 | 0.9836 | 0.9854 | 0.9827 | 0.9818 | 0.9836 | 0.9921 | 0.9910 | 0.9932 |
| **Ethnicity** | | | | | | | | | |
| White | Ref | – | – | Ref | – | – | Ref | – | – |
| Mixed-race | 1.1530 | 0.9729 | 1.3724 | 0.9368 | 0.7890 | 1.1172 | 1.1223 | 0.9387 | 1.3474 |
| Asian | 1.0218 | 0.9227 | 1.1331 | 0.9252 | 0.8343 | 1.0273 | 1.2225 | 1.0962 | 1.3652 |
| Black | 0.8780 | 0.7906 | 0.9763 | 0.7200 | 0.6474 | 0.8017 | 1.1852 | 1.0591 | 1.3277 |
| Chinese | 1.7641 | 1.3404 | 2.3591 | 1.5038 | 1.1401 | 2.0150 | 1.6205 | 1.2207 | 2.1842 |
| Other | 0.9554 | 0.8252 | 1.1091 | 0.8340 | 0.7191 | 0.9697 | 1.2027 | 1.0301 | 1.4078 |
| **Highest qualification** | | | | | | | | | |
| None | Ref | – | – | Ref | – | – | Ref | – | – |
| O levels/GCSEs/CSEs | 1.5515 | 1.4891 | 1.6165 | 1.2693 | 1.2166 | 1.3241 | 0.9920 | 0.9485 | 1.0375 |
| A levels/NVQ/HND/HNC[2] | 1.3935 | 1.3364 | 1.4531 | 1.2067 | 1.1562 | 1.2594 | 0.8953 | 0.8554 | 0.9371 |
| Degree | 1.7157 | 1.6492 | 1.7849 | 1.4204 | 1.3635 | 1.4796 | 0.8550 | 0.8167 | 0.8950 |

*Note:* Bonferroni-adjusted (~99.9%) confidence intervals. OR = odds ratio; CI = confidence interval; GCSEs = general certificate of secondary education; CSE = certificate of secondary education; NVQ = national vocational qualification; HND = higher national diploma; HNC = higher national certificate. [1]Annual household income groups: very low (<£18 000), low (£18 000–30 999), middle (£31 000–51 999), high (£52 000–100 000) and very high (>£100 000). [2]also includes 'other professional qualifications'.
Model 1 – only individual explanatory variables.
Model 2 – adjusted for age and sex.
Model 3 – all explanatory variables.

Supplement e8-B. **Psychosocial factors associated with long-standing illness**

| | Model 1 | | | Model 2 | | | Model 3 | | |
|---|---|---|---|---|---|---|---|---|---|
| Term | OR | Bonferroni-corrected CI | | OR | Bonferroni-corrected CI | | OR | Bonferroni-corrected CI | |
| **Loneliness** | | | | | | | | | |
| Not lonely | Ref | – | – | Ref | – | – | Ref | – | – |
| Lonely | 0.5828 | 0.5539 | 0.6134 | 0.5658 | 0.5373 | 0.5958 | 0.7097 | 0.6722 | 0.7494 |
| **Social isolation** | | | | | | | | | |
| Not isolated | Ref | – | – | Ref | – | – | Ref | – | – |
| Isolated | 0.6542 | 0.6269 | 0.6828 | 0.6558 | 0.6281 | 0.6847 | 0.9185 | 0.8770 | 0.9621 |

*Note:* Bonferroni-adjusted (~99.9%) confidence intervals. OR = odds ratio; CI = confidence interval.

Model 1 – only individual explanatory variables.

Model 2 – adjusted for age and sex.

Model 3 – all explanatory variables.

| Supplement e8-C. **Lifestyle factors associated with long-standing illness** | | | | | | | | | |
|---|---|---|---|---|---|---|---|---|---|
| | Model 1 | | | Model 2 | | | Model 3 | | |
| Term | OR | Bonferroni-corrected CI | | OR | Bonferroni-corrected CI | | OR | Bonferroni-corrected CI | |
| **Sleep duration** (hours/day) | 0.9984 | 0.9866 | 1.0104 | 1.0142 | 1.0020 | 1.0265 | 1.0004 | 0.9882 | 1.0128 |
| **Physical activity** (days/week)[1] | | | | | | | | | |
| Walking | 1.0408 | 1.0341 | 1.0475 | 1.0505 | 1.0437 | 1.0573 | 1.0250 | 1.0175 | 1.0325 |
| Moderate activity | 1.0319 | 1.0263 | 1.0376 | 1.0428 | 1.0370 | 1.0486 | 0.9984 | 0.9916 | 1.0053 |
| Vigorous activity | 1.0917 | 1.0844 | 1.0991 | 1.0918 | 1.0844 | 1.0993 | 1.0641 | 1.0557 | 1.0727 |
| **Stair climbing frequency** | | | | | | | | | |
| None | Ref | – | – | Ref | – | – | Ref | – | – |
| 1-5/day | 1.2582 | 1.1962 | 1.3235 | 1.1326 | 1.0759 | 1.1923 | 1.1101 | 1.0522 | 1.1710 |
| 6-10/day | 1.6877 | 1.6100 | 1.7690 | 1.5338 | 1.4623 | 1.6086 | 1.2937 | 1.2307 | 1.3598 |
| 11-15/day | 1.8530 | 1.7600 | 1.9508 | 1.6718 | 1.5868 | 1.7612 | 1.3131 | 1.2434 | 1.3866 |
| 16-20/day | 1.9066 | 1.7941 | 2.0263 | 1.7183 | 1.6159 | 1.8275 | 1.3066 | 1.2255 | 1.3932 |
| 20+/day | 1.9605 | 1.8360 | 2.0939 | 1.7164 | 1.6061 | 1.8347 | 1.2975 | 1.2108 | 1.3907 |
| **Alcohol intake frequency** | | | | | | | | | |
| Never | 0.4952 | 0.4701 | 0.5216 | 0.4959 | 0.4704 | 0.5227 | 0.5533 | 0.5234 | 0.5850 |
| Special occasions | 0.6305 | 0.6028 | 0.6596 | 0.6119 | 0.5844 | 0.6406 | 0.7142 | 0.6809 | 0.7492 |
| 1-3/month | 0.8239 | 0.7876 | 0.8620 | 0.7846 | 0.7495 | 0.8213 | 0.8486 | 0.8096 | 0.8895 |
| 1-2/week | Ref | – | – | Ref | – | – | Ref | – | – |
| 3-4/week | 1.1360 | 1.0949 | 1.1786 | 1.1808 | 1.1376 | 1.2257 | 1.0925 | 1.0514 | 1.1353 |
| Daily/almost daily | 1.0289 | 0.9911 | 1.0682 | 1.1540 | 1.1108 | 1.1989 | 1.0744 | 1.0326 | 1.1180 |
| **BMI** ($kg/m^2$) | 0.9259 | 0.9234 | 0.9284 | 0.9276 | 0.9250 | 0.9301 | 0.9419 | 0.9392 | 0.9446 |
| **Smoking status** | | | | | | | | | |
| Never | Ref | – | – | Ref | – | – | Ref | – | – |
| Former | 0.7181 | 0.6989 | 0.7378 | 0.7898 | 0.7683 | 0.8119 | 0.7978 | 0.7749 | 0.8213 |
| Current | 0.7016 | 0.6723 | 0.7322 | 0.6864 | 0.6574 | 0.7168 | 0.7858 | 0.7506 | 0.8227 |

*Note:* Bonferroni-adjusted (~99.9%) confidence intervals. OR = odds ratio; CI = confidence interval; BMI = body mass index. [1]number of days per week engaging in these activities for 10+ minutes continuously.
Model 1 – only individual explanatory variables.
Model 2 – adjusted for age and sex.
Model 3 – all explanatory variables.

Supplement e8-D. **Environmental exposures associated with long-standing illness**

| | Model 1 | | | Model 2 | | | Model 3 | | |
|---|---|---|---|---|---|---|---|---|---|
| Term | OR | Bonferroni-corrected CI | | OR | Bonferroni-corrected CI | | OR | Bonferroni-corrected CI | |
| **$PM_{2.5}$** | 0.9254 | 0.9144 | 0.9367 | 0.9065 | 0.8955 | 0.9176 | 0.9677 | 0.9430 | 0.9932 |
| **$PM_{10}$** | 0.9814 | 0.9748 | 0.9880 | 0.9762 | 0.9696 | 0.9828 | 1.0019 | 0.9934 | 1.0105 |
| **$NO_2$** | 0.9917 | 0.9901 | 0.9933 | 0.9887 | 0.9870 | 0.9904 | 1.0025 | 0.9982 | 1.0068 |
| **$L_{den}$** | 0.9971 | 0.9942 | 1.0001 | 0.9958 | 0.9928 | 0.9988 | 1.0015 | 0.9979 | 1.0052 |
| **Greenspace** 1000m | 1.0017 | 1.0011 | 1.0022 | 1.0026 | 1.0020 | 1.0032 | 0.9997 | 0.9987 | 1.0007 |

*Note:* Bonferroni-adjusted (~99.9%) confidence intervals. OR = odds ratio; CI = confidence interval; PM = particulate matter; $NO_2$ = nitrogen dioxide; $L_{den}$ = day-evening-night noise level.
Model 1 – only individual explanatory variables.
Model 2 – adjusted for age and sex.
Model 3 – all explanatory variables.

**Supplement e9. Regression tables self-rated health**

Sociodemographic characteristics

Supplement e9-A. **Sociodemographic characteristics associated with self-rated health**

| | Model 1 | | | Model 2 | | | Model 3 | | |
|---|---|---|---|---|---|---|---|---|---|
| Term | OR | Bonferroni-corrected CI | | OR | Bonferroni-corrected CI | | OR | Bonferroni-corrected CI | |
| **Household income**[1] | | | | | | | | | |
| Very low | 0.4830 | 0.4670 | 0.4997 | 0.4399 | 0.4248 | 0.4555 | 0.6192 | 0.5960 | 0.6432 |
| Low | 0.8127 | 0.7873 | 0.8389 | 0.7668 | 0.7425 | 0.7919 | 0.8512 | 0.8234 | 0.8799 |
| Middle | Ref | – | – | Ref | – | – | Ref | – | – |
| High | 1.3055 | 1.2628 | 1.3497 | 1.3572 | 1.3125 | 1.4034 | 1.2102 | 1.1691 | 1.2526 |
| Very high | 1.9695 | 1.8690 | 2.0754 | 2.0726 | 1.9663 | 2.1846 | 1.6364 | 1.5491 | 1.7286 |
| **Sex** | | | | | | | | | |
| Female | Ref | – | – | Ref | – | – | Ref | – | – |
| Male | 0.8049 | 0.7867 | 0.8236 | – | – | – | 0.7836 | 0.7648 | 0.8030 |
| **Age** | 0.9954 | 0.9940 | 0.9968 | – | – | – | 1.0113 | 1.0097 | 1.0130 |
| **Multiple deprivation** | 0.9771 | 0.9762 | 0.9779 | 0.9768 | 0.9760 | 0.9777 | 0.9923 | 0.9913 | 0.9933 |
| **Ethnicity** | | | | | | | | | |
| White | Ref | – | – | Ref | – | – | Ref | – | – |
| Mixed-race | 0.8044 | 0.6916 | 0.9360 | 0.7644 | 0.6570 | 0.8898 | 0.9019 | 0.7729 | 1.0527 |
| Asian | 0.5267 | 0.4811 | 0.5768 | 0.5270 | 0.4813 | 0.5773 | 0.5995 | 0.5445 | 0.6602 |
| Black | 0.6258 | 0.5683 | 0.6893 | 0.6025 | 0.5469 | 0.6639 | 1.0896 | 0.9839 | 1.2069 |
| Chinese | 0.8293 | 0.6673 | 1.0318 | 0.7950 | 0.6396 | 0.9894 | 0.6783 | 0.5413 | 0.8506 |
| Other | 0.6770 | 0.5922 | 0.7745 | 0.6604 | 0.5774 | 0.7557 | 0.8903 | 0.7746 | 1.0235 |
| **Highest qualification** | | | | | | | | | |
| None | Ref | – | – | Ref | – | – | Ref | – | – |
| O levels/GCSEs/CSEs | 1.5682 | 1.5094 | 1.6292 | 1.5801 | 1.5193 | 1.6434 | 1.2420 | 1.1920 | 1.2941 |
| A levels/NVQ/HND/HNC[2] | 1.7160 | 1.6499 | 1.7849 | 1.7435 | 1.6753 | 1.8146 | 1.2876 | 1.2345 | 1.3429 |
| Degree | 2.5396 | 2.4468 | 2.6360 | 2.5944 | 2.4968 | 2.6958 | 1.5031 | 1.4412 | 1.5676 |

*Note:* Bonferroni-adjusted (~99.9%) confidence intervals. OR = odds ratio; CI = confidence interval; GCSEs = general certificate of secondary education; CSE = certificate of secondary education; NVQ = national vocational qualification; HND = higher national diploma; HNC = higher national certificate. For categorical explanatory variables the odds ratios indicate the changes in odds of reporting better self-rated health associated with the explanatory variable group relative to the reference group. Odds ratios for continuous explanatory variables indicate proportional odds ratios for a 1-unit increase in the explanatory variable on level of self-rated health. [1]Annual household income groups: very low (<£18 000), low (£18 000–30 999), middle (£31 000–51 999), high (£52 000–100 000) and very high (>£100 000). [2]also includes 'other professional qualifications'.
Model 1 – only individual explanatory variables.
Model 2 – adjusted for age and sex.
Model 3 – all explanatory variables.

Supplement e9-B. **Psychosocial factors associated with self-rated health**

| | Model 1 | | | Model 2 | | | Model 3 | | |
|---|---|---|---|---|---|---|---|---|---|
| Term | OR | Bonferroni-corrected CI | | OR | Bonferroni-corrected CI | | OR | Bonferroni-corrected CI | |
| **Loneliness** | | | | | | | | | |
| Not lonely | Ref | – | – | Ref | – | – | Ref | – | – |
| Lonely | 0.3590 | 0.3419 | 0.3769 | 0.3589 | 0.3419 | 0.3769 | 0.4921 | 0.4678 | 0.5176 |
| **Social isolation** | | | | | | | | | |
| Not isolated | Ref | – | – | Ref | – | – | Ref | – | – |
| Isolated | 0.5374 | 0.5159 | 0.5598 | 0.5394 | 0.5178 | 0.5619 | 0.8593 | 0.8233 | 0.8970 |

*Note:* Bonferroni-adjusted (~99.9%) confidence intervals. OR = odds ratio; CI = confidence interval. For categorical explanatory variables the odds ratios indicate the changes in odds of reporting better self-rated health associated with the explanatory variable group relative to the reference group.
Model 1 – only individual explanatory variables.
Model 2 – adjusted for age and sex.
Model 3 – all explanatory variables.

| Supplement e9-C. **Lifestyle factors associated with self-rated health** | | | | | | | | | |
|---|---|---|---|---|---|---|---|---|---|
| | Model 1 | | | Model 2 | | | Model 3 | | |
| Term | OR | Bonferroni-corrected CI | | OR | Bonferroni-corrected CI | | OR | Bonferroni-corrected CI | |
| **Sleep duration** (hours/day) | 1.0959 | 1.0836 | 1.1082 | 1.0958 | 1.0836 | 1.1082 | 1.0730 | 1.0609 | 1.0852 |
| **Physical activity** (days/week)[1] | | | | | | | | | |
| Walking | 1.1044 | 1.0979 | 1.1110 | 1.1044 | 1.0978 | 1.1110 | 1.0522 | 1.0454 | 1.0591 |
| Moderate activity | 1.1009 | 1.0954 | 1.1064 | 1.1029 | 1.0974 | 1.1085 | 1.0132 | 1.0070 | 1.0195 |
| Vigorous activity | 1.2033 | 1.1960 | 1.2106 | 1.2126 | 1.2052 | 1.2200 | 1.1668 | 1.1585 | 1.1751 |
| **Stair climbing frequency** | | | | | | | | | |
| None | Ref | – | – | Ref | – | – | Ref | – | – |
| 1-5/day | 1.0734 | 1.0225 | 1.1269 | 1.0717 | 1.0205 | 1.1254 | 1.0533 | 1.0019 | 1.1074 |
| 6-10/day | 1.5855 | 1.5157 | 1.6586 | 1.5775 | 1.5077 | 1.6506 | 1.2293 | 1.1733 | 1.2878 |
| 11-15/day | 1.8892 | 1.7997 | 1.9831 | 1.8694 | 1.7804 | 1.9628 | 1.2892 | 1.2260 | 1.3557 |
| 16-20/day | 2.0255 | 1.9150 | 2.1424 | 1.9988 | 1.8893 | 2.1146 | 1.2991 | 1.2258 | 1.3767 |
| 20+/day | 2.1944 | 2.0663 | 2.3305 | 2.1546 | 2.0282 | 2.2889 | 1.3652 | 1.2828 | 1.4529 |
| **Alcohol intake frequency** | | | | | | | | | |
| Never | 0.5340 | 0.5078 | 0.5615 | 0.5201 | 0.4945 | 0.5470 | 0.6624 | 0.6285 | 0.6981 |
| Special occasions | 0.6048 | 0.5799 | 0.6309 | 0.5743 | 0.5504 | 0.5992 | 0.7565 | 0.7240 | 0.7904 |
| 1-3/month | 0.8268 | 0.7934 | 0.8616 | 0.8007 | 0.7683 | 0.8345 | 0.9201 | 0.8821 | 0.9597 |
| 1-2/week | Ref | – | – | Ref | – | – | Ref | – | – |
| 3-4/week | 1.2045 | 1.1663 | 1.2441 | 1.2297 | 1.1906 | 1.2701 | 1.0520 | 1.0178 | 1.0875 |
| Daily/almost daily | 1.1495 | 1.1119 | 1.1884 | 1.2013 | 1.1617 | 1.2423 | 1.0264 | 0.9913 | 1.0628 |
| **BMI** ($kg/m^2$) | 0.8850 | 0.8827 | 0.8873 | 0.8860 | 0.8838 | 0.8883 | 0.9045 | 0.9020 | 0.9069 |
| **Smoking status** | | | | | | | | | |
| Never | Ref | – | – | Ref | – | – | Ref | – | – |
| Former | 0.7546 | 0.7361 | 0.7735 | 0.7741 | 0.7549 | 0.7938 | 0.8128 | 0.7919 | 0.8342 |
| Current | 0.4133 | 0.3973 | 0.4298 | 0.4192 | 0.4030 | 0.4361 | 0.5029 | 0.4825 | 0.5242 |

*Note:* Bonferroni-adjusted (~99.9%) confidence intervals. OR = odds ratio; CI = confidence interval; BMI = body mass index. For categorical explanatory variables the odds ratios indicate the changes in odds of reporting better self-rated health associated with the explanatory variable group relative to the reference group. Odds ratios for continuous explanatory variables indicate proportional odds ratios for a 1-unit increase in the explanatory variable on level of self-rated health. [1]number of days per week engaging in these activities for 10+ minutes continuously.
Model 1 – only individual explanatory variables.
Model 2 – adjusted for age and sex.
Model 3 – all explanatory variables.

Supplement e9-D. **Environmental exposures associated with self-rated health**

| | Model 1 | | | Model 2 | | | Model 3 | | |
|---|---|---|---|---|---|---|---|---|---|
| Term | OR | Bonferroni-corrected CI | | OR | Bonferroni-corrected CI | | OR | Bonferroni-corrected CI | |
| **$PM_{2.5}$** | 0.8860 | 0.8762 | 0.8958 | 0.8829 | 0.8732 | 0.8927 | 1.0283 | 1.0047 | 1.0525 |
| **$PM_{10}$** | 0.9628 | 0.9570 | 0.9687 | 0.9621 | 0.9562 | 0.9679 | 0.9979 | 0.9904 | 1.0055 |
| **$NO_2$** | 0.9843 | 0.9829 | 0.9858 | 0.9838 | 0.9823 | 0.9853 | 0.9948 | 0.9910 | 0.9986 |
| **$L_{den}$** | 0.9951 | 0.9924 | 0.9977 | 0.9950 | 0.9923 | 0.9976 | 1.0025 | 0.9993 | 1.0058 |
| **Greenspace** 1000m | 1.0039 | 1.0034 | 1.0044 | 1.0041 | 1.0035 | 1.0046 | 1.0007 | 0.9998 | 1.0015 |

*Note:* Bonferroni-adjusted (~99.9%) confidence intervals. OR = odds ratio; CI = confidence interval; PM = particulate matter; $NO_2$ = nitrogen dioxide; $L_{den}$ = day-evening-night noise level. Odds ratios indicate proportional odds ratios for a 1-unit increase in the explanatory variable on level of self-rated health.
Model 1 – only individual explanatory variables.
Model 2 – adjusted for age and sex.
Model 3 – all explanatory variables.

**Supplement e10. Regression tables health indicators**

Sociodemographic characteristics

Supplement e10-A. **Sociodemographic characteristics associated with health indicators**

| | Health status | | | Long-standing illness | | | Self-rated health | | |
|---|---|---|---|---|---|---|---|---|---|
| Term | OR | Bonferroni-corrected CI | | OR | Bonferroni-corrected CI | | OR[3] | Bonferroni-corrected CI | |
| **Household income**[1] | | | | | | | | | |
| Very low | 0.7447 | 0.7148 | 0.7759 | 0.6654 | 0.6384 | 0.6934 | 0.6192 | 0.5960 | 0.6432 |
| Low | 0.9196 | 0.8864 | 0.9541 | 0.8906 | 0.8581 | 0.9244 | 0.8512 | 0.8234 | 0.8799 |
| Middle | Ref | – | – | Ref | – | – | Ref | – | – |
| High | 1.0510 | 1.0096 | 1.0941 | 1.1171 | 1.0726 | 1.1634 | 1.2102 | 1.1691 | 1.2526 |
| Very high | 1.0472 | 0.9812 | 1.1181 | 1.2524 | 1.1703 | 1.3410 | 1.6364 | 1.5491 | 1.7286 |
| **Sex** | | | | | | | | | |
| Female | Ref | – | – | Ref | – | – | Ref | – | – |
| Male | 0.8847 | 0.8610 | 0.9090 | 0.7585 | 0.7380 | 0.7797 | 0.7836 | 0.7648 | 0.8030 |
| **Age** | 0.9528 | 0.9510 | 0.9546 | 0.9730 | 0.9711 | 0.9748 | 1.0113 | 1.0097 | 1.0130 |
| **Multiple deprivation** | 0.9952 | 0.9941 | 0.9963 | 0.9921 | 0.9910 | 0.9932 | 0.9923 | 0.9913 | 0.9933 |
| **Ethnicity** | | | | | | | | | |
| White | Ref | – | – | Ref | – | – | Ref | – | – |
| Mixed-race | 1.1037 | 0.9221 | 1.3273 | 1.1223 | 0.9387 | 1.3474 | 0.9019 | 0.7729 | 1.0527 |
| Asian | 1.0188 | 0.9149 | 1.1359 | 1.2225 | 1.0962 | 1.3652 | 0.5995 | 0.5445 | 0.6602 |
| Black | 1.3125 | 1.1657 | 1.4805 | 1.1852 | 1.0591 | 1.3277 | 1.0896 | 0.9839 | 1.2069 |
| Chinese | 1.8339 | 1.3646 | 2.5106 | 1.6205 | 1.2207 | 2.1842 | 0.6783 | 0.5413 | 0.8506 |
| Other | 1.2570 | 1.0727 | 1.4779 | 1.2027 | 1.0301 | 1.4078 | 0.8903 | 0.7746 | 1.0235 |
| **Highest qualification** | | | | | | | | | |
| None | Ref | – | – | Ref | – | – | Ref | – | – |
| O levels/GCSEs/CSEs | 1.0315 | 0.9870 | 1.0779 | 0.9920 | 0.9485 | 1.0375 | 1.2420 | 1.1920 | 1.2941 |
| A levels/NVQ/HND/HNC[2] | 0.9879 | 0.9445 | 1.0332 | 0.8953 | 0.8554 | 0.9371 | 1.2876 | 1.2345 | 1.3429 |
| Degree | 0.9558 | 0.9137 | 0.9997 | 0.8550 | 0.8167 | 0.8950 | 1.5031 | 1.4412 | 1.5676 |

*Note:* Estimates from Model 3 (i.e. including all explanatory variables). Bonferroni-adjusted (~99.9%) confidence intervals. OR = odds ratio; CI = confidence interval; GCSEs = general certificate of secondary education; CSE = certificate of secondary education; NVQ = national vocational qualification; HND = higher national diploma; HNC = higher national certificate. [1]Annual household income groups: very low (<£18 000), low (£18 000–30 999), middle (£31 000–51 999), high (£52 000–100 000) and very high (>£100 000). [2]also includes 'other professional qualifications'. [3]For categorical explanatory variables the odds ratios indicate the changes in odds of reporting better self-rated health associated with the explanatory variable group relative to the reference group. Odds ratios for continuous explanatory variables indicate proportional odds ratios for a 1-unit increase in the explanatory variable on level of self-rated health.

| Supplement e10-B. **Psychosocial factors associated with health indicators** | | | | | | | | | |
|---|---|---|---|---|---|---|---|---|---|
| | Health status | | | Long-standing illness | | | Self-rated health | | |
| Term | OR | Bonferroni-corrected CI | | OR | Bonferroni-corrected CI | | OR[1] | Bonferroni-corrected CI | |
| **Loneliness** | | | | | | | | | |
| Not lonely | Ref | – | – | Ref | – | – | Ref | – | – |
| Lonely | 0.8129 | 0.7697 | 0.8587 | 0.7097 | 0.6722 | 0.7494 | 0.4921 | 0.4678 | 0.5176 |
| **Social isolation** | | | | | | | | | |
| Not isolated | Ref | – | – | Ref | – | – | Ref | – | – |
| Isolated | 0.9496 | 0.9067 | 0.9947 | 0.9185 | 0.8770 | 0.9621 | 0.8593 | 0.8233 | 0.8970 |

*Note:* Estimates from Model 3 (i.e. including all explanatory variables). Bonferroni-adjusted (~99.9%) confidence intervals. OR = odds ratio; CI = confidence interval. [1]For categorical explanatory variables the odds ratios indicate the changes in odds of reporting better self-rated health associated with the explanatory variable group relative to the reference group.

Supplement e10-C. **Lifestyle factors associated with health indicators**

| | Health status | | | Long-standing illness | | | Self-rated health | | |
|---|---|---|---|---|---|---|---|---|---|
| Term | OR | Bonferroni-corrected CI | | OR | Bonferroni-corrected CI | | OR[2] | Bonferroni-corrected CI | |
| **Sleep duration** (hours/day) | 0.9748 | 0.9631 | 0.9868 | 1.0004 | 0.9882 | 1.0128 | 1.0730 | 1.0609 | 1.0852 |
| **Physical activity** (days/week)[1] | | | | | | | | | |
| Walking | 1.0105 | 1.0032 | 1.0178 | 1.0250 | 1.0175 | 1.0325 | 1.0522 | 1.0454 | 1.0591 |
| Moderate activity | 1.0032 | 0.9964 | 1.0100 | 0.9984 | 0.9916 | 1.0053 | 1.0132 | 1.0070 | 1.0195 |
| Vigorous activity | 1.0319 | 1.0238 | 1.0400 | 1.0641 | 1.0557 | 1.0727 | 1.1668 | 1.1585 | 1.1751 |
| **Stair climbing frequency** | | | | | | | | | |
| None | Ref | – | – | Ref | – | – | Ref | – | – |
| 1-5/day | 1.0704 | 1.0148 | 1.1289 | 1.1101 | 1.0522 | 1.1710 | 1.0533 | 1.0019 | 1.1074 |
| 6-10/day | 1.1756 | 1.1188 | 1.2352 | 1.2937 | 1.2307 | 1.3598 | 1.2293 | 1.1733 | 1.2878 |
| 11-15/day | 1.1714 | 1.1098 | 1.2363 | 1.3131 | 1.2434 | 1.3866 | 1.2892 | 1.2260 | 1.3557 |
| 16-20/day | 1.1650 | 1.0937 | 1.2410 | 1.3066 | 1.2255 | 1.3932 | 1.2991 | 1.2258 | 1.3767 |
| 20+/day | 1.1881 | 1.1097 | 1.2723 | 1.2975 | 1.2108 | 1.3907 | 1.3652 | 1.2828 | 1.4529 |
| **Alcohol intake frequency** | | | | | | | | | |
| Never | 0.6285 | 0.5946 | 0.6644 | 0.5533 | 0.5234 | 0.5850 | 0.6624 | 0.6285 | 0.6981 |
| Special occasions | 0.7792 | 0.7429 | 0.8174 | 0.7142 | 0.6809 | 0.7492 | 0.7565 | 0.7240 | 0.7904 |
| 1-3/month | 0.9129 | 0.8708 | 0.9570 | 0.8486 | 0.8096 | 0.8895 | 0.9201 | 0.8821 | 0.9597 |
| 1-2/week | Ref | – | – | Ref | – | – | Ref | – | – |
| 3-4/week | 1.0590 | 1.0198 | 1.0998 | 1.0925 | 1.0514 | 1.1353 | 1.0520 | 1.0178 | 1.0875 |
| Daily/almost daily | 1.0334 | 0.9939 | 1.0745 | 1.0744 | 1.0326 | 1.1180 | 1.0264 | 0.9913 | 1.0628 |
| **BMI** ($kg/m^2$) | 0.9683 | 0.9656 | 0.9711 | 0.9419 | 0.9392 | 0.9446 | 0.9045 | 0.9020 | 0.9069 |
| **Smoking status** | | | | | | | | | |
| Never | Ref | – | – | Ref | – | – | Ref | – | – |
| Former | 0.7922 | 0.7698 | 0.8152 | 0.7978 | 0.7749 | 0.8213 | 0.8128 | 0.7919 | 0.8342 |
| Current | 0.7518 | 0.7183 | 0.7869 | 0.7858 | 0.7506 | 0.8227 | 0.5029 | 0.4825 | 0.5242 |

*Note:* Estimates from Model 3 (i.e. including all explanatory variables). Bonferroni-adjusted (~99.9%) confidence intervals. OR = odds ratio; CI = confidence interval; BMI = body mass index. [1]number of days per week engaging in these activities for 10+ minutes continuously. [2]For categorical explanatory variables the odds ratios indicate the changes in odds of reporting better self-rated health associated with the explanatory variable group relative to the reference group. Odds ratios for continuous explanatory variables indicate proportional odds ratios for a 1-unit increase in the explanatory variable on level of self-rated health.

Supplement e10-D. **Environmental exposures associated with health indicators**

| | Health status | | | Long-standing illness | | | Self-rated health | | |
|---|---|---|---|---|---|---|---|---|---|
| Term | OR | Bonferroni-corrected CI | | OR | Bonferroni-corrected CI | | OR[1] | Bonferroni-corrected CI | |
| **$PM_{2.5}$** | 0.9656 | 0.9411 | 0.9908 | 0.9677 | 0.9430 | 0.9932 | 1.0283 | 1.0047 | 1.0525 |
| **$PM_{10}$** | 1.0019 | 0.9935 | 1.0104 | 1.0019 | 0.9934 | 1.0105 | 0.9979 | 0.9904 | 1.0055 |
| **$NO_2$** | 1.0036 | 0.9993 | 1.0078 | 1.0025 | 0.9982 | 1.0068 | 0.9948 | 0.9910 | 0.9986 |
| **$L_{den}$** | 0.9981 | 0.9945 | 1.0017 | 1.0015 | 0.9979 | 1.0052 | 1.0025 | 0.9993 | 1.0058 |
| **Greenspace** 1000m | 0.9995 | 0.9985 | 1.0004 | 0.9997 | 0.9987 | 1.0007 | 1.0007 | 0.9998 | 1.0015 |

*Note:* Estimates from Model 3 (i.e. including all explanatory variables). Bonferroni-adjusted (~99.9%) confidence intervals. OR = odds ratio; CI = confidence interval; PM = particulate matter; $NO_2$ = nitrogen dioxide; $L_{den}$ = day-evening-night noise level. [1]Odds ratios indicate proportional odds ratios for a 1-unit increase in the explanatory variable on level of self-rated health.

**Supplement e11. Standardised regression coefficients - tables**

Health status

| Supplement e11-A. **Standardised regression coefficients health status** | | | |
|---|---|---|---|
| Term | *β* | Bonferroni-corrected CI | |
| Sociodemographic characteristics | | | |
| **Household income**[1] | | | |
| Very low | -0.2948 | -0.3358 | -0.2538 |
| Low | -0.0838 | -0.1206 | -0.047 |
| Middle | Ref | – | – |
| High | 0.0497 | 0.0095 | 0.0899 |
| Very high | 0.0461 | -0.019 | 0.1116 |
| **Sex** | | | |
| Female | Ref | – | – |
| Male | -0.1225 | -0.1496 | -0.0954 |
| **Age** | -0.7801 | -0.811 | -0.7493 |
| **Multiple deprivation** | -0.1295 | -0.1591 | -0.0998 |
| **Ethnicity** | | | |
| White | Ref | – | – |
| Mixed-race | 0.0987 | -0.0811 | 0.2831 |
| Asian | 0.0187 | -0.0889 | 0.1274 |
| Black | 0.2719 | 0.1533 | 0.3924 |
| Chinese | 0.6065 | 0.3109 | 0.9205 |
| Other | 0.2287 | 0.0702 | 0.3906 |
| **Highest qualification** | | | |
| None | Ref | – | – |
| O levels/GCSEs/CSEs | 0.031 | -0.0131 | 0.075 |
| A levels/NVQ/HND/HNC[2] | -0.0122 | -0.0571 | 0.0326 |
| Degree | -0.0453 | -0.0902 | -0.0003 |
| Psychosocial factors | | | |
| **Loneliness** | | | |
| Not lonely | Ref | – | – |
| Lonely | -0.2072 | -0.2618 | -0.1524 |
| **Social isolation** | | | |
| Not isolated | Ref | – | – |
| Isolated | -0.0517 | -0.098 | -0.0053 |
| Lifestyle factors | | | |
| **Sleep duration** (hours/day) | -0.0538 | -0.0794 | -0.0281 |
| **Physical activity** (days/week)[3] | | | |
| Walking | 0.0406 | 0.0124 | 0.0687 |
| Moderate activity | 0.0147 | -0.0168 | 0.0462 |
| Vigorous activity | 0.1218 | 0.0914 | 0.1523 |
| **Stair climbing frequency** | | | |
| None | Ref | – | – |
| 1-5/day | 0.068 | 0.0147 | 0.1213 |
| 6-10/day | 0.1618 | 0.1122 | 0.2113 |
| 11-15/day | 0.1581 | 0.1042 | 0.2121 |
| 16-20/day | 0.1527 | 0.0896 | 0.2159 |
| 20+/day | 0.1724 | 0.1041 | 0.2408 |
| **Alcohol intake frequency** | | | |
| Never | -0.4644 | -0.5198 | -0.4089 |
| Special occasions | -0.2494 | -0.2972 | -0.2016 |
| 1-3/month | -0.0911 | -0.1384 | -0.0439 |
| 1-2/week | Ref | – | – |
| 3-4/week | 0.0574 | 0.0196 | 0.0952 |
| Daily/almost daily | 0.0329 | -0.0062 | 0.0718 |
| **BMI** ($kg/m^2$) | -0.3001 | -0.3267 | -0.2735 |
| **Smoking status** | | | |
| Never | Ref | – | – |
| Former | -0.233 | -0.2616 | -0.2043 |
| Current | -0.2853 | -0.3309 | -0.2396 |
| Environmental exposures | | | |
| **$PM_{2.5}$** | -0.0728 | -0.1261 | -0.0193 |
| **$PM_{10}$** | 0.007 | -0.0246 | 0.0388 |
| **$NO_2$** | 0.0538 | -0.0103 | 0.1177 |
| **$L_{den}$** | -0.0162 | -0.0466 | 0.0143 |
| **Greenspace** 1000m | -0.023 | -0.0652 | 0.0192 |

*Note: β* = Model 3 regression coefficients rescaled to have a mean equal to zero and, for numeric variables with more than two values, divided by two standard deviations. Bonferroni-adjusted (~99.9%) confidence intervals. CI = confidence interval; GCSEs = general certificate of secondary education; CSE = certificate of secondary education; NVQ = national vocational qualification; HND = higher national diploma; HNC = higher national certificate; BMI = body mass index; PM = particulate matter; $NO_2$ = nitrogen dioxide; $L_{den}$ = day-evening-night noise level. [1]Annual household income groups: very low (<£18 000), low (£18 000–30 999), middle (£31 000–51 999), high (£52 000–100 000) and very high (>£100 000). [2]also includes 'other professional qualifications'. [3]number of days per week engaging in these activities for 10+ minutes continuously.

Long-standing illness

Supplement e11-B. **Standardised regression coefficients for long-standing illness**

| Term | $\beta$ | Bonferroni-adjusted CI | |
|---|---|---|---|
| Sociodemographic characteristics | | | |
| **Household income**[1] | | | |
| Very low | -0.4074 | -0.4487 | -0.3661 |
| Low | -0.1158 | -0.153 | -0.0786 |
| Middle | Ref | – | – |
| High | 0.1107 | 0.0701 | 0.1514 |
| Very high | 0.2251 | 0.1573 | 0.2934 |
| **Sex** | | | |
| Female | Ref | – | – |
| Male | -0.2763 | -0.3038 | -0.2489 |
| **Age** | -0.4423 | -0.4729 | -0.4117 |
| **Multiple deprivation** | -0.2125 | -0.2421 | -0.1828 |
| **Ethnicity** | | | |
| White | Ref | – | – |
| Mixed-race | 0.1154 | -0.0632 | 0.2982 |
| Asian | 0.2009 | 0.0918 | 0.3113 |
| Black | 0.1699 | 0.0574 | 0.2835 |
| Chinese | 0.4827 | 0.1994 | 0.7812 |
| Other | 0.1846 | 0.0297 | 0.3421 |
| **Highest qualification** | | | |
| None | Ref | – | – |
| O levels/GCSEs/CSEs | -0.008 | -0.0529 | 0.0368 |
| A levels/NVQ/HND/HNC[2] | -0.1106 | -0.1562 | -0.0649 |
| Degree | -0.1567 | -0.2025 | -0.1109 |
| Psychosocial factors | | | |
| **Loneliness** | | | |
| Not lonely | Ref | – | – |
| Lonely | -0.3429 | -0.3971 | -0.2885 |
| **Social isolation** | | | |
| Not isolated | Ref | – | – |
| Isolated | -0.085 | -0.1313 | -0.0387 |
| Lifestyle factors | | | |
| **Sleep duration** (hours/day) | 0.0009 | -0.025 | 0.0268 |
| **Physical activity** (days/week)[3] | | | |
| Walking | 0.096 | 0.0677 | 0.1243 |
| Moderate activity | -0.0074 | -0.0393 | 0.0245 |
| Vigorous activity | 0.2413 | 0.2103 | 0.2724 |
| **Stair climbing frequency** | | | |
| None | Ref | – | – |
| 1-5/day | 0.1044 | 0.0509 | 0.1579 |
| 6-10/day | 0.2575 | 0.2076 | 0.3073 |
| 11-15/day | 0.2724 | 0.2179 | 0.3269 |
| 16-20/day | 0.2675 | 0.2034 | 0.3316 |
| 20+/day | 0.2605 | 0.1913 | 0.3298 |
| **Alcohol intake frequency** | | | |
| Never | -0.5918 | -0.6474 | -0.5361 |
| Special occasions | -0.3366 | -0.3844 | -0.2888 |
| 1-3/month | -0.1642 | -0.2113 | -0.1171 |
| 1-2/week | Ref | – | – |
| 3-4/week | 0.0885 | 0.0501 | 0.1269 |
| Daily/almost daily | 0.0718 | 0.032 | 0.1115 |
| **BMI** ($kg/m^2$) | -0.5586 | -0.5855 | -0.5317 |
| **Smoking status** | | | |
| Never | Ref | – | – |
| Former | -0.2259 | -0.255 | -0.1969 |
| Current | -0.2411 | -0.2868 | -0.1952 |
| Environmental exposures | | | |
| **$PM_{2.5}$** | -0.0682 | -0.1221 | -0.0142 |
| **$PM_{10}$** | 0.0072 | -0.0249 | 0.0393 |
| **$NO_2$** | 0.0381 | -0.0266 | 0.1029 |
| **$L_{den}$** | 0.0127 | -0.0181 | 0.0436 |
| **Greenspace** 1000m | -0.0133 | -0.0562 | 0.0295 |

*Note:* $\beta$ = Model 3 regression coefficients rescaled to have a mean equal to zero and, for numeric variables with more than two values, divided by two standard deviations. Bonferroni-adjusted (~99.9%) confidence intervals. CI = confidence interval; GCSEs = general certificate of secondary education; CSE = certificate of secondary education; NVQ = national vocational qualification; HND = higher national diploma; HNC = higher national certificate; BMI = body mass index; PM = particulate matter; $NO_2$ = nitrogen dioxide; $L_{den}$ = day-evening-night noise level. [1]Annual household income groups: very low (<£18 000), low (£18 000–30 999), middle (£31 000–51 999), high (£52 000–100 000) and very high (>£100 000). [2]also includes 'other professional qualifications'. [3]number of days per week engaging in these activities for 10+ minutes continuously.

Self-rated health

Supplement e11-C. **Standardised regression coefficients for self-rated health**

| Term | *β* | Bonferroni-corrected CI | |
|---|---|---|---|
| Sociodemographic characteristics | | | |
| **Household income**[1] | | | |
| Very low | -0.4794 | -0.5174 | -0.4412 |
| Low | -0.1611 | -0.1943 | -0.128 |
| Middle | Ref | – | – |
| High | 0.1908 | 0.1563 | 0.2252 |
| Very high | 0.4925 | 0.444 | 0.5477 |
| **Sex** | | | |
| Female | Ref | – | – |
| Male | -0.2438 | -0.2682 | -0.2194 |
| **Age** | 0.1819 | 0.1552 | 0.2086 |
| **Multiple deprivation** | -0.2054 | -0.2325 | -0.1783 |
| **Ethnicity** | | | |
| White | Ref | – | – |
| Mixed-race | -0.1034 | -0.2572 | 0.0508 |
| Asian | -0.5116 | -0.6077 | -0.4153 |
| Black | 0.0858 | -0.016 | 0.1879 |
| Chinese | -0.3881 | -0.6128 | -0.163 |
| Other | -0.1163 | -0.2553 | 0.0229 |
| **Highest qualification** | | | |
| None | Ref | – | – |
| O levels/GCSEs/CSEs | 0.2167 | 0.1761 | 0.2578 |
| A levels/NVQ/HND/HNC[2] | 0.2528 | 0.211 | 0.2947 |
| Degree | 0.4075 | 0.3655 | 0.4495 |
| Psychosocial factors | | | |
| **Loneliness** | | | |
| Not lonely | Ref | – | – |
| Lonely | -0.7092 | -0.7596 | -0.6587 |
| **Social isolation** | | | |
| Not isolated | Ref | – | – |
| Isolated | -0.1516 | -0.1944 | -0.1087 |
| Lifestyle factors | | | |
| **Sleep duration** (hours/day) | 0.1487 | 0.1248 | 0.1726 |
| **Physical activity** (days/week)[3] | | | |
| Walking | 0.198 | 0.1726 | 0.2234 |
| Moderate activity | 0.061 | 0.0326 | 0.0895 |
| Vigorous activity | 0.5988 | 0.5711 | 0.6266 |
| **Stair climbing frequency** | | | |
| None | Ref | – | – |
| 1-5/day | 0.052 | 0.0019 | 0.102 |
| 6-10/day | 0.2064 | 0.1599 | 0.2529 |
| 11-15/day | 0.2541 | 0.2037 | 0.3043 |
| 16-20/day | 0.2617 | 0.2037 | 0.3196 |
| 20+/day | 0.3113 | 0.2489 | 0.3738 |
| **Alcohol intake frequency** | | | |
| Never | -0.4119 | -0.4631 | -0.3607 |
| Special occasions | -0.2791 | -0.3229 | -0.2352 |
| 1-3/month | -0.0833 | -0.1254 | -0.0411 |
| 1-2/week | Ref | – | – |
| 3-4/week | 0.0507 | 0.0177 | 0.0837 |
| Daily/almost daily | 0.0261 | -0.0093 | 0.0609 |
| **BMI** (kg/m$^2$) | -0.9368 | -0.9616 | -0.912 |
| **Smoking status** | | | |
| Never | Ref | – | – |
| Former | -0.2073 | -0.2332 | -0.1813 |
| Current | -0.6873 | -0.7287 | -0.6459 |
| Environmental exposures | | | |
| **$PM_{2.5}$** | 0.0581 | 0.01 | 0.1064 |
| **$PM_{10}$** | -0.0077 | -0.0359 | 0.0206 |
| **$NO_2$** | -0.0784 | -0.1362 | -0.0207 |
| **$L_{den}$** | 0.0214 | -0.0059 | 0.0487 |
| **Greenspace** 1000m | 0.0289 | -0.0089 | 0.0666 |

*Note: β* = Model 3 regression coefficients rescaled to have a mean equal to zero and, for numeric variables with more than two values, divided by two standard deviations. Bonferroni-adjusted (~99.9%) confidence intervals. CI = confidence interval; GCSEs = general certificate of secondary education; CSE = certificate of secondary education; NVQ = national vocational qualification; HND = higher national diploma; HNC = higher national certificate; BMI = body mass index; PM = particulate matter; $NO_2$ = nitrogen dioxide; $L_{den}$ = day-evening-night noise level. [1]Annual household income groups: very low (<£18 000), low (£18 000–30 999), middle (£31 000–51 999), high (£52 000–100 000) and very high (>£100 000). [2]also includes 'other professional qualifications'. [3]number of days per week engaging in these activities for 10+ minutes continuously.

**Supplement e12. Standardised regression coefficients – plots**

Long-standing illness

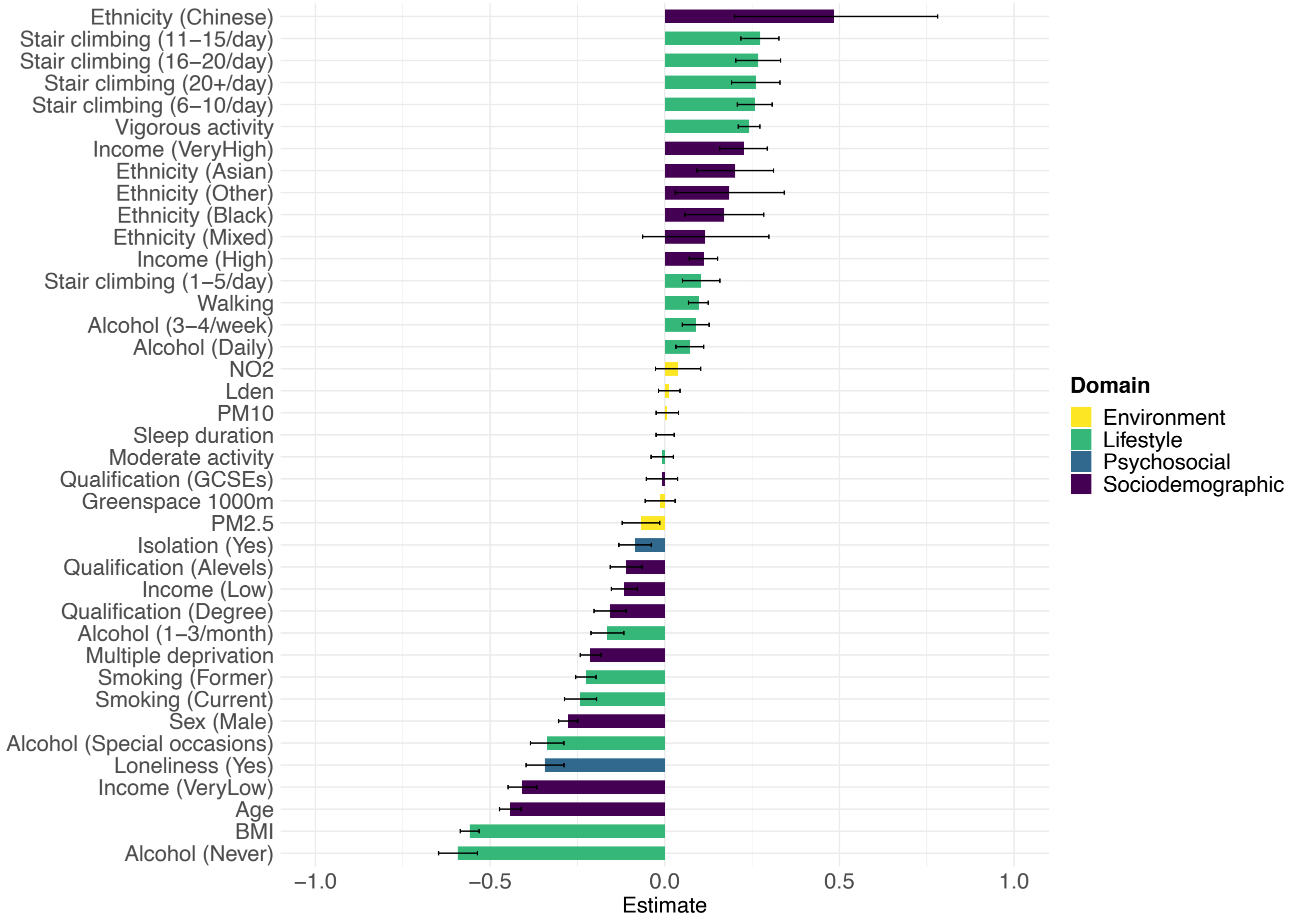


**Supplement e12-A.** Horizontal bar plot for long-standing illness. $\beta$ estimates and Bonferroni-adjusted (~99.9%) confidence intervals. Model 3 regression coefficients rescaled to have a mean equal to zero and, for numeric variables with more than two values, divided by two standard deviations.

Self-rated health

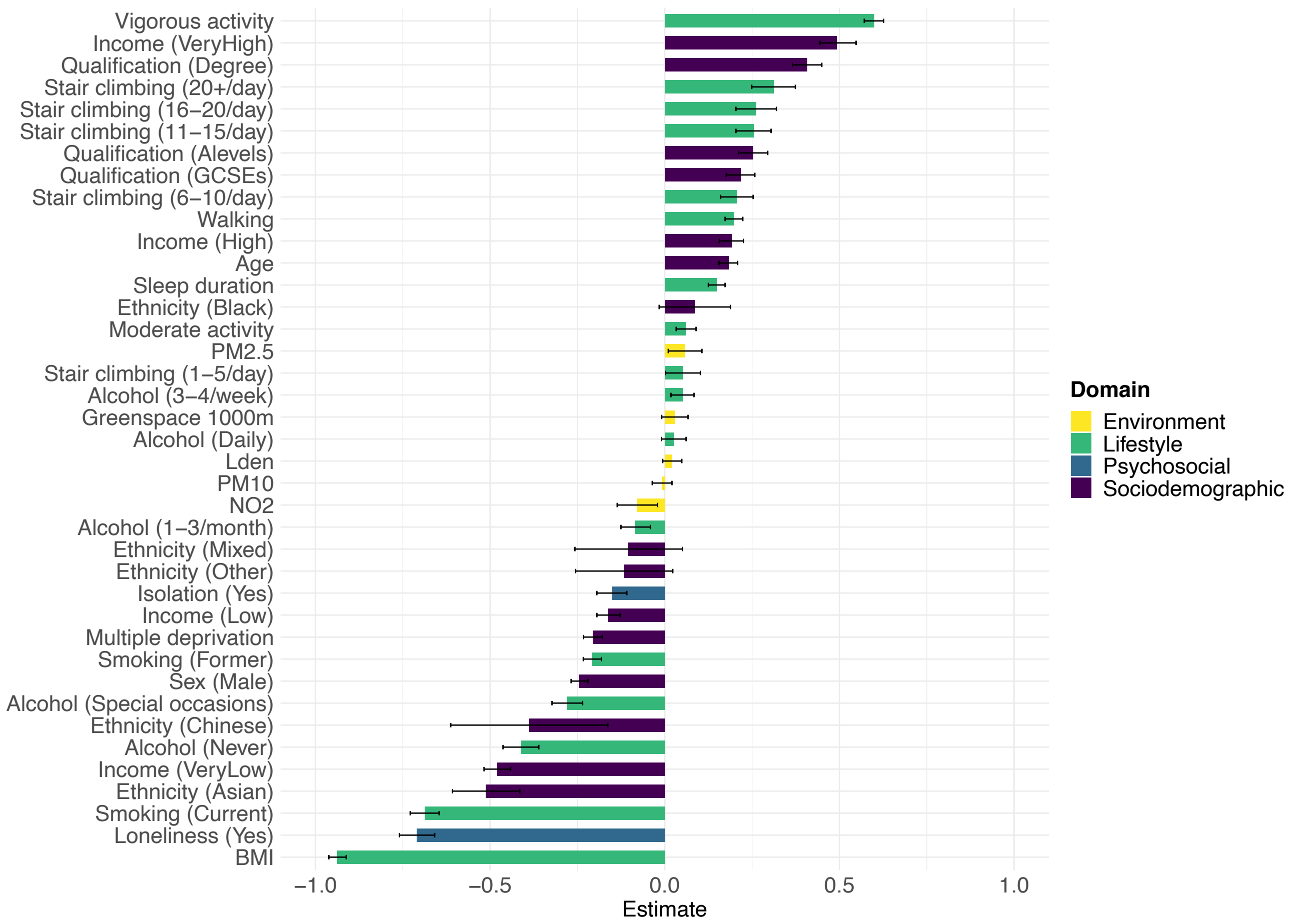


**Supplement e12-B.** Horizontal bar plot for self-rated health. $\beta$ estimates and Bonferroni-adjusted (~99.9%) confidence intervals. Model 3 regression coefficients rescaled to have a mean equal to zero and, for numeric variables with more than two values, divided by two standard deviations.

**Supplement e13. Baseline characteristics stratified by sex**

Table e13. **Baseline characteristics stratified by sex**

| | Female (*n* = 159 574) | Male (*n* = 147 804) | p value |
|---|---|---|---|
| Health indicators | | | |
| **Health status** | | | < 0.001 |
| Unhealthy | 47 016 (29.5%) | 48 161 (32.6%) | |
| Healthy | 112 558 (70.5%) | 99 643 (67.4%) | |
| **Long-standing illness** | | | < 0.001 |
| Yes | 44 607 (28.0%) | 49 150 (33.3%) | |
| No | 114 967 (72.0%) | 98 654 (66.7%) | |
| **Self-rated health** | | | < 0.001 |
| Poor | 4 991 (3.1%) | 6 075 (4.1%) | |
| Fair | 27 657 (17.3%) | 31 512 (21.3%) | |
| Good | 97 197 (60.9%) | 85 502 (57.8%) | |
| Excellent | 29 729 (18.6%) | 24 715 (16.7%) | |
| Sociodemographic characteristics | | | |
| **Age** | | | < 0.001 |
| Mean (SD) | 55.68 (7.96) | 56.56 (8.16) | |
| Range | 39.00-71.00 | 38.00-73.00 | |
| **Ethnicity** | | | < 0.001 |
| White | 152 293 (95.4%) | 141 272 (95.6%) | |
| Mixed-race | 1 105 (0.7%) | 661 (0.4%) | |
| Black | 2 445 (1.5%) | 1 812 (1.2%) | |
| Asian | 2 016 (1.3%) | 2 739 (1.9%) | |
| Chinese | 505 (0.3%) | 313 (0.2%) | |
| Other | 1 210 (0.8%) | 1 007 (0.7%) | |
| **Highest qualification** | | | < 0.001 |
| None | 19 497 (12.2%) | 20 331 (13.8%) | |
| O levels/GCSEs/CSEs | 47 467 (29.7%) | 36 981 (25.0%) | |
| A levels/NVQ/HND/HNC[1] | 36 545 (22.9%) | 36 039 (24.4%) | |
| Degree | 56 065 (35.1%) | 54 453 (36.8%) | |
| **Household income**[2] | | | < 0.001 |
| Very low | 41 713 (26.1%) | 40 625 (27.5%) | |
| Low | 35 968 (22.5%) | 27 131 (18.4%) | |
| Medium | 42 060 (26.4%) | 35 871 (24.3%) | |
| High | 31 507 (19.7%) | 34 599 (23.4%) | |
| Very high | 8 326 (5.2%) | 9 578 (6.5%) | |
| **Multiple deprivation** | | | < 0.001 |
| Mean (SD) | 16.65 (13.13) | 16.90 (13.54) | |
| Range | 0.61-82.00 | 0.61-82.00 | |
| Psychosocial factors | | | |
| **Loneliness** | | | < 0.001 |
| Not lonely | 150 898 (94.6%) | 139 003 (94.0%) | |
| Lonely | 8 676 (5.4%) | 8 801 (6.0%) | |
| **Social isolation** | | | < 0.001 |
| Not isolated | 146 366 (91.7%) | 134 565 (91.0%) | |
| Isolated | 13 208 (8.3%) | 13 239 (9.0%) | |
| Lifestyle factors | | | |
| **Smoking status** | | | < 0.001 |
| Never | 94 928 (59.5%) | 73 547 (49.8%) | |
| Former | 51 194 (32.1%) | 57 444 (38.9%) | |
| Current | 13 452 (8.4%) | 16 813 (11.4%) | |
| **Stair climbing frequency** | | | < 0.001 |
| None | 12 398 (7.8%) | 11 651 (7.9%) | |
| 1-5/day | 28 452 (17.8%) | 29 815 (20.2%) | |
| 6-10/day | 59 067 (37.0%) | 56 915 (38.5%) | |
| 11-15/day | 32 437 (20.3%) | 27 878 (18.9%) | |
| 16-20/day | 15 174 (9.5%) | 12 435 (8.4%) | |
| 20+/day | 12 046 (7.5%) | 9 110 (6.2%) | |
| **Alcohol intake frequency** | | | < 0.001 |
| Never | 41 206 (25.8%) | 37 571 (25.4%) | |
| Special occasions | 12 551 (7.9%) | 7 872 (5.3%) | |
| 1-3/month | 21 911 (13.7%) | 9 615 (6.5%) | |
| 1-2/week | 20 913 (13.1%) | 12 885 (8.7%) | |
| 3-4/week | 35 098 (22.0%) | 40 153 (27.2%) | |
| Daily/almost daily | 27 895 (17.5%) | 39 708 (26.9%) | |
| **Sleep duration** (hours/day) | | | < 0.001 |
| Mean (SD) | 7.18 (1.06) | 7.13 (1.04) | |
| Range | 1.00-20.00 | 1.00-20.00 | |
| **BMI** ($kg/m^2$) | | | < 0.001 |
| Mean (SD) | 26.85 (5.07) | 27.72 (4.13) | |
| Range | 12.80-67.30 | 12.80-63.40 | |
| **Walking** (days/week)[3] | | | < 0.001 |

| | | | |
|---|---|---|---|
| Mean (SD) | 5.43 (1.89) | 5.29 (2.00) | |
| Range | 0.00-7.00 | 0.00-7.00 | |
| **Moderate activity** (days/week)[3] | | | 0.352 |
| Mean (SD) | 3.59 (2.32) | 3.58 (2.32) | |
| Range | 0.00-7.00 | 0.00-7.00 | |
| **Vigorous activity** (days/week)[3] | | | < 0.001 |
| Mean (SD) | 1.70 (1.84) | 2.07 (2.02) | |
| Range | 0.00-7.00 | 0.00-7.00 | |
| Environmental exposures | | | |
| **$PM_{2.5}$** | | | 0.005 |
| Mean (SD) | 9.96 (1.03) | 9.95 (1.04) | |
| Range | 8.17-19.89 | 8.17-21.25 | |
| **$PM_{10}$** | | | 0.159 |
| Mean (SD) | 16.19 (1.87) | 16.18 (1.89) | |
| Range | 11.78-30.65 | 11.78-30.65 | |
| **$NO_2$** | | | < 0.001 |
| Mean (SD) | 26.49 (7.53) | 26.38 (7.59) | |
| Range | 12.93-107.47 | 12.93-108.49 | |
| **$L_{den}$** | | | 0.174 |
| Mean (SD) | 56.00 (4.21) | 56.02 (4.27) | |
| Range | 51.55-89.29 | 51.55-86.50 | |
| **Greenspace** 1000m | | | < 0.001 |
| Mean (SD) | 45.27 (21.82) | 45.78 (21.71) | |
| Range | 4.49-99.18 | 4.54-99.19 | |

*Note:* GCSEs = general certificate of secondary education; CSE = certificate of secondary education; NVQ = national vocational qualification; HND = higher national diploma; HNC = higher national certificate; BMI = body mass index; PM = particulate matter; $NO_2$ = nitrogen dioxide; $L_{den}$ = day-evening-night noise level. [1]also includes 'other professional qualifications'. [2]Annual household income groups: very low (<£18 000), low (£18 000–30 999), middle (£31 000–51 999), high (£52 000–100 000) and very high (>£100 000). [3]number of days per week engaging in these activities for 10+ minutes continuously.

**Supplement e14. Baseline characteristics stratified by age**

Table e14. **Baseline characteristics stratified by age**

| | Below 65 (*n* = 254 361) | 65 and above (*n* = 53 017) | p value |
|---|---|---|---|
| Health indicators | | | |
| **Health status** | | | < 0.001 |
| Unhealthy | 71 234 (28.0%) | 23 943 (45.2%) | |
| Healthy | 183 127 (72.0%) | 29 074 (54.8%) | |
| **Long-standing illness** | | | < 0.001 |
| Yes | 73 408 (28.9%) | 20 349 (38.4%) | |
| No | 180 953 (71.1%) | 32 668 (61.6%) | |
| **Self-rated health** | | | < 0.001 |
| Poor | 9 382 (3.7%) | 1 684 (3.2%) | |
| Fair | 48 636 (19.1%) | 10 533 (19.9%) | |
| Good | 150 135 (59.0%) | 32 564 (61.4%) | |
| Excellent | 46 208 (18.2%) | 8 236 (15.5%) | |
| Sociodemographic characteristics | | | |
| **Age** | | | < 0.001 |
| Mean (SD) | 53.86 (7.00) | 66.87 (1.48) | |
| Range | 38.00 - 64.00 | 65.00 - 73.00 | |
| **Sex** | | | < 0.001 |
| Female | 135 416 (53.2%) | 24 158 (45.6%) | |
| Male | 118 945 (46.8%) | 28 859 (54.4%) | |
| **Ethnicity** | | | < 0.001 |
| White | 241 835 (95.1%) | 51 730 (97.6%) | |
| Mixed-race | 1 641 (0.6%) | 125 (0.2%) | |
| Black | 3 943 (1.6%) | 314 (0.6%) | |
| Asian | 4 167 (1.6%) | 588 (1.1%) | |
| Chinese | 758 (0.3%) | 60 (0.1%) | |
| Other | 2 017 (0.8%) | 200 (0.4%) | |
| **Highest qualification** | | | < 0.001 |
| None | 25 954 (10.2%) | 13 874 (26.2%) | |
| O levels/GCSEs/CSEs | 72 006 (28.3%) | 12 442 (23.5%) | |
| A levels/NVQ/HND/HNC[1] | 60 115 (23.6%) | 12 469 (23.5%) | |
| Degree | 96 286 (37.9%) | 14 232 (26.8%) | |
| **Household income**[2] | | | < 0.001 |
| Very low | 72 706 (28.6%) | 9 632 (18.2%) | |
| Low | 42 889 (16.9%) | 20 210 (38.1%) | |
| Medium | 59 348 (23.3%) | 18 583 (35.1%) | |
| High | 62 369 (24.5%) | 3 737 (7.0%) | |
| Very high | 17 049 (6.7%) | 855 (1.6%) | |
| **Multiple deprivation** | | | < 0.001 |
| Mean (SD) | 16.95 (13.44) | 15.91 (12.78) | |
| Range | 0.61 - 82.00 | 0.61 - 82.00 | |
| Psychosocial factors | | | |
| **Loneliness** | | | < 0.001 |
| Not lonely | 239 515 (94.2%) | 50 386 (95.0%) | |
| Lonely | 14 846 (5.8%) | 2 631 (5.0%) | |
| **Social isolation** | | | 0.002 |
| Not isolated | 232 297 (91.3%) | 48 634 (91.7%) | |
| Isolated | 22 064 (8.7%) | 4 383 (8.3%) | |
| Lifestyle factors | | | |
| **Smoking status** | | | < 0.001 |
| Never | 142 883 (56.2%) | 25 592 (48.3%) | |
| Former | 84 862 (33.4%) | 23 776 (44.8%) | |
| Current | 26 616 (10.5%) | 3 649 (6.9%) | |
| **Stair climbing frequency** | | | < 0.001 |
| None | 17 222 (6.8%) | 6 827 (12.9%) | |
| 1-5/day | 49 187 (19.3%) | 9 080 (17.1%) | |
| 6-10/day | 96 576 (38.0%) | 19 406 (36.6%) | |
| 11-15/day | 50 249 (19.8%) | 10 066 (19.0%) | |
| 16-20/day | 23 039 (9.1%) | 4 570 (8.6%) | |
| 20+/day | 18 088 (7.1%) | 3 068 (5.8%) | |
| **Alcohol intake frequency** | | | < 0.001 |
| Never | 66 613 (26.2%) | 12 164 (22.9%) | |
| Special occasions | 16 139 (6.3%) | 4 284 (8.1%) | |
| 1-3/month | 25 472 (10.0%) | 6 054 (11.4%) | |
| 1-2/week | 28 784 (11.3%) | 5 014 (9.5%) | |
| 3-4/week | 63 767 (25.1%) | 11 484 (21.7%) | |
| Daily/almost daily | 53 586 (21.1%) | 14 017 (26.4%) | |
| **Sleep duration** (hours/day) | | | < 0.001 |
| Mean (SD) | 7.13 (1.04) | 7.32 (1.11) | |
| Range | 1.00 - 20.00 | 1.00 - 18.00 | |
| **BMI** (kg/m$^2$) | | | < 0.001 |

| | | | |
|---|---|---|---|
| Mean (SD) | 27.24 (4.75) | 27.40 (4.25) | |
| Range | 12.80 - 67.30 | 12.80 - 56.50 | |
| **Walking** (days/week)[3] | | | < 0.001 |
| Mean (SD) | 5.31 (1.97) | 5.63 (1.78) | |
| Range | 0.00 - 7.00 | 0.00 - 7.00 | |
| **Moderate activity** (days/week)[3] | | | < 0.001 |
| Mean (SD) | 3.52 (2.31) | 3.93 (2.32) | |
| Range | 0.00 - 7.00 | 0.00 - 7.00 | |
| **Vigorous activity** (days/week)[3] | | | < 0.001 |
| Mean (SD) | 1.89 (1.93) | 1.80 (1.98) | |
| Range | 0.00 - 7.00 | 0.00 - 7.00 | |
| Environmental exposures | | | |
| **$PM_{2.5}$** | | | < 0.001 |
| Mean (SD) | 9.97 (1.04) | 9.88 (1.01) | |
| Range | 8.17 - 20.19 | 8.17 - 21.25 | |
| **$PM_{10}$** | | | < 0.001 |
| Mean (SD) | 16.20 (1.88) | 16.13 (1.85) | |
| Range | 11.78 - 30.65 | 11.78 - 27.62 | |
| **$NO_2$** | | | < 0.001 |
| Mean (SD) | 26.54 (7.59) | 25.92 (7.40) | |
| Range | 12.93 - 108.49 | 12.93 - 107.07 | |
| **$L_{den}$** | | | < 0.001 |
| Mean (SD) | 56.03 (4.25) | 55.92 (4.21) | |
| Range | 51.55 - 89.29 | 51.55 - 86.28 | |
| **Greenspace** 1000m | | | < 0.001 |
| Mean (SD) | 45.27 (21.77) | 46.69 (21.72) | |
| Range | 4.49 - 99.19 | 5.34 - 99.18 | |

*Note:* GCSEs = general certificate of secondary education; CSE = certificate of secondary education; NVQ = national vocational qualification; HND = higher national diploma; HNC = higher national certificate; BMI = body mass index; PM = particulate matter; $NO_2$ = nitrogen dioxide; $L_{den}$ = day-evening-night noise level. [1]also includes 'other professional qualifications'. [2]Annual household income groups: very low (<£18 000), low (£18 000–30 999), middle (£31 000–51 999), high (£52 000–100 000) and very high (>£100 000). [3]number of days per week engaging in these activities for 10+ minutes continuously.

**Supplement e15. Regression tables health status stratified by sex**

Sociodemographic characteristics

Supplement e15-A. **Sociodemographic characteristics associated with health status stratified by sex**

| | All participants | | | Male | | | Female | | | Interaction term | |
|---|---|---|---|---|---|---|---|---|---|---|---|
| Term | OR | Bonferroni-corrected CI | | OR | Bonferroni-corrected CI | | OR | Bonferroni-corrected CI | | $p_{Bonf.}$ | $p_{BH}$ |
| **Household income**[1] | | | | | | | | | | | |
| Very low | 0.7447 | 0.7148 | 0.7759 | 0.6953 | 0.6545 | 0.7386 | 0.7794 | 0.7368 | 0.8245 | <0.0001 | <0.0001 |
| Low | 0.9196 | 0.8864 | 0.9541 | 0.9052 | 0.8586 | 0.9544 | 0.9326 | 0.8858 | 0.9818 | 0.0006 | 0.0001 |
| Middle | Ref | – | – | Ref | – | – | Ref | – | – | – | – |
| High | 1.0510 | 1.0096 | 1.0941 | 1.0507 | 0.9934 | 1.1113 | 1.0438 | 0.9852 | 1.1060 | 0.4472 | 0.0225 |
| Very high | 1.0472 | 0.9812 | 1.1181 | 1.0541 | 0.9640 | 1.1535 | 1.0257 | 0.9327 | 1.1292 | 0.1482 | 0.0087 |
| **Sex** | | | | | | | | | | | |
| Female | Ref | – | – | Ref | – | – | Ref | – | – | – | – |
| Male | 0.8847 | 0.8610 | 0.9090 | – | – | – | – | – | – | – | – |
| **Age** | 0.9528 | 0.9510 | 0.9546 | 0.9441 | 0.9414 | 0.9467 | 0.9608 | 0.9583 | 0.9634 | <0.0001 | <0.0001 |
| **Multiple deprivation** | 0.9952 | 0.9941 | 0.9963 | 0.9943 | 0.9927 | 0.9959 | 0.9962 | 0.9946 | 0.9977 | <0.0001 | <0.0001 |
| **Ethnicity** | | | | | | | | | | | |
| White | Ref | – | – | Ref | – | – | Ref | – | – | – | – |
| Mixed-race | 1.1037 | 0.9221 | 1.3273 | 1.0296 | 0.7706 | 1.3907 | 1.1729 | 0.9339 | 1.4851 | >0.9999 | 0.6739 |
| Asian | 1.0188 | 0.9149 | 1.1359 | 0.9194 | 0.7985 | 1.0605 | 1.1865 | 1.0024 | 1.4096 | 0.1043 | 0.0065 |
| Black | 1.3125 | 1.1657 | 1.4805 | 1.3079 | 1.0905 | 1.5754 | 1.3178 | 1.1274 | 1.5455 | >0.9999 | 0.9911 |
| Chinese | 1.8339 | 1.3646 | 2.5106 | 1.7054 | 1.0800 | 2.8050 | 1.9827 | 1.3537 | 3.0032 | >0.9999 | 0.8143 |
| Other | 1.2570 | 1.0727 | 1.4779 | 1.3268 | 1.0478 | 1.6919 | 1.2131 | 0.9805 | 1.5104 | >0.9999 | 0.3444 |
| **Highest qualification** | | | | | | | | | | | |
| None | Ref | – | – | Ref | – | – | Ref | – | – | – | – |
| O levels/GCSEs/CSEs | 1.0315 | 0.9870 | 1.0779 | 0.9946 | 0.9330 | 1.0603 | 1.0482 | 0.9858 | 1.1146 | 0.0078 | 0.0006 |
| A levels/NVQ/HND/HNC[2] | 0.9879 | 0.9445 | 1.0332 | 0.9710 | 0.9114 | 1.0344 | 1.0033 | 0.9410 | 1.0697 | 0.0037 | 0.0004 |
| Degree | 0.9558 | 0.9137 | 0.9997 | 0.9400 | 0.8816 | 1.0021 | 0.9624 | 0.9029 | 1.0257 | <0.0001 | <0.0001 |

*Note:* Estimates from Model 3 (i.e. including all explanatory variables). Bonferroni-adjusted (~99.9%) confidence intervals. OR = odds ratio; CI = confidence interval; GCSEs = general certificate of secondary education; CSE = certificate of secondary education; NVQ = national vocational qualification; HND = higher national diploma; HNC = higher national certificate. [1]Annual household income groups: very low (<£18 000), low (£18 000–30 999), middle (£31 000–51 999), high (£52 000–100 000) and very high (>£100 000). [2]also includes 'other professional qualifications'.

Psychosocial factors

| Supplement e15-B. **Psychosocial factors associated with health status stratified by sex** | | | | | | | | | | | |
|---|---|---|---|---|---|---|---|---|---|---|---|
| | All participants | | | Male | | | Female | | | Interaction term | |
| Term | OR | Bonferroni-corrected CI | | OR | Bonferroni-corrected CI | | OR | Bonferroni-corrected CI | | $p_{Bonf.}$ | $p_{BH}$ |
| **Loneliness** | | | | | | | | | | | |
| Not lonely | Ref | – | – | Ref | – | – | Ref | – | – | – | – |
| Lonely | 0.8129 | 0.7697 | 0.8587 | 0.7688 | 0.7112 | 0.8313 | 0.8538 | 0.7904 | 0.9227 | 0.0083 | 0.0006 |
| **Social isolation** | | | | | | | | | | | |
| Not isolated | Ref | – | – | Ref | – | – | Ref | – | – | – | – |
| Isolated | 0.9496 | 0.9067 | 0.9947 | 0.9307 | 0.8711 | 0.9946 | 0.9653 | 0.9044 | 1.0306 | 0.0818 | 0.0055 |

*Note:* Estimates from Model 3 (i.e. including all explanatory variables). Bonferroni-adjusted (~99.9%) confidence intervals. OR = odds ratio; CI = confidence interval.

Lifestyle factors

Supplement e15-C. **Lifestyle factors associated with health status stratified by sex**

| | All participants | | | Male | | | Female | | | Interaction term | |
|---|---|---|---|---|---|---|---|---|---|---|---|
| Term | OR | Bonferroni-corrected CI | | OR | Bonferroni-corrected CI | | OR | Bonferroni-corrected CI | | $p_{Bonf.}$ | $p_{BH}$ |
| **Sleep duration** (hours/day) | 0.9748 | 0.9631 | 0.9868 | 0.9635 | 0.9465 | 0.9808 | 0.9947 | 0.9782 | 1.0116 | <0.0001 | <0.0001 |
| **Physical activity** (days/week)[1] | | | | | | | | | | | |
| Walking | 1.0105 | 1.0032 | 1.0178 | 1.0062 | 0.9960 | 1.0164 | 1.0160 | 1.0055 | 1.0266 | >0.9999 | 0.1722 |
| Moderate activity | 1.0032 | 0.9964 | 1.0100 | 1.0093 | 0.9992 | 1.0195 | 0.9979 | 0.9887 | 1.0071 | 0.8784 | 0.0351 |
| Vigorous activity | 1.0319 | 1.0238 | 1.0400 | 1.0319 | 1.0205 | 1.0433 | 1.0279 | 1.0164 | 1.0395 | 0.0064 | 0.0006 |
| **Stair climbing frequency** | | | | | | | | | | | |
| None | Ref | – | – | Ref | – | – | Ref | – | – | – | – |
| 1-5/day | 1.0704 | 1.0148 | 1.1289 | 1.0604 | 0.9827 | 1.1442 | 1.0778 | 0.9998 | 1.1617 | >0.9999 | 0.1995 |
| 6-10/day | 1.1756 | 1.1188 | 1.2352 | 1.1657 | 1.0854 | 1.2517 | 1.1836 | 1.1044 | 1.2683 | 0.5750 | 0.0261 |
| 11-15/day | 1.1714 | 1.1098 | 1.2363 | 1.1553 | 1.0684 | 1.2491 | 1.1898 | 1.1038 | 1.2823 | 0.6537 | 0.0284 |
| 16-20/day | 1.1650 | 1.0937 | 1.2410 | 1.1425 | 1.0422 | 1.2525 | 1.1971 | 1.0971 | 1.3063 | >0.9999 | 0.1028 |
| 20+/day | 1.1881 | 1.1097 | 1.2723 | 1.2364 | 1.1172 | 1.3688 | 1.1660 | 1.0627 | 1.2797 | 0.0004 | <0.0001 |
| **Alcohol intake frequency** | | | | | | | | | | | |
| Never | 0.6285 | 0.5946 | 0.6644 | 0.6097 | 0.5586 | 0.6656 | 0.6279 | 0.5842 | 0.6749 | 0.7799 | 0.0325 |
| Special occasions | 0.7792 | 0.7429 | 0.8174 | 0.7591 | 0.7003 | 0.8231 | 0.7720 | 0.7270 | 0.8199 | >0.9999 | 0.0621 |
| 1-3/month | 0.9129 | 0.8708 | 0.9570 | 0.8829 | 0.8204 | 0.9503 | 0.9244 | 0.8689 | 0.9836 | >0.9999 | 0.1105 |
| 1-2/week | Ref | – | – | Ref | – | – | Ref | – | – | – | – |
| 3-4/week | 1.0590 | 1.0198 | 1.0998 | 1.0837 | 1.0280 | 1.1424 | 1.0342 | 0.9794 | 1.0922 | 0.4505 | 0.0225 |
| Daily/almost daily | 1.0334 | 0.9939 | 1.0745 | 1.0604 | 1.0054 | 1.1183 | 0.9982 | 0.9419 | 1.0579 | >0.9999 | 0.0392 |
| **BMI** (kg/m$^2$) | 0.9683 | 0.9656 | 0.9711 | 0.9564 | 0.9520 | 0.9608 | 0.9746 | 0.9711 | 0.9782 | <0.0001 | <0.0001 |
| **Smoking status** | | | | | | | | | | | |
| Never | Ref | – | – | Ref | – | – | Ref | – | – | – | – |
| Former | 0.7922 | 0.7698 | 0.8152 | 0.7694 | 0.7384 | 0.8018 | 0.8366 | 0.8036 | 0.8711 | <0.0001 | <0.0001 |
| Current | 0.7518 | 0.7183 | 0.7869 | 0.7812 | 0.7337 | 0.8321 | 0.7188 | 0.6725 | 0.7685 | 0.0747 | 0.0053 |

*Note:* Estimates from Model 3 (i.e. including all explanatory variables). Bonferroni-adjusted (~99.9%) confidence intervals. OR = odds ratio; CI = confidence interval; BMI = body mass index. [1]number of days per week engaging in these activities for 10+ minutes continuously.

Supplement e15-D. **Environmental exposures associated with health status stratified by sex**

| | All participants | | | Male | | | Female | | | Interaction term | |
|---|---|---|---|---|---|---|---|---|---|---|---|
| Term | OR | Bonferroni-corrected CI | | OR | Bonferroni-corrected CI | | OR | Bonferroni-corrected CI | | $p_{Bonf.}$ | $p_{BH}$ |
| **$PM_{2.5}$** | 0.9656 | 0.9411 | 0.9908 | 0.9614 | 0.9266 | 0.9976 | 0.9690 | 0.9347 | 1.0045 | 0.1946 | 0.0108 |
| **$PM_{10}$** | 1.0019 | 0.9935 | 1.0104 | 1.0012 | 0.9892 | 1.0134 | 1.0029 | 0.9911 | 1.0148 | >0.9999 | 0.1722 |
| **$NO_2$** | 1.0036 | 0.9993 | 1.0078 | 1.0010 | 0.9949 | 1.0071 | 1.0060 | 1.0001 | 1.0120 | 0.4953 | 0.0236 |
| **$L_{den}$** | 0.9981 | 0.9945 | 1.0017 | 1.0009 | 0.9957 | 1.0061 | 0.9955 | 0.9905 | 1.0005 | >0.9999 | 0.5176 |
| **Greenspace** 1000m | 0.9995 | 0.9985 | 1.0004 | 0.9986 | 0.9973 | 1.0000 | 1.0003 | 0.9989 | 1.0017 | >0.9999 | 0.7593 |

*Note:* Estimates from Model 3 (i.e. including all explanatory variables). Bonferroni-adjusted (~99.9%) confidence intervals. OR = odds ratio; CI = confidence interval; PM = particulate matter; $NO_2$ = nitrogen dioxide; $L_{den}$ = day-evening-night noise level.

**Supplement e16. Confidence interval plots health status stratified by sex**

Sociodemographic and psychosocial factors

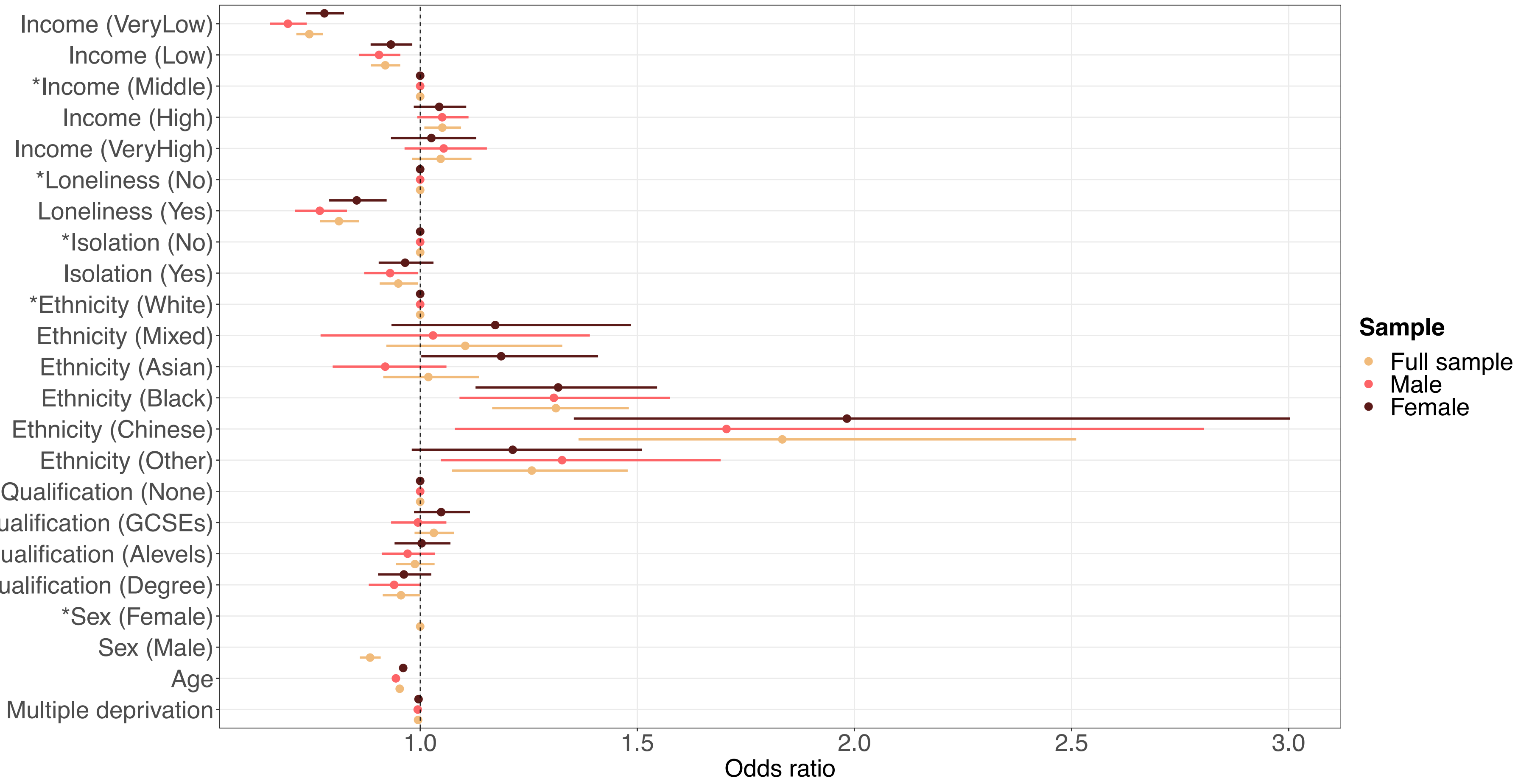


**Supplement e16-A.** Sociodemographic characteristics and psychosocial factors associated with health status, stratified by sex. Confidence interval plot (odds ratio ± Bonferroni-adjusted (~99.9%) confidence intervals) for Model 3 (i.e. including all explanatory variables). GCSEs = general certificate of secondary education. [*]Indicates reference group for categorical explanatory variables. Annual household income groups: very low (<£18 000), low (£18 000–30 999), middle (£31 000–51 999), high (£52 000–100 000) and very high (>£100 000). 'GCSEs' also includes O levels and certificate of secondary education (CSE). 'A levels' also includes national vocational qualification (NVQ), higher national diploma (HND), higher national certificate (HNC) and 'other professional qualifications'.

Lifestyle factors

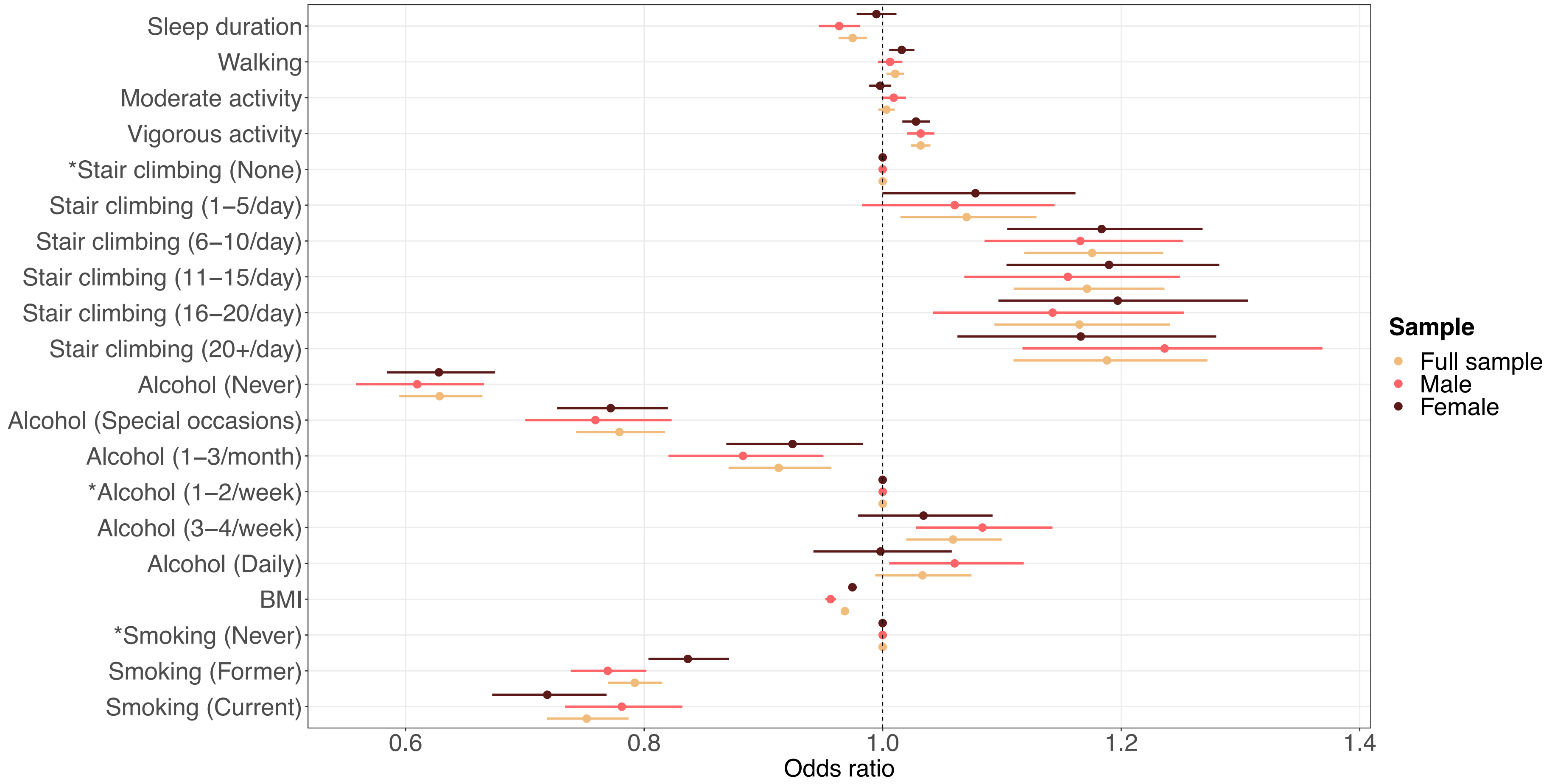


**Supplement e16-B.** Lifestyle factors associated with health status, stratified by sex. Confidence interval plot (odds ratio ± Bonferroni-adjusted (~99.9%) confidence intervals) for Model 3 (i.e. including all explanatory variables). BMI = body mass index. *Indicates reference group for categorical explanatory variables.

Environmental exposures

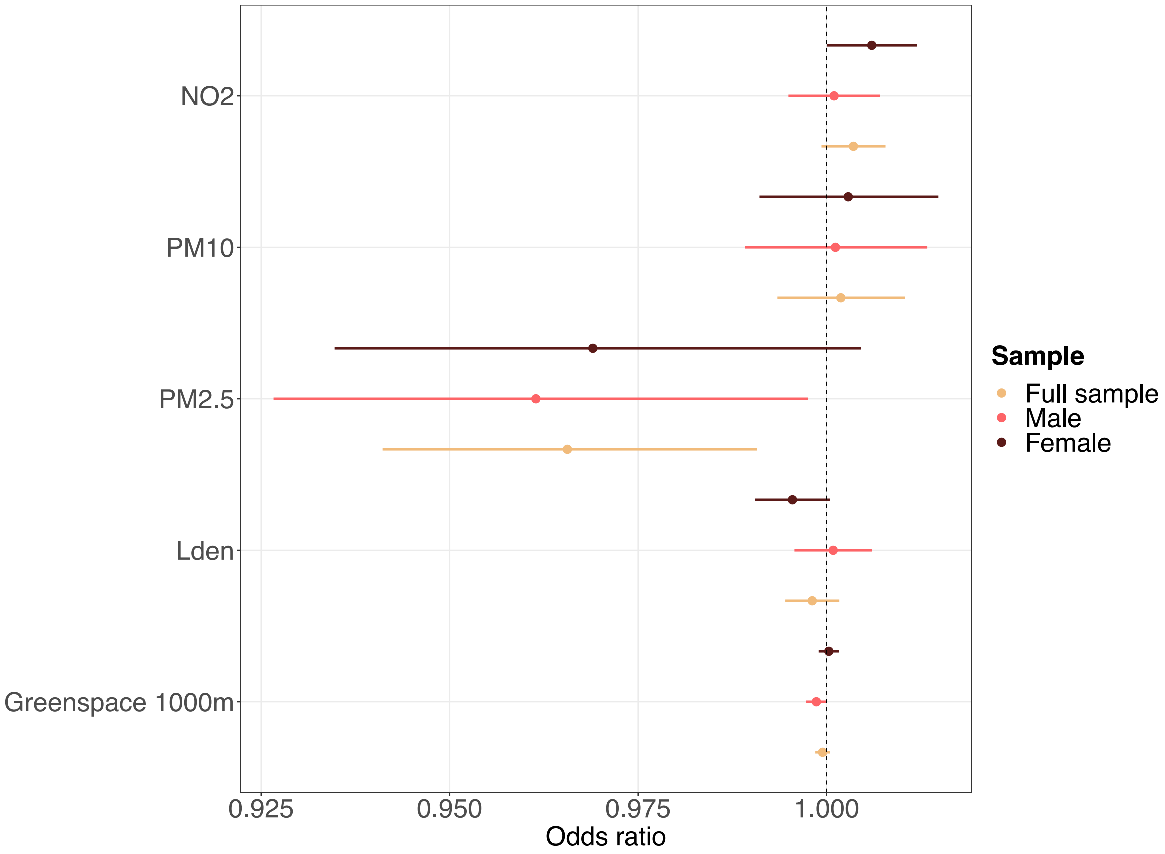


**Supplement e16-C.** Environmental exposures associated with health status, stratified by sex. Confidence interval plot (odds ratio ± Bonferroni-adjusted (~99.9%) confidence intervals) for Model 3 (i.e. including all explanatory variables). PM = particulate matter; $NO_2$ = nitrogen dioxide; $L_{den}$ = day-evening-night noise level.

**Supplement e17. Regression tables health status stratified by age**

Sociodemographic characteristics

Supplement e17-A. **Sociodemographic characteristics associated with health status stratified by age**

| | All participants | | | Below 65 | | | 65 and above | | | Interaction term | |
|---|---|---|---|---|---|---|---|---|---|---|---|
| Term | OR | Bonferroni-corrected CI | | OR | Bonferroni-corrected CI | | OR | Bonferroni-corrected CI | | $p_{Bonf.}$ | $p_{BH}$ |
| **Household income**[1] | | | | | | | | | | | |
| Very low | 0.7447 | 0.7148 | 0.7759 | 0.7132 | 0.6807 | 0.7471 | 0.8509 | 0.7764 | 0.9326 | <0.0001 | <0.0001 |
| Low | 0.9196 | 0.8864 | 0.9541 | 0.9249 | 0.8875 | 0.9639 | 0.9432 | 0.8666 | 1.0264 | >0.9999 | 0.7976 |
| Middle | Ref | – | – | Ref | – | – | Ref | – | – | – | – |
| High | 1.0510 | 1.0096 | 1.0941 | 1.0509 | 1.0070 | 1.0967 | 0.9563 | 0.8415 | 1.0872 | 0.2171 | 0.0155 |
| Very high | 1.0472 | 0.9812 | 1.1181 | 1.0428 | 0.9741 | 1.1169 | 0.9791 | 0.7734 | 1.2424 | >0.9999 | 0.2886 |
| **Sex** | | | | | | | | | | | |
| Female | Ref | – | – | Ref | – | – | Ref | – | – | – | – |
| Male | 0.8847 | 0.8610 | 0.9090 | 0.9270 | 0.8992 | 0.9557 | 0.7548 | 0.7101 | 0.8023 | <0.0001 | <0.0001 |
| **Age** | 0.9528 | 0.9510 | 0.9546 | 0.9553 | 0.9530 | 0.9575 | 0.9436 | 0.9255 | 0.9620 | – | – |
| **Multiple deprivation** | 0.9952 | 0.9941 | 0.9963 | 0.9951 | 0.9939 | 0.9963 | 0.9958 | 0.9933 | 0.9984 | >0.9999 | 0.1995 |
| **Ethnicity** | | | | | | | | | | | |
| White | Ref | – | – | Ref | – | – | Ref | – | – | – | – |
| Mixed-race | 1.1037 | 0.9221 | 1.3273 | 1.1227 | 0.9296 | 1.3634 | 1.0573 | 0.5866 | 1.9315 | >0.9999 | 0.6005 |
| Asian | 1.0188 | 0.9149 | 1.1359 | 1.0654 | 0.9472 | 1.2003 | 0.8668 | 0.6575 | 1.1426 | <0.0001 | <0.0001 |
| Black | 1.3125 | 1.1657 | 1.4805 | 1.3410 | 1.1832 | 1.5233 | 1.2654 | 0.8693 | 1.8516 | >0.9999 | 0.9840 |
| Chinese | 1.8339 | 1.3646 | 2.5106 | 1.8577 | 1.3604 | 2.5919 | 1.9177 | 0.7868 | 5.2841 | >0.9999 | 0.7976 |
| Other | 1.2570 | 1.0727 | 1.4779 | 1.2936 | 1.0925 | 1.5381 | 1.1064 | 0.6937 | 1.7755 | >0.9999 | 0.0544 |
| **Highest qualification** | | | | | | | | | | | |
| None | Ref | – | – | Ref | – | – | Ref | – | – | – | – |
| O levels/GCSEs/CSEs | 1.0315 | 0.9870 | 1.0779 | 1.0315 | 0.9786 | 1.0873 | 1.0540 | 0.9688 | 1.1466 | >0.9999 | 0.1995 |
| A levels/NVQ/HND/HNC[2] | 0.9879 | 0.9445 | 1.0332 | 0.9907 | 0.9388 | 1.0454 | 1.0009 | 0.9197 | 1.0892 | 0.8449 | 0.0422 |
| Degree | 0.9558 | 0.9137 | 0.9997 | 0.9517 | 0.9022 | 1.0039 | 1.0103 | 0.9246 | 1.1040 | 0.8264 | 0.0422 |

*Note:* Estimates from Model 3 (i.e. including all explanatory variables). Bonferroni-adjusted (~99.9%) confidence intervals. OR = odds ratio; CI = confidence interval; GCSEs = general certificate of secondary education; CSE = certificate of secondary education; NVQ = national vocational qualification; HND = higher national diploma; HNC = higher national certificate. [1]Annual household income groups: very low (<£18 000), low (£18 000–30 999), middle (£31 000–51 999), high (£52 000–100 000) and very high (>£100 000). [2]also includes 'other professional qualifications'.

Psychosocial factors

Supplement e17-B. **Psychosocial factors associated with health status stratified by age**

| | All participants | | | Below 65 | | | 65 and above | | | Interaction term | |
|---|---|---|---|---|---|---|---|---|---|---|---|
| Term | OR | Bonferroni-corrected CI | | OR | Bonferroni-corrected CI | | OR | Bonferroni-corrected CI | | $p_{Bonf.}$ | $p_{BH}$ |
| **Loneliness** | | | | | | | | | | | |
| Not lonely | Ref | – | – | Ref | – | – | Ref | – | – | – | – |
| Lonely | 0.8129 | 0.7697 | 0.8587 | 0.7959 | 0.7497 | 0.8452 | 0.8958 | 0.7844 | 1.0231 | 0.3908 | 0.0244 |
| **Social isolation** | | | | | | | | | | | |
| Not isolated | Ref | – | – | Ref | – | – | Ref | – | – | – | – |
| Isolated | 0.9496 | 0.9067 | 0.9947 | 0.9443 | 0.8970 | 0.9942 | 0.9625 | 0.8650 | 1.0711 | 0.0112 | 0.0009 |

*Note:* Estimates from Model 3 (i.e. including all explanatory variables). Bonferroni-adjusted (~99.9%) confidence intervals. OR = odds ratio; CI = confidence interval.

Lifestyle factors

Supplement e17-C. **Lifestyle factors associated with health status stratified by age**

| | All participants | | | Below 65 | | | 65 and above | | | Interaction term | |
|---|---|---|---|---|---|---|---|---|---|---|---|
| Term | OR | Bonferroni-corrected CI | | OR | Bonferroni-corrected CI | | OR | Bonferroni-corrected CI | | $p_{Bonf.}$ | $p_{BH}$ |
| **Sleep duration** (hours/day) | 0.9748 | 0.9631 | 0.9868 | 0.9784 | 0.9650 | 0.9920 | 0.9787 | 0.9537 | 1.0043 | >0.9999 | 0.1995 |
| **Physical activity** (days/week)[1] | | | | | | | | | | | |
| Walking | 1.0105 | 1.0032 | 1.0178 | 1.0084 | 1.0003 | 1.0165 | 1.0242 | 1.0067 | 1.0421 | <0.0001 | <0.0001 |
| Moderate activity | 1.0032 | 0.9964 | 1.0100 | 1.0026 | 0.9949 | 1.0103 | 1.0057 | 0.9913 | 1.0204 | >0.9999 | 0.1763 |
| Vigorous activity | 1.0319 | 1.0238 | 1.0400 | 1.0338 | 1.0246 | 1.0432 | 1.0241 | 1.0075 | 1.0410 | >0.9999 | 0.1536 |
| **Stair climbing frequency** | | | | | | | | | | | |
| None | Ref | – | – | Ref | – | – | Ref | – | – | – | – |
| 1-5/day | 1.0704 | 1.0148 | 1.1289 | 1.1064 | 1.0395 | 1.1774 | 0.9614 | 0.8649 | 1.0686 | <0.0001 | <0.0001 |
| 6-10/day | 1.1756 | 1.1188 | 1.2352 | 1.2041 | 1.1355 | 1.2766 | 1.1106 | 1.0115 | 1.2194 | <0.0001 | <0.0001 |
| 11-15/day | 1.1714 | 1.1098 | 1.2363 | 1.1985 | 1.1249 | 1.2767 | 1.1195 | 1.0079 | 1.2434 | 0.0004 | <0.0001 |
| 16-20/day | 1.1650 | 1.0937 | 1.2410 | 1.1772 | 1.0942 | 1.2665 | 1.1735 | 1.0324 | 1.3343 | 0.4709 | 0.0277 |
| 20+/day | 1.1881 | 1.1097 | 1.2723 | 1.2150 | 1.1236 | 1.3139 | 1.1410 | 0.9861 | 1.3210 | 0.0128 | 0.0010 |
| **Alcohol intake frequency** | | | | | | | | | | | |
| Never | 0.6285 | 0.5946 | 0.6644 | 0.6145 | 0.5772 | 0.6543 | 0.6783 | 0.6023 | 0.7636 | 0.0021 | 0.0002 |
| Special occasions | 0.7792 | 0.7429 | 0.8174 | 0.7720 | 0.7317 | 0.8146 | 0.8078 | 0.7273 | 0.8972 | 0.0052 | 0.0005 |
| 1-3/month | 0.9129 | 0.8708 | 0.9570 | 0.9015 | 0.8557 | 0.9499 | 0.9684 | 0.8669 | 1.0820 | >0.9999 | 0.0775 |
| 1-2/week | Ref | – | – | Ref | – | – | Ref | – | – | – | – |
| 3-4/week | 1.0590 | 1.0198 | 1.0998 | 1.0552 | 1.0117 | 1.1006 | 1.0730 | 0.9841 | 1.1698 | >0.9999 | 0.2894 |
| Daily/almost daily | 1.0334 | 0.9939 | 1.0745 | 1.0310 | 0.9864 | 1.0776 | 1.0607 | 0.9751 | 1.1538 | >0.9999 | 0.1995 |
| **BMI** (kg/m$^2$) | 0.9683 | 0.9656 | 0.9711 | 0.9694 | 0.9663 | 0.9724 | 0.9625 | 0.9557 | 0.9693 | <0.0001 | <0.0001 |
| **Smoking status** | | | | | | | | | | | |
| Never | Ref | – | – | Ref | – | – | Ref | – | – | – | – |
| Former | 0.7922 | 0.7698 | 0.8152 | 0.8054 | 0.7796 | 0.8320 | 0.7632 | 0.7175 | 0.8117 | <0.0001 | <0.0001 |
| Current | 0.7518 | 0.7183 | 0.7869 | 0.7423 | 0.7066 | 0.7799 | 0.8400 | 0.7460 | 0.9459 | 0.6816 | 0.0379 |

*Note:* Estimates from Model 3 (i.e. including all explanatory variables). Bonferroni-adjusted (~99.9%) confidence intervals. OR = odds ratio; CI = confidence interval; BMI = body mass index. [1]number of days per week engaging in these activities for 10+ minutes continuously.

Supplement e17-D. **Environmental exposures associated with health status stratified by age**

| | All participants | | | Below 65 | | | 65 and above | | | Interaction term | |
|---|---|---|---|---|---|---|---|---|---|---|---|
| Term | OR | Bonferroni-corrected CI | | OR | Bonferroni-corrected CI | | OR | Bonferroni-corrected CI | | $p_{Bonf.}$ | $p_{BH}$ |
| **$PM_{2.5}$** | 0.9656 | 0.9411 | 0.9908 | 0.9641 | 0.9367 | 0.9922 | 0.9640 | 0.9099 | 1.0213 | >0.9999 | 0.2963 |
| **$PM_{10}$** | 1.0019 | 0.9935 | 1.0104 | 1.0006 | 0.9912 | 1.0101 | 1.0064 | 0.9876 | 1.0255 | >0.9999 | 0.2092 |
| **$NO_2$** | 1.0036 | 0.9993 | 1.0078 | 1.0037 | 0.9990 | 1.0085 | 1.0043 | 0.9949 | 1.0138 | >0.9999 | 0.8340 |
| **$L_{den}$** | 0.9981 | 0.9945 | 1.0017 | 0.9990 | 0.9950 | 1.0030 | 0.9946 | 0.9866 | 1.0026 | 0.3084 | 0.0206 |
| **Greenspace** 1000m | 0.9995 | 0.9985 | 1.0004 | 0.9995 | 0.9984 | 1.0006 | 0.9994 | 0.9972 | 1.0015 | >0.9999 | 0.2912 |

*Note:* Estimates from Model 3 (i.e. including all explanatory variables). Bonferroni-adjusted (~99.9%) confidence intervals. OR = odds ratio; CI = confidence interval; PM = particulate matter; $NO_2$ = nitrogen dioxide; $L_{den}$ = day-evening-night noise level.

**Supplement e18. Confidence interval plots health status stratified by age**

Sociodemographic and psychosocial factors

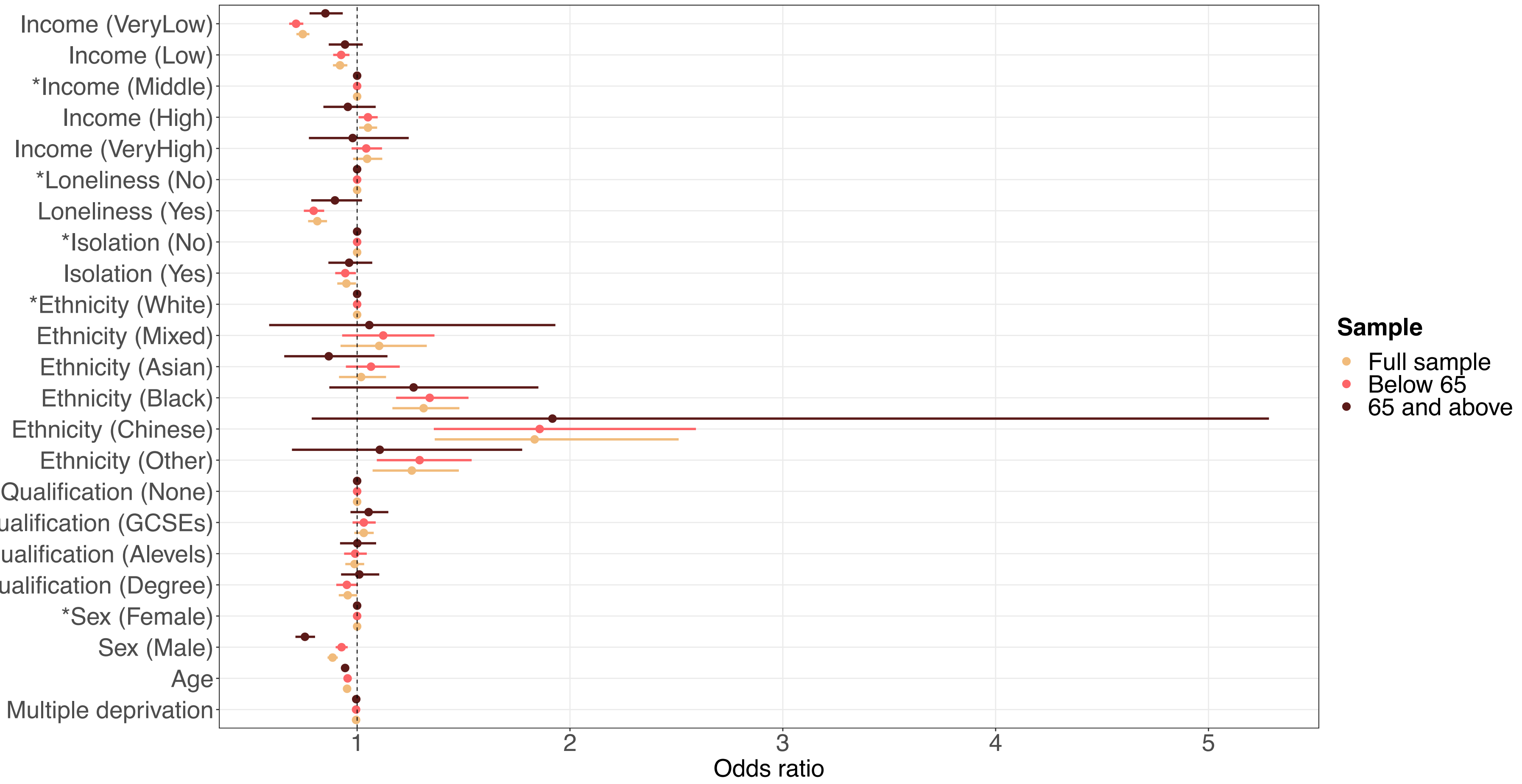


**Supplement e18-A.** Sociodemographic characteristics and psychosocial factors associated with health status, stratified by age. Confidence interval plot (odds ratio ± Bonferroni-adjusted (~99.9%) confidence intervals) for Model 3 (i.e. including all explanatory variables). GCSEs = general certificate of secondary education. *Indicates reference group for categorical explanatory variables. Annual household income groups: very low (<£18 000), low (£18 000–30 999), middle (£31 000–51 999), high (£52 000–100 000) and very high (>£100 000). 'GCSEs' also includes O levels and certificate of secondary education (CSE). 'A levels' also includes national vocational qualification (NVQ), higher national diploma (HND), higher national certificate (HNC) and 'other professional qualifications'.

Lifestyle factors

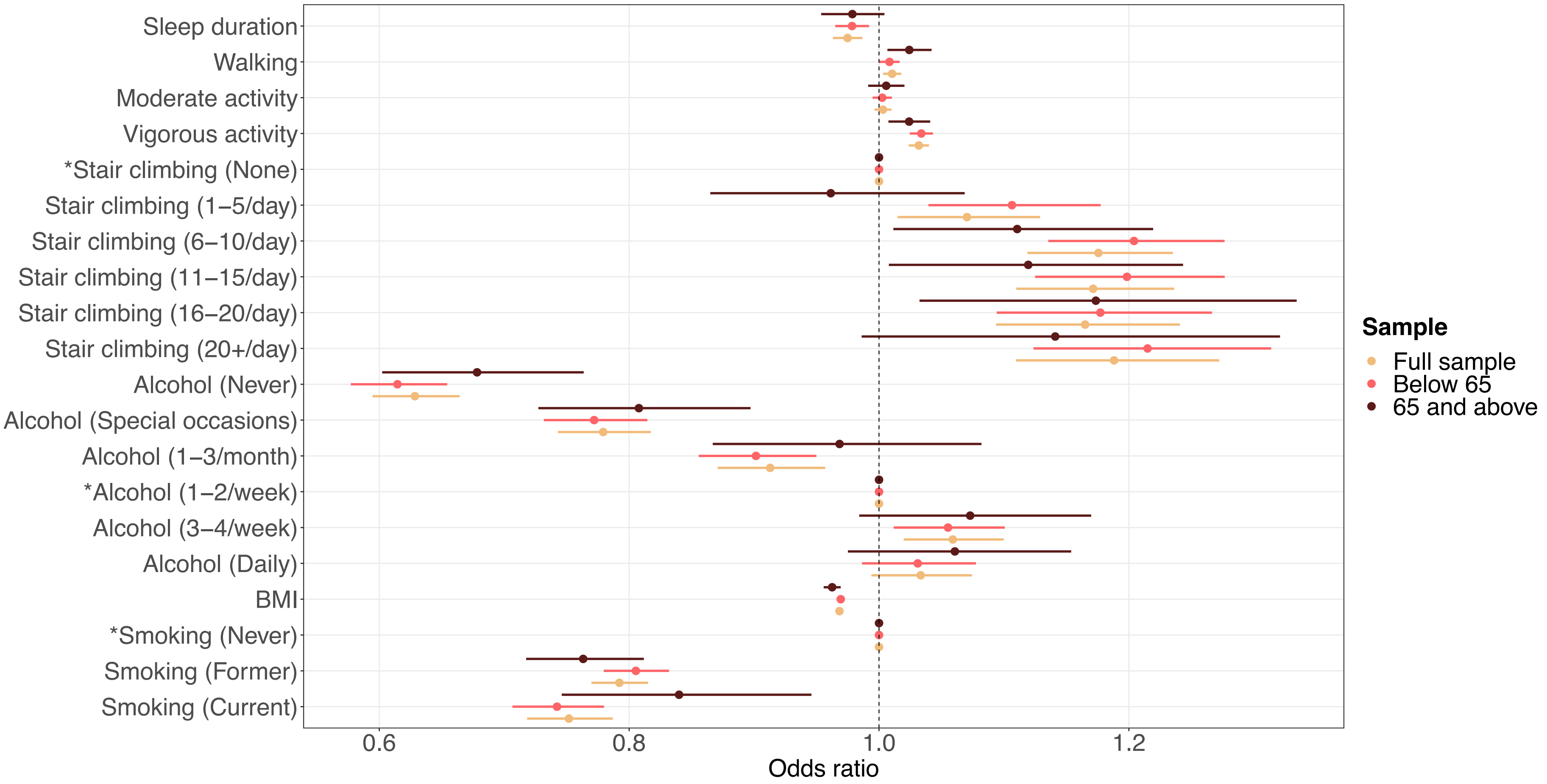


**Supplement e18-B.** Lifestyle factors associated with health status, stratified by age. Confidence interval plot (odds ratio ± Bonferroni-adjusted (~99.9%) confidence intervals) for Model 3 (i.e. including all explanatory variables). BMI = body mass index. *Indicates reference group for categorical explanatory variables.

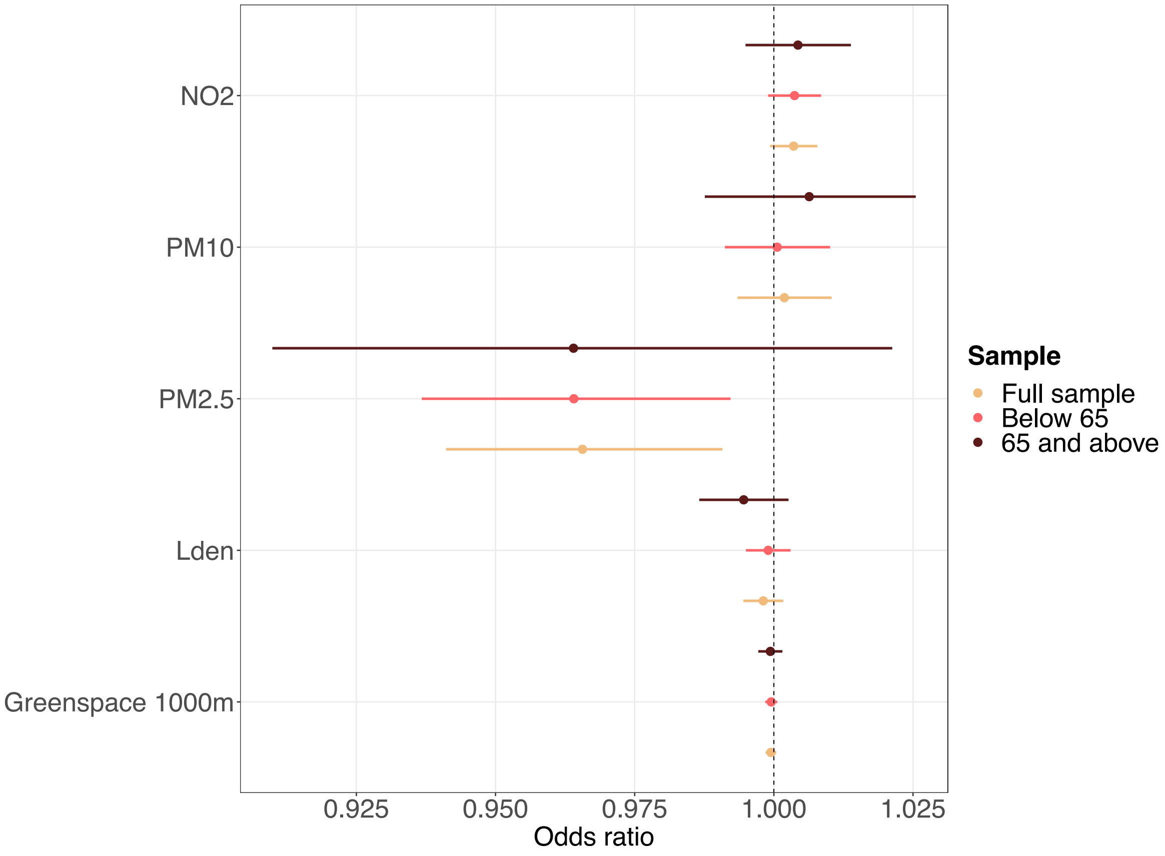


**Supplement e18-C.** Environmental exposures associated with health status, stratified by age. Confidence interval plot (odds ratio ± Bonferroni-adjusted (~99.9%) confidence intervals) for Model 3 (i.e. including all explanatory variables). PM = particulate matter; $NO_2$ = nitrogen dioxide; $L_{den}$ = day-evening-night noise level.

**Supplement e19. Regression tables long-standing illness stratified by sex**

Sociodemographic characteristics

Supplement e19-A. **Sociodemographic characteristics associated with long-standing illness stratified by sex**

| | All participants | | | Male | | | Female | | | Interaction term | |
|---|---|---|---|---|---|---|---|---|---|---|---|
| Term | OR | Bonferroni-corrected CI | | OR | Bonferroni-corrected CI | | OR | Bonferroni-corrected CI | | $p_{Bonf.}$ | $p_{BH}$ |
| **Household income**[1] | | | | | | | | | | | |
| Very low | 0.6654 | 0.6384 | 0.6934 | 0.6067 | 0.5714 | 0.6442 | 0.7141 | 0.6743 | 0.7563 | <0.0001 | <0.0001 |
| Low | 0.8906 | 0.8581 | 0.9244 | 0.8593 | 0.8154 | 0.9056 | 0.9228 | 0.8752 | 0.9729 | 0.0001 | <0.0001 |
| Middle | Ref | – | – | Ref | – | – | Ref | – | – | – | – |
| High | 1.1171 | 1.0726 | 1.1634 | 1.1170 | 1.0568 | 1.1807 | 1.1068 | 1.0426 | 1.1752 | >0.9999 | 0.1703 |
| Very high | 1.2524 | 1.1703 | 1.3410 | 1.2639 | 1.1547 | 1.3847 | 1.2112 | 1.0934 | 1.3436 | 0.9984 | 0.0605 |
| **Sex** | | | | | | | | | | | |
| Female | Ref | – | – | Ref | – | – | Ref | – | – | – | – |
| Male | 0.7585 | 0.7380 | 0.7797 | – | – | – | – | – | – | – | – |
| **Age** | 0.9730 | 0.9711 | 0.9748 | 0.9684 | 0.9658 | 0.9710 | 0.9775 | 0.9748 | 0.9801 | <0.0001 | <0.0001 |
| **Multiple deprivation** | 0.9921 | 0.9910 | 0.9932 | 0.9919 | 0.9903 | 0.9934 | 0.9924 | 0.9908 | 0.9940 | 0.3315 | 0.0255 |
| **Ethnicity** | | | | | | | | | | | |
| White | Ref | – | – | Ref | – | – | Ref | – | – | – | – |
| Mixed-race | 1.1223 | 0.9387 | 1.3474 | 1.1439 | 0.8594 | 1.5376 | 1.1235 | 0.8952 | 1.4203 | >0.9999 | 0.5334 |
| Asian | 1.2225 | 1.0962 | 1.3652 | 1.1664 | 1.0117 | 1.3474 | 1.2903 | 1.0886 | 1.5350 | >0.9999 | 0.9754 |
| Black | 1.1852 | 1.0591 | 1.3277 | 1.4492 | 1.2161 | 1.7330 | 1.0314 | 0.8905 | 1.1966 | <0.0001 | <0.0001 |
| Chinese | 1.6205 | 1.2207 | 2.1842 | 1.5633 | 1.0145 | 2.4879 | 1.7049 | 1.1788 | 2.5373 | >0.9999 | 0.9159 |
| Other | 1.2027 | 1.0301 | 1.4078 | 1.2967 | 1.0326 | 1.6367 | 1.1288 | 0.9151 | 1.3993 | >0.9999 | 0.0605 |
| **Highest qualification** | | | | | | | | | | | |
| None | Ref | – | – | Ref | – | – | Ref | – | – | – | – |
| O levels/GCSEs/CSEs | 0.9920 | 0.9485 | 1.0375 | 0.9806 | 0.9199 | 1.0453 | 0.9820 | 0.9212 | 1.0466 | 0.0149 | 0.0014 |
| A levels/NVQ/HND/HNC[2] | 0.8953 | 0.8554 | 0.9371 | 0.9294 | 0.8723 | 0.9902 | 0.8574 | 0.8023 | 0.9161 | <0.0001 | <0.0001 |
| Degree | 0.8550 | 0.8167 | 0.8950 | 0.9002 | 0.8442 | 0.9599 | 0.8028 | 0.7514 | 0.8576 | <0.0001 | <0.0001 |

*Note:* Estimates from Model 3 (i.e. including all explanatory variables). Bonferroni-adjusted (~99.9%) confidence intervals. OR = odds ratio; CI = confidence interval; GCSEs = general certificate of secondary education; CSE = certificate of secondary education; NVQ = national vocational qualification; HND = higher national diploma; HNC = higher national certificate. [1]Annual household income groups: very low (<£18 000), low (£18 000–30 999), middle (£31 000–51 999), high (£52 000–100 000) and very high (>£100 000). [2]also includes 'other professional qualifications'.

Psychosocial factors

| Supplement e19-B. **Psychosocial factors associated with long-standing illness stratified by sex** | | | | | | | | | | |
|---|---|---|---|---|---|---|---|---|---|---|
| | All participants | | | Male | | | Female | | | Interaction term | |
| Term | OR | Bonferroni-corrected CI | | OR | Bonferroni-corrected CI | | OR | Bonferroni-corrected CI | | $p_{Bonf.}$ | $p_{BH}$ |
| **Loneliness** | | | | | | | | | | | |
| Not lonely | Ref | – | – | Ref | – | – | Ref | – | – | – | – |
| Lonely | 0.7097 | 0.6722 | 0.7494 | 0.6682 | 0.6191 | 0.7214 | 0.7526 | 0.6964 | 0.8136 | 0.0037 | 0.0005 |
| **Social isolation** | | | | | | | | | | | |
| Not isolated | Ref | – | – | Ref | – | – | Ref | – | – | – | – |
| Isolated | 0.9185 | 0.8770 | 0.9621 | 0.8990 | 0.8422 | 0.9599 | 0.9325 | 0.8731 | 0.9963 | >0.9999 | 0.0605 |

*Note:* Estimates from Model 3 (i.e. including all explanatory variables). Bonferroni-adjusted (~99.9%) confidence intervals. OR = odds ratio; CI = confidence interval.

| Supplement e19-C. **Lifestyle factors associated with long-standing illness stratified by sex** | | | | | | | | | | | |
|---|---|---|---|---|---|---|---|---|---|---|---|
| | All participants | | | Male | | | Female | | | Interaction term | |
| Term | OR | Bonferroni-corrected CI | | OR | Bonferroni-corrected CI | | OR | Bonferroni-corrected CI | | $p_{Bonf.}$ | $p_{BH}$ |
| **Sleep duration** (hours/day) | 1.0004 | 0.9882 | 1.0128 | 1.0000 | 0.9825 | 1.0179 | 1.0087 | 0.9915 | 1.0262 | >0.9999 | 0.0605 |
| **Physical activity** (days/week)[1] | | | | | | | | | | | |
| Walking | 1.0250 | 1.0175 | 1.0325 | 1.0173 | 1.0071 | 1.0275 | 1.0342 | 1.0233 | 1.0453 | 0.0177 | 0.0015 |
| Moderate activity | 0.9984 | 0.9916 | 1.0053 | 1.0097 | 0.9997 | 1.0199 | 0.9893 | 0.9799 | 0.9987 | 0.3576 | 0.0255 |
| Vigorous activity | 1.0641 | 1.0557 | 1.0727 | 1.0618 | 1.0502 | 1.0736 | 1.0636 | 1.0512 | 1.0761 | >0.9999 | 0.0721 |
| **Stair climbing frequency** | | | | | | | | | | | |
| None | Ref | – | – | Ref | – | – | Ref | – | – | – | – |
| 1-5/day | 1.1101 | 1.0522 | 1.1710 | 1.0747 | 0.9963 | 1.1592 | 1.1463 | 1.0625 | 1.2366 | >0.9999 | 0.9829 |
| 6-10/day | 1.2937 | 1.2307 | 1.3598 | 1.2203 | 1.1365 | 1.3100 | 1.3692 | 1.2762 | 1.4688 | >0.9999 | 0.2991 |
| 11-15/day | 1.3131 | 1.2434 | 1.3866 | 1.2388 | 1.1459 | 1.3392 | 1.3897 | 1.2873 | 1.5001 | >0.9999 | 0.4917 |
| 16-20/day | 1.3066 | 1.2255 | 1.3932 | 1.2230 | 1.1158 | 1.3406 | 1.3984 | 1.2784 | 1.5300 | >0.9999 | 0.3744 |
| 20+/day | 1.2975 | 1.2108 | 1.3907 | 1.2701 | 1.1483 | 1.4053 | 1.3355 | 1.2141 | 1.4695 | >0.9999 | 0.3002 |
| **Alcohol intake frequency** | | | | | | | | | | | |
| Never | 0.5533 | 0.5234 | 0.5850 | 0.5987 | 0.5488 | 0.6532 | 0.5192 | 0.4827 | 0.5584 | 0.0152 | 0.0014 |
| Special occasions | 0.7142 | 0.6809 | 0.7492 | 0.7400 | 0.6833 | 0.8016 | 0.6913 | 0.6506 | 0.7347 | >0.9999 | 0.2105 |
| 1-3/month | 0.8486 | 0.8096 | 0.8895 | 0.8393 | 0.7811 | 0.9020 | 0.8477 | 0.7962 | 0.9027 | >0.9999 | 0.9159 |
| 1-2/week | Ref | – | – | Ref | – | – | Ref | – | – | – | – |
| 3-4/week | 1.0925 | 1.0514 | 1.1353 | 1.1021 | 1.0459 | 1.1612 | 1.0875 | 1.0274 | 1.1512 | >0.9999 | 0.3744 |
| Daily/almost daily | 1.0744 | 1.0326 | 1.1180 | 1.0780 | 1.0224 | 1.1366 | 1.0766 | 1.0129 | 1.1444 | >0.9999 | 0.9331 |
| **BMI** ($kg/m^2$) | 0.9419 | 0.9392 | 0.9446 | 0.9390 | 0.9347 | 0.9434 | 0.9437 | 0.9402 | 0.9472 | 0.0071 | 0.0008 |
| **Smoking status** | | | | | | | | | | | |
| Never | Ref | – | – | Ref | – | – | Ref | – | – | – | – |
| Former | 0.7978 | 0.7749 | 0.8213 | 0.7711 | 0.7401 | 0.8034 | 0.8382 | 0.8040 | 0.8739 | <0.0001 | <0.0001 |
| Current | 0.7858 | 0.7506 | 0.8227 | 0.8126 | 0.7637 | 0.8648 | 0.7519 | 0.7024 | 0.8051 | >0.9999 | 0.1703 |

*Note:* Estimates from Model 3 (i.e. including all explanatory variables). Bonferroni-adjusted (~99.9%) confidence intervals. OR = odds ratio; CI = confidence interval; BMI = body mass index. [1]number of days per week engaging in these activities for 10+ minutes continuously.

Supplement e19-D. **Environmental exposures associated with long-standing illness stratified by sex**

| | All participants | | | Male | | | Female | | | Interaction term | |
|---|---|---|---|---|---|---|---|---|---|---|---|
| Term | OR | Bonferroni-corrected CI | | OR | Bonferroni-corrected CI | | OR | Bonferroni-corrected CI | | $p_{Bonf.}$ | $p_{BH}$ |
| **$PM_{2.5}$** | 0.9677 | 0.9430 | 0.9932 | 0.9629 | 0.9283 | 0.9987 | 0.9713 | 0.9362 | 1.0079 | >0.9999 | 0.0721 |
| **$PM_{10}$** | 1.0019 | 0.9934 | 1.0105 | 1.0004 | 0.9884 | 1.0125 | 1.0039 | 0.9917 | 1.0162 | >0.9999 | 0.3238 |
| **$NO_2$** | 1.0025 | 0.9982 | 1.0068 | 1.0011 | 0.9951 | 1.0071 | 1.0042 | 0.9981 | 1.0104 | >0.9999 | 0.3424 |
| **$L_{den}$** | 1.0015 | 0.9979 | 1.0052 | 1.0037 | 0.9986 | 1.0089 | 0.9994 | 0.9942 | 1.0046 | >0.9999 | 0.4917 |
| **Greenspace** 1000m | 0.9997 | 0.9987 | 1.0007 | 0.9992 | 0.9978 | 1.0006 | 1.0001 | 0.9988 | 1.0015 | >0.9999 | 0.9754 |

*Note:* Estimates from Model 3 (i.e. including all explanatory variables). Bonferroni-adjusted (~99.9%) confidence intervals. OR = odds ratio; CI = confidence interval; PM = particulate matter; $NO_2$ = nitrogen dioxide; $L_{den}$ = day-evening-night noise level.

**Supplement e20. Confidence interval plots long-standing illness stratified by sex**

Sociodemographic and psychosocial factors

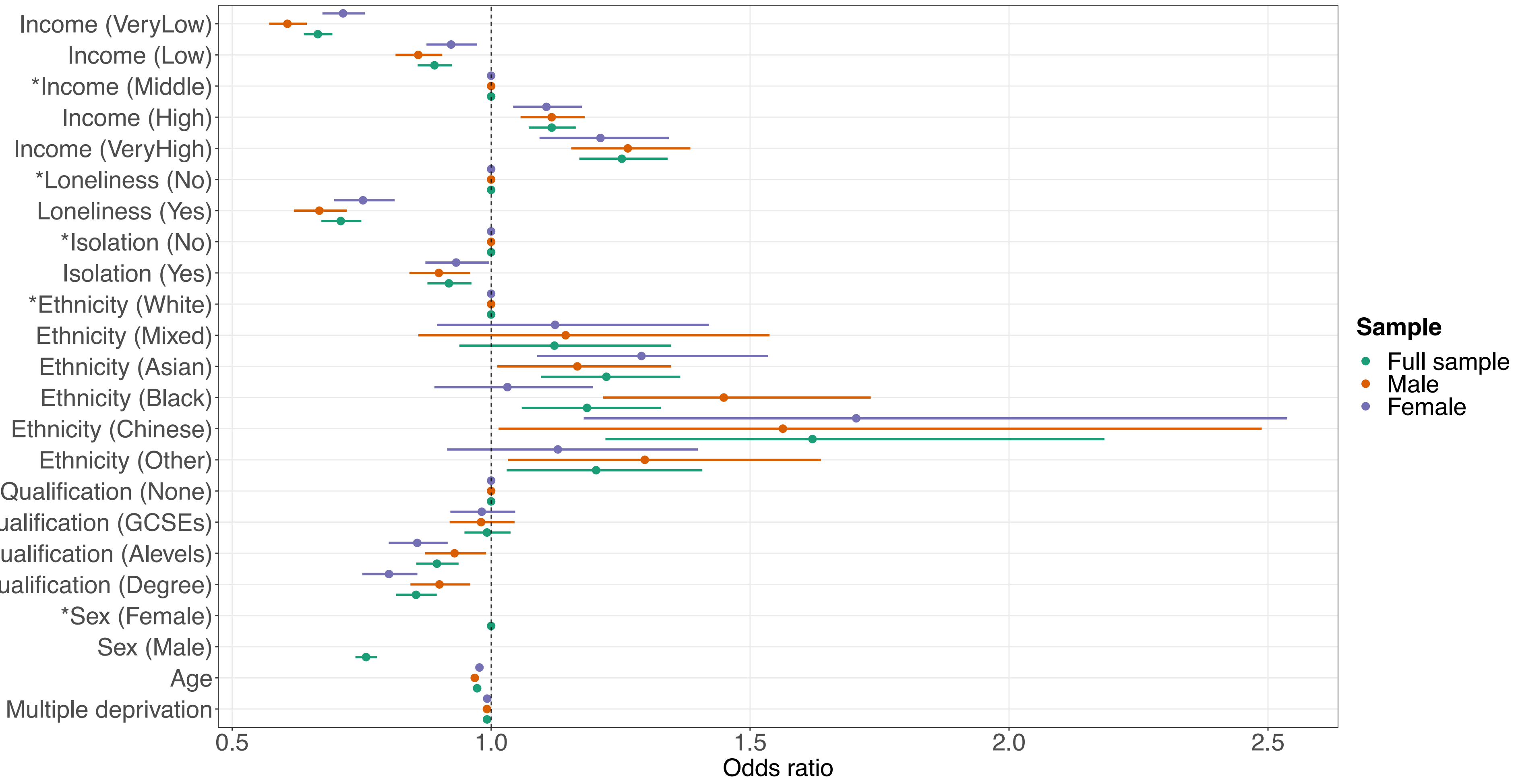


**Supplement e20-A.** Sociodemographic characteristics and psychosocial factors associated with long-standing illness, stratified by sex. Confidence interval plot (odds ratio ± Bonferroni-adjusted (~99.9%) confidence intervals) for Model 3 (i.e. including all explanatory variables). GCSEs = general certificate of secondary education. *Indicates reference group for categorical explanatory variables. Annual household income groups: very low (<£18 000), low (£18 000–30 999), middle (£31 000–51 999), high (£52 000–100 000) and very high (>£100 000). 'GCSEs' also includes O levels and certificate of secondary education (CSE). 'A levels' also includes national vocational qualification (NVQ), higher national diploma (HND), higher national certificate (HNC) and 'other professional qualifications'.

Lifestyle factors

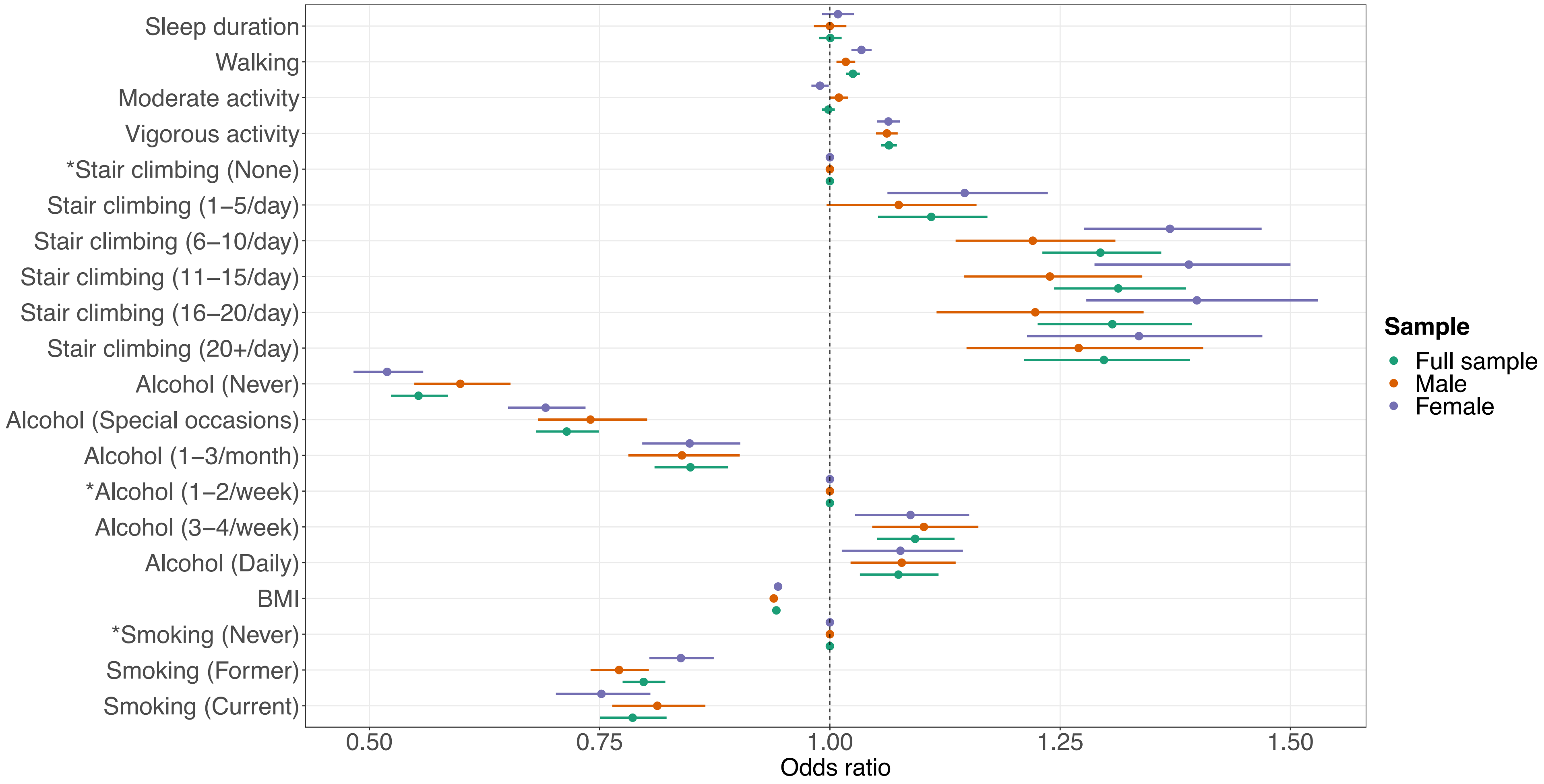


**Supplement e20-B.** Lifestyle factors associated with long-standing illness, stratified by sex. Confidence interval plot (odds ratio ± Bonferroni-adjusted (~99.9%) confidence intervals) for Model 3 (i.e. including all explanatory variables). BMI = body mass index. *Indicates reference group for categorical explanatory variables.

Environmental exposures

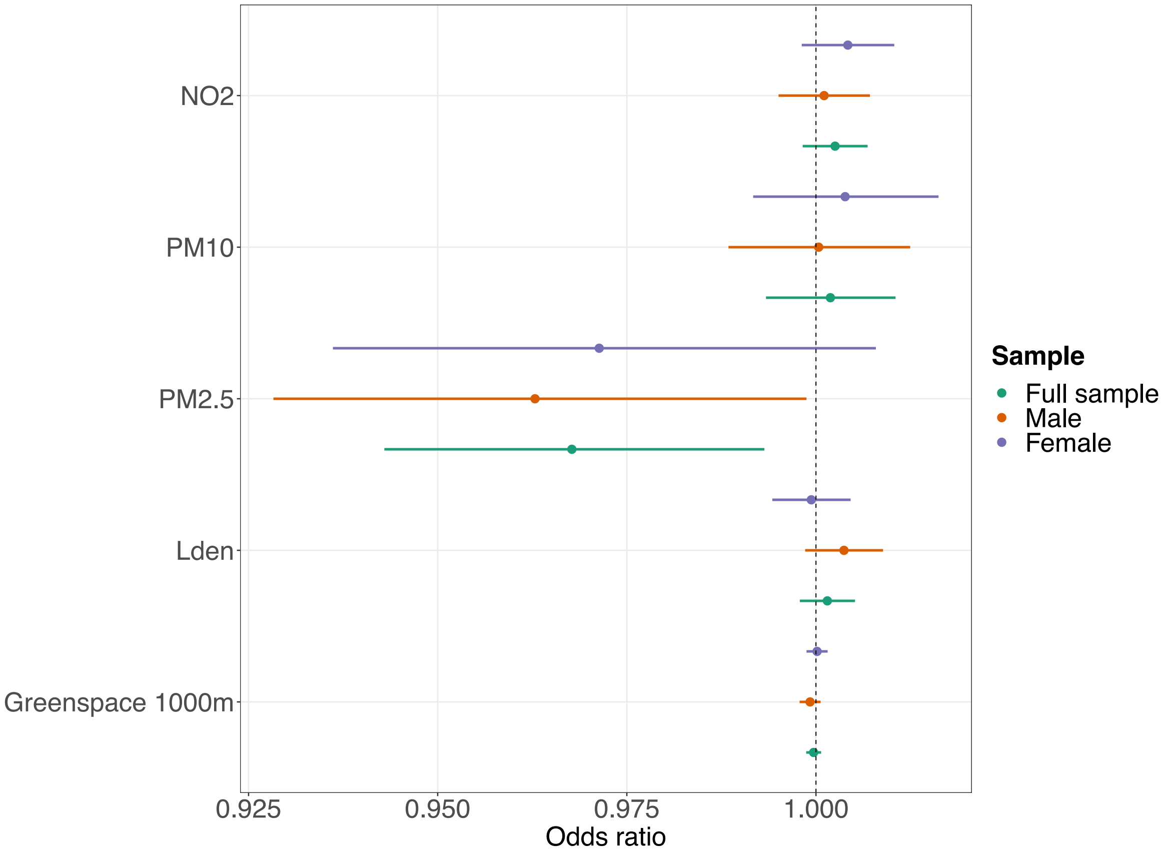


**Supplement e20-C.** Environmental exposures associated with long-standing illness, stratified by sex. Confidence interval plot (odds ratio ± Bonferroni-adjusted (~99.9%) confidence intervals) for Model 3 (i.e. including all explanatory variables). PM = particulate matter; $NO_2$ = nitrogen dioxide; $L_{den}$ = day-evening-night noise level.

**Supplement e21. Regression tables long-standing illness stratified by age**

Sociodemographic characteristics

Supplement e21-A. **Sociodemographic characteristics associated with long-standing illness stratified by age**

| | All participants | | | Below 65 | | | 65 and above | | | Interaction term | |
|---|---|---|---|---|---|---|---|---|---|---|---|
| Term | OR | Bonferroni-corrected CI | | OR | Bonferroni-corrected CI | | OR | Bonferroni-corrected CI | | $p_{Bonf.}$ | $p_{BH}$ |
| **Household income**[1] | | | | | | | | | | | |
| Very low | 0.6654 | 0.6384 | 0.6934 | 0.6115 | 0.5837 | 0.6405 | 0.8485 | 0.7712 | 0.9334 | <0.0001 | <0.0001 |
| Low | 0.8906 | 0.8581 | 0.9244 | 0.8888 | 0.8527 | 0.9263 | 0.9497 | 0.8691 | 1.0375 | 0.1447 | 0.0072 |
| Middle | Ref | – | – | Ref | – | – | Ref | – | – | – | – |
| High | 1.1171 | 1.0726 | 1.1634 | 1.1103 | 1.0637 | 1.1590 | 1.0561 | 0.9228 | 1.2096 | >0.9999 | 0.0776 |
| Very high | 1.2524 | 1.1703 | 1.3410 | 1.2450 | 1.1599 | 1.3372 | 1.1402 | 0.8881 | 1.4724 | >0.9999 | 0.0720 |
| **Sex** | | | | | | | | | | | |
| Female | Ref | – | – | Ref | – | – | Ref | – | – | – | – |
| Male | 0.7585 | 0.7380 | 0.7797 | 0.7670 | 0.7439 | 0.7909 | 0.7216 | 0.6770 | 0.7690 | <0.0001 | <0.0001 |
| **Age** | 0.9730 | 0.9711 | 0.9748 | 0.9708 | 0.9685 | 0.9730 | 0.9752 | 0.9558 | 0.9950 | – | – |
| **Multiple deprivation** | 0.9921 | 0.9910 | 0.9932 | 0.9922 | 0.9910 | 0.9935 | 0.9919 | 0.9893 | 0.9945 | >0.9999 | 0.6879 |
| **Ethnicity** | | | | | | | | | | | |
| White | Ref | – | – | Ref | – | – | Ref | – | – | – | – |
| Mixed-race | 1.1223 | 0.9387 | 1.3474 | 1.0957 | 0.9097 | 1.3257 | 1.4796 | 0.7911 | 2.9027 | >0.9999 | 0.9750 |
| Asian | 1.2225 | 1.0962 | 1.3652 | 1.2957 | 1.1502 | 1.4621 | 0.9176 | 0.6924 | 1.2196 | <0.0001 | <0.0001 |
| Black | 1.1852 | 1.0591 | 1.3277 | 1.2185 | 1.0824 | 1.3735 | 0.9258 | 0.6330 | 1.3562 | 0.0015 | 0.0001 |
| Chinese | 1.6205 | 1.2207 | 2.1842 | 1.6305 | 1.2120 | 2.2309 | 1.7656 | 0.6992 | 5.1866 | >0.9999 | 0.3689 |
| Other | 1.2027 | 1.0301 | 1.4078 | 1.2814 | 1.0861 | 1.5165 | 0.7729 | 0.4805 | 1.2433 | <0.0001 | <0.0001 |
| **Highest qualification** | | | | | | | | | | | |
| None | Ref | – | – | Ref | – | – | Ref | – | – | – | – |
| O levels/GCSEs/CSEs | 0.9920 | 0.9485 | 1.0375 | 1.0113 | 0.9588 | 1.0665 | 1.0088 | 0.9242 | 1.1011 | <0.0001 | <0.0001 |
| A levels/NVQ/HND/HNC[2] | 0.8953 | 0.8554 | 0.9371 | 0.9185 | 0.8699 | 0.9697 | 0.8943 | 0.8193 | 0.9760 | <0.0001 | <0.0001 |
| Degree | 0.8550 | 0.8167 | 0.8950 | 0.8775 | 0.8312 | 0.9261 | 0.8600 | 0.7843 | 0.9430 | <0.0001 | <0.0001 |

*Note:* Estimates from Model 3 (i.e. including all explanatory variables). Bonferroni-adjusted (~99.9%) confidence intervals. OR = odds ratio; CI = confidence interval; GCSEs = general certificate of secondary education; CSE = certificate of secondary education; NVQ = national vocational qualification; HND = higher national diploma; HNC = higher national certificate. [1]Annual household income groups: very low (<£18 000), low (£18 000–30 999), middle (£31 000–51 999), high (£52 000–100 000) and very high (>£100 000). [2]also includes 'other professional qualifications'.

| Supplement e21-B. **Psychosocial factors associated with long-standing illness stratified by age** | | | | | | | | | | | |
|---|---|---|---|---|---|---|---|---|---|---|---|
| | All participants | | | Below 65 | | | 65 and above | | | Interaction term | |
| Term | OR | Bonferroni-corrected CI | | OR | Bonferroni-corrected CI | | OR | Bonferroni-corrected CI | | $p_{Bonf.}$ | $p_{BH}$ |
| **Loneliness** | | | | | | | | | | | |
| Not lonely | Ref | – | – | Ref | – | – | Ref | – | – | – | – |
| Lonely | 0.7097 | 0.6722 | 0.7494 | 0.7077 | 0.6670 | 0.7511 | 0.7328 | 0.6401 | 0.8392 | >0.9999 | 0.0776 |
| **Social isolation** | | | | | | | | | | | |
| Not isolated | Ref | – | – | Ref | – | – | Ref | – | – | – | – |
| Isolated | 0.9185 | 0.8770 | 0.9621 | 0.9168 | 0.8711 | 0.9650 | 0.9448 | 0.8468 | 1.0546 | 0.0001 | <0.0001 |

*Note:* Estimates from Model 3 (i.e. including all explanatory variables). Bonferroni-adjusted (~99.9%) confidence intervals. OR = odds ratio; CI = confidence interval.

Supplement e21-C. **Lifestyle factors associated with long-standing illness stratified by age**

| | All participants | | | Below 65 | | | 65 and above | | | Interaction term | |
|---|---|---|---|---|---|---|---|---|---|---|---|
| Term | OR | Bonferroni-corrected CI | | OR | Bonferroni-corrected CI | | OR | Bonferroni-corrected CI | | $p_{Bonf.}$ | $p_{BH}$ |
| **Sleep duration** (hours/day) | 1.0004 | 0.9882 | 1.0128 | 1.0010 | 0.9872 | 1.0150 | 1.0031 | 0.9766 | 1.0302 | 0.0758 | 0.0042 |
| **Physical activity** (days/week)[1] | | | | | | | | | | | |
| Walking | 1.0250 | 1.0175 | 1.0325 | 1.0215 | 1.0134 | 1.0297 | 1.0441 | 1.0257 | 1.0628 | <0.0001 | <0.0001 |
| Moderate activity | 0.9984 | 0.9916 | 1.0053 | 0.9958 | 0.9881 | 1.0036 | 1.0081 | 0.9931 | 1.0232 | <0.0001 | <0.0001 |
| Vigorous activity | 1.0641 | 1.0557 | 1.0727 | 1.0665 | 1.0569 | 1.0762 | 1.0551 | 1.0372 | 1.0733 | >0.9999 | 0.9750 |
| **Stair climbing frequency** | | | | | | | | | | | |
| None | Ref | – | – | Ref | – | – | Ref | – | – | – | – |
| 1-5/day | 1.1101 | 1.0522 | 1.1710 | 1.1256 | 1.0576 | 1.1979 | 1.0512 | 0.9437 | 1.1708 | <0.0001 | <0.0001 |
| 6-10/day | 1.2937 | 1.2307 | 1.3598 | 1.2962 | 1.2222 | 1.3745 | 1.2988 | 1.1801 | 1.4292 | 0.0002 | <0.0001 |
| 11-15/day | 1.3131 | 1.2434 | 1.3866 | 1.3104 | 1.2296 | 1.3964 | 1.3528 | 1.2139 | 1.5075 | 0.0441 | 0.0026 |
| 16-20/day | 1.3066 | 1.2255 | 1.3932 | 1.2799 | 1.1890 | 1.3778 | 1.4677 | 1.2837 | 1.6790 | >0.9999 | 0.0631 |
| 20+/day | 1.2975 | 1.2108 | 1.3907 | 1.2935 | 1.1957 | 1.3996 | 1.3680 | 1.1744 | 1.5952 | >0.9999 | 0.1379 |
| **Alcohol intake frequency** | | | | | | | | | | | |
| Never | 0.5533 | 0.5234 | 0.5850 | 0.5390 | 0.5062 | 0.5739 | 0.6084 | 0.5390 | 0.6867 | 0.2955 | 0.0141 |
| Special occasions | 0.7142 | 0.6809 | 0.7492 | 0.7070 | 0.6702 | 0.7458 | 0.7452 | 0.6691 | 0.8299 | 0.0211 | 0.0013 |
| 1-3/month | 0.8486 | 0.8096 | 0.8895 | 0.8408 | 0.7984 | 0.8855 | 0.8828 | 0.7877 | 0.9896 | 0.1011 | 0.0053 |
| 1-2/week | Ref | – | – | Ref | – | – | Ref | – | – | – | – |
| 3-4/week | 1.0925 | 1.0514 | 1.1353 | 1.0794 | 1.0345 | 1.1262 | 1.1667 | 1.0659 | 1.2770 | >0.9999 | 0.9558 |
| Daily/almost daily | 1.0744 | 1.0326 | 1.1180 | 1.0720 | 1.0252 | 1.1211 | 1.1248 | 1.0303 | 1.2279 | >0.9999 | 0.4071 |
| **BMI** ($kg/m^2$) | 0.9419 | 0.9392 | 0.9446 | 0.9425 | 0.9396 | 0.9455 | 0.9392 | 0.9324 | 0.9461 | <0.0001 | <0.0001 |
| **Smoking status** | | | | | | | | | | | |
| Never | Ref | – | – | Ref | – | – | Ref | – | – | – | – |
| Former | 0.7978 | 0.7749 | 0.8213 | 0.8114 | 0.7853 | 0.8385 | 0.7592 | 0.7120 | 0.8094 | <0.0001 | <0.0001 |
| Current | 0.7858 | 0.7506 | 0.8227 | 0.7824 | 0.7447 | 0.8222 | 0.8565 | 0.7578 | 0.9687 | >0.9999 | 0.2871 |

*Note:* Estimates from Model 3 (i.e. including all explanatory variables). Bonferroni-adjusted (~99.9%) confidence intervals. OR = odds ratio; CI = confidence interval; BMI = body mass index. [1]number of days per week engaging in these activities for 10+ minutes continuously.

Environmental exposures

Supplement e21-D. **Environmental exposures associated with long-standing illness stratified by age**

| | All participants | | | Below 65 | | | 65 and above | | | Interaction term | |
|---|---|---|---|---|---|---|---|---|---|---|---|
| Term | OR | Bonferroni-corrected CI | | OR | Bonferroni-corrected CI | | OR | Bonferroni-corrected CI | | $p_{Bonf.}$ | $p_{BH}$ |
| **$PM_{2.5}$** | 0.9677 | 0.9430 | 0.9932 | 0.9704 | 0.9428 | 0.9988 | 0.9517 | 0.8966 | 1.0102 | >0.9999 | 0.2850 |
| **$PM_{10}$** | 1.0019 | 0.9934 | 1.0105 | 1.0016 | 0.9921 | 1.0112 | 1.0028 | 0.9834 | 1.0226 | >0.9999 | 0.9425 |
| **$NO_2$** | 1.0025 | 0.9982 | 1.0068 | 1.0025 | 0.9977 | 1.0073 | 1.0036 | 0.9938 | 1.0134 | >0.9999 | 0.3335 |
| **$L_{den}$** | 1.0015 | 0.9979 | 1.0052 | 1.0020 | 0.9980 | 1.0061 | 0.9995 | 0.9911 | 1.0079 | >0.9999 | 0.0776 |
| **Greenspace** 1000m | 0.9997 | 0.9987 | 1.0007 | 0.9998 | 0.9987 | 1.0009 | 0.9995 | 0.9972 | 1.0018 | >0.9999 | 0.6879 |

*Note:* Estimates from Model 3 (i.e. including all explanatory variables). Bonferroni-adjusted (~99.9%) confidence intervals. OR = odds ratio; CI = confidence interval; PM = particulate matter; $NO_2$ = nitrogen dioxide; $L_{den}$ = day-evening-night noise level.

**Supplement e22. Confidence interval plots long-standing illness stratified by age**

Sociodemographic and psychosocial factors

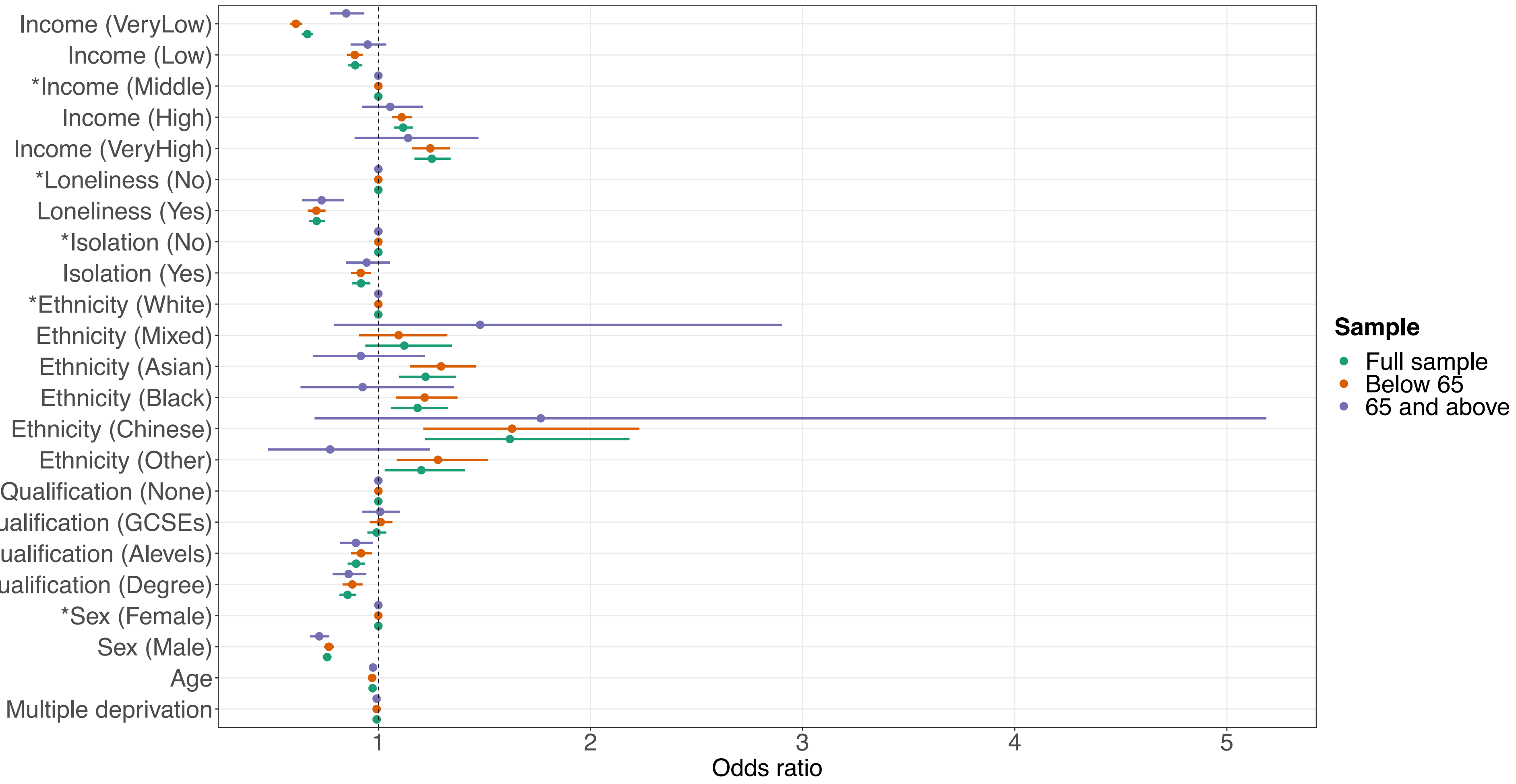


**Supplement e22-A.** Sociodemographic characteristics and psychosocial factors associated with long-standing illness, stratified by age. Confidence interval plot (odds ratio ± Bonferroni-adjusted (~99.9%) confidence intervals) for Model 3 (i.e. including all explanatory variables). GCSEs = general certificate of secondary education. *Indicates reference group for categorical explanatory variables. Annual household income groups: very low (<£18 000), low (£18 000–30 999), middle (£31 000–51 999), high (£52 000–100 000) and very high (>£100 000). 'GCSEs' also includes O levels and certificate of secondary education (CSE). 'A levels' also includes national vocational qualification (NVQ), higher national diploma (HND), higher national certificate (HNC) and 'other professional qualifications'.

Lifestyle factors

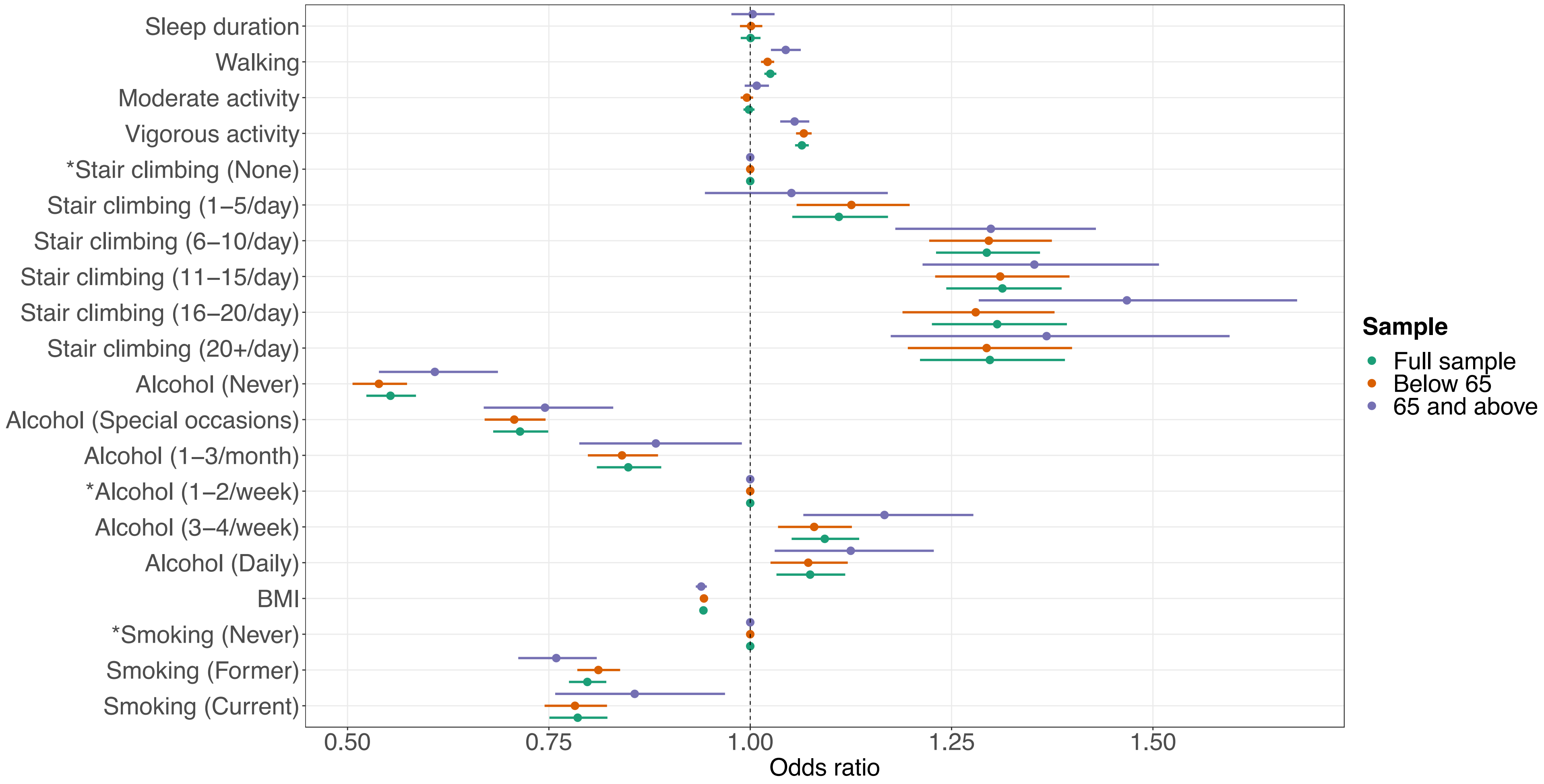


**Supplement e22-B.** Lifestyle factors associated with long-standing illness, stratified by age. Confidence interval plot (odds ratio ± Bonferroni-adjusted (~99.9%) confidence intervals) for Model 3 (i.e. including all explanatory variables). BMI = body mass index. *Indicates reference group for categorical explanatory variables.

Environmental exposures

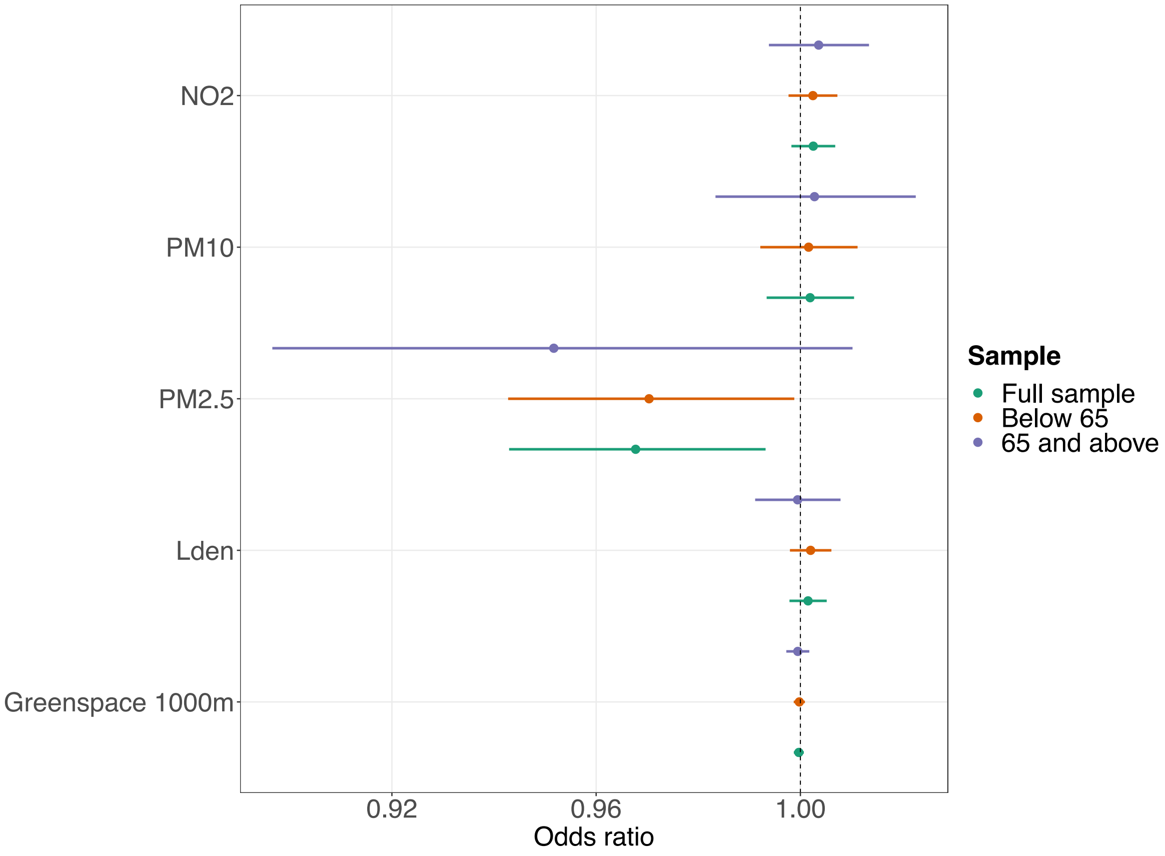


**Supplement e22-C.** Environmental exposures associated with long-standing illness, stratified by age. Confidence interval plot (odds ratio ± Bonferroni-adjusted (~99.9%) confidence intervals) for Model 3 (i.e. including all explanatory variables). PM = particulate matter; $NO_2$ = nitrogen dioxide; $L_{den}$ = day-evening-night noise level.

**Supplement e23. Regression tables self-rated health stratified by sex**

Sociodemographic characteristics

Supplement e23-A. **Sociodemographic characteristics associated with self-rated health stratified by sex**

| | All participants | | | Male | | | Female | | | Interaction term | |
|---|---|---|---|---|---|---|---|---|---|---|---|
| Term | OR | Bonferroni-corrected CI | | OR | Bonferroni-corrected CI | | OR | Bonferroni-corrected CI | | $p_{Bonf.}$ | $p_{BH}$ |
| **Household income**[1] | | | | | | | | | | | |
| Very low | 0.6192 | 0.5960 | 0.6432 | 0.5733 | 0.5418 | 0.6065 | 0.6505 | 0.6175 | 0.6852 | <0.0001 | <0.0001 |
| Low | 0.8512 | 0.8234 | 0.8799 | 0.8300 | 0.7912 | 0.8706 | 0.8650 | 0.8259 | 0.9059 | 0.1481 | 0.0067 |
| Middle | Ref | – | – | Ref | – | – | Ref | – | – | – | – |
| High | 1.2102 | 1.1691 | 1.2526 | 1.2497 | 1.1910 | 1.3113 | 1.1722 | 1.1156 | 1.2318 | 0.0002 | <0.0001 |
| Very high | 1.6364 | 1.5491 | 1.7286 | 1.7228 | 1.5975 | 1.8580 | 1.5552 | 1.4361 | 1.6842 | <0.0001 | <0.0001 |
| **Sex** | | | | | | | | | | | |
| Female | Ref | – | – | Ref | – | – | Ref | – | – | – | – |
| Male | 0.7836 | 0.7648 | 0.8030 | – | – | – | – | – | – | – | – |
| **Age** | 1.0113 | 1.0097 | 1.0130 | 1.0118 | 1.0094 | 1.0142 | 1.0108 | 1.0084 | 1.0132 | <0.0001 | <0.0001 |
| **Multiple deprivation** | 0.9923 | 0.9913 | 0.9933 | 0.9915 | 0.9901 | 0.9930 | 0.9932 | 0.9917 | 0.9946 | <0.0001 | <0.0001 |
| **Ethnicity** | | | | | | | | | | | |
| White | Ref | – | – | Ref | – | – | Ref | – | – | – | – |
| Mixed-race | 0.9019 | 0.7729 | 1.0527 | 1.0413 | 0.8098 | 1.3399 | 0.8229 | 0.6762 | 1.0020 | 0.5618 | 0.0208 |
| Asian | 0.5995 | 0.5445 | 0.6602 | 0.6139 | 0.5407 | 0.6970 | 0.5739 | 0.4948 | 0.6658 | 0.8350 | 0.0288 |
| Black | 1.0896 | 0.9839 | 1.2069 | 1.4325 | 1.2258 | 1.6745 | 0.8748 | 0.7643 | 1.0017 | <0.0001 | <0.0001 |
| Chinese | 0.6783 | 0.5413 | 0.8506 | 0.7109 | 0.4948 | 1.0236 | 0.6773 | 0.5069 | 0.9063 | >0.9999 | 0.2264 |
| Other | 0.8903 | 0.7746 | 1.0235 | 1.1508 | 0.9361 | 1.4154 | 0.7210 | 0.5972 | 0.8710 | <0.0001 | <0.0001 |
| **Highest qualification** | | | | | | | | | | | |
| None | Ref | – | – | Ref | – | – | Ref | – | – | – | – |
| O levels/GCSEs/CSEs | 1.2420 | 1.1920 | 1.2941 | 1.2168 | 1.1471 | 1.2908 | 1.2412 | 1.1716 | 1.3149 | 0.7710 | 0.0275 |
| A levels/NVQ/HND/HNC[2] | 1.2876 | 1.2345 | 1.3429 | 1.2761 | 1.2030 | 1.3536 | 1.2892 | 1.2138 | 1.3692 | 0.0201 | 0.0010 |
| Degree | 1.5031 | 1.4412 | 1.5676 | 1.5429 | 1.4537 | 1.6376 | 1.4286 | 1.3456 | 1.5166 | <0.0001 | <0.0001 |

*Note:* Estimates from Model 3 (i.e. including all explanatory variables). Bonferroni-adjusted (~99.9%) confidence intervals. OR = odds ratio; CI = confidence interval; GCSEs = general certificate of secondary education; CSE = certificate of secondary education; NVQ = national vocational qualification; HND = higher national diploma; HNC = higher national certificate. For categorical explanatory variables the odds ratios indicate the changes in odds of reporting better self-rated health associated with the explanatory variable group relative to the reference group. Odds ratios for continuous explanatory variables indicate proportional odds ratios for a 1-unit increase in the explanatory variable on level of self-rated health. [1]Annual household income groups: very low (<£18 000), low (£18 000–30 999), middle (£31 000–51 999), high (£52 000–100 000) and very high (>£100 000). [2]also includes 'other professional qualifications'.

Psychosocial factors

Supplement e23-B. **Psychosocial factors associated with self-rated health stratified by sex**

| | All participants | | | Male | | | Female | | | Interaction term | |
|---|---|---|---|---|---|---|---|---|---|---|---|
| Term | OR | Bonferroni-corrected CI | | OR | Bonferroni-corrected CI | | OR | Bonferroni-corrected CI | | $p_{Bonf.}$ | $p_{BH}$ |
| **Loneliness** | | | | | | | | | | | |
| Not lonely | Ref | – | – | Ref | – | – | Ref | – | – | – | – |
| Lonely | 0.4921 | 0.4678 | 0.5176 | 0.4901 | 0.4564 | 0.5263 | 0.4971 | 0.4626 | 0.5342 | 0.0172 | 0.0009 |
| **Social isolation** | | | | | | | | | | | |
| Not isolated | Ref | – | – | Ref | – | – | Ref | – | – | – | – |
| Isolated | 0.8593 | 0.8233 | 0.8970 | 0.8311 | 0.7821 | 0.8831 | 0.8805 | 0.8286 | 0.9358 | <0.0001 | <0.0001 |

*Note:* Estimates from Model 3 (i.e. including all explanatory variables). Bonferroni-adjusted (~99.9%) confidence intervals. OR = odds ratio; CI = confidence interval. For categorical explanatory variables the odds ratios indicate the changes in odds of reporting better self-rated health associated with the explanatory variable group relative to the reference group.

Lifestyle factors

Supplement e23-C. **Lifestyle factors associated with self-rated health stratified by sex**

| | All participants | | | Male | | | Female | | | Interaction term | |
|---|---|---|---|---|---|---|---|---|---|---|---|
| Term | OR | Bonferroni-corrected CI | | OR | Bonferroni-corrected CI | | OR | Bonferroni-corrected CI | | $p_{Bonf.}$ | $p_{BH}$ |
| **Sleep duration** (hours/day) | 1.0730 | 1.0609 | 1.0852 | 1.0685 | 1.0510 | 1.0864 | 1.0812 | 1.0644 | 1.0983 | >0.9999 | 0.1448 |
| **Physical activity** (days/week)[1] | | | | | | | | | | | |
| Walking | 1.0522 | 1.0454 | 1.0591 | 1.0477 | 1.0382 | 1.0573 | 1.0577 | 1.0477 | 1.0677 | 0.1603 | 0.0070 |
| Moderate activity | 1.0132 | 1.0070 | 1.0195 | 1.0256 | 1.0164 | 1.0350 | 1.0050 | 0.9966 | 1.0134 | <0.0001 | <0.0001 |
| Vigorous activity | 1.1668 | 1.1585 | 1.1751 | 1.1766 | 1.1648 | 1.1886 | 1.1506 | 1.1390 | 1.1624 | <0.0001 | <0.0001 |
| **Stair climbing frequency** | | | | | | | | | | | |
| None | Ref | – | – | Ref | – | – | Ref | – | – | – | – |
| 1-5/day | 1.0533 | 1.0019 | 1.1074 | 1.0459 | 0.9744 | 1.1226 | 1.0578 | 0.9854 | 1.1355 | >0.9999 | 0.5955 |
| 6-10/day | 1.2293 | 1.1733 | 1.2878 | 1.2207 | 1.1422 | 1.3045 | 1.2290 | 1.1511 | 1.3121 | 0.4963 | 0.0191 |
| 11-15/day | 1.2892 | 1.2260 | 1.3557 | 1.3136 | 1.2220 | 1.4119 | 1.2642 | 1.1785 | 1.3562 | 0.0001 | <0.0001 |
| 16-20/day | 1.2991 | 1.2258 | 1.3767 | 1.2994 | 1.1944 | 1.4135 | 1.3008 | 1.2004 | 1.4097 | 0.0168 | 0.0009 |
| 20+/day | 1.3652 | 1.2828 | 1.4529 | 1.4561 | 1.3287 | 1.5956 | 1.3054 | 1.1986 | 1.4217 | <0.0001 | <0.0001 |
| **Alcohol intake frequency** | | | | | | | | | | | |
| Never | 0.6624 | 0.6285 | 0.6981 | 0.7264 | 0.6688 | 0.7890 | 0.6123 | 0.5717 | 0.6558 | 0.0033 | 0.0002 |
| Special occasions | 0.7565 | 0.7240 | 0.7904 | 0.7772 | 0.7214 | 0.8373 | 0.7253 | 0.6861 | 0.7666 | >0.9999 | 0.6945 |
| 1-3/month | 0.9201 | 0.8821 | 0.9597 | 0.9173 | 0.8591 | 0.9795 | 0.9101 | 0.8610 | 0.9621 | >0.9999 | 0.4250 |
| 1-2/week | Ref | – | – | Ref | – | – | Ref | – | – | – | – |
| 3-4/week | 1.0520 | 1.0178 | 1.0875 | 1.0396 | 0.9926 | 1.0888 | 1.0700 | 1.0203 | 1.1222 | >0.9999 | 0.5955 |
| Daily/almost daily | 1.0264 | 0.9913 | 1.0628 | 0.9998 | 0.9535 | 1.0484 | 1.0609 | 1.0074 | 1.1172 | >0.9999 | 0.3959 |
| **BMI** (kg/m$^2$) | 0.9045 | 0.9020 | 0.9069 | 0.8908 | 0.8870 | 0.8946 | 0.9119 | 0.9087 | 0.9150 | <0.0001 | <0.0001 |
| **Smoking status** | | | | | | | | | | | |
| Never | Ref | – | – | Ref | – | – | Ref | – | – | – | – |
| Former | 0.8128 | 0.7919 | 0.8342 | 0.7768 | 0.7484 | 0.8063 | 0.8619 | 0.8309 | 0.8941 | <0.0001 | <0.0001 |
| Current | 0.5029 | 0.4825 | 0.5242 | 0.5007 | 0.4733 | 0.5297 | 0.5005 | 0.4706 | 0.5322 | 0.0015 | 0.0001 |

*Note:* Estimates from Model 3 (i.e. including all explanatory variables). Bonferroni-adjusted (~99.9%) confidence intervals. OR = odds ratio; CI = confidence interval; BMI = body mass index. For categorical explanatory variables the odds ratios indicate the changes in odds of reporting better self-rated health associated with the explanatory variable group relative to the reference group. Odds ratios for continuous explanatory variables indicate proportional odds ratios for a 1-unit increase in the explanatory variable on level of self-rated health. [1]number of days per week engaging in these activities for 10+ minutes continuously.

Supplement e23-D. **Environmental exposures associated with self-rated health stratified by sex**

| | All participants | | | Male | | | Female | | | Interaction term | |
|---|---|---|---|---|---|---|---|---|---|---|---|
| Term | OR | Bonferroni-corrected CI | | OR | Bonferroni-corrected CI | | OR | Bonferroni-corrected CI | | $p_{Bonf.}$ | $p_{BH}$ |
| **$PM_{2.5}$** | 1.0283 | 1.0047 | 1.0525 | 1.0267 | 0.9933 | 1.0612 | 1.0289 | 0.9959 | 1.0630 | 0.0031 | 0.0002 |
| **$PM_{10}$** | 0.9979 | 0.9904 | 1.0055 | 0.9959 | 0.9852 | 1.0067 | 1.0003 | 0.9898 | 1.0110 | 0.2851 | 0.0119 |
| **$NO_2$** | 0.9948 | 0.9910 | 0.9986 | 0.9955 | 0.9901 | 1.0009 | 0.9945 | 0.9892 | 0.9998 | 0.3695 | 0.0148 |
| **$L_{den}$** | 1.0025 | 0.9993 | 1.0058 | 1.0016 | 0.9970 | 1.0063 | 1.0035 | 0.9989 | 1.0080 | >0.9999 | 0.1412 |
| **Greenspace** 1000m | 1.0007 | 0.9998 | 1.0015 | 1.0010 | 0.9998 | 1.0023 | 1.0004 | 0.9992 | 1.0016 | >0.9999 | 0.0889 |

*Note:* Estimates from Model 3 (i.e. including all explanatory variables). Bonferroni-adjusted (~99.9%) confidence intervals. OR = odds ratio; CI = confidence interval; PM = particulate matter; $NO_2$ = nitrogen dioxide; $L_{den}$ = day-evening-night noise level. Odds ratios indicate proportional odds ratios for a 1-unit increase in the explanatory variable on level of self-rated health.

**Supplement e24. Confidence interval plots self-rated health stratified by sex**

Sociodemographic and psychosocial factors

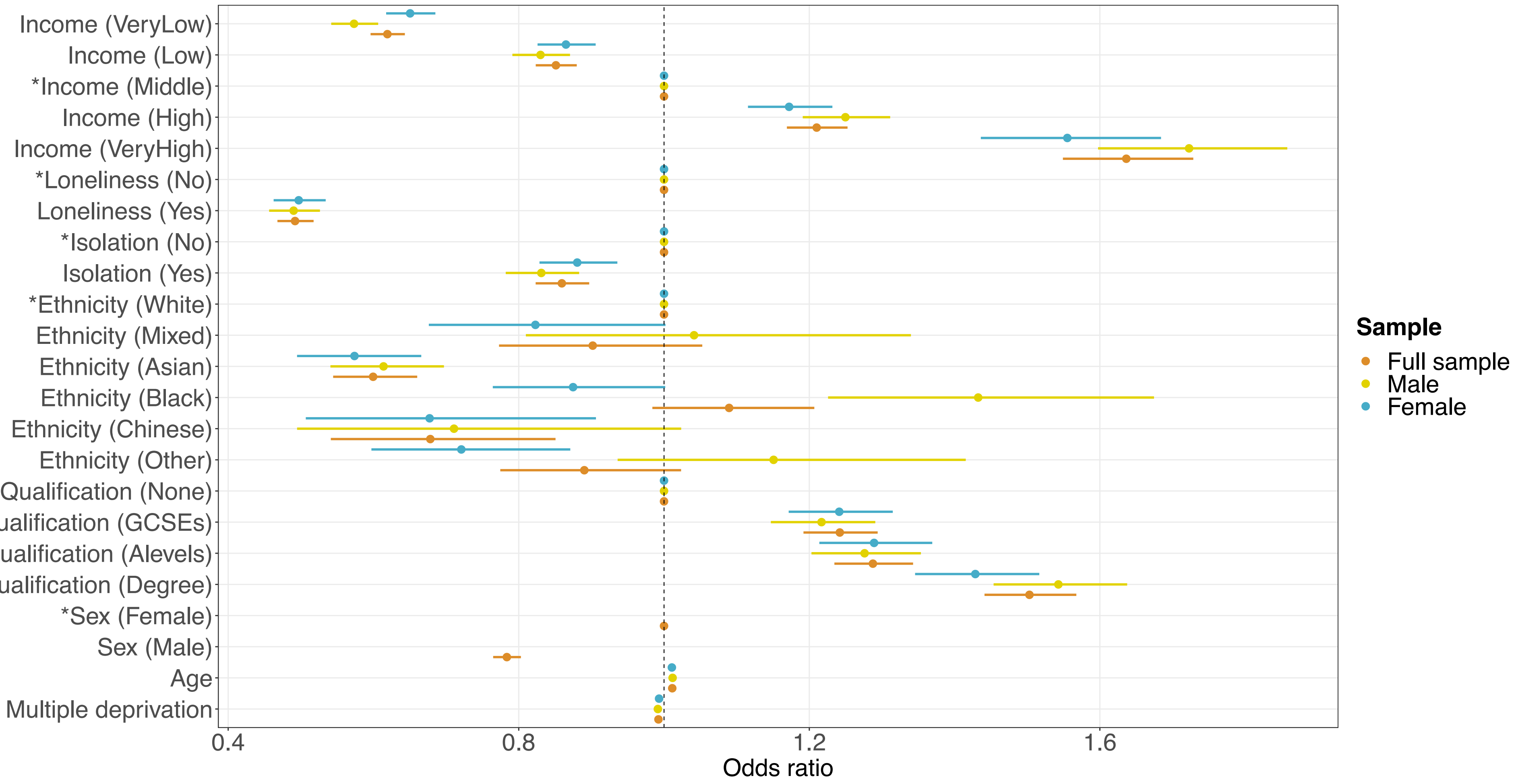


**Supplement e24-A.** Sociodemographic characteristics and psychosocial factors associated with self-rated health, stratified by sex. Confidence interval plot (odds ratio ± Bonferroni-adjusted (~99.9%) confidence intervals) for Model 3 (i.e. including all explanatory variables). GCSEs = general certificate of secondary education. *Indicates reference group for categorical explanatory variables. Annual household income groups: very low (<£18 000), low (£18 000–30 999), middle (£31 000–51 999), high (£52 000–100 000) and very high (>£100 000). 'GCSEs' also includes O levels and certificate of secondary education (CSE). 'A levels' also includes national vocational qualification (NVQ), higher national diploma (HND), higher national certificate (HNC) and 'other professional qualifications'.

Lifestyle factors

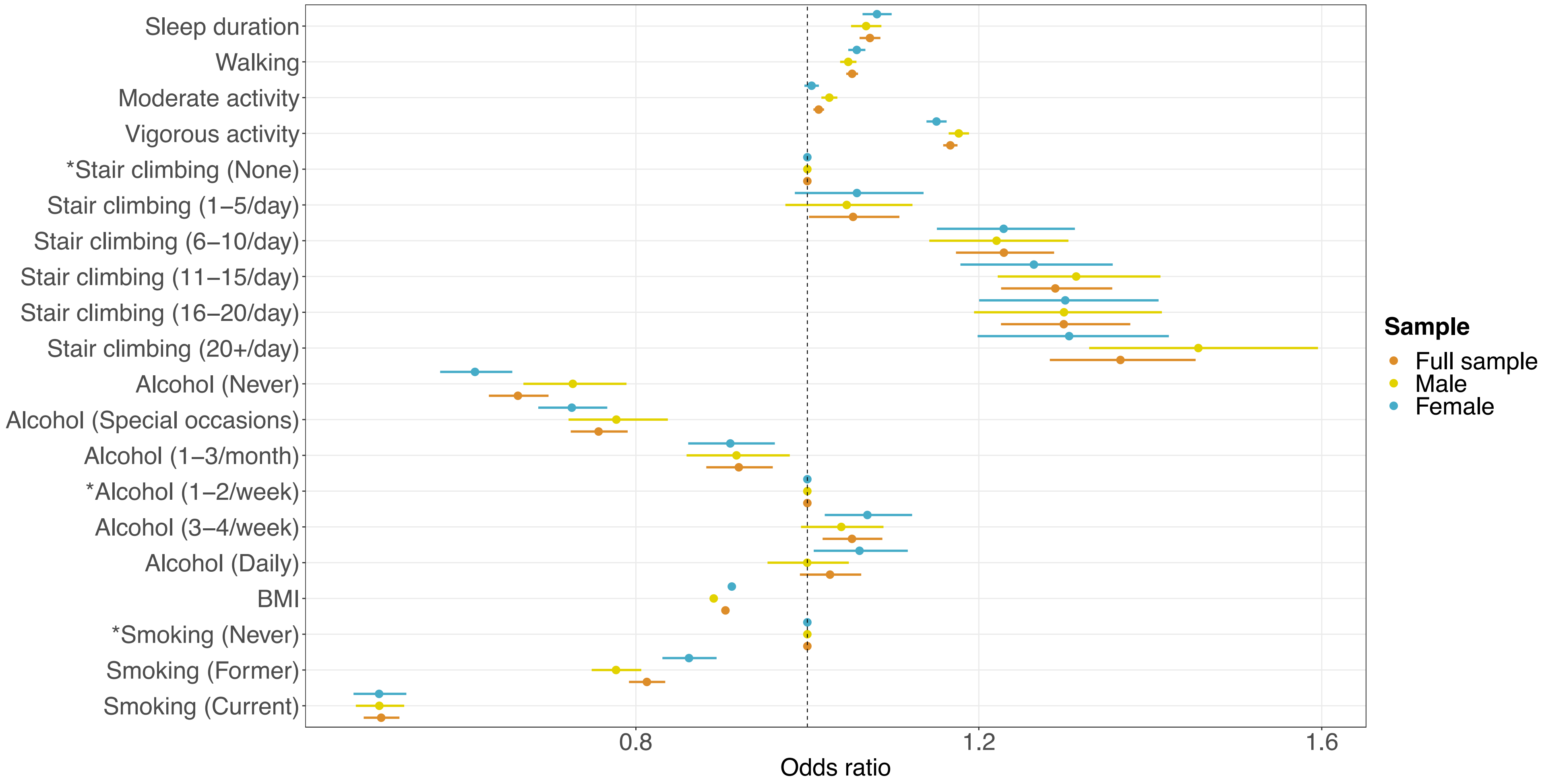


**Supplement e24-B.** Lifestyle factors associated with self-rated health, stratified by sex. Confidence interval plot (odds ratio ± Bonferroni-adjusted (~99.9%) confidence intervals) for Model 3 (i.e. including all explanatory variables). BMI = body mass index. *Indicates reference group for categorical explanatory variables.

Environmental exposures

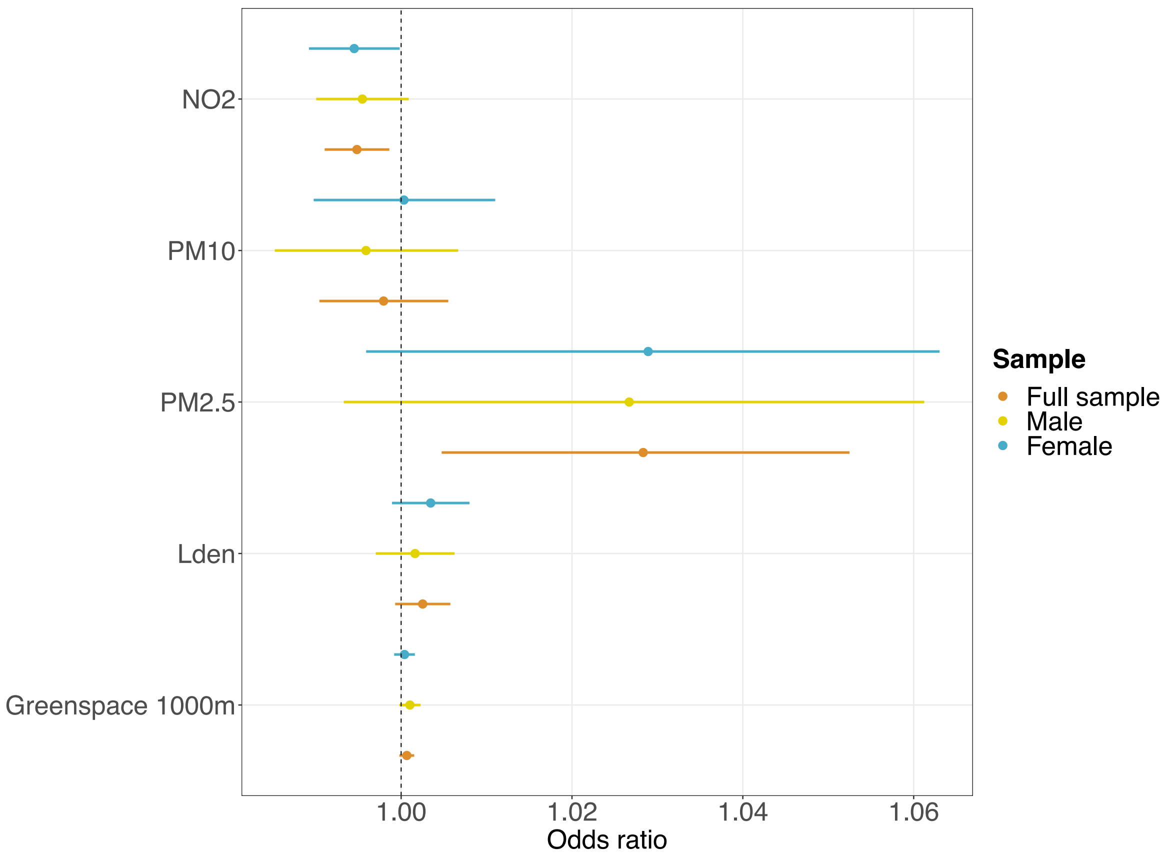


**Supplement e24-C.** Environmental exposures associated with self-rated health, stratified by sex. Confidence interval plot (odds ratio ± Bonferroni-adjusted (~99.9%) confidence intervals) for Model 3 (i.e. including all explanatory variables). PM = particulate matter; $NO_2$ = nitrogen dioxide; $L_{den}$ = day-evening-night noise level.

**Supplement e25. Regression tables self-rated health stratified by age**

Sociodemographic characteristics

Supplement e25-A. **Sociodemographic characteristics associated with self-rated health stratified by age**

| | All participants | | | Below 65 | | | 65 and above | | | Interaction term | |
|---|---|---|---|---|---|---|---|---|---|---|---|
| Term | OR | Bonferroni-corrected CI | | OR | Bonferroni-corrected CI | | OR | Bonferroni-corrected CI | | $p_{Bonf.}$ | $p_{BH}$ |
| **Household income**[1] | | | | | | | | | | | |
| Very low | 0.6192 | 0.5960 | 0.6432 | 0.5873 | 0.5627 | 0.6130 | 0.7057 | 0.6442 | 0.7729 | <0.0001 | <0.0001 |
| Low | 0.8512 | 0.8234 | 0.8799 | 0.8561 | 0.8255 | 0.8879 | 0.8614 | 0.7922 | 0.9367 | 0.6464 | 0.0249 |
| Middle | Ref | – | – | Ref | – | – | Ref | – | – | – | – |
| High | 1.2102 | 1.1691 | 1.2526 | 1.2062 | 1.1635 | 1.2504 | 1.1177 | 0.9844 | 1.2691 | 0.0288 | 0.0014 |
| Very high | 1.6364 | 1.5491 | 1.7286 | 1.6386 | 1.5483 | 1.7341 | 1.2259 | 0.9692 | 1.5498 | 0.0004 | <0.0001 |
| **Sex** | | | | | | | | | | | |
| Female | Ref | – | – | Ref | – | – | Ref | – | – | – | – |
| Male | 0.7836 | 0.7648 | 0.8030 | 0.7808 | 0.7601 | 0.8019 | 0.8118 | 0.7643 | 0.8622 | <0.0001 | <0.0001 |
| **Age** | 1.0113 | 1.0097 | 1.0130 | 1.0120 | 1.0010 | 1.0140 | 0.9978 | 0.9789 | 1.0171 | – | – |
| **Multiple deprivation** | 0.9923 | 0.9913 | 0.9933 | 0.9922 | 0.9911 | 0.9933 | 0.9938 | 0.9913 | 0.9963 | >0.9999 | 0.8416 |
| **Ethnicity** | | | | | | | | | | | |
| White | Ref | – | – | Ref | – | – | Ref | – | – | – | – |
| Mixed-race | 0.9019 | 0.7729 | 1.0527 | 0.9212 | 0.7851 | 1.0811 | 0.7125 | 0.3969 | 1.2876 | >0.9999 | 0.3609 |
| Asian | 0.5995 | 0.5445 | 0.6602 | 0.6297 | 0.5680 | 0.6983 | 0.4443 | 0.3399 | 0.5817 | <0.0001 | <0.0001 |
| Black | 1.0896 | 0.9839 | 1.2069 | 1.1503 | 1.0343 | 1.2796 | 0.6069 | 0.4243 | 0.8709 | <0.0001 | <0.0001 |
| Chinese | 0.6783 | 0.5413 | 0.8506 | 0.6780 | 0.5366 | 0.8573 | 0.6584 | 0.2825 | 1.5576 | >0.9999 | 0.5891 |
| Other | 0.8903 | 0.7746 | 1.0235 | 0.9262 | 0.8005 | 1.0719 | 0.6205 | 0.3924 | 0.9868 | <0.0001 | <0.0001 |
| **Highest qualification** | | | | | | | | | | | |
| None | Ref | – | – | Ref | – | – | Ref | – | – | – | – |
| O levels/GCSEs/CSEs | 1.2420 | 1.1920 | 1.2941 | 1.2790 | 1.2187 | 1.3423 | 1.2593 | 1.1587 | 1.3688 | <0.0001 | <0.0001 |
| A levels/NVQ/HND/HNC[2] | 1.2876 | 1.2345 | 1.3429 | 1.3358 | 1.2713 | 1.4036 | 1.2424 | 1.1423 | 1.3514 | <0.0001 | <0.0001 |
| Degree | 1.5031 | 1.4412 | 1.5676 | 1.5655 | 1.4905 | 1.6443 | 1.4066 | 1.2878 | 1.5364 | <0.0001 | <0.0001 |

*Note:* Estimates from Model 3 (i.e. including all explanatory variables). Bonferroni-adjusted (~99.9%) confidence intervals. OR = odds ratio; CI = confidence interval; GCSEs = general certificate of secondary education; CSE = certificate of secondary education; NVQ = national vocational qualification; HND = higher national diploma; HNC = higher national certificate. For categorical explanatory variables the odds ratios indicate the changes in odds of reporting better self-rated health associated with the explanatory variable group relative to the reference group. Odds ratios for continuous explanatory variables indicate proportional odds ratios for a 1-unit increase in the explanatory variable on level of self-rated health. [1]Annual household income groups: very low (<£18 000), low (£18 000–30 999), middle (£31 000–51 999), high (£52 000–100 000) and very high (>£100 000). [2]also includes 'other professional qualifications'.

Psychosocial factors

| Supplement e25-B. **Psychosocial factors associated with self-rated health stratified by age** | | | | | | | | | | |
|---|---|---|---|---|---|---|---|---|---|---|
| | All participants | | | Below 65 | | | 65 and above | | | Interaction term | |
| Term | OR | Bonferroni-corrected CI | | OR | Bonferroni-corrected CI | | OR | Bonferroni-corrected CI | | $p_{Bonf.}$ | $p_{BH}$ |
| **Loneliness** | | | | | | | | | | | |
| Not lonely | Ref | – | – | Ref | – | – | Ref | – | – | – | – |
| Lonely | 0.4921 | 0.4678 | 0.5176 | 0.4853 | 0.4594 | 0.5127 | 0.5460 | 0.4798 | 0.6216 | 0.0016 | 0.0001 |
| **Social isolation** | | | | | | | | | | | |
| Not isolated | Ref | – | – | Ref | – | – | Ref | – | – | – | – |
| Isolated | 0.8593 | 0.8233 | 0.8970 | 0.8528 | 0.8137 | 0.8938 | 0.9057 | 0.8149 | 1.0067 | 0.0334 | 0.0015 |

*Note:* Estimates from Model 3 (i.e. including all explanatory variables). Bonferroni-adjusted (~99.9%) confidence intervals. OR = odds ratio; CI = confidence interval. For categorical explanatory variables the odds ratios indicate the changes in odds of reporting better self-rated health associated with the explanatory variable group relative to the reference group.

| Supplement e25-C. **Lifestyle factors associated with self-rated health stratified by age** | | | | | | | | | | | |
|---|---|---|---|---|---|---|---|---|---|---|---|
| | All participants | | | Below 65 | | | 65 and above | | | Interaction term | |
| Term | OR | Bonferroni-corrected CI | | OR | Bonferroni-corrected CI | | OR | Bonferroni-corrected CI | | $p_{Bonf.}$ | $p_{BH}$ |
| **Sleep duration** (hours/day) | 1.0730 | 1.0609 | 1.0852 | 1.0790 | 1.0654 | 1.0927 | 1.0453 | 1.0185 | 1.0727 | 0.0193 | 0.0010 |
| **Physical activity** (days/week)[1] | | | | | | | | | | | |
| Walking | 1.0522 | 1.0454 | 1.0591 | 1.0474 | 1.0400 | 1.0548 | 1.0836 | 1.0652 | 1.1024 | 0.0005 | <0.0001 |
| Moderate activity | 1.0132 | 1.0070 | 1.0195 | 1.0112 | 1.0044 | 1.0181 | 1.0218 | 1.0073 | 1.0365 | >0.9999 | 0.3609 |
| Vigorous activity | 1.1668 | 1.1585 | 1.1751 | 1.1745 | 1.1652 | 1.1839 | 1.1352 | 1.1168 | 1.1540 | <0.0001 | <0.0001 |
| **Stair climbing frequency** | | | | | | | | | | | |
| None | Ref | – | – | Ref | – | – | Ref | – | – | – | – |
| 1-5/day | 1.0533 | 1.0019 | 1.1074 | 1.0797 | 1.0192 | 1.1437 | 0.9617 | 0.8660 | 1.0679 | 0.0006 | <0.0001 |
| 6-10/day | 1.2293 | 1.1733 | 1.2878 | 1.2422 | 1.1767 | 1.3114 | 1.2165 | 1.1086 | 1.3348 | 0.4794 | 0.0192 |
| 11-15/day | 1.2892 | 1.2260 | 1.3557 | 1.2980 | 1.2249 | 1.3755 | 1.3092 | 1.1794 | 1.4532 | >0.9999 | 0.0663 |
| 16-20/day | 1.2991 | 1.2258 | 1.3767 | 1.3031 | 1.2198 | 1.3921 | 1.3533 | 1.1920 | 1.5364 | >0.9999 | 0.3178 |
| 20+/day | 1.3652 | 1.2828 | 1.4529 | 1.3693 | 1.2767 | 1.4687 | 1.4508 | 1.2551 | 1.6770 | >0.9999 | 0.0405 |
| **Alcohol intake frequency** | | | | | | | | | | | |
| Never | 0.6624 | 0.6285 | 0.6981 | 0.6645 | 0.6265 | 0.7048 | 0.6660 | 0.5922 | 0.7490 | 0.0324 | 0.0015 |
| Special occasions | 0.7565 | 0.7240 | 0.7904 | 0.7546 | 0.7189 | 0.7921 | 0.7807 | 0.7037 | 0.8662 | >0.9999 | 0.0689 |
| 1-3/month | 0.9201 | 0.8821 | 0.9597 | 0.9123 | 0.8716 | 0.9549 | 0.9756 | 0.8743 | 1.0887 | 0.8692 | 0.0322 |
| 1-2/week | Ref | – | – | Ref | – | – | Ref | – | – | – | – |
| 3-4/week | 1.0520 | 1.0178 | 1.0875 | 1.0384 | 1.0017 | 1.0765 | 1.1298 | 1.0374 | 1.2304 | >0.9999 | 0.3609 |
| Daily/almost daily | 1.0264 | 0.9913 | 1.0628 | 1.0044 | 0.9665 | 1.0437 | 1.1712 | 1.0773 | 1.2732 | <0.0001 | <0.0001 |
| **BMI** ($kg/m^2$) | 0.9045 | 0.9020 | 0.9069 | 0.9025 | 0.8999 | 0.9052 | 0.9180 | 0.9116 | 0.9245 | <0.0001 | <0.0001 |
| **Smoking status** | | | | | | | | | | | |
| Never | Ref | – | – | Ref | – | – | Ref | – | – | – | – |
| Former | 0.8128 | 0.7919 | 0.8342 | 0.8170 | 0.7938 | 0.8409 | 0.7808 | 0.7343 | 0.8302 | >0.9999 | 0.6825 |
| Current | 0.5029 | 0.4825 | 0.5242 | 0.4964 | 0.4748 | 0.5188 | 0.5844 | 0.5196 | 0.6574 | <0.0001 | <0.0001 |

*Note:* Estimates from Model 3 (i.e. including all explanatory variables). Bonferroni-adjusted (~99.9%) confidence intervals. OR = odds ratio; CI = confidence interval; BMI = body mass index. For categorical explanatory variables the odds ratios indicate the changes in odds of reporting better self-rated health associated with the explanatory variable group relative to the reference group. Odds ratios for continuous explanatory variables indicate proportional odds ratios for a 1-unit increase in the explanatory variable on level of self-rated health. [1]number of days per week engaging in these activities for 10+ minutes continuously.

Supplement e25-D. **Environmental exposures associated with self-rated health stratified by age**

| | All participants | | | Below 65 | | | 65 and above | | | Interaction term | |
|---|---|---|---|---|---|---|---|---|---|---|---|
| Term | OR | Bonferroni-corrected CI | | OR | Bonferroni-corrected CI | | OR | Bonferroni-corrected CI | | $p_{Bonf.}$ | $p_{BH}$ |
| **$PM_{2.5}$** | 1.0283 | 1.0047 | 1.0525 | 1.0290 | 1.0031 | 1.0555 | 1.0195 | 0.9631 | 1.0793 | 0.0001 | <0.0001 |
| **$PM_{10}$** | 0.9979 | 0.9904 | 1.0055 | 0.9959 | 0.9877 | 1.0042 | 1.0074 | 0.9889 | 1.0264 | >0.9999 | 0.4056 |
| **$NO_2$** | 0.9948 | 0.9910 | 0.9986 | 0.9959 | 0.9917 | 1.0000 | 0.9908 | 0.9816 | 1.0000 | <0.0001 | <0.0001 |
| **$L_{den}$** | 1.0025 | 0.9993 | 1.0058 | 1.0032 | 0.9996 | 1.0067 | 0.9996 | 0.9917 | 1.0076 | 0.0007 | <0.0001 |
| **Greenspace** 1000m | 1.0007 | 0.9998 | 1.0015 | 1.0008 | 0.9999 | 1.0018 | 1.0000 | 0.9978 | 1.0021 | 0.0566 | 0.0024 |

*Note:* Estimates from Model 3 (i.e. including all explanatory variables). Bonferroni-adjusted (~99.9%) confidence intervals. OR = odds ratio; CI = confidence interval; PM = particulate matter; $NO_2$ = nitrogen dioxide; $L_{den}$ = day-evening-night noise level. Odds ratios indicate proportional odds ratios for a 1-unit increase in the explanatory variable on level of self-rated health.

**Supplement e26. Confidence interval plots self-rated health stratified by age**

Sociodemographic and psychosocial factors

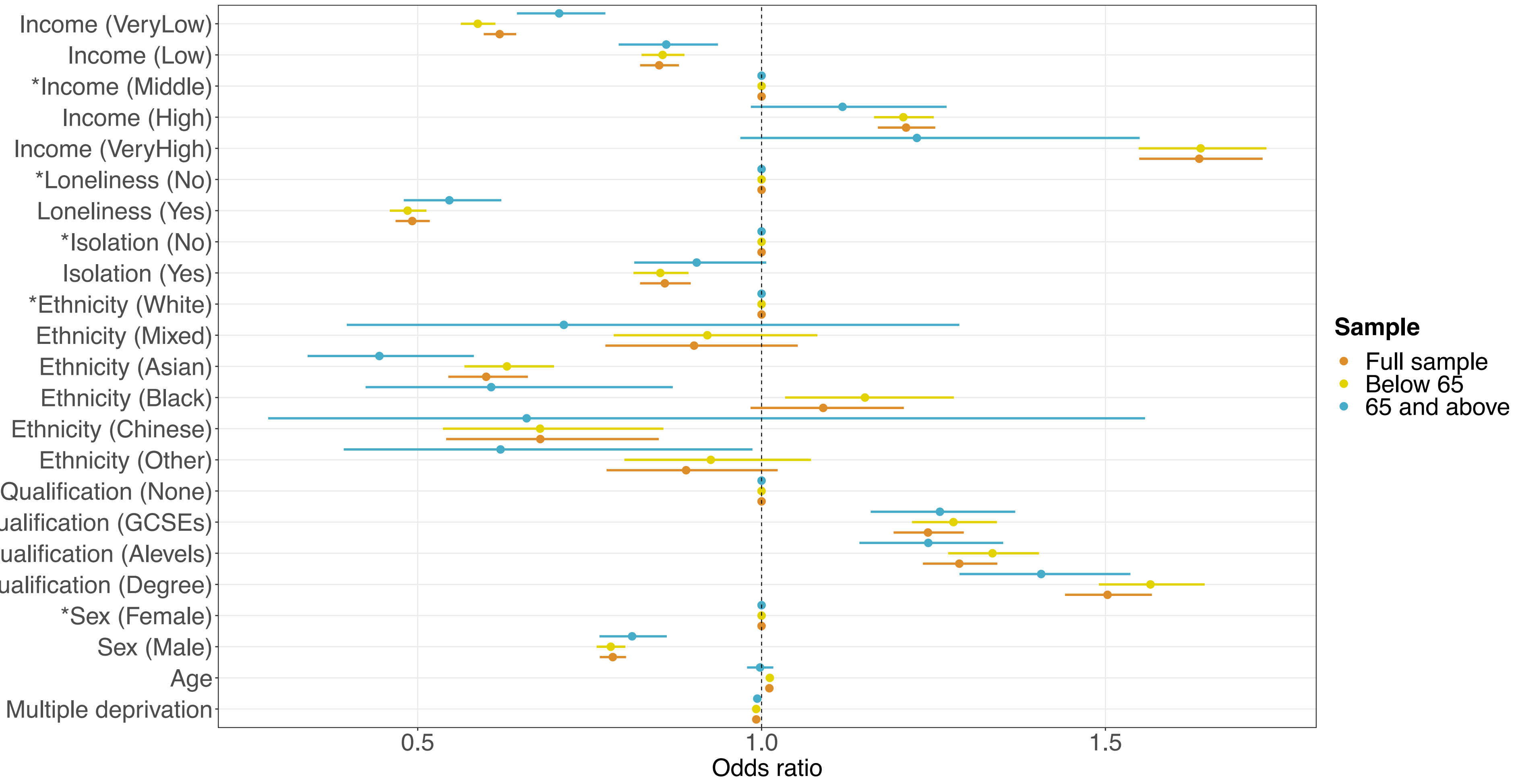


**Supplement e26-A.** Sociodemographic characteristics and psychosocial factors associated with self-rated health, stratified by age. Confidence interval plot (odds ratio ± Bonferroni-adjusted (~99.9%) confidence intervals) for Model 3 (i.e. including all explanatory variables). GCSEs = general certificate of secondary education. *Indicates reference group for categorical explanatory variables. Annual household income groups: very low (<£18 000), low (£18 000–30 999), middle (£31 000–51 999), high (£52 000–100 000) and very high (>£100 000). 'GCSEs' also includes O levels and certificate of secondary education (CSE). 'A levels' also includes national vocational qualification (NVQ), higher national diploma (HND), higher national certificate (HNC) and 'other professional qualifications'.

Lifestyle factors

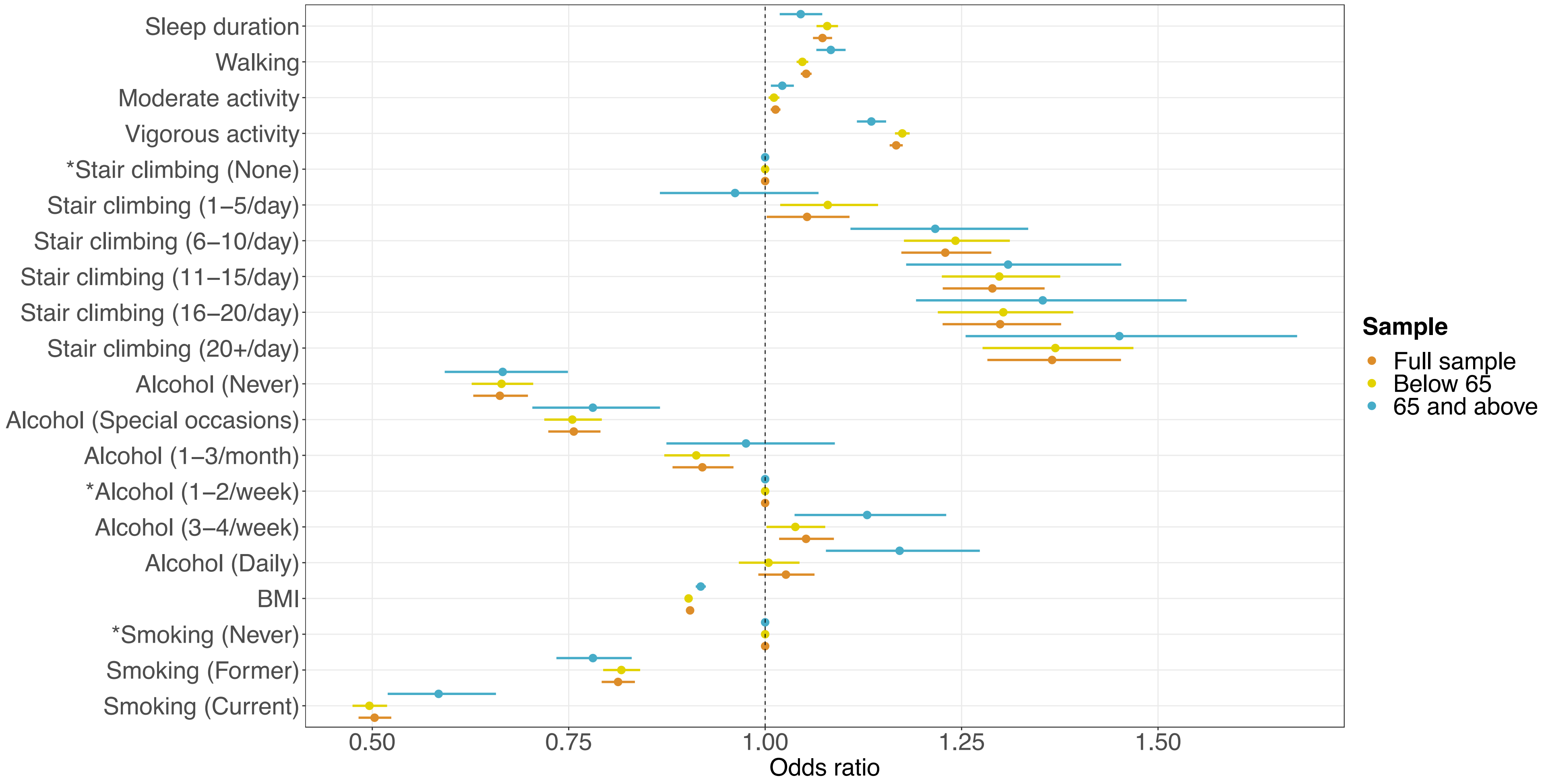


**Supplement e26-B.** Lifestyle factors associated with self-rated health, stratified by age. Confidence interval plot (odds ratio ± Bonferroni-adjusted (~99.9%) confidence intervals) for Model 3 (i.e. including all explanatory variables). BMI = body mass index. *Indicates reference group for categorical explanatory variables.

Environmental exposures

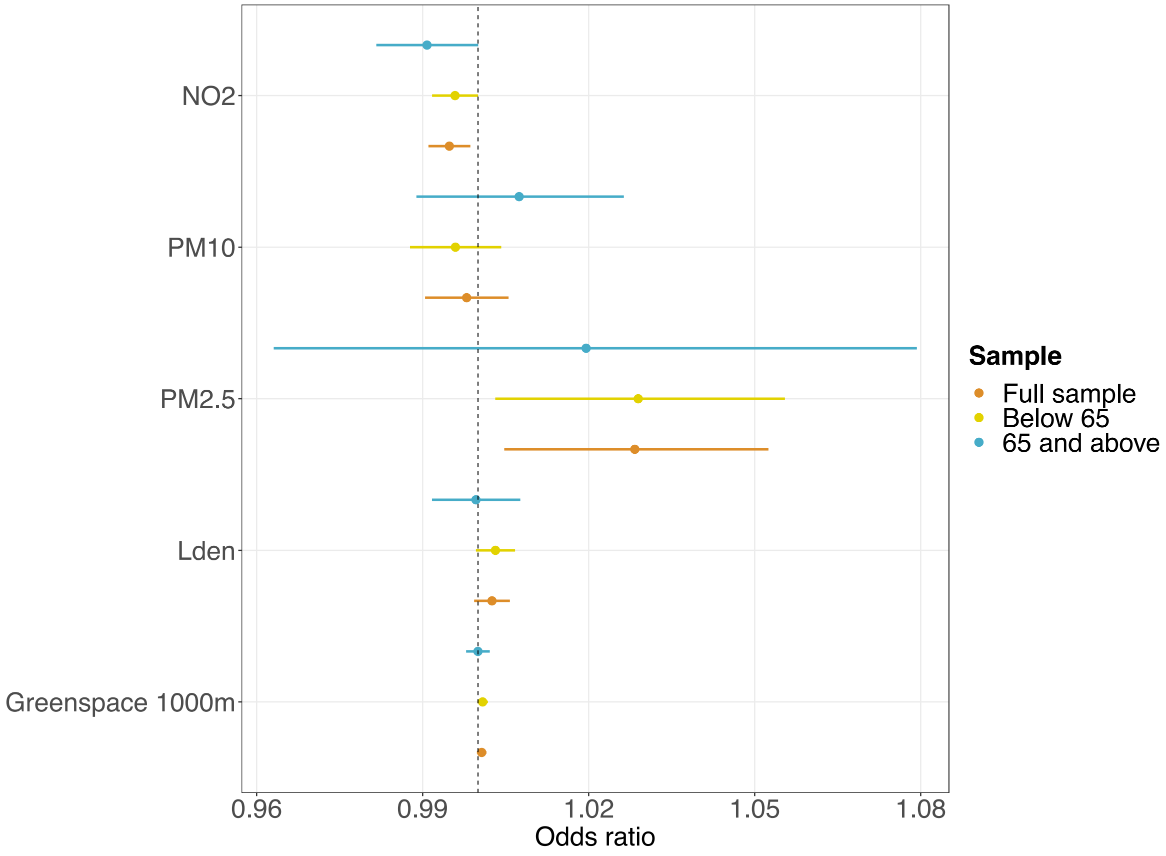


**Supplement e26-C.** Environmental exposures associated with self-rated health, stratified by age. Confidence interval plot (odds ratio ± Bonferroni-adjusted (~99.9%) confidence intervals) for Model 3 (i.e. including all explanatory variables). PM = particulate matter; $NO_2$ = nitrogen dioxide; $L_{den}$ = day-evening-night noise level.

**Supplement e27. Baseline characteristics prospective samples**

Supplement e27. **Baseline characteristics of prospective samples**

| | Analytical sample ($N$ = 307 378) | Follow-up t1 ($N$ = 16 058) | Follow-up t2 ($N$ = 32 617) |
|---|---|---|---|
| **Self-rated health** | | | |
| Poor | 11 066 (3.6%) | 469 (2.9%) | 677 (2.1%) |
| Fair | 59 169 (19.2%) | 2 834 (17.6%) | 5 197 (15.9%) |
| Good | 182 699 (59.4%) | 9 790 (61.0%) | 20 653 (63.3%) |
| Excellent | 54 444 (17.7%) | 2 965 (18.5%) | 6 090 (18.7%) |
| Sociodemographic characteristics | | | |
| **Age** | | | |
| Mean (SD) | 56.10 (8.07) | 56.92 (7.39) | 54.84 (7.46) |
| Range | 38.00-73.00 | 40.00-73.00 | 40.00-70.00 |
| **Sex** | | | |
| Female | 159 574 (51.9%) | 7 951 (49.5%) | 16 344 (50.1%) |
| Male | 147 804 (48.1%) | 8 107 (50.5%) | 16 273 (49.9%) |
| **Ethnicity** | | | |
| White | 293 565 (95.5%) | 15 748 (98.1%) | 31 768 (97.4%) |
| Mixed-race | 1 766 (0.6%) | 58 (0.4%) | 139 (0.4%) |
| Black | 4 257 (1.4%) | 63 (0.4%) | 191 (0.6%) |
| Asian | 4 755 (1.5%) | 84 (0.5%) | 298 (0.9%) |
| Chinese | 818 (0.3%) | 37 (0.2%) | 82 (0.3%) |
| Other | 2 217 (0.7%) | 68 (0.4%) | 139 (0.4%) |
| **Highest qualification** | | | |
| None | 39 828 (13.0%) | 1 156 (7.2%) | 1 854 (5.7%) |
| O levels/GCSEs/CSEs | 84 448 (27.5%) | 3 620 (22.5%) | 7 748 (23.8%) |
| A levels/NVQ/HND/HNC[1] | 72 584 (23.6%) | 3 819 (23.8%) | 7 608 (23.3%) |
| Degree | 110 518 (36.0%) | 7 463 (46.5%) | 15 407 (47.2%) |
| **Household income**[2] | | | |
| Very low | 82 338 (26.8%) | 4 679 (29.1%) | 9 910 (30.4%) |
| Low | 63 099 (20.5%) | 2 562 (16.0%) | 3 824 (11.7%) |
| Medium | 77 931 (25.4%) | 4 412 (27.5%) | 7 308 (22.4%) |
| High | 66 106 (21.5%) | 3 595 (22.4%) | 9 231 (28.3%) |
| Very high | 17 904 (5.8%) | 810 (5.0%) | 2 344 (7.2%) |
| **Multiple deprivation** | | | |
| Mean (SD) | 16.77 (13.33) | 16.51 (12.93) | 15.49 (12.16) |
| Range | 0.61-82.00 | 1.12-81.59 | 0.61-81.59 |
| Psychosocial factors | | | |
| **Loneliness** | | | |
| Not lonely | 289 901 (94.3%) | 15 303 (95.3%) | 31 145 (95.5%) |
| Lonely | 17 477 (5.7%) | 755 (4.7%) | 1 472 (4.5%) |
| **Social isolation** | | | |
| Not isolated | 280 931 (91.4%) | 14 930 (93.0%) | 30 337 (93.0%) |
| Isolated | 26 447 (8.6%) | 1 128 (7.0%) | 2 280 (7.0%) |
| Lifestyle factors | | | |
| **Smoking status** | | | |
| Never | 168 475 (54.8%) | 9 478 (59.0%) | 19 674 (60.3%) |
| Former | 108 638 (35.3%) | 5 596 (34.8%) | 10 910 (33.4%) |
| Current | 30 265 (9.8%) | 984 (6.1%) | 2 033 (6.2%) |
| **Stair climbing frequency** | | | |
| None | 24 049 (7.8%) | 1 042 (6.5%) | 1 977 (6.1%) |
| 1-5/day | 58 267 (19.0%) | 2 670 (16.6%) | 5 476 (16.8%) |
| 6-10/day | 115 982 (37.7%) | 6 165 (38.4%) | 12 777 (39.2%) |
| 11-15/day | 60 315 (19.6%) | 3 421 (21.3%) | 6 939 (21.3%) |
| 16-20/day | 27 609 (9.0%) | 1 600 (10.0%) | 3 141 (9.6%) |
| 20+/day | 21 156 (6.9%) | 1 160 (7.2%) | 2 307 (7.1%) |
| **Alcohol intake frequency** | | | |
| Never | 78 777 (25.6%) | 3 978 (24.8%) | 8 327 (25.5%) |
| Special occasions | 20 423 (6.6%) | 822 (5.1%) | 1 457 (4.5%) |
| 1-3/month | 31 526 (10.3%) | 1 343 (8.4%) | 2 569 (7.9%) |
| 1-2/week | 33 798 (11.0%) | 1 699 (10.6%) | 3 514 (10.8%) |
| 3-4/week | 75 251 (24.5%) | 4 496 (28.0%) | 9 291 (28.5%) |
| Daily/almost daily | 67 603 (22.0%) | 3 720 (23.2%) | 7 459 (22.9%) |
| **Sleep duration** (hours/day) | | | |
| Mean (SD) | 7.16 (1.06) | 7.20 (0.98) | 7.16 (0.96) |
| Range | 1.00-20.00 | 1.00-15.00 | 1.00-18.00 |
| **BMI** ($kg/m^2$) | | | |
| Mean (SD) | 27.27 (4.66) | 26.83 (4.46) | 26.68 (4.27) |
| Range | 12.80-67.30 | 15.80-56.60 | 15.20-63.60 |
| **Walking** (days/week)[3] | | | |
| Mean (SD) | 5.36 (1.95) | 5.17 (2.02) | 5.21 (2.00) |
| Range | 0.00-7.00 | 0.00-7.00 | 0.00-7.00 |
| **Moderate activity** (days/week)[3] | | | |
| Mean (SD) | 3.59 (2.32) | 3.45 (2.28) | 3.44 (2.26) |
| Range | 0.00-7.00 | 0.00-7.00 | 0.00-7.00 |
| **Vigorous activity** (days/week)[3] | | | |
| Mean (SD) | 1.88 (1.94) | 1.83 (1.85) | 1.89 (1.84) |
| Range | 0.00-7.00 | 0.00-7.00 | 0.00-7.00 |

| Environmental exposures | | | |
|---|---|---|---|
| **$PM_{2.5}$** | | | |
| Mean (SD) | 9.95 (1.04) | 9.96 (1.03) | 9.92 (1.04) |
| Range | 8.17-21.25 | 8.17-19.65 | 8.17-19.65 |
| **$PM_{10}$** | | | |
| Mean (SD) | 16.19 (1.88) | 15.92 (1.85) | 16.00 (1.85) |
| Range | 11.78-30.65 | 11.78-25.38 | 11.78-26.33 |
| **$NO_2$** | | | |
| Mean (SD) | 26.43 (7.56) | 25.99 (6.68) | 25.72 (7.15) |
| Range | 12.93-108.49 | 12.93-89.08 | 12.93-89.55 |
| **$L_{den}$** | | | |
| Mean (SD) | 56.01 (4.24) | 56.12 (4.33) | 55.98 (4.19) |
| Range | 51.55-89.29 | 51.57-81.36 | 51.56-81.67 |
| **Greenspace** 1000m | | | |
| Mean (SD) | 45.52 (21.77) | 48.37 (20.62) | 47.95 (21.77) |
| Range | 4.49-99.19 | 8.90-98.29 | 4.96-98.98 |

*Note:* All variables assessed at baseline except self-rated health in the third and fourth column assessed at t1 and t2, respectively. GCSEs = general certificate of secondary education; CSE = certificate of secondary education; NVQ = national vocational qualification; HND = higher national diploma; HNC = higher national certificate; BMI = body mass index; PM = particulate matter; $NO_2$ = nitrogen dioxide; $L_{den}$ = day-evening-night noise level. [1]also includes 'other professional qualifications'. [2]Annual household income groups: very low (<£18 000), low (£18 000–30 999), middle (£31 000–51 999), high (£52 000–100 000) and very high (>£100 000). [3]number of days per week engaging in these activities for 10+ minutes continuously.

**Supplement e28. Regression tables self-rated health t1**

Sociodemographic characteristics

Supplement e28-A. **Sociodemographic characteristics at baseline associated with self-rated health at t1**

| | Model 1 | | | Model 2 | | | Model 3 | | |
|---|---|---|---|---|---|---|---|---|---|
| Term | OR | Bonferroni-corrected CI | | OR | Bonferroni-corrected CI | | OR | Bonferroni-corrected CI | |
| **Household income**[1] | | | | | | | | | |
| Very low | 0.6337 | 0.5411 | 0.7421 | 0.6173 | 0.5256 | 0.7251 | 0.7396 | 0.6234 | 0.8773 |
| Low | 0.8404 | 0.7346 | 0.9613 | 0.8254 | 0.7201 | 0.9461 | 0.8789 | 0.7645 | 1.0103 |
| Middle | Ref | – | – | Ref | – | – | Ref | – | – |
| High | 1.3217 | 1.1460 | 1.5244 | 1.3423 | 1.1625 | 1.5501 | 1.2876 | 1.1111 | 1.4924 |
| Very high | 1.7091 | 1.3381 | 2.1817 | 1.7355 | 1.3573 | 2.2179 | 1.5147 | 1.1769 | 1.9488 |
| **Sex** | | | | | | | | | |
| Female | Ref | – | – | Ref | – | – | Ref | – | – |
| Male | 0.8676 | 0.7842 | 0.9598 | – | – | – | 0.8864 | 0.7965 | 0.9862 |
| **Age** | 0.9905 | 0.9837 | 0.9974 | – | – | – | 1.0006 | 0.9927 | 1.0085 |
| **Multiple deprivation** | 0.9853 | 0.9814 | 0.9892 | 0.9847 | 0.9808 | 0.9887 | 0.9964 | 0.9920 | 1.0009 |
| **Ethnicity** | | | | | | | | | |
| White | Ref | – | – | Ref | – | – | Ref | – | – |
| Mixed-race | 0.5693 | 0.2410 | 1.3718 | 0.5348 | 0.2258 | 1.2915 | 0.7006 | 0.2942 | 1.6803 |
| Asian | 0.6917 | 0.3451 | 1.4062 | 0.6640 | 0.3309 | 1.3514 | 0.6824 | 0.3359 | 1.3994 |
| Black | 0.9514 | 0.4177 | 2.1879 | 0.8885 | 0.3899 | 2.0446 | 1.5064 | 0.6503 | 3.5222 |
| Chinese | 0.8760 | 0.3190 | 2.4515 | 0.8202 | 0.2984 | 2.2975 | 0.7685 | 0.2734 | 2.1899 |
| Other | 0.6262 | 0.2924 | 1.3657 | 0.6158 | 0.2876 | 1.3422 | 0.6499 | 0.2965 | 1.4386 |
| **Highest qualification** | | | | | | | | | |
| None | Ref | – | – | Ref | – | – | Ref | – | – |
| O levels/GCSEs/CSEs | 1.2827 | 1.0335 | 1.5911 | 1.2291 | 0.9870 | 1.5296 | 1.0969 | 0.8765 | 1.3722 |
| A levels/NVQ/HND/HNC[2] | 1.3799 | 1.1130 | 1.7098 | 1.3360 | 1.0748 | 1.6597 | 1.1282 | 0.9018 | 1.4110 |
| Degree | 1.8163 | 1.4821 | 2.2247 | 1.7446 | 1.4179 | 2.1455 | 1.2253 | 0.9856 | 1.5230 |

*Note:* Bonferroni-adjusted (~99.9%) confidence intervals. OR = odds ratio; CI = confidence interval; GCSEs = general certificate of secondary education; CSE = certificate of secondary education; NVQ = national vocational qualification; HND = higher national diploma; HNC = higher national certificate. For categorical explanatory variables the odds ratios indicate the changes in odds of reporting better self-rated health associated with the explanatory variable group relative to the reference group. Odds ratios for continuous explanatory variables indicate proportional odds ratios for a 1-unit increase in the explanatory variable on level of self-rated health. [1]Annual household income groups: very low (<£18 000), low (£18 000–30 999), middle (£31 000–51 999), high (£52 000–100 000) and very high (>£100 000). [2]also includes 'other professional qualifications'.
Model 1 – only individual explanatory variables; Model 2 – adjusted for age and sex; Model 3 – all explanatory variables. *All models adjusted for number of days between t0 and t1.

| Supplement e28-B. **Psychosocial factors at baseline associated with self-rated health at t1** | | | | | | | | | |
|---|---|---|---|---|---|---|---|---|---|
| | Model 1 | | | Model 2 | | | Model 3 | | |
| Term | OR | Bonferroni-corrected CI | | OR | Bonferroni-corrected CI | | OR | Bonferroni-corrected CI | |
| **Loneliness** | | | | | | | | | |
| Not lonely | Ref | – | – | Ref | – | – | Ref | – | – |
| Lonely | 0.4145 | 0.3277 | 0.5249 | 0.4120 | 0.3256 | 0.5218 | 0.5321 | 0.4178 | 0.6782 |
| **Social isolation** | | | | | | | | | |
| Not isolated | Ref | – | – | Ref | – | – | Ref | – | – |
| Isolated | 0.5918 | 0.4851 | 0.7225 | 0.5928 | 0.4857 | 0.7240 | 0.8688 | 0.7065 | 1.0688 |

*Note:* Bonferroni-adjusted (~99.9%) confidence intervals. OR = odds ratio; CI = confidence interval. Odds ratios indicate the changes in odds of reporting better self-rated health associated with the explanatory variable group relative to the reference group.
Model 1 – only individual explanatory variables.
Model 2 – adjusted for age and sex.
Model 3 – all explanatory variables.
*All models adjusted for number of days between t0 and t1.

Supplement e28-C. **Lifestyle factors at baseline associated with self-rated health at t1**

| | Model 1 | | | Model 2 | | | Model 3 | | |
|---|---|---|---|---|---|---|---|---|---|
| Term | OR | Bonferroni-corrected CI | | OR | Bonferroni-corrected CI | | OR | Bonferroni-corrected CI | |
| **Sleep duration** (hours/day) | 1.0941 | 1.0380 | 1.1534 | 1.0998 | 1.0431 | 1.1595 | 1.1018 | 1.0447 | 1.1621 |
| **Physical activity** (days/week)[1] | | | | | | | | | |
| Walking | 1.0986 | 1.0711 | 1.1268 | 1.1016 | 1.0740 | 1.1300 | 1.0589 | 1.0300 | 1.0886 |
| Moderate activity | 1.0855 | 1.0615 | 1.1100 | 1.0893 | 1.0651 | 1.1141 | 1.0083 | 0.9809 | 1.0364 |
| Vigorous activity | 1.1803 | 1.1478 | 1.2138 | 1.1857 | 1.1528 | 1.2195 | 1.1313 | 1.0947 | 1.1691 |
| **Stair climbing frequency** | | | | | | | | | |
| None | Ref | – | – | Ref | – | – | Ref | – | – |
| 1-5/day | 0.9367 | 0.7417 | 1.1823 | 0.9110 | 0.7204 | 1.1514 | 0.9228 | 0.7272 | 1.1707 |
| 6-10/day | 1.3028 | 1.0521 | 1.6122 | 1.2673 | 1.0224 | 1.5701 | 1.1083 | 0.8904 | 1.3791 |
| 11-15/day | 1.5277 | 1.2186 | 1.9146 | 1.4856 | 1.1840 | 1.8635 | 1.1678 | 0.9259 | 1.4725 |
| 16-20/day | 1.5523 | 1.2038 | 2.0013 | 1.5070 | 1.1676 | 1.9445 | 1.1308 | 0.8714 | 1.4673 |
| 20+/day | 1.7904 | 1.3621 | 2.3528 | 1.7285 | 1.3137 | 2.2738 | 1.2175 | 0.9191 | 1.6126 |
| **Alcohol intake frequency** | | | | | | | | | |
| Never | 0.6707 | 0.5239 | 0.8592 | 0.6636 | 0.5182 | 0.8506 | 0.7134 | 0.5541 | 0.9188 |
| Special occasions | 0.6566 | 0.5369 | 0.8034 | 0.6363 | 0.5197 | 0.7793 | 0.7828 | 0.6362 | 0.9634 |
| 1-3/month | 0.7801 | 0.6480 | 0.9392 | 0.7642 | 0.6346 | 0.9204 | 0.8287 | 0.6864 | 1.0006 |
| 1-2/week | Ref | – | – | Ref | – | – | Ref | – | – |
| 3-4/week | 1.1624 | 1.0113 | 1.3362 | 1.1800 | 1.0265 | 1.3567 | 1.0416 | 0.9034 | 1.2010 |
| Daily/almost daily | 1.0895 | 0.9415 | 1.2607 | 1.1385 | 0.9828 | 1.3189 | 1.0369 | 0.8910 | 1.2067 |
| **BMI** (kg/m$^2$) | 0.8924 | 0.8820 | 0.9029 | 0.8931 | 0.8827 | 0.9036 | 0.9091 | 0.8981 | 0.9202 |
| **Smoking status** | | | | | | | | | |
| Never | Ref | – | – | Ref | – | – | Ref | – | – |
| Former | 0.7472 | 0.6705 | 0.8327 | 0.7679 | 0.6880 | 0.8570 | 0.8176 | 0.7296 | 0.9161 |
| Current | 0.4688 | 0.3785 | 0.5811 | 0.4708 | 0.3799 | 0.5839 | 0.5221 | 0.4186 | 0.6516 |

*Note:* Bonferroni-adjusted (~99.9%) confidence intervals. OR = odds ratio; CI = confidence interval; BMI = body mass index. For categorical explanatory variables the odds ratios indicate the changes in odds of reporting better self-rated health associated with the explanatory variable group relative to the reference group. Odds ratios for continuous explanatory variables indicate proportional odds ratios for a 1-unit increase in the explanatory variable on level of self-rated health. [1]number of days per week engaging in these activities for 10+ minutes continuously.
Model 1 – only individual explanatory variables.
Model 2 – adjusted for age and sex.
Model 3 – all explanatory variables.
*All models adjusted for number of days between t0 and t1.

Supplement e28-D. **Environmental exposures at baseline associated with self-rated health at t1**

| | Model 1 | | | Model 2 | | | Model 3 | | |
|---|---|---|---|---|---|---|---|---|---|
| Term | OR | Bonferroni-corrected CI | | OR | Bonferroni-corrected CI | | OR | Bonferroni-corrected CI | |
| **$PM_{2.5}$** | 0.9111 | 0.8667 | 0.9579 | 0.9046 | 0.8603 | 0.9511 | 1.0122 | 0.9023 | 1.136 |
| **$PM_{10}$** | 0.9648 | 0.9386 | 0.9918 | 0.9636 | 0.9374 | 0.9906 | 0.9942 | 0.9601 | 1.030 |
| **$NO_2$** | 0.9844 | 0.9766 | 0.9923 | 0.9831 | 0.9753 | 0.9910 | 0.9896 | 0.9703 | 1.009 |
| **$L_{den}$** | 0.9928 | 0.9814 | 1.0044 | 0.9924 | 0.9810 | 1.0040 | 1.0025 | 0.9888 | 1.016 |
| **Greenspace** 1000m | 1.0015 | 0.9989 | 1.0040 | 1.0018 | 0.9992 | 1.0044 | 0.9975 | 0.9935 | 1.001 |

*Note:* Bonferroni-adjusted (~99.9%) confidence intervals. OR = odds ratio; CI = confidence interval; PM = particulate matter; $NO_2$ = nitrogen dioxide; $L_{den}$ = day-evening-night noise level. Odds ratios for continuous explanatory variables indicate proportional odds ratios for a 1-unit increase in the explanatory variable on level of self-rated health.
Model 1 – only individual explanatory variables.
Model 2 – adjusted for age and sex.
Model 3 – all explanatory variables.
*All models adjusted for number of days between t0 and t1.

**Supplement e29. Regression tables self-rated health t2**

Sociodemographic characteristics

Supplement e29-A. **Sociodemographic characteristics at baseline associated with self-rated health at t2**

| | Model 1 | | | Model 2 | | | Model 3 | | |
|---|---|---|---|---|---|---|---|---|---|
| Term | OR | Bonferroni-corrected CI | | OR | Bonferroni-corrected CI | | OR | Bonferroni-corrected CI | |
| **Household income**[1] | | | | | | | | | |
| Very low | 0.6433 | 0.5680 | 0.7288 | 0.6003 | 0.5291 | 0.6812 | 0.7042 | 0.6166 | 0.8041 |
| Low | 0.8741 | 0.7905 | 0.9666 | 0.8314 | 0.7510 | 0.9204 | 0.8647 | 0.7795 | 0.9591 |
| Middle | Ref | – | – | Ref | – | – | Ref | – | – |
| High | 1.2214 | 1.1114 | 1.3423 | 1.2625 | 1.1482 | 1.3884 | 1.1531 | 1.0460 | 1.2713 |
| Very high | 1.7805 | 1.5338 | 2.0665 | 1.8603 | 1.6013 | 2.1608 | 1.5583 | 1.3344 | 1.8196 |
| **Sex** | | | | | | | | | |
| Female | Ref | – | – | Ref | – | – | Ref | – | – |
| Male | 0.8796 | 0.8184 | 0.9453 | – | – | – | 0.9112 | 0.8441 | 0.9836 |
| **Age** | 1.0027 | 0.9978 | 1.0076 | – | – | – | 1.0121 | 1.0065 | 1.0176 |
| **Multiple deprivation** | 0.9833 | 0.9804 | 0.9862 | 0.9833 | 0.9803 | 0.9862 | 0.9947 | 0.9914 | 0.9981 |
| **Ethnicity** | | | | | | | | | |
| White | Ref | – | – | Ref | – | – | Ref | – | – |
| Mixed-race | 0.9761 | 0.5584 | 1.7081 | 0.9690 | 0.5542 | 1.6963 | 1.0936 | 0.6267 | 1.9098 |
| Asian | 0.5379 | 0.3710 | 0.7837 | 0.5578 | 0.3844 | 0.8132 | 0.5071 | 0.3454 | 0.7469 |
| Black | 0.6076 | 0.3797 | 0.9797 | 0.6139 | 0.3831 | 0.9911 | 0.8968 | 0.5543 | 1.4566 |
| Chinese | 0.6796 | 0.3389 | 1.3796 | 0.6817 | 0.3399 | 1.3840 | 0.5738 | 0.2805 | 1.1857 |
| Other | 0.8768 | 0.5001 | 1.5421 | 0.8729 | 0.4976 | 1.5361 | 0.8945 | 0.5040 | 1.5911 |
| **Highest qualification** | | | | | | | | | |
| None | Ref | – | – | Ref | – | – | Ref | – | – |
| O levels/GCSEs/CSEs | 1.1908 | 1.0069 | 1.4077 | 1.2177 | 1.0275 | 1.4425 | 1.0646 | 0.8946 | 1.2666 |
| A levels/NVQ/HND/HNC[2] | 1.3584 | 1.1481 | 1.6066 | 1.3899 | 1.1732 | 1.6460 | 1.1558 | 0.9706 | 1.3762 |
| Degree | 1.8206 | 1.5513 | 2.1357 | 1.8744 | 1.5941 | 2.2032 | 1.3026 | 1.0988 | 1.5439 |

*Note:* Bonferroni-adjusted (~99.9%) confidence intervals. OR = odds ratio; CI = confidence interval; GCSEs = general certificate of secondary education; CSE = certificate of secondary education; NVQ = national vocational qualification; HND = higher national diploma; HNC = higher national certificate. For categorical explanatory variables the odds ratios indicate the changes in odds of reporting better self-rated health associated with the explanatory variable group relative to the reference group. Odds ratios for continuous explanatory variables indicate proportional odds ratios for a 1-unit increase in the explanatory variable on level of self-rated health. [1]Annual household income groups: very low (<£18 000), low (£18 000–30 999), middle (£31 000–51 999), high (£52 000–100 000) and very high (>£100 000). [2]also includes 'other professional qualifications'.
Model 1 – only individual explanatory variables; Model 2 – adjusted for age and sex; Model 3 – all explanatory variables. *All models adjusted for number of days between t0 and t2.

| Supplement e29-B. **Psychosocial factors at baseline associated with self-rated health at t2** | | | | | | | | | |
|---|---|---|---|---|---|---|---|---|---|
| | Model 1 | | | Model 2 | | | Model 3 | | |
| Term | OR | Bonferroni-corrected CI | | OR | Bonferroni-corrected CI | | OR | Bonferroni-corrected CI | |
| **Loneliness** | | | | | | | | | |
| Not lonely | Ref | – | – | Ref | – | – | Ref | – | – |
| Lonely | 0.3934 | 0.3318 | 0.4669 | 0.3950 | 0.3331 | 0.4687 | 0.5103 | 0.4281 | 0.6086 |
| **Social isolation** | | | | | | | | | |
| Not isolated | Ref | – | – | Ref | – | – | Ref | – | – |
| Isolated | 0.6071 | 0.5271 | 0.6995 | 0.6099 | 0.5295 | 0.7027 | 0.8303 | 0.7172 | 0.9615 |

*Note:* Bonferroni-adjusted (~99.9%) confidence intervals. OR = odds ratio; CI = confidence interval. For categorical explanatory variables the odds ratios indicate the changes in odds of reporting better self-rated health associated with the explanatory variable group relative to the reference group.
Model 1 – only individual explanatory variables.
Model 2 – adjusted for age and sex.
Model 3 – all explanatory variables.
*All models adjusted for number of days between t0 and t2.

Supplement e29-C. **Lifestyle factors at baseline associated with self-rated health at t2**

| | Model 1 | | | Model 2 | | | Model 3 | | |
|---|---|---|---|---|---|---|---|---|---|
| Term | OR | Bonferroni-corrected CI | | OR | Bonferroni-corrected CI | | OR | Bonferroni-corrected CI | |
| **Sleep duration** (hours/day) | 1.1006 | 1.0589 | 1.1438 | 1.0969 | 1.0553 | 1.1401 | 1.0756 | 1.0345 | 1.1183 |
| **Physical activity** (days/week)[1] | | | | | | | | | |
| Walking | 1.0756 | 1.0561 | 1.0953 | 1.0736 | 1.0542 | 1.0934 | 1.0289 | 1.0087 | 1.0495 |
| Moderate activity | 1.0788 | 1.0617 | 1.0963 | 1.0777 | 1.0604 | 1.0952 | 1.0029 | 0.9832 | 1.0230 |
| Vigorous activity | 1.1661 | 1.1431 | 1.1896 | 1.1723 | 1.1491 | 1.1961 | 1.1278 | 1.1015 | 1.1547 |
| **Stair climbing frequency** | | | | | | | | | |
| None | Ref | – | – | Ref | – | – | Ref | – | – |
| 1-5/day | 0.9418 | 0.7937 | 1.1173 | 0.9570 | 0.8059 | 1.1362 | 0.9539 | 0.8009 | 1.1360 |
| 6-10/day | 1.1940 | 1.0202 | 1.3973 | 1.2108 | 1.0336 | 1.4180 | 1.0481 | 0.8920 | 1.2315 |
| 11-15/day | 1.3456 | 1.1398 | 1.5883 | 1.3606 | 1.1517 | 1.6072 | 1.0756 | 0.9072 | 1.2752 |
| 16-20/day | 1.5273 | 1.2671 | 1.8408 | 1.5393 | 1.2762 | 1.8563 | 1.1764 | 0.9715 | 1.4245 |
| 20+/day | 1.5307 | 1.2540 | 1.8683 | 1.5464 | 1.2657 | 1.8891 | 1.1442 | 0.9325 | 1.4038 |
| **Alcohol intake frequency** | | | | | | | | | |
| Never | 0.7111 | 0.5895 | 0.8583 | 0.6996 | 0.5798 | 0.8446 | 0.8048 | 0.6646 | 0.9748 |
| Special occasions | 0.6698 | 0.5781 | 0.7762 | 0.6456 | 0.5568 | 0.7487 | 0.7999 | 0.6880 | 0.9300 |
| 1-3/month | 0.8298 | 0.7278 | 0.9461 | 0.8148 | 0.7145 | 0.9293 | 0.9051 | 0.7924 | 1.0338 |
| 1-2/week | Ref | – | – | Ref | – | – | Ref | – | – |
| 3-4/week | 1.1309 | 1.0254 | 1.2472 | 1.1380 | 1.0318 | 1.2552 | 1.0085 | 0.9126 | 1.1145 |
| Daily/almost daily | 1.0750 | 0.9689 | 1.1927 | 1.0902 | 0.9816 | 1.2107 | 0.9799 | 0.8795 | 1.0917 |
| **BMI** ($kg/m^2$) | 0.8848 | 0.8771 | 0.8926 | 0.8846 | 0.8769 | 0.8924 | 0.8992 | 0.8911 | 0.9074 |
| **Smoking status** | | | | | | | | | |
| Never | Ref | – | – | Ref | – | – | Ref | – | – |
| Former | 0.7758 | 0.7176 | 0.8385 | 0.7744 | 0.7158 | 0.8378 | 0.8330 | 0.7680 | 0.9035 |
| Current | 0.4923 | 0.4228 | 0.5733 | 0.5010 | 0.4302 | 0.5837 | 0.5724 | 0.4893 | 0.6698 |

*Note:* Bonferroni-adjusted (~99.9%) confidence intervals. OR = odds ratio; CI = confidence interval; BMI = body mass index. For categorical explanatory variables the odds ratios indicate the changes in odds of reporting better self-rated health associated with the explanatory variable group relative to the reference group. Odds ratios for continuous explanatory variables indicate proportional odds ratios for a 1-unit increase in the explanatory variable on level of self-rated health. [1]number of days per week engaging in these activities for 10+ minutes continuously.
Model 1 – only individual explanatory variables.
Model 2 – adjusted for age and sex.
Model 3 – all explanatory variables.
*All models adjusted for number of days between t0 and t2.

Supplement e29-D. **Environmental exposures at baseline associated with self-rated health at t2**

| | Model 1 | | | Model 2 | | | Model 3 | | |
|---|---|---|---|---|---|---|---|---|---|
| Term | OR | Bonferroni-corrected CI | | OR | Bonferroni-corrected CI | | OR | Bonferroni-corrected CI | |
| **$PM_{2.5}$** | 0.9205 | 0.8889 | 0.9531 | 0.9213 | 0.8896 | 0.9540 | 0.9855 | 0.9140 | 1.063 |
| **$PM_{10}$** | 0.9821 | 0.9631 | 1.0014 | 0.9825 | 0.9635 | 1.0018 | 1.0174 | 0.9936 | 1.042 |
| **$NO_2$** | 0.9890 | 0.9840 | 0.9940 | 0.9891 | 0.9841 | 0.9942 | 0.9998 | 0.9872 | 1.013 |
| **$L_{den}$** | 0.9920 | 0.9835 | 1.0006 | 0.9921 | 0.9836 | 1.0007 | 0.9976 | 0.9878 | 1.008 |
| **Greenspace** 1000m | 1.0021 | 1.0004 | 1.0037 | 1.0021 | 1.0004 | 1.0037 | 1.0004 | 0.9978 | 1.003 |

*Note:* Bonferroni-adjusted (~99.9%) confidence intervals. OR = odds ratio; CI = confidence interval; PM = particulate matter; $NO_2$ = nitrogen dioxide; $L_{den}$ = day-evening-night noise level. Odds ratios for continuous explanatory variables indicate proportional odds ratios for a 1-unit increase in the explanatory variable on level of self-rated health.
Model 1 – only individual explanatory variables.
Model 2 – adjusted for age and sex.
Model 3 – all explanatory variables.
*All models adjusted for number of days between t0 and t2.

**Supplement e30. Additional analyses**

Descriptive statistics

Supplement e30. **Descriptive statistics additional analyses**

| Variable | Statistics |
|---|---|
| **Body fat percentage** | Mean (SD) |
| | 30.85 (8.46) |
| | Range |
| | 5-65.2 |
| **Alcohol intake frequency** | *n* (%) |
| Lifetime abstainers | 10 599 (3.45) |
| Current abstainers | 9 792 (3.19) |
| Special occasions | 31 526 (10.26) |
| 1-3/month | 33 798 (11.00) |
| 1-2/week | 78 777 (25.63) |
| 3-4/week | 75 251 (24.48) |
| Daily/almost daily | 67 603 (22.00) |
| **Current tobacco smoking** | *n* (%) |
| "Yes, on most or all days" | 21 778 (7.09) |
| "Only occasionally" | 8 487 (2.76) |
| "No" | 277 107 (90.15) |
| **Metabolic Equivalent Task minutes/week** | Median (IQR) |
| Walking | 693 (1089) |
| Moderate activity | 480 (1080) |
| Vigorous activity | 240 (960) |
| **$PM_{2.5}$** | *n* (%) |
| ≤10 $\mu g/m^3$ | 167 631 (54.54) |
| >10 $\mu g/m^3$ | 139 747 (45.46) |
| **$PM_{10}$** | *n* (%) |
| ≤20 $\mu g/m^3$ | 290 684 (94.57) |
| >20 $\mu g/m^3$ | 16 694 (5.43) |
| **$NO_2$** | *n* (%) |
| ≤40 $\mu g/m^3$ | 294 402 (95.78) |
| >40 $\mu g/m^3$ | 12 976 (4.22) |
| **$L_{den}$** | *n* (%) |
| ≤53 decibels | 61 017 (19.85) |
| >53 decibels | 246 361 (80.15) |
| **Greenspace 300m** | Mean (SD) |
| | 35.65 (23.46) |
| | Range |
| | 0.227-99.180 |

*Note:* SD = standard deviation; IQR = interquartile range; PM = particulate matter; $NO_2$ = nitrogen dioxide; $L_{den}$ = day-evening-night noise level.

**Supplement e31. Fitted probabilities**

Sociodemographic characteristics

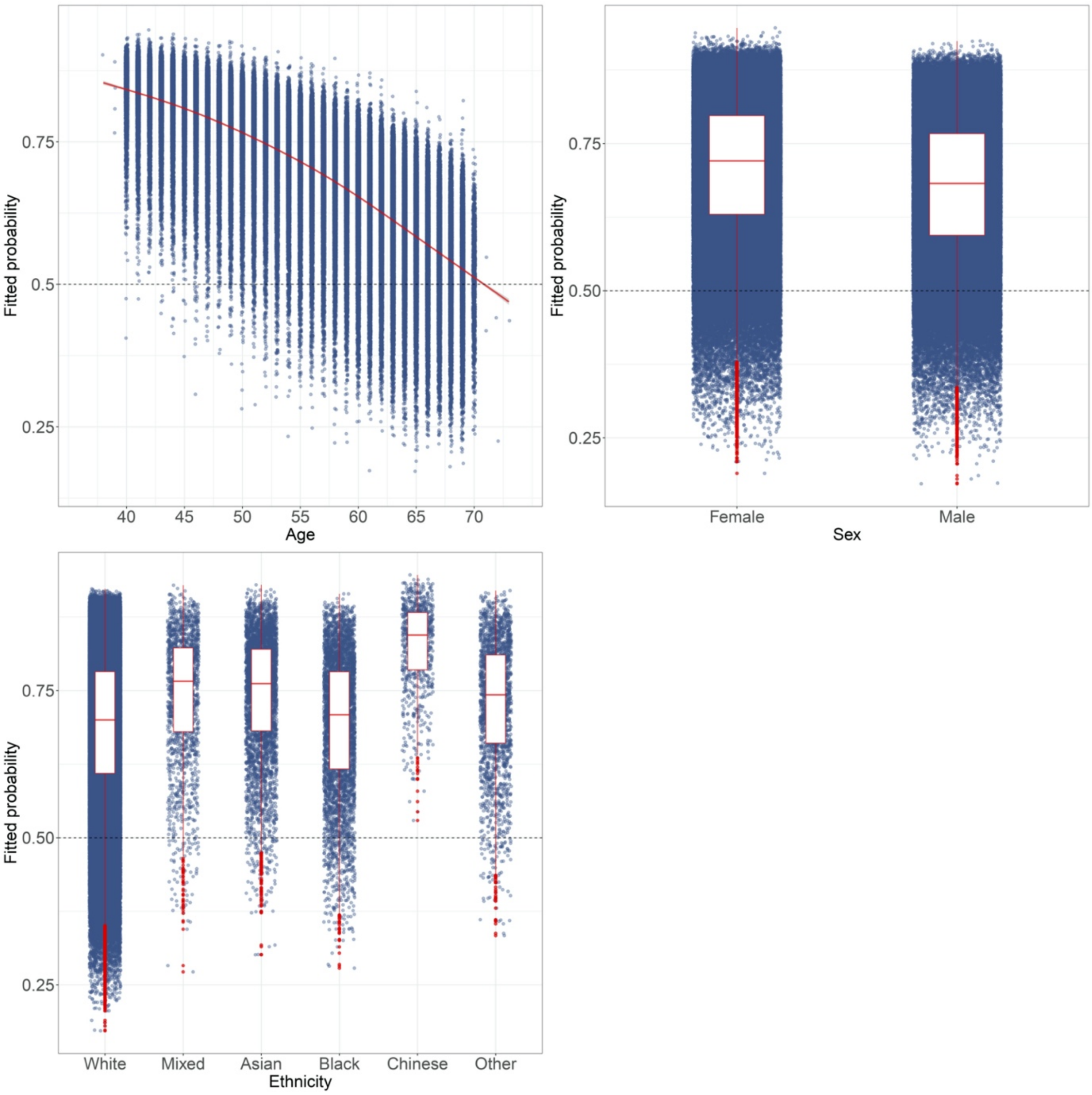


**Supplement e31-A.** Model 3 (i.e. including all explanatory variables) predicted probabilities for health status (healthy: ≥ 0.50; unhealthy <0.50) by sociodemographic characteristics. Figure includes generalised additive model curve with 95% confidence interval for continuous explanatory variables. Box-plot elements: centre line = median; box limits = 25th and 75th percentile; whiskers = 1.5x interquartile range; points = outliers.

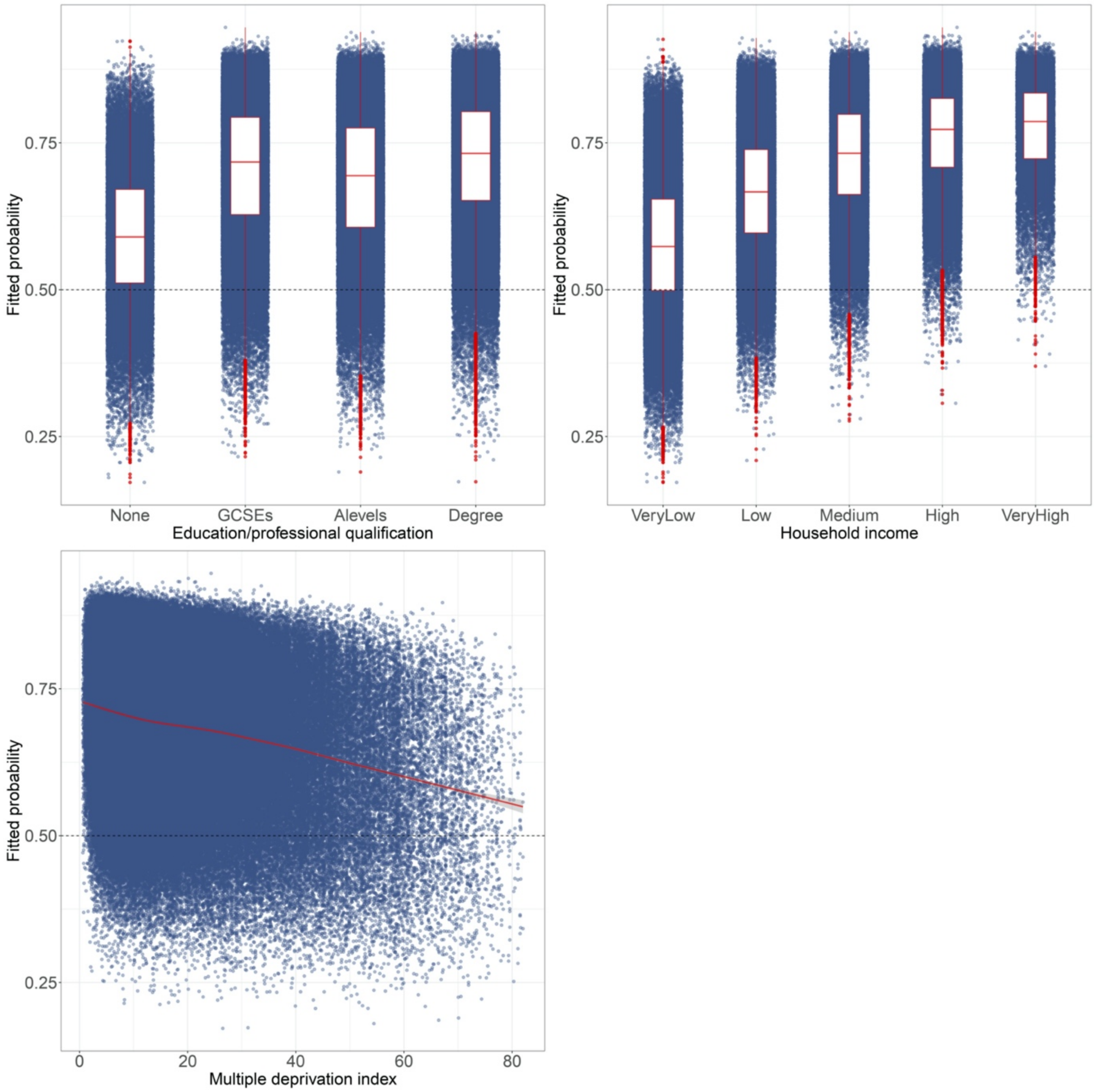


**Supplement e31-B.** Model 3 (i.e. including all explanatory variables) predicted probabilities for health status (healthy: ≥ 0.50; unhealthy <0.50) by sociodemographic characteristics. Figure includes generalised additive model curve with 95% confidence interval for continuous explanatory variables. Box-plot elements: centre line = median; box limits = 25th and 75th percentile; whiskers = 1.5x interquartile range; points = outliers.

Psychosocial factors

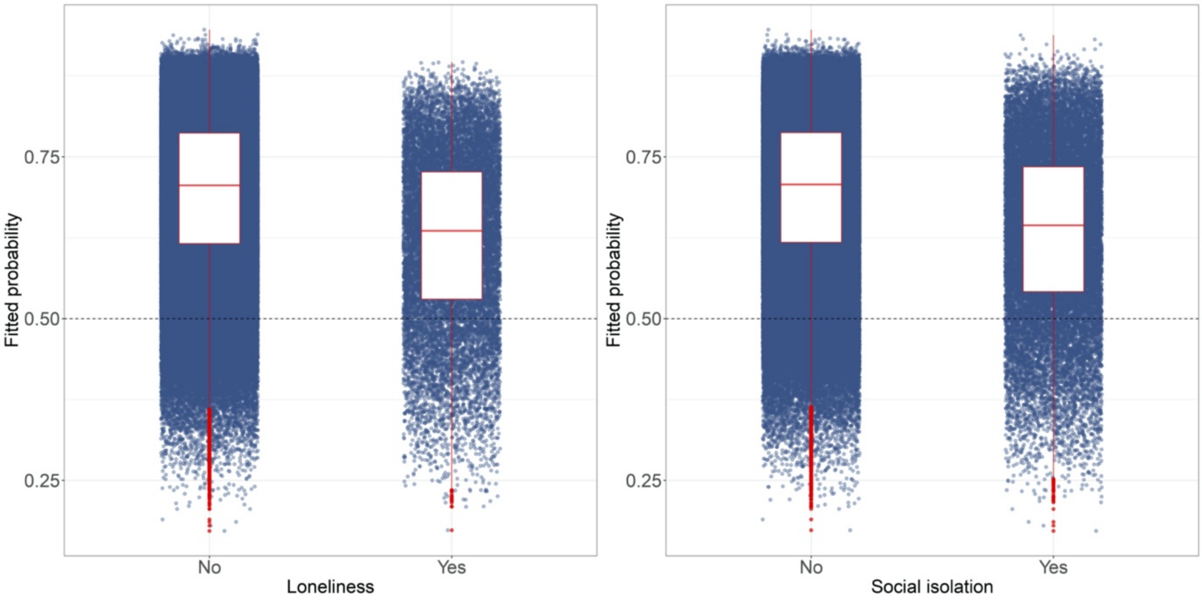


**Supplement e31-C.** Model 3 (i.e. including all explanatory variables) predicted probabilities for health status (healthy: ≥ 0.50; unhealthy <0.50) by psychosocial factors. Box-plot elements: centre line = median; box limits = 25th and 75th percentile; whiskers = 1.5x interquartile range; points = outliers.

Lifestyle factors

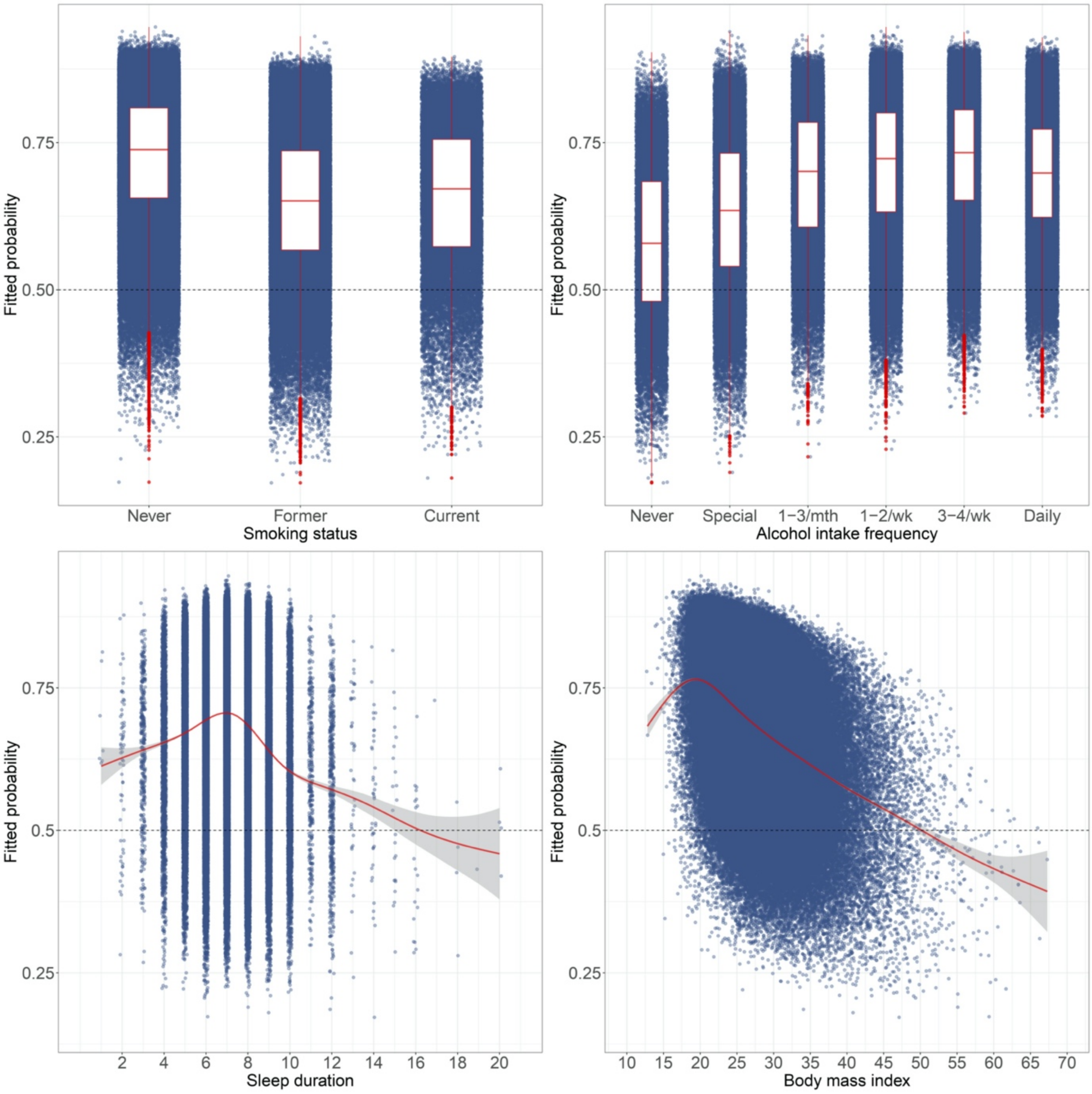


**Supplement e31-D.** Model 3 (i.e. including all explanatory variables) predicted probabilities for health status (healthy: ≥ 0.50; unhealthy <0.50) by lifestyle factors. Figure includes generalised additive model curve with 95% confidence interval for continuous explanatory variables. Box-plot elements: centre line = median; box limits = 25th and 75th percentile; whiskers = 1.5x interquartile range; points = outliers.

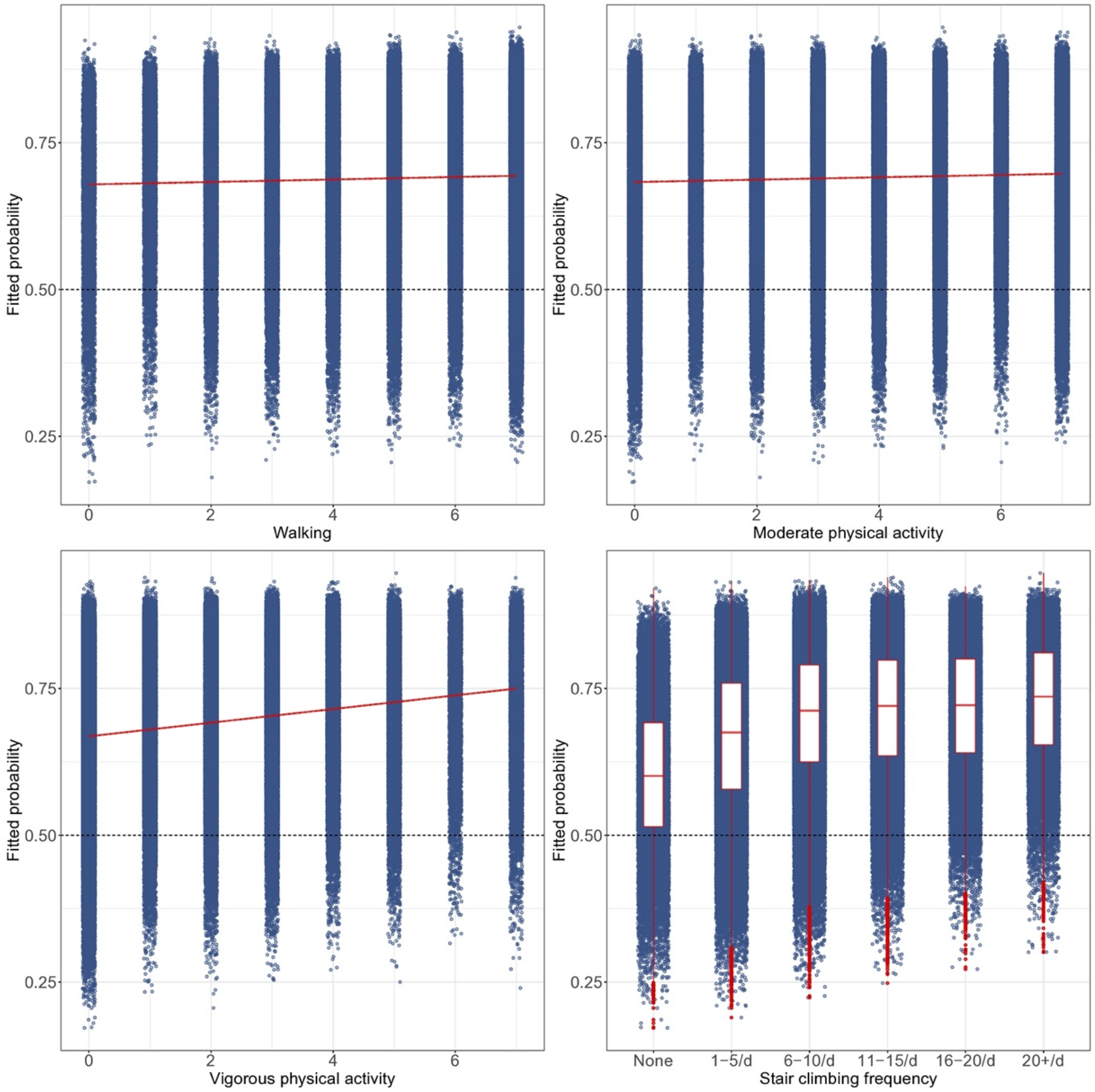


**Supplement e31-E.** Model 3 (i.e. including all explanatory variables) predicted probabilities for health status (healthy: ≥ 0.50; unhealthy <0.50) by lifestyle factors. Figure includes generalised additive model curve with 95% confidence interval for continuous explanatory variables. Box-plot elements: centre line = median; box limits = 25th and 75th percentile; whiskers = 1.5x interquartile range; points = outliers.

Environmental exposures

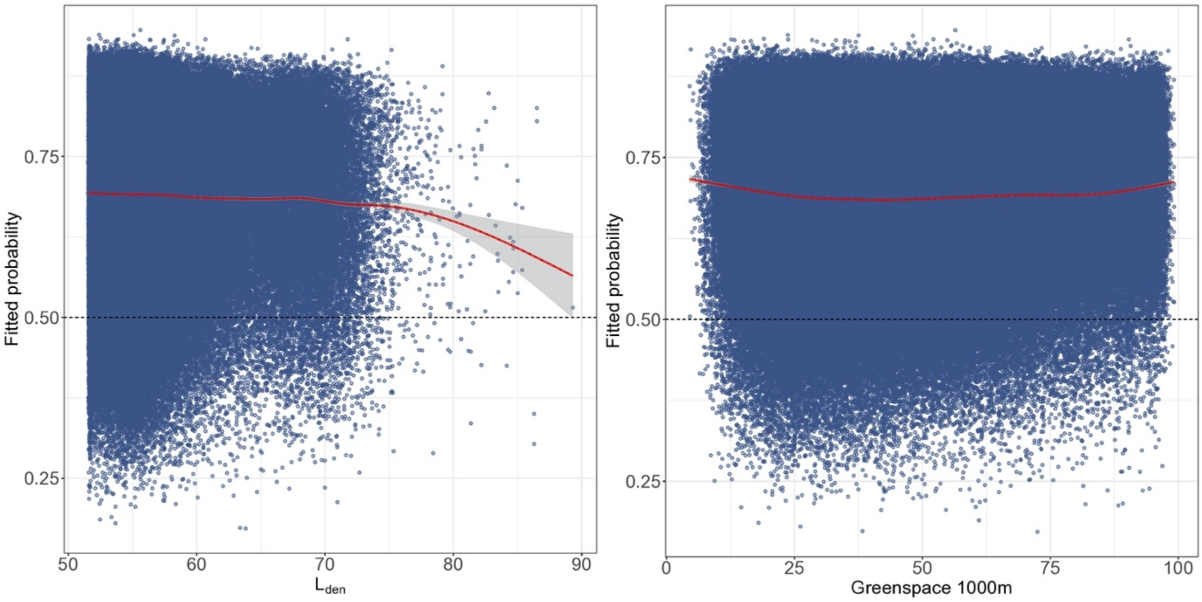


**Supplement e31-F.** Model 3 (i.e. including all explanatory variables) predicted probabilities for health status (healthy: ≥ 0.50; unhealthy <0.50) by environmental exposures. Figure includes generalised additive model curve with 95% confidence interval. $L_{den}$ = day-evening-night noise level.

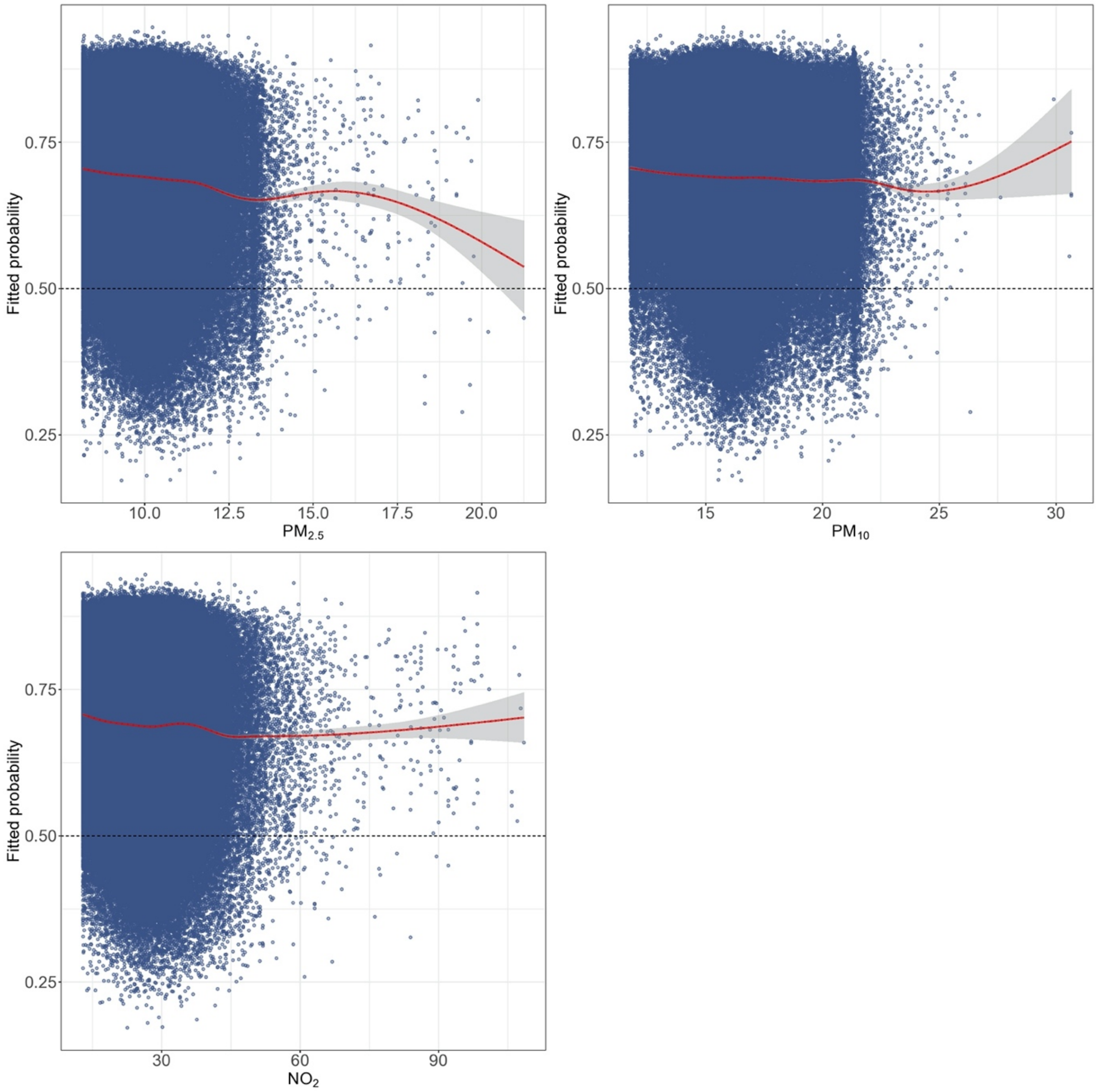


**Supplement e31-G.** Model 3 (i.e. including all explanatory variables) predicted probabilities for health status (healthy: ≥ 0.50; unhealthy <0.50) by environmental exposures. Figure includes generalised additive model curve with 95% confidence interval. PM = particulate matter; $NO_2$ = nitrogen dioxide.
